# Pre-MEASURE: *FLT3*-ITD and mutated *NPM1* measurable residual disease before allogeneic transplant in adults with AML in first remission

**DOI:** 10.1101/2022.10.21.22281322

**Authors:** Laura W Dillon, Gege Gui, Niveditha Ravindra, Zoë Wong, Georgia Andrew, Devdeep Mukherjee, Scott L. Zeger, Firas El Chaer, Stephen Spellman, Alan Howard, Karen Chen, Jeffery Auletta, Steven M. Devine, Antonio Martin Jimenez Jimenez, Marcos J.G. De Lima, Mark Robert Litzow, Partow Kebriaei, Wael Saber, Daniel Jordan Weisdorf, Kristin M. Page, Christopher S. Hourigan

**Author notes:** LWD and GG contributed equally to this work. **Corresponding Author:** Christopher S. Hourigan DM DPhil FRCP, Laboratory of Myeloid Malignancies, Hematology Branch, National Heart, Lung, and Blood Institute, National Institutes of Health, Bethesda, Maryland 20814-1476.

## Abstract

**Background:** Prevention of relapse for adults with Acute Myeloid Leukemia (AML) in first remission (CR1) is the most common indication for allogeneic hematopoietic cell transplantation (alloHCT). Measurable residual disease (MRD) detection has been associated with higher relapse rates, but testing is not standardized. Establishment of validated AML MRD criteria would allow harmonization across centers facilitating clinical trials and generalizable practice guidelines.

**Methods:** Patients aged 18 or older who underwent first alloHCT for *FLT3, NPM1, IDH1, IDH2* and/or *KIT* mutated AML in CR1 were eligible for this multicenter study. Residual mutations detected in remission using ultra-deep error-corrected next-generation DNA-sequencing (NGS-MRD) associated with increased relapse risk were validated in a second independent cohort. The impact of baseline characteristics on NGS-MRD results was evaluated.

**Results:** Pre-conditioning CR1 blood samples from 1075 patients were tested. Detection of residual *NPM1* and/or *FLT3*-ITD mutations before alloHCT was associated with worse outcomes after transplantation in both discovery (patients transplanted 2013-2017) and validation (patients transplanted 2018-2019) cohorts. In multivariate Cox regression analysis detection of residual *NPM1*m (relapse; HR: 4.9, 3.5-7.0, P<0.0001, survival; HR: 2.9, 2.2-4.0, P<0.0001), residual *FLT3*-ITD (relapse: HR: 4.2, 3.1-5.7, P<0.0001, survival; HR: 2.6, 2.0-3.6, P<0.0001), and receipt of non-melphalan containing reduced intensity conditioning were associated with inferior clinical outcomes.

**Conclusions:** Detection of residual *NPM1* and/or *FLT3*-ITD mutations in the blood of patients undergoing alloHCT for AML in CR1 is associated with increased relapse and worse survival. In those undergoing reduced intensity conditioning, melphalan may reduce the risk associated with testing NGS-MRD positive prior to transplantation.

## Introduction

Acute myeloid leukemia (AML) is a term given to a rare, highly fatal, group of blood cancers^1^. The genetic etiology of AML has been well characterized^2-5^, and genomic analysis performed at the time of AML diagnostic evaluation can help predict response to therapy^6-8^, identify patients eligible for specific molecularly targeted therapies^9-12^, and stratify patients for risk of subsequent relapse and death in those who achieve an initial complete remission to therapy^3,13,14^.

Maintenance of initial remission in AML is the most common indication for allogeneic hematopoietic cell transplantation (alloHCT) and generally recommended in first remission for all except for those unable, unwilling, or with the lowest expected rates of relapse after chemotherapy (“favorable-risk” disease)^14-17^. Despite this role as consolidative therapy, disease recurrence after alloHCT is common with multiple potentially associated factors identified including transplant type^18,19^, immunological mechanisms^20-23^, and suboptimal remission status^24,25^.

In 2017 the treatment response criteria for AML was updated^26^ in recognition of the growing evidence that results of measurable residual disease (MRD) testing can provide important additional stratification for subsequent relapse and survival outcomes in patients within cytomorphological complete remission, including prior to alloHCT^27-32^. However, there is currently no harmonized standard method for AML MRD testing^27^, and the evidence for the utility of novel approaches such as genetic sequencing^33-37^ has been insufficient to motivate widespread clinical adoption.

In this “Pre-MEASURE” study, we performed targeted ultra-deep DNA-sequencing to test the hypothesis that detection of residual AML-associated mutations in the blood of patients in first remission prior to alloHCT would be associated with increased relapse and worse survival after transplantation.

## Methods

### STUDY DESIGN

The study was designed by the senior and first two authors, who wrote the manuscript with input from the other authors. The authors vouch for the completeness and accuracy of the data and analysis. No one who is not an author contributed to the manuscript. There was no commercial support for the study.

### CLINICAL COHORT

To be included in the study, patients must be 18 years or older and have undergone first alloHCT for *NPM1, FLT3, IDH1, IDH2* and/or *KIT* mutated AML in CR1 at a Center for International Blood and Marrow Transplant Research (CIBMTR) reporting site. In addition, patients must have provided written informed consent for participation in the CIBMTR database (NCT01166009) and repository (NCT04920474) protocols, have a remission blood sample collected within 100 days prior to transplant available in the CIBMTR repository, and have relapse and survival data available within the CIBMTR database.

### NEXT-GENERATION SEQUENCING FOR RESIDUAL MUTATION DETECTION

Targeted error-corrected DNA sequencing was performed on 500ng of genomic DNA using a custom panel covering hotspot regions within the five genes of interest. Libraries were generated using an automated liquid handling workflow with pre-and post-PCR separation and subjected to sequencing using unique dual indices on an Illumina NovaSeq 6000. Further details on library preparation and variant detection are provided in the Supplemental Appendix.

### STATISTICAL ANALYSIS

Clinical data of the 1075 patients included age, sex, race, hematopoietic cell transplant specific comorbidity (HCT-CI), Karnofsky performance status (KPS), secondary AML, European LeukemiaNet (ELN) 2017 risk group, baseline genetics before transplant (*IDH1, IDH2, KIT, NPM1, FLT3-TKD, FLT3*-ITD), conditioning regimen, graft type, donor group, anti-thymocyte globulin (ATG) usage, and site reported MRD status. Day of transplant was considered as day 0, with the primary outcomes being overall survival (OS) and cumulative incidence of relapse (CIR) with non-relapse mortality (NRM) as a competing risk. Relapse-free survival (RFS) was the secondary outcome. Kaplan-Meier estimation and log rank tests were used to calculate OS and RFS endpoints; Fine-Gray regression models were used to examine CIR with NRM as a competing risk. Gene combinations for NGS-MRD were determined by the discovery cohort and validated in the second cohort. Cox proportional hazards regression models with forward selection or Lasso penalty were used to estimate the relative risks of clinical events and the proportional hazards assumptions were validated. Further details are provided in the in the Supplemental Appendix. The pre-specified statistical analysis plan was registered with OSF (https://osf.io/a6epx).

## Results

### PATIENTS

Among the 1075 patients in this analysis, 454 were transplanted between March 2013 and December 2017 (discovery cohort), and 621 were transplanted between January 2018 and February 2019 (validation cohort) (Table 1, Fig. S1. in Supplemental Appendix, available with the full text of this article at NEJM.org). The two cohorts were similar for most patient, leukemic, and transplant factors however in the later validation cohort mutations in *IDH* genes at AML diagnosis were more commonly reported and cord blood transplants were less likely to have been received.

**Table 1.**
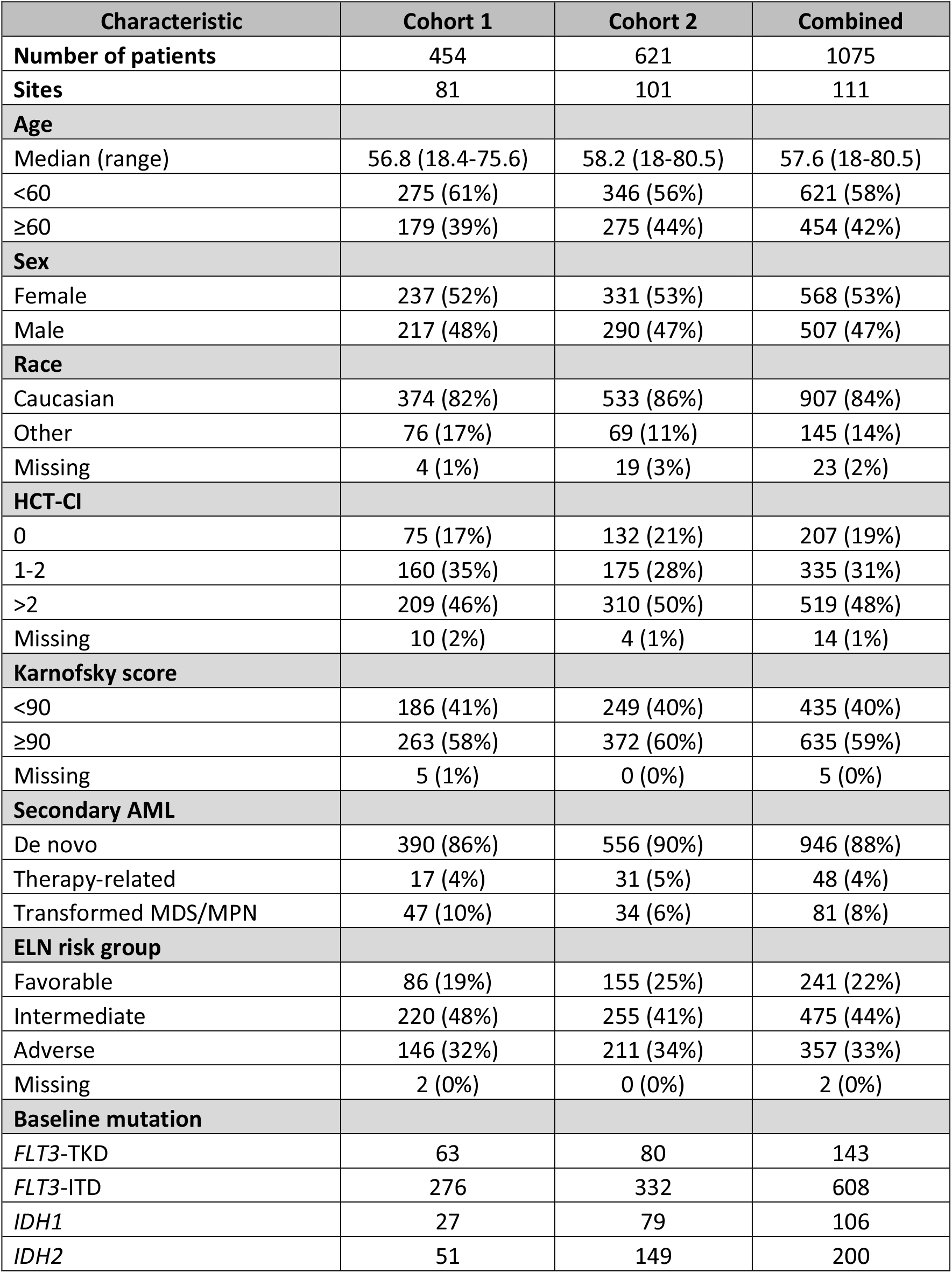
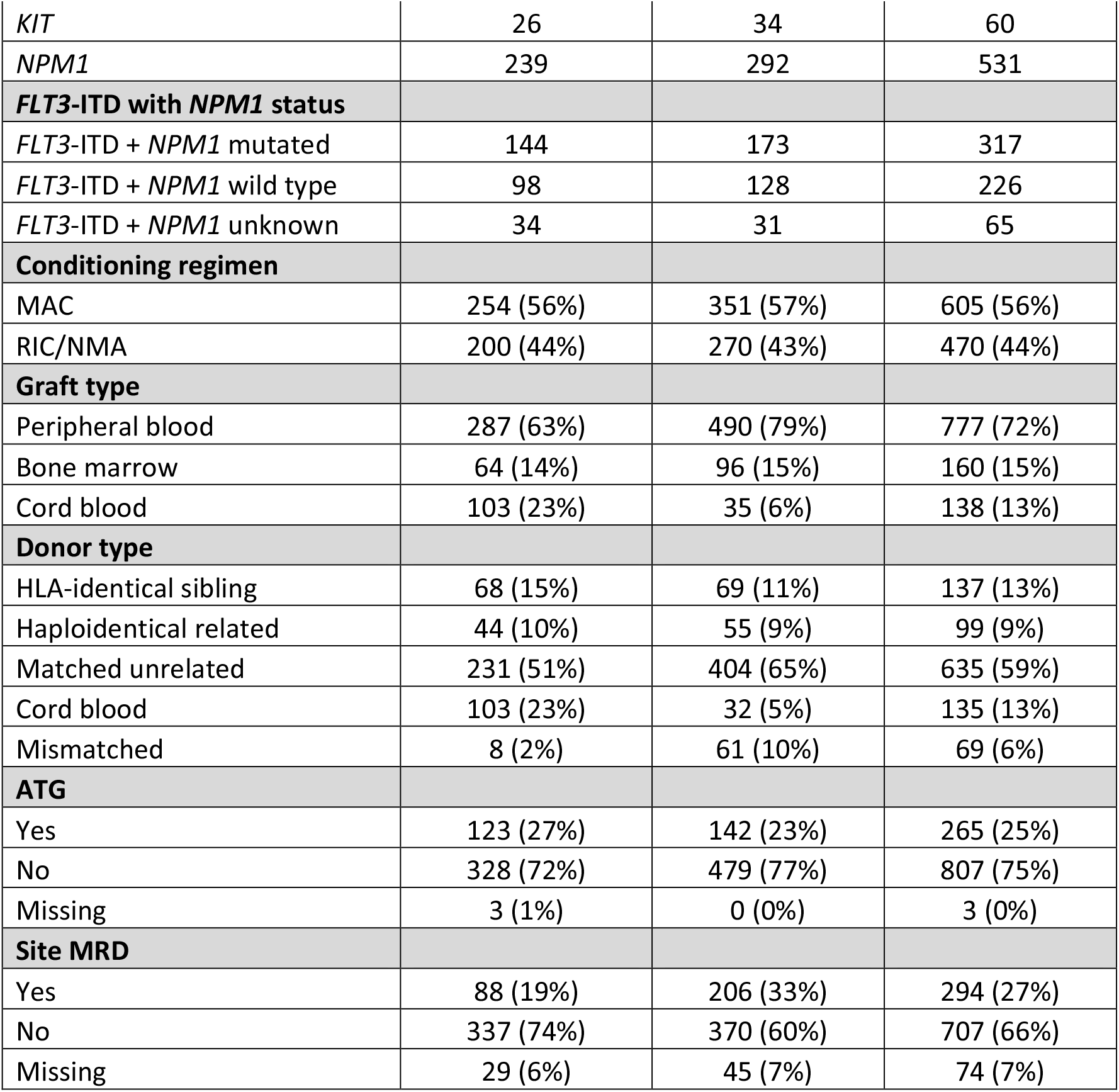
Patient Clinical Characteristics.

| Characteristic | Cohort 1 | Cohort 2 | Combined |
| --- | --- | --- | --- |
| **Number of patients** | 454 | 621 | 1075 |
| **Sites** | 81 | 101 | 111 |
| **Age** |  |  |  |
| Median (range) | 56.8 (18.4-75.6) | 58.2 (18-80.5) | 57.6 (18-80.5) |
| <60 | 275 (61%) | 346 (56%) | 621 (58%) |
| ≥60 | 179 (39%) | 275 (44%) | 454 (42%) |
| **Sex** |  |  |  |
| Female | 237 (52%) | 331 (53%) | 568 (53%) |
| Male | 217 (48%) | 290 (47%) | 507 (47%) |
| **Race** |  |  |  |
| Caucasian | 374 (82%) | 533 (86%) | 907 (84%) |
| Other | 76 (17%) | 69 (11%) | 145 (14%) |
| Missing | 4 (1%) | 19 (3%) | 23 (2%) |
| **HCT-CI** |  |  |  |
| 0 | 75 (17%) | 132 (21%) | 207 (19%) |
| 1-2 | 160 (35%) | 175 (28%) | 335 (31%) |
| >2 | 209 (46%) | 310 (50%) | 519 (48%) |
| Missing | 10 (2%) | 4 (1%) | 14 (1%) |
| **Karnofsky score** |  |  |  |
| <90 | 186 (41%) | 249 (40%) | 435 (40%) |
| ≥90 | 263 (58%) | 372 (60%) | 635 (59%) |
| Missing | 5 (1%) | 0 (0%) | 5 (0%) |
| **Secondary AML** |  |  |  |
| De novo | 390 (86%) | 556 (90%) | 946 (88%) |
| Therapy-related | 17 (4%) | 31 (5%) | 48 (4%) |
| Transformed MDS/MPN | 47 (10%) | 34 (6%) | 81 (8%) |
| **ELN risk group** |  |  |  |
| Favorable | 86 (19%) | 155 (25%) | 241 (22%) |
| Intermediate | 220 (48%) | 255 (41%) | 475 (44%) |
| Adverse | 146 (32%) | 211 (34%) | 357 (33%) |
| Missing | 2 (0%) | 0 (0%) | 2 (0%) |
| **Baseline mutation** |  |  |  |
| *FLT3*-TKD | 63 | 80 | 143 |
| *FLT3*-ITD | 276 | 332 | 608 |
| *IDH1* | 27 | 79 | 106 |
| *IDH2* | 51 | 149 | 200 |
| *KIT* | 26 | 34 | 60 |
| *NPM1* | 239 | 292 | 531 |
| ***FLT3*-ITD with *NPM1* status** |  |  |  |
| *FLT3*-ITD + *NPM1* mutated | 144 | 173 | 317 |
| *FLT3*-ITD + *NPM1* wild type | 98 | 128 | 226 |
| *FLT3*-ITD + *NPM1* unknown | 34 | 31 | 65 |
| **Conditioning regimen** |  |  |  |
| MAC | 254 (56%) | 351 (57%) | 605 (56%) |
| RIC/NMA | 200 (44%) | 270 (43%) | 470 (44%) |
| **Graft type** |  |  |  |
| Peripheral blood | 287 (63%) | 490 (79%) | 777 (72%) |
| Bone marrow | 64 (14%) | 96 (15%) | 160 (15%) |
| Cord blood | 103 (23%) | 35 (6%) | 138 (13%) |
| **Donor type** |  |  |  |
| HLA-identical sibling | 68 (15%) | 69 (11%) | 137 (13%) |
| Haploidentical related | 44 (10%) | 55 (9%) | 99 (9%) |
| Matched unrelated | 231 (51%) | 404 (65%) | 635 (59%) |
| Cord blood | 103 (23%) | 32 (5%) | 135 (13%) |
| Mismatched | 8 (2%) | 61 (10%) | 69 (6%) |
| **ATG** |  |  |  |
| Yes | 123 (27%) | 142 (23%) | 265 (25%) |
| No | 328 (72%) | 479 (77%) | 807 (75%) |
| Missing | 3 (1%) | 0 (0%) | 3 (0%) |
| **Site MRD** |  |  |  |
| Yes | 88 (19%) | 206 (33%) | 294 (27%) |
| No | 337 (74%) | 370 (60%) | 707 (66%) |
| Missing | 29 (6%) | 45 (7%) | 74 (7%) |

The relapse rate was 21% at 1 year in both cohorts. Patients with secondary AML, ELN adverse or intermediate risk, or receiving reduced intensity conditioning had higher relapse while age, sex, race, HCT-CI, KPS, graft source, donor type, and ATG usage did not stratify for relapse risk. Results of MRD testing performed at the local transplant centers did not correlate with overall survival (Fig. S2 in the Supplemental Appendix).

In the discovery cohort, 131 of 454 patients had a mutation in *FLT3, NPM1, IDH1, IDH2*, or *KIT* detected in CR1 blood by NGS-MRD. The total number of mutations detected in this first cohort was 177. In the validation cohort, 188 of 621 patient pre-alloHCT remission blood samples had a mutation in *FLT3, NPM1, IDH1, IDH2*, or *KIT* detected by NGS-MRD. The total number of mutations detected in this second cohort was 254. The most common mutations detected were in *IDH2, FLT3*-ITD, and *NPM1*. Detected variants were validated using digital droplet PCR (Fig. S3 in the Supplementary Appendix).

### NGS-MRD ASSOCIATED WITH RELAPSE AND SURVIVAL

Detection of a mutation in pre-alloHCT remission blood was associated with increased CIR, lower RFS and OS and was not associated with NRM (Fig. S4 in the Supplemental Appendix). Univariate cox regression analysis identified detection of *FLT3*-ITD (HR, 3.1; 95% CI: 2.1-4.8; P<0.0001) or mutated *NPM1* (NPM1m, HR, 4.5; 95% CI: 3.0-6.7, P<0.0001) as associated with increased relapse in the discovery cohort and was selected as the NGS-MRD definition (Fig. S5 in the Supplemental Appendix).

Detection of *NPM1*m and/or *FLT3*-ITD NGS-MRD in pre-alloHCT blood was able to stratify both cohorts, individually and combined, for CIR, RFS, and OS but not NRM (Fig. S6 in the Supplemental Appendix). In total 822 patients in the combined training and validation cohorts were known to have *NPM1*m and/or *FLT3*-ITD mutated AML and served as the clinically relevant subset for subsequent analysis on the impact of detecting persistence of these mutations in remission. The 142 of these patients (17.3%) testing positive for residual *NPM1m* and/or *FLT3*-ITD had NRM rates similar to those testing negative. However, detection of these mutations was associated with significantly higher 3-year relapse rate than no detection (62% vs. 23%; HR, 4.0; 95% CI, 3.1 to 5.2; P <0.0001), as well as with significantly lower 3-year rates of relapse-free survival (25% vs. 59%; HR for relapse or death, 2.8; 95% CI, 2.2 to 3.5; P <0.0001) and overall survival (36% vs. 66%; HR for death: 2.5; 95% CI, 2.0 to 3.2; P <0.0001) (Figure 1, Fig. S6 in the Supplemental Appendix).

**Figure 1.**
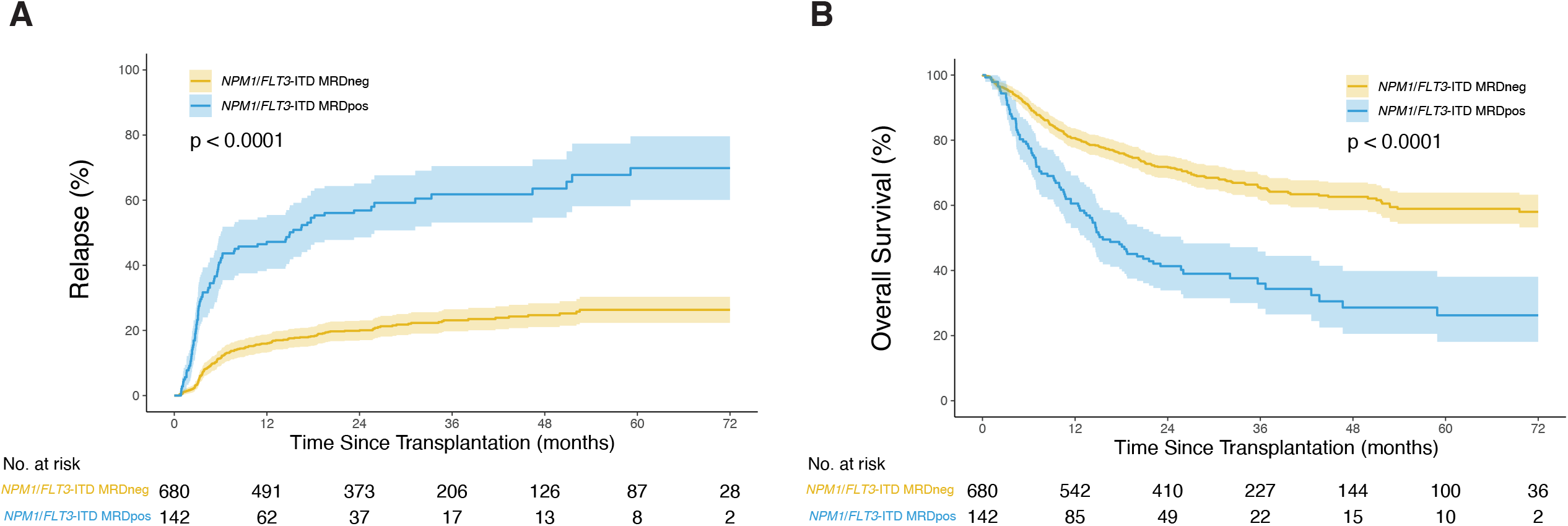
Impact of *NPM1* and *FLT3*-ITD NGS-MRD status on clinical outcomes. Patients with *NPM1* and/or *FLT3*-ITD mutated acute myeloid leukemia (n=822) with measurable residual disease (MRD) defined by persistent mutations detectable by next-generation sequencing (NGS) in remission blood prior to transplant (*NPM1*/*FLT3*-ITD MRDpos) had significantly (A) higher rates of relapse (3-year, *P* < 0.0001) and (B) decreased overall survival (3-year, *P* < 0.0001) after transplantation compared to patients with no mutations detected (*NPM1*/*FLT3*-ITD MRDneg). Data viewed at 72 months.

### NGS-MRD IN MOLECULAR SUBGROUPS

Patients in this study were often known to have multiple genes of interest mutated at the time of initial AML diagnosis (median: 1, range: 1-4) (Figure 2).

**Figure 2.**
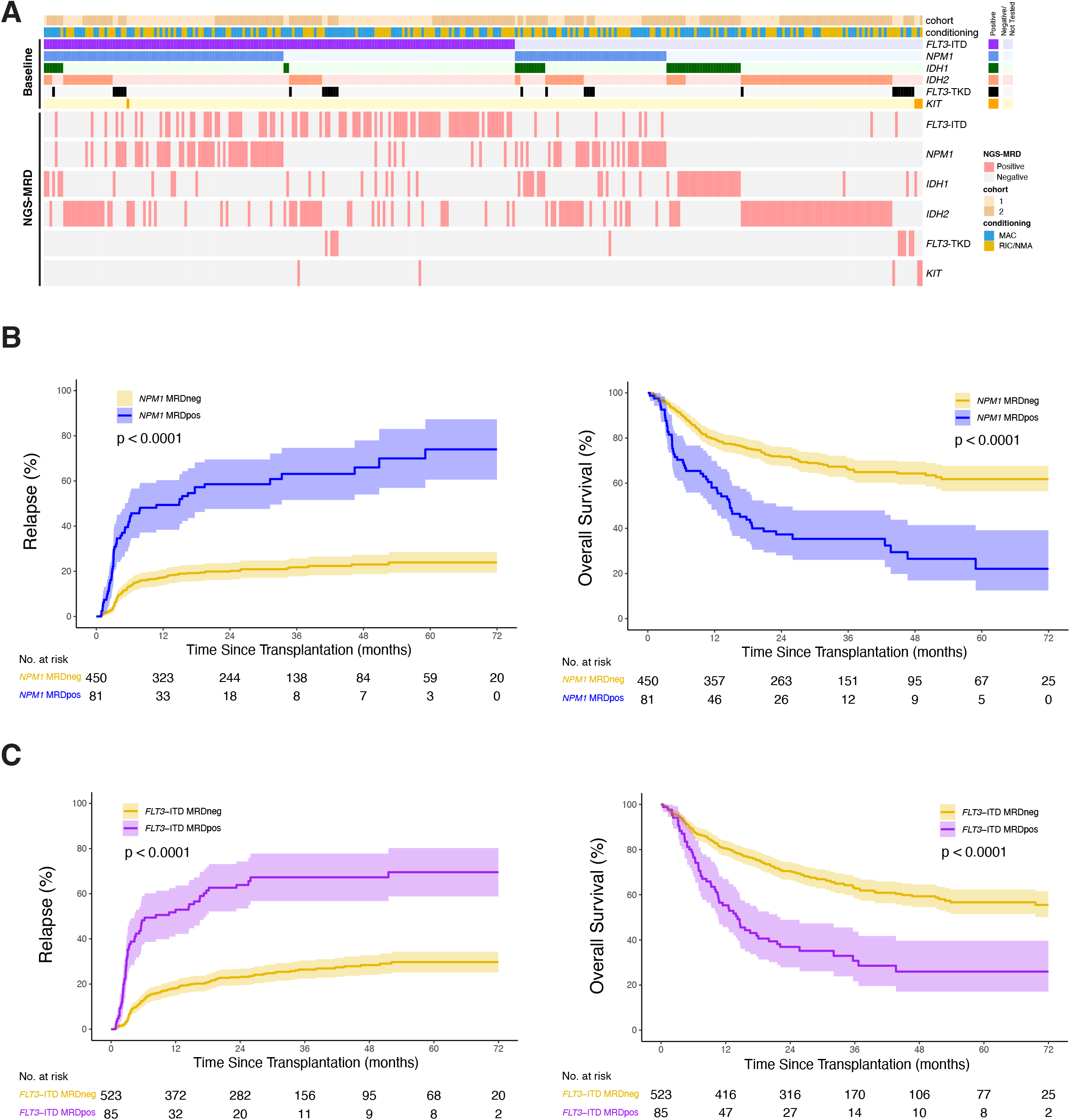
Detection and impact of residual mutations on clinical outcomes by gene. (A) Heatmap illustrating for each patient (n=1,075 patients) their cohort assignment, conditioning intensity, reported baseline mutation status, and residual mutation detection status by next generation sequencing (NGS-MRD) for each gene. Patients are sorted by the mutations reported at baseline (positive or negative/not tested). (B) Patients with *NPM1* mutated AML (n=531) with residual *NPM1* mutations detectable by NGS prior to transplant (*NPM1* MRDpos) had significantly higher rates of relapse (3-year, *P* < 0.0001) and decreased overall survival (3-year, *P* < 0.0001) compared to patients with no *NPM1* mutations detected (*NPM1* MRDneg). (C) Patients with *FLT3*-ITD mutated AML (n=608) with residual *FLT3*-ITD mutations detectable by NGS prior to transplant (*FLT3*-ITD MRDpos) had significantly higher rates of relapse (3-year, *P* < 0.0001) and decreased overall survival (3-year, *P* < 0.0001) compared to patients with no *FLT3*-ITD mutations detected (*FLT3*-ITD MRDneg). Data viewed at 72 months.

The 3-year relapse rate for all 531 patients with *NPM1*-mutated AML was 28%. Residual *NPM1*m in pre-transplant remission blood samples was detectable in 81 (15.3%) of these patients and associated with statistically significantly higher 3-year rates of relapse versus those without residual *NPM1*m detected (63% vs. 22%; HR, 4.5; 95% CI, 3.2 to 6.2; P <0.0001), as well as with significantly lower 3-year rates of relapse-free survival (23% vs. 61%; HR for relapse or death, 3.2; 95% CI, 2.4 to 4.2; P <0.0001) and overall survival (35% vs. 66%; HR for death, 2.9; 95% CI, 2.1 to 3.9; P <0.0001).

The 3-year relapse rate for all 608 patients with *FLT3*-ITD-mutated AML was 32%. Residual *FLT3*-ITD was detectable in pre-transplant remission blood samples from 85 (14%) of these patients and associated with significantly higher 3-year rates of relapse versus those without residual *FLT3*-ITD detected (67% vs. 26%; HR, 4.1; 95% CI, 3.0 to 5.6; P <0.0001), as well as with significantly lower 3-year rates of relapse-free survival (20% vs. 56%; HR, 2.9; 95% CI, 2.2 to 3.8; P <0.0001) and overall survival (31% vs. 63%; HR, 2.6; 95% CI, 1.9 to 3.5; P <0.0001).

NGS-MRD for *NPM1m* and *FLT3*-ITD outperformed reported local site MRD determinations for relapse and survival prognostication (Fig. S7 in the Supplemental Appendix) and had utility for relapse prognostication in both younger and older adults (Fig. S8 in the Supplemental Appendix).

For those patients with *NPM1* and *FLT3*-ITD co-mutated AML, detection in pre-transplant remission blood of *NPM1*m alone, *FLT3*-ITD alone, or both was associated with increased post-transplant relapse (Fig. S9 in the Supplemental Appendix).

### IMPACT OF CONDITIONING INTENSITY

There is strong evidence that patients who test MRD positive prior to alloHCT with reduced intensity conditioning (RIC) have high relapse and poor survival^38^ and that in younger adult patients, conditioning intensification may improve these outcomes^28,39^. In this study patients were not randomly assigned to conditioning intensities and treating physicians were aware of pre-transplantation patient and disease factors. NGS-MRD positivity was associated with increased relapse and worse survival (Figure 3A) that appeared partially mitigated in younger patients (<60 years) who received high intensity conditioning (3-year relapse rate 53% vs. 78%, HR for death, 2.0; 95% CI, 1.0 to 3.7; P=0.04) (Figure 3B, Fig. S10 in the Supplemental Appendix).

**Figure 3.**
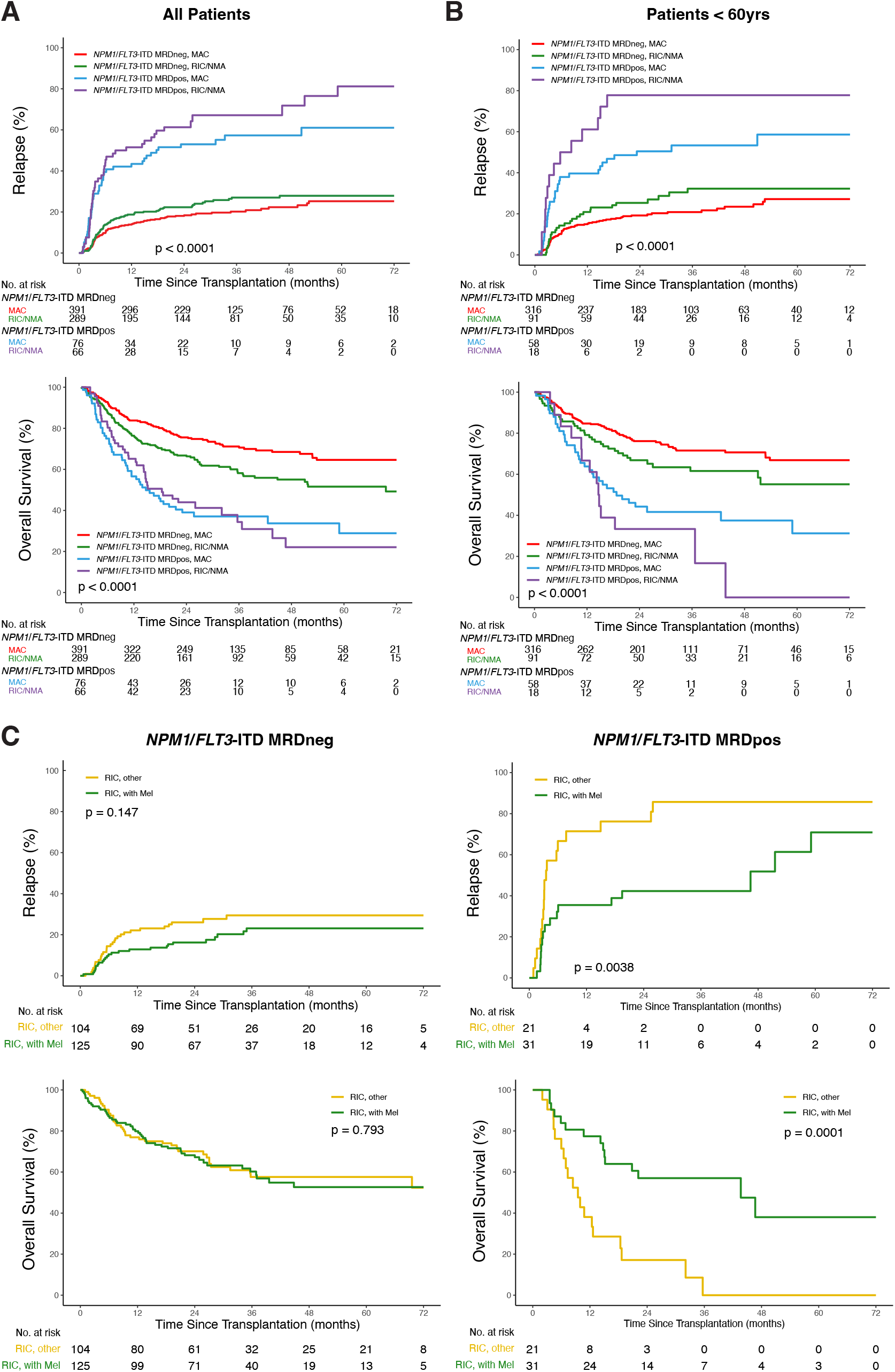
Impact of conditioning intensity and NGS-MRD status on clinical outcomes. (A) In patients with *NPM1* and/or *FLT3*-ITD mutated AML (n=822), differences in rates of relapse (*P* < 0.0001) and overall survival (*P* < 0.0001) were identified between subgroups defined by conditioning intensity and next-generation sequencing (NGS) based measurable residual disease (MRD) status. Increased rates of relapse and decreased overall survival were observed in patients with residual mutations detectable by NGS in remission blood prior to transplant (*NPM1*/*FLT3*-ITD MRDpos) compared to patients with no mutations detected (*NPM1*/*FLT3*-ITD MRDneg), regardless of conditioning intensity. (B) In patients with *NPM1* and/or *FLT3*-ITD mutated AML aged less than 60 years (n=483), the use of myeloablative (MAC) vs reduced intensity (RIC) or non-myeloablative (NMA) conditioning regimens resulted in significantly decreased rates of relapse (*P* = 0.04) within the *NPM1*/*FLT3*-ITD MRDpos group. (C) In patients with *NPM1* and/or *FLT3*-ITD mutated AML receiving RIC (n=281), the use of melphalan (Mel) resulted in significantly decreased rates of relapse (*P* = 0.00038) and increased overall survival (*P* = 0.0001) in the *NPM1*/*FLT3*-ITD MRDpos group (right) but not in the *NPM1*/*FLT3*-ITD MRDneg group (left).

Based on the premise that patients allocated to RIC would be more similar to each other than to those receiving myeloablative conditioning in this non-randomized study, we compared outcomes in patients receiving melphalan-containing RIC regimens to other forms of RIC (excluding non-myeloablative regimens, Table S1 in the Supplemental Appendix). There was no difference in relapse or survival in those who were NGS-MRD negative, but significantly lower relapse and improved survival in NGS-MRD positive patients receiving melphalan-based RIC (Figure 3C).

In multivariate analyses NGS-MRD status was the most important factor for both relapse and overall survival although receipt of RIC that did not contain melphalan was associated with a higher rate of relapse (Figure 4). In subgroups defined by baseline *NPM1* and *FLT3*-ITD mutational status NGS-MRD remained the most important factor regardless of variant allele fraction threshold selected. *NPM1* mutations or *FLT3*-ITD detected in remission using the current ELN recommended threshold of 0.1% VAF was associated with increased relapse and decreased survival, lowering this threshold ten-fold further to 0.01% VAF remained prognostic and doubled the number of high-risk NGS-MRD positive patients identified (Fig. S11 in the Supplemental Appendix). No difference in clinical outcomes were observed for those receiving post-transplant maintenance therapy, regardless of NGS-MRD status (Fig. S12 in the Supplemental Appendix).

**Figure 4.**
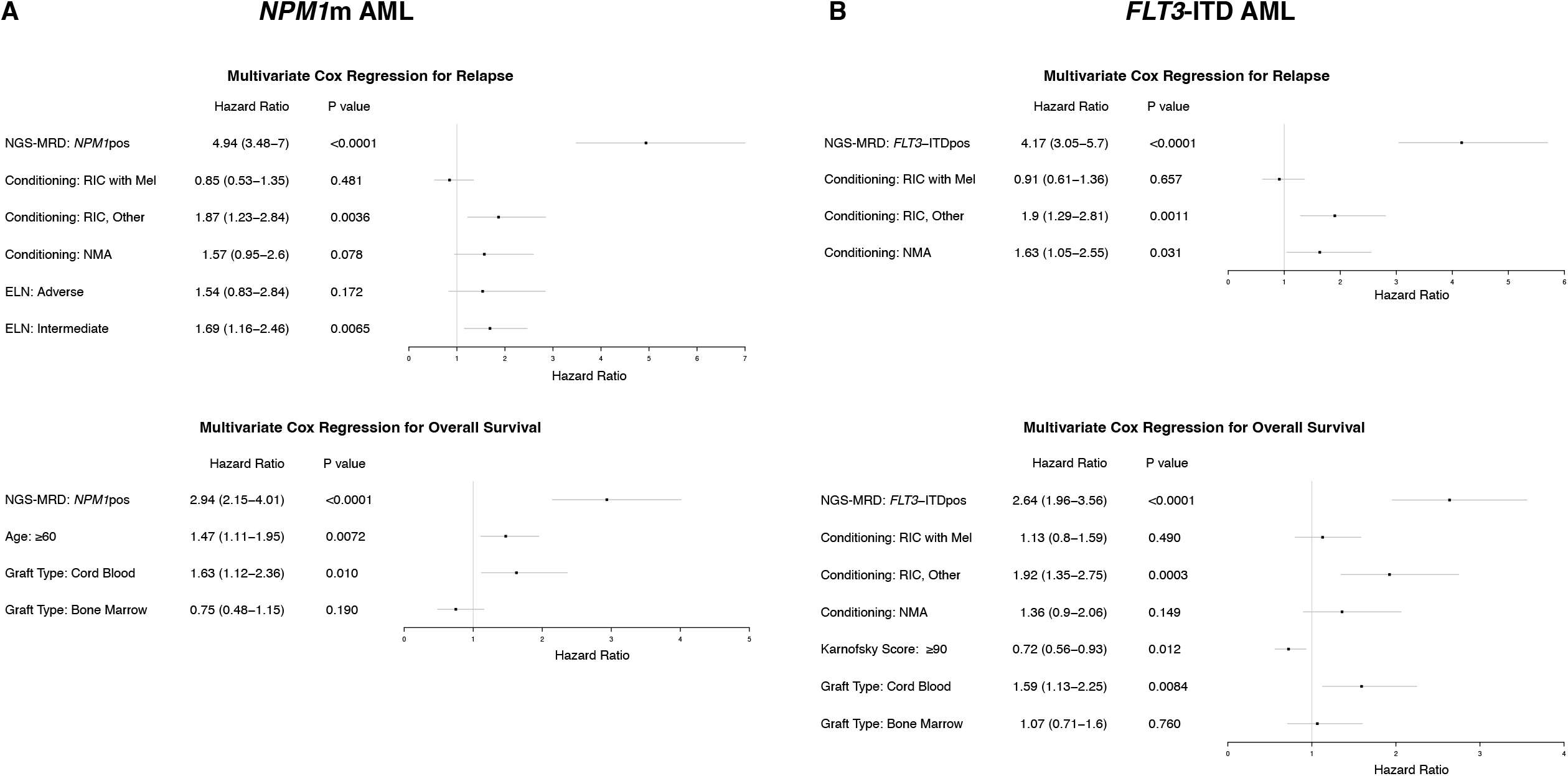
Multivariate Cox regression analysis. (A) Residual *NPM1*m in patients with *NPM1*-mutated AML (n=531). (B) Residual *FLT3*-ITD in patients with *FLT3*-ITD mutated AML (n=608). Models were selected stepwise with next-generation sequencing measurable residual disease (NGS-MRD) status and baseline characteristics including age, sex, race, hematopoietic cell transplant specific comorbidity index, Karnofsky performance status, secondary AML, ELN risk group, baseline genetics before transplant (*IDH1, IDH2, KIT, NPM1, FLT3-TKD, FLT3-*ITD), conditioning regimen, graft type, donor group, anti-thymocyte globulin usage.

## Discussion

A personalized medicine approach to the care of patients with AML would require not only comprehensive clinical and genetic profiling at initial diagnosis for a precisely optimized initial therapeutic approach, but also iterative re-assessment and therapy adjustment during and after planned treatment. The ability to detect residual leukemia in patients in an apparent “complete” remission after treatment using AML MRD would be a powerful tool, providing the measured result was interpretable in the context of large clinical datasets supporting both the prognostic implications and the clinical utility of any proposed intervention. Current evidence for AML MRD comes from individual centers or clinical trials with a consistent approach but unclear generalizability due to test inaccessibility, or large and highly representative registry studies of limited clinical applicability due to heterogenous testing methodologies^24,27,28,32,39^. Here we report the results of a large systematic study demonstrating that the detection by NGS of persistent *NPM1*m or *FLT3*-ITD mutations in the blood of adult patients with AML in CR1 before alloHCT is associated with statistically significant increased relapse and decreased survival than those testing negative.

A powerful motivating factor for the clinical adoption of DNA-sequencing for AML MRD is the ability to establish a single test with coverage of multiple potential mutations within targets; a prior study using quantitative polymerase chain reaction reported the need to establish 27 different mutation-specific assays for *NPM1*m AML MRD^40^. We show excellent correlation of DNA mutation quantification between NGS-MRD and ddPCR suggesting harmonized thresholds could be established. While prognostic models based on baseline characteristics can be developed to stratify entire cohorts of patients based on relapse or survival probabilities, we show here that NGS-MRD testing of blood in AML patients with *NPM1*m and/or *FLT3*-ITD AML at a key landmark during treatment can identify differential risk between individuals otherwise placed in the same baseline risk-classification.

Consistent with prior reports^28,39^, we observed a benefit for myeloablative over reduced intensity conditioning in younger patients with NGS-MRD prior to alloHCT (Figure 3B). However, many patients with AML are not able to tolerate myeloablative conditioning, limiting applicability. Attempts to augment RIC to reduce the risk of relapse, particularly in those testing MRD positive, have been unsuccessful^38^. Large registry-level retrospective analyses have reported that the use of melphalan containing RIC alloHCT regimens for AML is associated with lower relapse rates than busulfan-based regimens^41,42^. We did not find evidence of a difference in relapse for patients testing NGS-MRD negative prior to RIC alloHCT when comparing those transplanted with or without melphalan. In contrast, there was a statistically significant difference in relapse and survival for who those with persistently detectable *NPM1* or *FLT3*-ITD mutations based on the RIC regimen received (Figure 3C). This observation, which should be confirmed in a prospective randomized trial, could have important implications for alloHCT conditioning selection.

This study has limitations. It is unknown how results of NGS-MRD testing on bone marrow would differ from blood. It is also unknown if centralized flow cytometry MRD based testing would be concordant or complementary to NGS-MRD. While MRD results reported by local transplant sites to CIBMTR were not associated with overall survival our study did not have access to details of the testing performed; this question is now being examined by an ongoing CIBMTR initiative. Due to strict pre-specified criteria four potential NGS-MRD targets were not selected for validation based on results of our discovery cohort, future work will explore the prognostic implications of detecting these mutations. Only a minority (10%) of patients were reported as having received post-transplant maintenance therapy, while there is evidence this approach may reduce relapse including in those MRD positive after transplant^43,44^, no benefit of post-transplant maintenance therapy was observed for any subset in our study. Randomized phase 3 clinical trial evidence for the efficacy of FLT3-inhibitor post-alloHCT maintenance is forthcoming (NCT02997202). While this study provides evidence in support of NGS-MRD based assessment of blood in patients with AML with *NPM1*m and/or *FLT3*-ITD prior to alloHCT, it is unknown how serial testing after transplantation would further improve the performance characteristics of NGS-MRD testing. This question will be addressed by a prospective national clinical protocol, MEASURE (NCT05224661).

The European Leukemia Network guidelines have recognized, since 2017, that MRD negativity in CR represents a higher category of response after treatment for AML than CR alone^26^. The U.S. Food and Drug Administration has already accepted that achievement of MRD levels of less than 0.01% represents important evidence of drug efficacy in patients with acute leukemia^45^. This study confirms that standard, using blood-based NGS-MRD testing, for the specific situation of adult patients with *NPM1* and/or *FLT3*-ITD mutated AML in CR1 prior to alloHCT. We show that approximately 1 in 6 of such patients are in a high-risk subgroup with MRD-detectable above this threshold, with serious post-transplantation outcomes not adequately addressed by the current clinical standard of care. Adults with persistence of *NPM1* and/or *FLT3*-ITD mutations in cytomorphological remission after initial treatment for AML therefore represent patients with unmet medical need, who should be offered enrolment in a therapeutic clinical trial wherever possible.

In conclusion, detection of residual *NPM1* and/or *FLT3*-ITD mutations in the blood of patients with AML in CR1 prior to alloHCT is associated with increased relapse and worse survival compared with those testing negative. Myeloablative and melphalan-containing reduced-intensity conditioning regimens may partially mitigate this risk, but novel therapeutic approaches are needed. This study establishes the clinical utility of NGS-MRD testing of blood to identify a high-risk group of patients with AML in CR1 prior to alloHCT who have a clear unmet medical need.

## Data Availability

All data produced in the present work are contained in the manuscript or available online. FASTQ files are available in the NCBI Small Reads Archive (SRA) (Accession: PRJNA834073). The pre-specified statistical analysis plan was registered with OSF: Gui, G., Dillon, L., & Hourigan, C. (2022, January 7). Allogeneic Transplantation for Acute Myeloid Leukemia with Genomic Evidence of Residual Disease (https://osf.io/a6epx).

## Acknowledgements

This work was supported in part by the Intramural Research Program of the National Heart, Lung, and Blood Institute and by the National Institutes of Health Director’s Challenge Innovation Award.

Sequencing was performed in the NHLBI Intramural DNA Sequencing and Genomics Core. Digital droplet PCR was performed in the NCI Intramural CCR Genomics Core.

The National Heart, Lung, and Blood Institute receives research funding for the laboratory of Dr. Hourigan from Sellas and from the Foundation of the NIH AML MRD Biomarkers Consortium.

The CIBMTR is supported primarily by Public Health Service U24CA076518 from the National Cancer Institute (NCI), the National Heart, Lung and Blood Institute (NHLBI) and the National Institute of Allergy and Infectious Diseases (NIAID); U24HL138660 and U24HL157560 from NHLBI and NCI; U24CA233032 from the NCI; OT3HL147741 and U01HL128568 from the NHLBI; HHSH250201700005C, HHSH250201700006C, and HHSH250201700007C from the Health Resources and Services Administration (HRSA); and N00014-20-1-2832 and N00014-21-1-2954 from the Office of Naval Research.

The views expressed in this article do not reflect the official policy or position of the National Institute of Health, the Department of the Navy, the Department of Defense, Health Resources and Services Administration (HRSA) or any other agency of the U.S. Government.

## Conflict of Interest Statements

Hourigan: The National Heart, Lung, and Blood Institute receives research funding for the laboratory of Dr. Hourigan from Sellas and from the Foundation of the NIH AML MRD Biomarkers Consortium.

De Lima: Consultant for Pfizer, BMS, Research collaboration with Miltenyi Biotec

Devine: Full time employee of NMDP. Advisor to Sanofi, Magenta, Orca Bio Jimenez

Jimenez: Abbvie

Weisdorf: Research support from FATE therapeutics (unrelated). Previous research support from Incyte (unrelated).

All other authors report no potential conflict of interest.

## Notes

### Author Declarations

Patients provided written informed consent to participate in the Center for International Blood and Marrow Transplant Research research protocol, which was approved by the National Marrow Donor Program institutional review board.

## References

1. Döhner H, Weisdorf DJ, Bloomfield CD. Acute Myeloid Leukemia. N Engl J Med 2015;373(12):1136-52. (In eng). DOI: 10.1056/NEJMra1406184.

2. Ley TJ, Miller C, Ding L, et al. Genomic and epigenomic landscapes of adult de novo acute myeloid leukemia. N Engl J Med 2013;368(22):2059-74. (In eng). DOI: 10.1056/NEJMoa1301689.

3. Papaemmanuil E, Gerstung M, Bullinger L, et al. Genomic Classification and Prognosis in Acute Myeloid Leukemia. N Engl J Med 2016;374(23):2209-2221. (In eng). DOI: 10.1056/NEJMoa1516192.

4. Tyner JW, Tognon CE, Bottomly D, et al. Functional genomic landscape of acute myeloid leukaemia. Nature 2018;562(7728):526-531. (In eng). DOI: 10.1038/s41586-018-0623-z.

5. Bottomly D, Long N, Schultz AR, et al. Integrative analysis of drug response and clinical outcome in acute myeloid leukemia. Cancer Cell 2022;40(8):850–864 e9. DOI: 10.1016/j.ccell.2022.07.002.

6. Patel JP, Gönen M, Figueroa ME, et al. Prognostic relevance of integrated genetic profiling in acute myeloid leukemia. N Engl J Med 2012;366(12):1079-89. (In eng). DOI: 10.1056/NEJMoa1112304.

7. Schlenk RF, Döhner K, Krauter J, et al. Mutations and treatment outcome in cytogenetically normal acute myeloid leukemia. N Engl J Med 2008;358(18):1909-18. (In eng). DOI: 10.1056/NEJMoa074306.

8. Duncavage EJ, Schroeder MC, O’Laughlin M, et al. Genome Sequencing as an Alternative to Cytogenetic Analysis in Myeloid Cancers. N Engl J Med 2021;384(10):924-935. (In eng). DOI: 10.1056/NEJMoa2024534.

9. Stone RM, Mandrekar SJ, Sanford BL, et al. Midostaurin plus Chemotherapy for Acute Myeloid Leukemia with a FLT3 Mutation. N Engl J Med 2017;377(5):454-464. (In eng). DOI: 10.1056/NEJMoa1614359.

10. DiNardo CD, Stein EM, de Botton S, et al. Durable Remissions with Ivosidenib in IDH1-Mutated Relapsed or Refractory AML. N Engl J Med 2018;378(25):2386-2398. (In eng). DOI: 10.1056/NEJMoa1716984.

11. Perl AE, Martinelli G, Cortes JE, et al. Gilteritinib or Chemotherapy for Relapsed or Refractory FLT3-Mutated AML. N Engl J Med 2019;381(18):1728-1740. (In eng). DOI: 10.1056/NEJMoa1902688.

12. Montesinos P, Recher C, Vives S, et al. Ivosidenib and Azacitidine in IDH1-Mutated Acute Myeloid Leukemia. N Engl J Med 2022;386(16):1519-1531. (In eng). DOI: 10.1056/NEJMoa2117344.

13. Tazi Y, Arango-Ossa JE, Zhou Y, et al. Unified classification and risk-stratification in Acute Myeloid Leukemia. Nat Commun 2022;13(1):4622. (In eng). DOI: 10.1038/s41467-022-32103-8.

14. Döhner H, Wei AH, Appelbaum FR, et al. Diagnosis and Management of AML in Adults: 2022 ELN Recommendations from an International Expert Panel. Blood 2022 (In eng). DOI: 10.1182/blood.2022016867.

15. Koreth J, Schlenk R, Kopecky KJ, et al. Allogeneic stem cell transplantation for acute myeloid leukemia in first complete remission: systematic review and meta-analysis of prospective clinical trials. Jama 2009;301(22):2349-61. (In eng). DOI: 10.1001/jama.2009.813.

16. Cornelissen JJ, Versluis J, Passweg JR, et al. Comparative therapeutic value of post-remission approaches in patients with acute myeloid leukemia aged 40-60 years. Leukemia 2015;29(5):1041-50. (In eng). DOI: 10.1038/leu.2014.332.

17. Pollyea DA, Bixby D, Perl A, et al. NCCN Guidelines Insights: Acute Myeloid Leukemia, Version 2.2021. J Natl Compr Canc Netw 2021;19(1):16-27. (In eng). DOI: 10.6004/jnccn.2021.0002.

18. Scott BL, Pasquini MC, Logan BR, et al. Myeloablative Versus Reduced-Intensity Hematopoietic Cell Transplantation for Acute Myeloid Leukemia and Myelodysplastic Syndromes. J Clin Oncol 2017;35(11):1154-1161. (In eng). DOI: 10.1200/jco.2016.70.7091.

19. Milano F, Gooley T, Wood B, et al. Cord-Blood Transplantation in Patients with Minimal Residual Disease. N Engl J Med 2016;375(10):944-53. (In eng). DOI: 10.1056/NEJMoa1602074.

20. Venstrom JM, Pittari G, Gooley TA, et al. HLA-C-dependent prevention of leukemia relapse by donor activating KIR2DS1. N Engl J Med 2012;367(9):805-16. (In eng). DOI: 10.1056/NEJMoa1200503.

21. Christopher MJ, Petti AA, Rettig MP, et al. Immune Escape of Relapsed AML Cells after Allogeneic Transplantation. N Engl J Med 2018;379(24):2330-2341. (In eng). DOI: 10.1056/NEJMoa1808777.

22. Vago L, Perna SK, Zanussi M, et al. Loss of mismatched HLA in leukemia after stem-cell transplantation. N Engl J Med 2009;361(5):478-88. (In eng). DOI: 10.1056/NEJMoa0811036.

23. Toffalori C, Zito L, Gambacorta V, et al. Immune signature drives leukemia escape and relapse after hematopoietic cell transplantation. Nat Med 2019;25(4):603-611. (In eng). DOI: 10.1038/s41591-019-0400-z.

24. Buckley SA, Wood BL, Othus M, et al. Minimal residual disease prior to allogeneic hematopoietic cell transplantation in acute myeloid leukemia: a meta-analysis. Haematologica 2017;102(5):865-873. (In eng). DOI: 10.3324/haematol.2016.159343.

25. Thol F, Gabdoulline R, Liebich A, et al. Measurable residual disease monitoring by NGS before allogeneic hematopoietic cell transplantation in AML. Blood 2018;132(16):1703-1713. (In eng). DOI: 10.1182/blood-2018-02-829911.

26. Döhner H, Estey E, Grimwade D, et al. Diagnosis and management of AML in adults: 2017 ELN recommendations from an international expert panel. Blood 2017;129(4):424-447. (In eng). DOI: 10.1182/blood-2016-08-733196.

27. Heuser M, Freeman SD, Ossenkoppele GJ, et al. 2021 Update Measurable Residual Disease in Acute Myeloid Leukemia: European LeukemiaNet Working Party Consensus Document. Blood 2021. DOI: 10.1182/blood.2021013626.

28. Hourigan CS, Dillon LW, Gui G, et al. Impact of Conditioning Intensity of Allogeneic Transplantation for Acute Myeloid Leukemia With Genomic Evidence of Residual Disease. J Clin Oncol 2020;38(12):1273-1283. (In eng). DOI: 10.1200/jco.19.03011.

29. Araki D, Wood BL, Othus M, et al. Allogeneic Hematopoietic Cell Transplantation for Acute Myeloid Leukemia: Time to Move Toward a Minimal Residual Disease-Based Definition of Complete Remission? J Clin Oncol 2016;34(4):329–36. (In eng). DOI: 10.1200/jco.2015.63.3826.

30. Hourigan CS, Gale RP, Gormley NJ, Ossenkoppele GJ, Walter RB. Measurable residual disease testing in acute myeloid leukaemia. Leukemia 2017;31(7):1482–1490. DOI: 10.1038/leu.2017.113.

31. Jongen-Lavrencic M, Grob T, Hanekamp D, et al. Molecular Minimal Residual Disease in Acute Myeloid Leukemia. N Engl J Med 2018;378(13):1189-1199. (In eng). DOI: 10.1056/NEJMoa1716863.

32. Short NJ, Zhou S, Fu C, et al. Association of Measurable Residual Disease With Survival Outcomes in Patients With Acute Myeloid Leukemia: A Systematic Review and Meta-analysis. JAMA Oncol 2020;6(12):1890-9. (In eng). DOI: 10.1001/jamaoncol.2020.4600.

33. Levis MJ, Perl AE, Altman JK, et al. A next-generation sequencing-based assay for minimal residual disease assessment in AML patients with FLT3-ITD mutations. Blood Adv 2018;2(8):825-831. (In eng). DOI: 10.1182/bloodadvances.2018015925.

34. Dillon R, Hills R, Freeman S, et al. Molecular MRD status and outcome after transplantation in NPM1-mutated AML. Blood 2020;135(9):680-688. (In eng). DOI: 10.1182/blood.2019002959.

35. Liang EC, Chen C, Lu R, Mannis GN, Muffly L. Measurable residual disease status and FLT3 inhibitor therapy in patients with FLT3-ITD mutated AML following allogeneic hematopoietic cell transplantation. Bone Marrow Transplant 2021;56(12):3091-3093. (In eng). DOI: 10.1038/s41409-021-01475-8.

36. Loo S, Dillon R, Ivey A, et al. Pre-transplant FLT3-ITD MRD assessed by high-sensitivity PCR-NGS determines post-transplant clinical outcome. Blood 2022 (In eng). DOI: 10.1182/blood.2022016567.

37. Murdock HM, Kim HT, Denlinger N, et al. Impact of diagnostic genetics on remission MRD and transplantation outcomes in older patients with AML. Blood 2022;139(24):3546-3557. (In eng). DOI: 10.1182/blood.2021014520.

38. Craddock C, Jackson A, Loke J, et al. Augmented Reduced-Intensity Regimen Does Not Improve Postallogeneic Transplant Outcomes in Acute Myeloid Leukemia. J Clin Oncol 2020:Jco2002308. (In eng). DOI: 10.1200/jco.20.02308.

39. Gilleece MH, Labopin M, Yakoub-Agha I, et al. Measurable residual disease, conditioning regimen intensity, and age predict outcome of allogeneic hematopoietic cell transplantation for acute myeloid leukemia in first remission: A registry analysis of 2292 patients by the Acute Leukemia Working Party European Society of Blood and Marrow Transplantation. Am J Hematol 2018;93(9):1142–1152. DOI: 10.1002/ajh.25211.

40. Ivey A, Hills RK, Simpson MA, et al. Assessment of Minimal Residual Disease in Standard-Risk AML. N Engl J Med 2016;374(5):422-33. (In eng). DOI: 10.1056/NEJMoa1507471.

41. Zhou Z, Nath R, Cerny J, et al. Reduced intensity conditioning for acute myeloid leukemia using melphalan-vs busulfan-based regimens: a CIBMTR report. Blood Adv 2020;4(13):3180-3190. (In eng). DOI: 10.1182/bloodadvances.2019001266.

42. Baron F, Labopin M, Peniket A, et al. Reduced-intensity conditioning with fludarabine and busulfan versus fludarabine and melphalan for patients with acute myeloid leukemia: a report from the Acute Leukemia Working Party of the European Group for Blood and Marrow Transplantation. Cancer 2015;121(7):1048-55. (In eng). DOI: 10.1002/cncr.29163.

43. Burchert A, Bug G, Fritz LV, et al. Sorafenib Maintenance After Allogeneic Hematopoietic Stem Cell Transplantation for Acute Myeloid Leukemia With FLT3–Internal Tandem Duplication Mutation (SORMAIN). Journal of Clinical Oncology 2020;38(26):2993–3002. DOI: 10.1200/jco.19.03345.

44. Platzbecker U, Middeke JM, Sockel K, et al. Measurable residual disease-guided treatment with azacitidine to prevent haematological relapse in patients with myelodysplastic syndrome and acute myeloid leukaemia (RELAZA2): an open-label, multicentre, phase 2 trial. Lancet Oncol 2018;19(12):1668-1679. (In eng). DOI: 10.1016/s1470-2045(18)30580-1.

45. Food and Drug Administration. Acute Myeloid Leukemia: Developing Drugs and Biological Products for Treatment. In: U.S. Department of Health and Human Services, ed. https://www.fda.gov/regulatory-information/search-fda-guidance-documents/acute-myeloid-leukemia-developing-drugs-and-biological-products-treatment 2020.

