## Supplementary Appendix for "Pre-MEASURE: *FLT3*-ITD and mutated *NPM1* measurable residual disease before allogeneic transplant in adults with AML in first remission"

#### Table of Contents


| | |
|---|---|
| Supplementary methods | Page 2 |
| Figure S1. CONSORT diagram. | Page 10 |
| Figure S2. Association of baseline patient characteristics and site MRD on clinical outcomes | Page 11 |
| Figure S3. Detection of residual mutations in the blood of AML patients during complete remission. | Page 16 |
| Figure S4. Association of pre-transplant residual mutations on clinical outcomes. | Page 19 |
| Figure S5. Univariate cox regression for relapse. | Page 20 |
| Figure S6. Association of NGS-MRD status for *NPM1* and/or *FLT3*-ITD on clinical outcomes. | Page 23 |
| Figure S7. Association of site MRD and NGS-MRD status on clinical outcomes for patients with baseline *FLT3*-ITD and/or *NPM1* mutations. | Page 25 |
| Figure S8. Association of age and NGS-MRD status for *NPM1* and/or *FLT3*-ITD on clinical outcomes. | Page 28 |
| Figure S9. Association of persistent mutations by gene on clinical outcomes. | Page 29 |
| Figure S10. Association of conditioning intensity, age, and NGS-MRD status for *NPM1* and/or *FLT3*-ITD on clinical outcomes. | Page 31 |
| Figure S11. Association of *NPM1* and/or *FLT3*-ITD mutational status and NGS-MRD VAF on clinical outcomes. | Page 32 |
| Figure S12. Association of post-transplant maintenance therapy and NGS-MRD status on clinical outcomes. | Page 36 |
| Table S1. Regions of interest in targeted DNA-sequencing panel. | Page 38 |
| Table S2. Sample sequencing summary (n = 1,075). | Page 39 |
| Table S3. Variants detected by next-generation sequencing in the blood of AML patients prior to conditioning | Page 40 |

#### SUPPLEMENTARY METHODS

##### Samples and DNA isolation

Post-treatment, pre-conditioning baseline whole blood samples were collected a median of 9 days (range 0-81) prior to the date of transplantation, with 95% collected within 40 days of transplant. High quality genomic DNA (gDNA) was extracted from 1,098 available frozen 1mL pre-conditioning whole blood aliquots (cohort #1 n=468, cohort #2 n=630) using the chemagic DNA Blood 2k kit H24 (cat# CMG-1097, PerkinElmer Health Sciences, Inc., Shelton, CT) on the chemagic360 instrument (PerkinElmer Health Sciences, Inc., Shelton, CT). DNA concentration was measured using the Quant-iT PicoGreen dsDNA Assay kit (cat# P11496, Thermo Fisher Scientific, Waltham, MA) on the Synergy LX microplate reader (BioTek Instruments, Inc., Winooski, VT). A median of 15.7-μg of gDNA (range 0.21-94.1-μg) was isolated from pre-conditioning whole blood and patients with at least 1-μg available were carried forward for analysis (n=1,075).

##### Targeted next-generation sequencing (NGS)

A custom anchored multiplex PCR based targeted DNA sequencing panel (VariantPlex, ArcherDx, Boulder, CO) was designed with coverage of recurrent hotspot mutation regions of *FLT3*, *IDH1*, *IDH2*, *KIT* and *NPM1* (**Table S1**).

Libraries were prepared according to manufacturer's instructions, with modifications, utilizing pre- and post-PCR separation on liquid handling robots. 500-ng of gDNA isolated from whole blood was brought to 50-μL using 10mM Tris-HCl pH 8.0 and was subjected to DNA fragmentation, end repair, A-tailing, purification using SPRIselect reagent (cat# B23318, Beckman Coulter, Inc., Brea, CA), and ligation with a universal ArcherDx molecular barcode (MBC) adapter, which tags each DNA molecule with a unique molecular index (UMI) and allows for unidirectional amplification of the sample using gene-specific primers. Following molecular barcode ligation, the libraries were subjected to two rounds of nested PCR for target enrichment. DNA fragmentation, end repair, cleanup after end repair, ligation step 1, cleanup after ligation step 1, MBC adapter incorporation, ligation step 2, cleanup after ligation step 2, and setup of first PCR were all performed on a Sciclone G3 NGS Workstation (PerkinElmer Health Sciences, Inc., Shelton, CT). Cleanup after first PCR, second PCR, and cleanup after second PCR were all performed on a Zephyr G3 NGS Workstation (PerkinElmer Health Sciences, Inc., Shelton, CT). Incubations and PCR reactions were performed on a

Mastercycler X50a (Eppendorf AG, Hamburg, Germany). For the first PCR, amplification was performed as follows: 95°C for 3 minutes; 16 cycles of 95°C for 30 seconds, 65°C for 5 minutes; 72°C for 3 minutes. For the second PCR, amplification was performed as follows: 95°C for 3 minutes; 22 cycles of 95°C for 30 seconds, 65°C for 5 minutes; 72°C for 3 minutes.

The resulting libraries were subjected to paired-end 150-bp sequencing on a NovaSeq 6000 (Illumina, San Diego, CA), according to manufacturer's instructions. Libraries were pooled for sequencing such that each sample had a unique dual index. A total of 15.7 billion read pairs were acquired across the 1,075 AML pre-conditioning patient samples, with an average of 12 million read pairs per sample (**Table S2**). FASTQ files are available in the NCBI Small Reads Archive (SRA) (Accession: PRJNA834073).

**Next generation sequencing data analysis**

Raw sequencing FASTQ files were analyzed using the Archer Analysis software version 6.2.7, using default settings, except for the deep shallow threshold set to 3 (defining a deep error-correctable bin as ≥3 reads per molecular barcode/UMI). DNA structural variation and SNP-InDel pipelines were utilized.

_Preprocessing_

Pre-processing steps include read cleaning, deduplication and error correction, read mapping, *de novo* and targeted variant calling, and variant annotation. During read cleaning, all reads were filtered based on Illumina's base quality calls and presence of the appropriate Archer chemistry read signatures. Additionally, pairs of reads were aligned to determine overlap between R1 and R2. If overlap occurred, any discordant bases between R1 and R2 were converted to N's. Next, de-duplication and error correction of PCR duplicates was performed by extracting UMI read families and generating a single high-fidelity consensus read for use in downstream analysis. Any singleton reads were used as is, but not considered a deep bin (≥3 reads per molecular barcode). All reads (consensus and singleton) were then mapped to the human genome version hg19 (build GRCh37) using a combination of Bowtie2 and BWA-Mem. For the 1,075 pre-conditioning libraries, there was an average of 12 million mapped read pairs with an average on-target deduplication ratio of 6.6 (**Table S2**). Across all gene-specific primers (GSP2) in the panel, there was a mean de-duplicated depth of 22,253 (**Table S2**).

Variant calling was then performed on the aligned reads using *de novo* and targeted variant calling algorithms. *De novo* variant calling was performed using both Freebayes and LoFreq algorithms, with a minimum allele frequency for variant calling set to 0.0001. To improve coverage of hotspot mutations, targeted variant calling was performed by providing a VCF file with recurrent variants to guarantee metrics were computed and supplied as an output in the bioinformatic pipeline. Annotation of variants was performed using the Variant Effect Predictor (VEP) algorithm.

*FLT3*-ITD mutations were detected using a structural variation algorithm. Beginning with deduplicated reads, clustering was performed for each gene specific primer, beginning with R2 reads and then assembled with their companion R1 reads using Velvet, creating a set of contigs for each R2/R1 cluster. Contigs were grouped together by target gene and merged based on pairwise local alignment, repeating until no further merges could take place. Contigs with low coverage, short length, or alignment to known transcripts or the reference genome were removed. The remaining sequences were annotated using NCBI blastn and those contigs sharing a breakpoint were merged. For each structural variant remaining, reads with coverage spanning the primary breakpoint by at least 20 bp were used to calculate the frequency of the mutation.

<u>*Error modeling*</u>

Following pre-processing, an outlier detection algorithm was performed to determine positions that have more alternate observations than would be expected due to error. In the outlier detection algorithm, a position-specific background error model based on a binomial distribution was generated for all variants called in a sample utilizing consensus reads that originated from multiple PCR duplicates (i.e. deep bins with ≥3 reads per molecular barcode/UMI). This background error model, called the Normal Data Set (ND), was generated using a set of 5 wild-type libraries made from Genome in a Bottle gDNA controls (Coriell Institute for Medical Research, Camden, NJ). For each variant called in a sample, a *P* value was calculated based on the ND background error model. Replicate control DNA libraries were included during library preparation to ensure continued validity of the ND background error model against batch effects, reagent lot variability, and sequencing run differences.

<u>*Variant filtering*</u>

***IDH1, IDH2, KIT, FLT3-TKD* variants**

Variants in *IDH1*, *IDH2*, *KIT*, and *FLT3* tyrosine kinase domain (TKD) called by the *de novo* (LoFreq and Freebayes) and targeted (Vision) variant algorithms underwent a first round of filtering to: 1) remove anomalies generated as the result of library preparation and sequencing; and 2) select for variants associated with leukemic progression. The following filter conditions were applied in step 1:

1. Deep Alternate Observations (DAO) ≥ 9
   a. The number of consensus reads that come from deep molecular families must be greater than or equal to 9. A molecular family is considered deep if there are greater than or equal to 3 reads per UMI.
2. Unique Start for Alternate Observation (UAO) ≥ 5
   a. The total number of unique starts across all reads that contained the alternate observations must be greater than or equal to 5.
3. gnomAD AF ≤ 0.001
   a. The population frequency of the variant in gnomAD database must not exceed 0.001.
4. VEP Consequence = coding_sequence_variant, feature_elongation, feature_truncation, frameshift_variant, incomplete_terminal_codon_variant, inframe_deletion, inframe_insertion, missense_variant, protein_altering_variant, start_lost, stop_gained, stop_lost, transcript_ablation, transcript_amplification, splice_acceptor_variant, or splice_donor_variant, splice_region_variant
   a. The VEP consequence of any known isoform affected by the variant must include one of these consequences.
5. Has Sample Strand Bias = No
   a. There must be no significant bias from primers that prime off of opposing strands.
6. Has Sequencing Direction Bias = No
   a. There must be no significant bias from read pairs when using Illumina's paired-end chemistry
7. HRUN < 9
   a. The variant must not be flanked by a homopolymer run of greater than 8 bases.
8. ND background noise (BN) ≤ 0.005
   a. The ND error model position-specific background noise for all read families must be less than or equal to 0.005.

9. ND Deep Allele Frequency (DAF) outlier P value ≤ 0.0001
    a. The ND background error model P value for deep consensus read families must be less than or equal to 0.0001.
10. Median distance from start site > 20
    a. The median distance of the variant to the start site generated by random ligation of the molecular barcode adapter must be greater than 20.
11. VAF ≥ 0.0001

***NPM1***

Variants within the *NPM1* gene identified by the targeted (Vision) variant algorithm had the following filters applied:

1. Alternate Observations (AO) ≥ 1
2. VEP Consequence = frameshift_variant
3. VAF ≥ 0.0001

***FLT3-ITD***

Variants resulting in internal tandem duplications (ITD) within exons 14-15 of the *FLT3* gene were identified by either the structural variation algorithm (as described above) or by the *de novo* variant algorithm. For the structural variation algorithm, a minimum of 5 breakpoint-spanning reads were required to support the structural variation. For short ITDs (<20bp) identified by the *de novo* variant algorithm the following filters were applied:

1. Alternate Observations (AO) ≥ 1
2. VEP Consequence = inframe_insertion
3. VAF ≥ 0.0001

All remaining variants underwent manual curation to select pathogenic or likely pathogenic mutations and carried forward for further analysis (**Table S3**).

**Digital droplet polymerase chain reaction (ddPCR)**

A subset of 279 of 408 variants (68%), with NGS detectable VAFs ranging from 0.01 to 63%, across 260 patient samples were orthogonally validated using ddPCR (**Table S3**). Allele-specific fluorescent TaqMan probe assays labeled with FAM or HEX were used for the detection of each mutation according to manufacturer recommendations (ddPCR mutation detection assay, cat# 10049550, Bio-Rad Laboratories, Inc., Hercules, CA). For pre-transplant samples, 110 to 900-ng of gDNA was evaluated, depending on the anticipated allele frequency and/or assay validation. In short, gDNA was combined with a final working concentration of 1X ddPCR mutation detection assay primers/probes, 1X ddPCR Supermix for Probes (no dUTP) (cat# 863023, Bio-Rad Laboratories, Inc., Hercules, CA), and 5U MseI or HindIII restriction enzyme (cat# R0104S or R0525L, New England BioLabs, Inc., Ipswich, MA) in a 22-μL reaction and allowed to incubate for 3 minutes at room temperature. Emulsions were generated using the Automated Droplet Generator (Bio-Rad Laboratories, Inc., Hercules, CA). The emulsion was incubated in a thermocycler using a heated lid set to 105°C and a ramp rate of 2°C per second as follows: 95°C for 10 minutes; 40 cycles of 94°C for 30 seconds, 55°C for 1 minutes; 98°C for 10 minutes. Endpoint fluorescence was measured using the QX200 system and analyzed using QuantaSoft software (Bio-Rad Laboratories, Inc., Hercules, CA). Positive and negative control samples were used to apply thresholds and limit of detection cut-offs using a 95% confidence interval based on a Poisson distribution.

**Conditioning intensity**

Patients were divided into 3 categories of conditioning treatment regimens prior to hematopoietic cell transplantation using the following criteria:

1. Myeloablative conditioning (MAC)
    a. Total body irradiation (TBI) >500 cGy, non-fractionated or TBI >800 cGy, fractionated
    b. Busulfan and Cyclophosphamide
    c. Busulfan and Melphalan
    d. Busulfan and Thiotepa
    e. Cyclophosphamide and Thiotepa
    f. Busulfan (≥9 mg/kg, oral or ≥7.2 mg/kg, IV) and Fludarabine
2. Reduced intensity conditioning (RIC)
    a. TBI ≤500 cGy but >200cGy, non-fractionated or TBI ≤800 cGy but >200cGy, fractionated
    b. Melphalan and Fludarabine

    c. Busulfan (<9 mg/kg, oral or <7.2 mg/kg, IV) and Fludarabine
    d. Cyclophosphamide and BNCU and Etoposide (VP-16) (CBV)
    e. BCNU + Etoposide (VP-16) + Cytarabine (Ara-C) + Melphalan (BEAM)
3. Non-myeloablative conditioning (NMA)
    a. TBI = 200cGy
    b. Fludarabine with or without Cyclophosphamide
    c. Total lymphoid irradiation (TLI) alone

**Statistical analysis**

Clinical data of the 1075 patients were obtained from International Blood and Marrow Transplant Research (CIBMTR). The patient baseline characteristics included age, sex, race, hematopoietic cell transplant specific comorbidity (HCT-CI), Karnofsky score, secondary AML, ELN 2017 risk group, baseline genetics before transplant (*IDH1*, *IDH2*, *KIT*, *NPM1*, *FLT3*-TKD, *FLT3*-ITD), conditioning regimen, graft type, donor group, anti-thymocyte globulin (ATG) usage. Data on site MRD status for patients were obtained when available. The characteristics were compared between cohort 1 and cohort 2 using student's t-test or Mann-Whitney U test for continuous variables, and chi-squared test or Fisher's exact test for categorical variables.

Day of transplant was considered as day 0 for summarizing the clinical endpoints. The primary outcomes were overall survival (OS) and cumulative incidence of relapse (CIR) with non-relapse mortality (NRM) as a competing risk. The secondary outcome was relapse-free survival (RFS), defined as the time from day of transplant until the date of morphologic relapse or death from any cause. Kaplan-Meier estimation and log rank tests were used to calculate OS and RFS endpoints; Fine-Gray regression models were used to examine CIR with NRM as a competing risk. The cumulative rates of clinical endpoints were estimated at important clinical time points including 3 months, 6 months, 1 year, 2 years, and 3 years with multiple Z-test for point-wise p-value calculation.

Optimal combinations of NGS detected mutations on different genes for predicting relapse events were determined by cohort 1 and validated in cohort 2. Cox proportional hazards regression models were used to estimate the relative risks of clinical events given the baseline characteristics and NGS-MRD status and the proportional hazards assumptions were assessed. All clinical characteristics were included in the model selection with a forward selection process, and Cox models with Lasso penalty were also fitted. Potential interactions between NGS-MRD status and conditioning intensity, baseline genetics were explored. The prognostic value of tracking mutations in different genes were estimated by comparing the clinical outcomes of the NGS-MRD status in stratified cohorts by baseline genetics.

The pre-specified statistical analysis plan was registered with OSF: Gui, G., Dillon, L., & Hourigan, C. (2022, January 7). Allogeneic Transplantation for Acute Myeloid Leukemia with Genomic Evidence of Residual Disease (https://osf.io/a6epx).

**Figure S1. CONSORT diagram.** The criteria used for selection of patients for pre-transplant next-generation sequencing measurable residual disease (NGS-MRD) analysis. alloHCT, allogeneic hematopoietic cell transplantation; CIBMTR, Center for International Blood and Marrow Transplant Research; gDNA, genomic DNA.

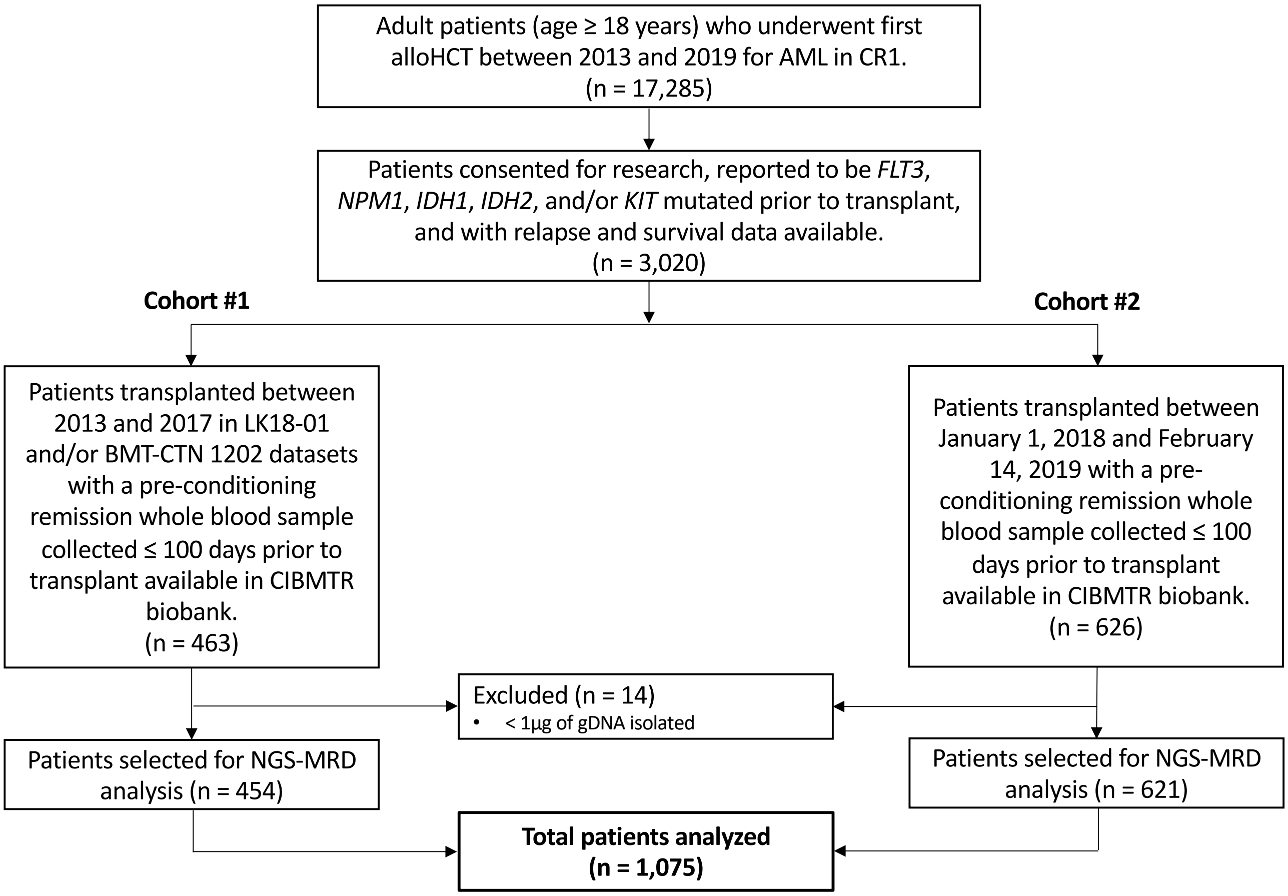

**Figure S2. Association of baseline patient characteristics and site MRD on clinical outcomes.**

**i. Baseline patient characteristics.** Rates of non-relapse mortality (NRM, top left), relapse (top right), relapse-free survival (RFS, bottom left), and overall survival (OS, bottom right) censored at 72 months are shown by gene based on baseline patient characteristics (A) age, (B) sex, (C) race, (D) HCT-CI, (E) Karnofsky score, (F) Secondary AML, (G) ELN risk group, (H) conditioning regimen, (I) graft type, (J) donor type, and (K) anti-thymocyte globulin (ATG). Point estimates at different time points are shown in the table (far right). Overall P values: Gray's test for NRM and relapse; log-rank test for DFS and OS. P values for pointwise estimations at different time points: z-test. Confidence interval, CI; Probability, prob; Month, mo; Year, yr.

A

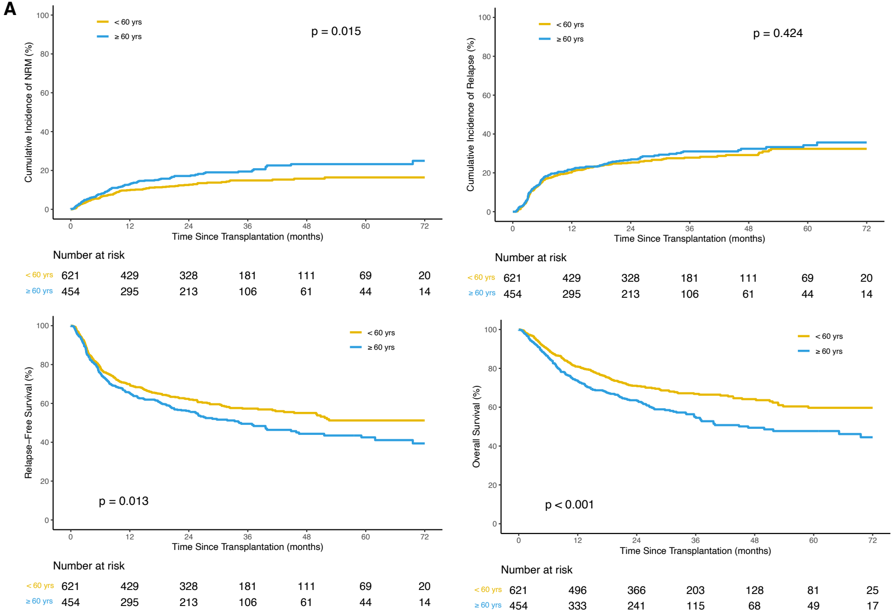


##### Age

| Outcomes | Prob (95% CI) | Prob (95% CI) | p-value |
|---|---|---|---|
| NRM | <60yrs=621 | ≥60yrs=454 | 0.015 |
| 3 mo | 4(2-5)% | 5(3-7)% | 0.298 |
| 6 mo | 6(4-8)% | 8(5-10)% | 0.369 |
| 1 yr | 10(8-12)% | 13(10-16)% | 0.136 |
| 2 yr | 13(10-15)% | 17(14-21)% | 0.037 |
| 3 yr | 15(12-18)% | 19(16-23)% | 0.065 |
| Relapse | <60yrs=621 | ≥60yrs=454 | 0.424 |
| 3 mo | 7(5-9)% | 6(4-9)% | 0.888 |
| 6 mo | 15(12-18)% | 17(13-20)% | 0.526 |
| 1 yr | 20(17-24)% | 22(18-26)% | 0.553 |
| 2 yr | 25(22-29)% | 27(23-31)% | 0.57 |
| 3 yr | 28(24-32)% | 31(26-36)% | 0.284 |
| RFS | <60yrs=621 | ≥60yrs=454 | 0.013 |
| 3 mo | 90(87-92)% | 89(85-91)% | 0.57 |
| 6 mo | 78(75-81)% | 76(71-79)% | 0.271 |
| 1 yr | 70(66-73)% | 65(61-70)% | 0.125 |
| 2 yr | 62(58-66)% | 56(51-61)% | 0.043 |
| 3 yr | 57(53-61)% | 50(44-54)% | 0.019 |
| OS | <60yrs=621 | ≥60yrs=454 | <0.001 |
| 3 mo | 96(95-98)% | 93(91-95)% | 0.027 |
| 6 mo | 90(87-92)% | 86(82-89)% | 0.061 |
| 1 yr | 81(78-84)% | 74(69-78)% | 0.006 |
| 2 yr | 71(67-74)% | 64(59-68)% | 0.011 |
| 3 yr | 67(63-71)% | 55(50-60)% | <0.001 |

B

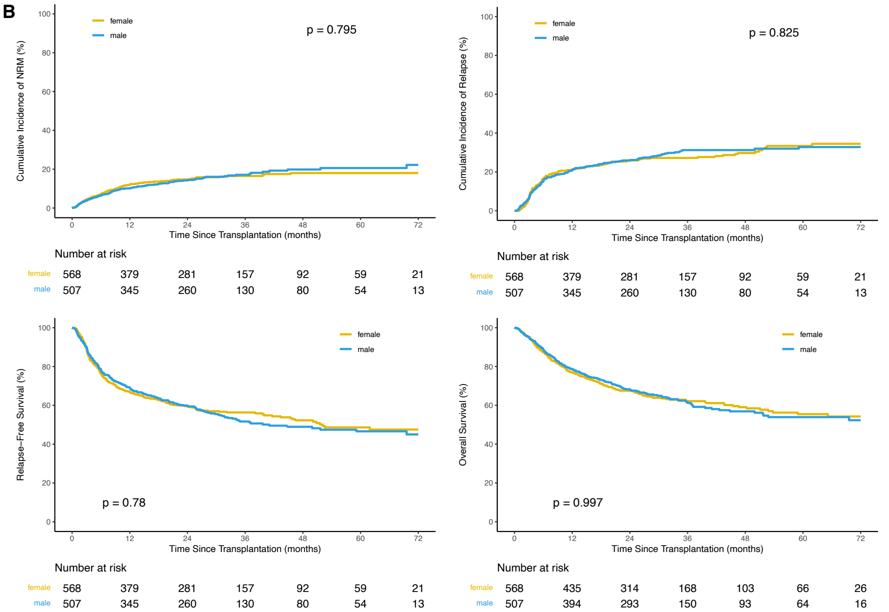


##### Sex

| Outcomes | Prob (95% CI) | Prob (95% CI) | p-value |
|---|---|---|---|
| NRM | female=568 | male=507 | 0.795 |
| 3 mo | 4(3-6)% | 4(2-5)% | 0.588 |
| 6 mo | 7(5-9)% | 7(4-9)% | 0.647 |
| 1 yr | 12(9-15)% | 10(7-13)% | 0.322 |
| 2 yr | 15(12-18)% | 14(11-17)% | 0.813 |
| 3 yr | 16(13-20)% | 17(14-21)% | 0.796 |
| Relapse | female=568 | male=507 | 0.825 |
| 3 mo | 7(4-9)% | 7(4-9)% | 0.997 |
| 6 mo | 17(14-20)% | 15(12-18)% | 0.544 |
| 1 yr | 21(18-24)% | 21(17-25)% | 1 |
| 2 yr | 26(22-29)% | 26(22-30)% | 0.948 |
| 3 yr | 27(23-31)% | 31(27-36)% | 0.17 |
| RFS | female=568 | male=507 | 0.78 |
| 3 mo | 89(86-91)% | 90(87-92)% | 0.726 |
| 6 mo | 76(72-80)% | 78(74-82)% | 0.42 |
| 1 yr | 67(63-71)% | 69(65-73)% | 0.506 |
| 2 yr | 60(55-64)% | 60(55-64)% | 0.911 |
| 3 yr | 56(52-60)% | 52(47-56)% | 0.154 |
| OS | female=568 | male=507 | 0.997 |
| 3 mo | 95(93-97)% | 95(93-97)% | 0.881 |
| 6 mo | 87(84-90)% | 89(85-91)% | 0.536 |
| 1 yr | 77(73-80)% | 79(75-82)% | 0.492 |
| 2 yr | 68(63-71)% | 68(64-72)% | 0.789 |
| 3 yr | 62(58-66)% | 62(57-66)% | 0.866 |

C

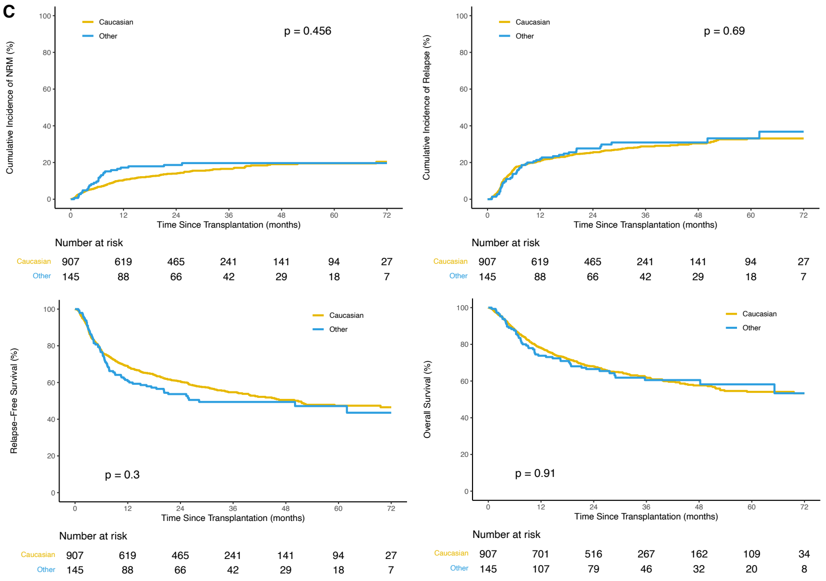


#### Race

| Outcomes | Prob (95% CI) | Prob (95% CI) | p-value |
|---|---|---|---|
| NRM | Caucasian=907 | Other=145 | 0.456 |
| 3 mo | 4(3-5)% | 5(1-8)% | 0.652 |
| 6 mo | 7(5-8)% | 10(5-14)% | 0.226 |
| 1 yr | 10(8-12)% | 17(11-23)% | 0.035 |
| 2 yr | 14(12-16)% | 19(12-25)% | 0.173 |
| 3 yr | 17(14-19)% | 20(13-26)% | 0.384 |
| Relapse | Caucasian=907 | Other=145 | 0.69 |
| 3 mo | 6(5-8)% | 6(2-10)% | 0.931 |
| 6 mo | 16(14-19)% | 14(8-19)% | 0.436 |
| 1 yr | 21(18-23)% | 22(15-29)% | 0.727 |
| 2 yr | 25(23-28)% | 28(20-35)% | 0.564 |
| 3 yr | 29(26-32)% | 31(23-39)% | 0.619 |
| RFS | Caucasian=907 | Other=145 | 0.3 |
| 3 mo | 90(87-91)% | 89(83-93)% | 0.81 |
| 6 mo | 77(74-80)% | 77(69-83)% | 0.852 |
| 1 yr | 69(66-72)% | 61(52-68)% | 0.057 |
| 2 yr | 61(57-64)% | 54(45-61)% | 0.116 |
| 3 yr | 55(51-58)% | 49(41-58)% | 0.258 |
| OS | Caucasian=907 | Other=145 | 0.91 |
| 3 mo | 95(94-96)% | 95(90-98)% | 0.964 |
| 6 mo | 88(86-90)% | 88(81-92)% | 0.928 |
| 1 yr | 78(75-81)% | 74(66-80)% | 0.258 |
| 2 yr | 68(65-71)% | 67(58-74)% | 0.711 |
| 3 yr | 62(58-65)% | 61(51-69)% | 0.753 |

D

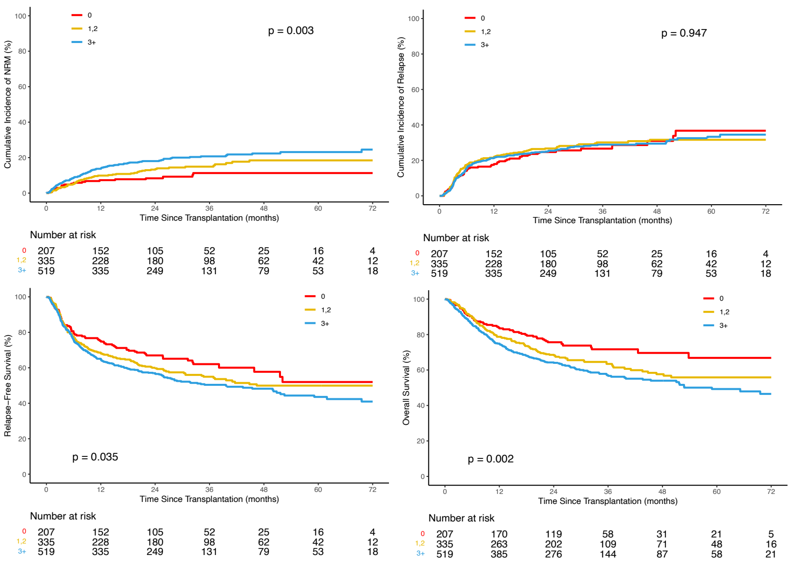


#### HCT-CI

| Outcomes | Prob (95% CI) | Prob (95% CI) | Prob (95% CI) | p-value |
|---|---|---|---|---|
| NRM | 0=207 | 1,2=335 | 3+=519 | 0.003 |
| 3 mo | 3(1-5)% | 3(1-5)% | 5(3-7)% | 0.139 |
| 6 mo | 6(3-9)% | 5(3-7)% | 9(6-11)% | 0.098 |
| 1 yr | 7(4-11)% | 10(7-13)% | 14(11-17)% | 0.021 |
| 2 yr | 8(5-12)% | 13(10-17)% | 18(15-21)% | <0.001 |
| 3 yr | 11(6-16)% | 15(11-19)% | 21(17-24)% | 0.007 |
| Relapse | 0=207 | 1,2=335 | 3+=519 | 0.947 |
| 3 mo | 7(4-11)% | 7(4-9)% | 6(4-8)% | 0.873 |
| 6 mo | 15(10-19)% | 17(13-21)% | 16(12-19)% | 0.661 |
| 1 yr | 17(12-23)% | 22(17-26)% | 21(18-25)% | 0.388 |
| 2 yr | 25(19-31)% | 27(22-31)% | 25(21-29)% | 0.821 |
| 3 yr | 27(20-33)% | 30(25-35)% | 29(25-33)% | 0.709 |
| RFS | 0=207 | 1,2=335 | 3+=519 | 0.035 |
| 3 mo | 90(85-93)% | 90(87-93)% | 88(85-91)% | 0.625 |
| 6 mo | 80(74-85)% | 78(73-82)% | 76(72-79)% | 0.491 |
| 1 yr | 75(69-81)% | 68(63-73)% | 65(61-69)% | 0.018 |
| 2 yr | 67(60-73)% | 60(55-65)% | 57(53-61)% | 0.042 |
| 3 yr | 62(54-69)% | 55(49-60)% | 50(46-55)% | 0.029 |
| OS | 0=207 | 1,2=335 | 3+=519 | 0.002 |
| 3 mo | 97(93-98)% | 97(94-98)% | 93(91-95)% | 0.052 |
| 6 mo | 89(84-93)% | 90(86-93)% | 86(82-88)% | 0.15 |
| 1 yr | 84(78-88)% | 79(74-83)% | 75(71-78)% | 0.017 |
| 2 yr | 76(69-81)% | 69(63-73)% | 64(60-68)% | 0.009 |
| 3 yr | 72(64-78)% | 63(58-69)% | 57(52-62)% | 0.002 |

E

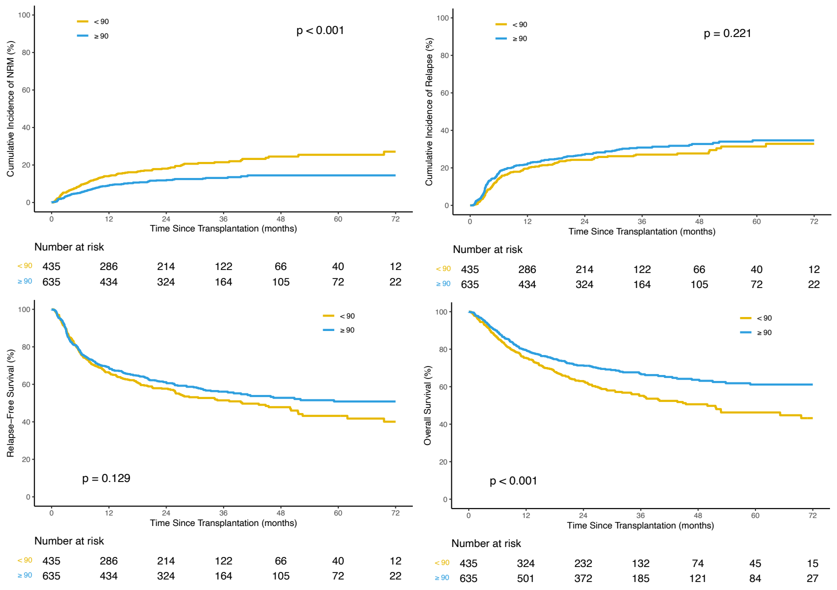


#### Karnofsky Score

| Outcomes | Prob (95% CI) | Prob (95% CI) | p-value |
|---|---|---|---|
| NRM | <90=435 | ≥90=635 | <0.001 |
| 3 mo | 5(3-7)% | 3(2-5)% | 0.124 |
| 6 mo | 9(6-12)% | 5(4-7)% | 0.028 |
| 1 yr | 14(11-17)% | 9(7-11)% | 0.013 |
| 2 yr | 18(14-22)% | 12(9-14)% | 0.006 |
| 3 yr | 21(17-26)% | 13(10-16)% | <0.001 |
| Relapse | <90=435 | ≥90=635 | 0.221 |
| 3 mo | 5(3-7)% | 8(6-10)% | 0.092 |
| 6 mo | 14(11-17)% | 17(14-20)% | 0.138 |
| 1 yr | 20(16-23)% | 22(19-25)% | 0.307 |
| 2 yr | 24(20-28)% | 27(24-31)% | 0.308 |
| 3 yr | 27(23-31)% | 31(27-35)% | 0.207 |
| RFS | <90=435 | ≥90=635 | 0.129 |
| 3 mo | 90(86-92)% | 89(86-91)% | 0.785 |
| 6 mo | 77(73-81)% | 77(74-80)% | 0.917 |
| 1 yr | 66(62-71)% | 69(65-72)% | 0.404 |
| 2 yr | 58(53-62)% | 61(57-65)% | 0.258 |
| 3 yr | 51(46-56)% | 56(52-60)% | 0.165 |
| OS | <90=435 | ≥90=635 | <0.001 |
| 3 mo | 94(92-96)% | 96(94-97)% | 0.417 |
| 6 mo | 86(83-89)% | 89(86-91)% | 0.16 |
| 1 yr | 75(71-79)% | 80(76-83)% | 0.107 |
| 2 yr | 63(58-67)% | 71(68-75)% | 0.005 |
| 3 yr | 55(50-60)% | 67(63-71)% | <0.001 |

F

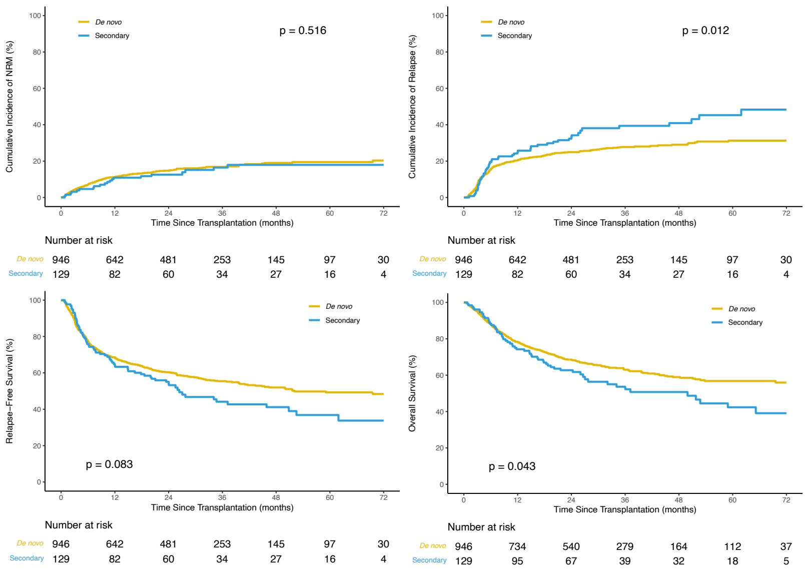


#### Secondary AML

| Outcomes | Prob (95% CI) | Prob (95% CI) | p-value |
|---|---|---|---|
| NRM | *De novo*=946 | Secondary=129 | 0.516 |
| 3 mo | 4(3-6)% | 3(0-6)% | 0.499 |
| 6 mo | 7(6-9)% | 5(1-8)% | 0.217 |
| 1 yr | 11(9-13)% | 11(5-16)% | 0.946 |
| 2 yr | 15(12-17)% | 13(7-18)% | 0.497 |
| 3 yr | 17(14-19)% | 16(9-23)% | 0.927 |
| Relapse | *De novo*=946 | Secondary=129 | 0.012 |
| 3 mo | 7(5-8)% | 4(1-7)% | 0.114 |
| 6 mo | 15(13-18)% | 19(13-26)% | 0.275 |
| 1 yr | 20(18-23)% | 25(17-32)% | 0.265 |
| 2 yr | 25(22-28)% | 32(24-41)% | 0.092 |
| 3 yr | 28(25-31)% | 39(30-49)% | 0.018 |
| RFS | *De novo*=946 | Secondary=129 | 0.083 |
| 3 mo | 89(87-91)% | 93(87-96)% | 0.094 |
| 6 mo | 77(75-80)% | 76(67-82)% | 0.706 |
| 1 yr | 68(65-71)% | 64(55-72)% | 0.336 |
| 2 yr | 60(57-63)% | 55(46-63)% | 0.262 |
| 3 yr | 55(52-59)% | 44(35-53)% | 0.028 |
| OS | *De novo*=946 | Secondary=129 | 0.043 |
| 3 mo | 95(93-96)% | 96(91-98)% | 0.553 |
| 6 mo | 88(86-90)% | 88(81-93)% | 0.875 |
| 1 yr | 78(76-81)% | 74(66-81)% | 0.311 |
| 2 yr | 69(65-71)% | 63(54-71)% | 0.202 |
| 3 yr | 63(60-66)% | 54(44-63)% | 0.07 |

G

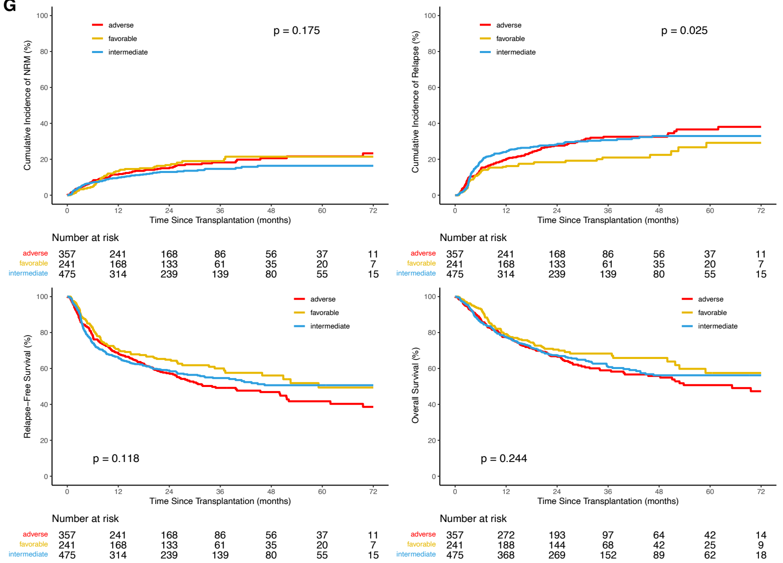


#### ELN risk group

| Outcomes | Prob (95% CI) | Prob (95% CI) | Prob (95% CI) | p-value |
|---|---|---|---|---|
| NRM | Favorable=241 | Intermediate=475 | Adverse=357 | 0.175 |
| 3 mo | 3(1-6)% | 4(3-6)% | 4(2-6)% | 0.751 |
| 6 mo | 5(2-8)% | 7(5-9)% | 8(5-11)% | 0.319 |
| 1 yr | 13(9-18)% | 10(7-12)% | 12(8-15)% | 0.343 |
| 2 yr | 17(12-22)% | 13(10-16)% | 15(11-19)% | 0.378 |
| 3 yr | 19(14-24)% | 15(11-18)% | 18(14-23)% | 0.263 |
| Relapse | Favorable=241 | Intermediate=475 | Adverse=357 | 0.025 |
| 3 mo | 5(3-8)% | 5(3-7)% | 9(6-12)% | 0.107 |
| 6 mo | 13(9-17)% | 19(15-22)% | 14(10-18)% | 0.068 |
| 1 yr | 16(11-20)% | 24(20-28)% | 20(16-24)% | 0.027 |
| 2 yr | 18(13-23)% | 28(24-32)% | 28(23-32)% | 0.005 |
| 3 yr | 21(15-27)% | 31(26-35)% | 33(27-38)% | 0.006 |
| RFS | Favorable=241 | Intermediate=475 | Adverse=357 | 0.118 |
| 3 mo | 91(87-94)% | 90(87-93)% | 87(83-90)% | 0.171 |
| 6 mo | 82(77-86)% | 74(70-78)% | 78(73-82)% | 0.04 |
| 1 yr | 71(65-76)% | 66(62-70)% | 68(63-73)% | 0.452 |
| 2 yr | 65(58-71)% | 59(54-63)% | 57(52-62)% | 0.159 |
| 3 yr | 60(53-66)% | 55(50-59)% | 49(43-55)% | 0.055 |
| OS | Favorable=241 | Intermediate=475 | Adverse=357 | 0.244 |
| 3 mo | 96(93-98)% | 95(93-97)% | 94(91-96)% | 0.465 |
| 6 mo | 93(89-96)% | 86(82-89)% | 87(83-90)% | 0.005 |
| 1 yr | 79(73-84)% | 78(74-81)% | 77(73-81)% | 0.868 |
| 2 yr | 71(64-76)% | 67(63-71)% | 67(62-71)% | 0.605 |
| 3 yr | 68(62-74)% | 61(56-66)% | 59(53-64)% | 0.075 |

H

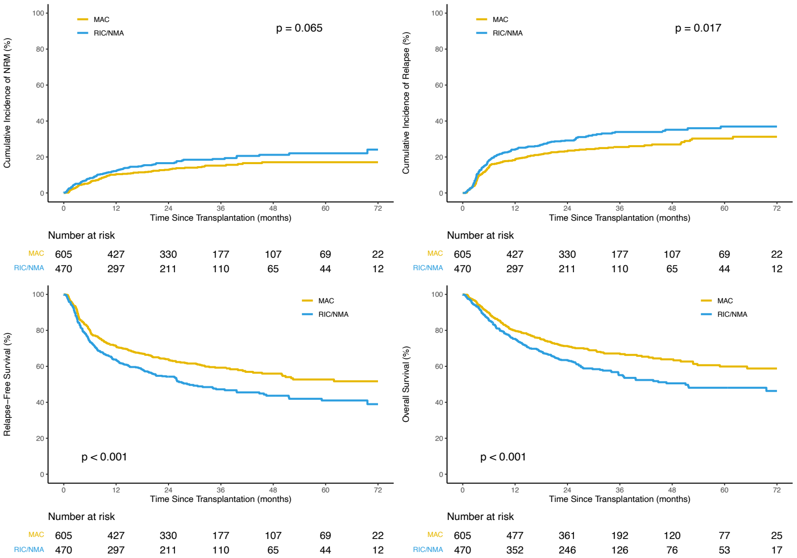


#### Conditioning

| Outcomes | Prob (95% CI) | Prob (95% CI) | p-value |
|---|---|---|---|
| NRM | MAC=605 | RIC/NMA=470 | 0.065 |
| 3 mo | 3(2-5)% | 5(3-7)% | 0.15 |
| 6 mo | 6(4-8)% | 8(6-11)% | 0.114 |
| 1 yr | 10(8-13)% | 12(9-15)% | 0.34 |
| 2 yr | 13(10-16)% | 17(13-20)% | 0.092 |
| 3 yr | 15(12-18)% | 19(15-23)% | 0.135 |
| Relapse | MAC=605 | RIC/NMA=470 | 0.017 |
| 3 mo | 5(4-7)% | 8(6-11)% | 0.072 |
| 6 mo | 14(11-17)% | 18(15-22)% | 0.089 |
| 1 yr | 18(15-21)% | 24(20-28)% | 0.02 |
| 2 yr | 23(20-27)% | 29(25-33)% | 0.027 |
| 3 yr | 26(22-29)% | 34(29-39)% | 0.005 |
| RFS | MAC=605 | RIC/NMA=470 | <0.001 |
| 3 mo | 91(89-93)% | 87(83-90)% | 0.017 |
| 6 mo | 80(77-83)% | 74(69-77)% | 0.014 |
| 1 yr | 71(68-75)% | 64(59-68)% | 0.007 |
| 2 yr | 64(60-68)% | 54(50-59)% | 0.001 |
| 3 yr | 59(55-63)% | 47(42-52)% | <0.001 |
| OS | MAC=605 | RIC/NMA=470 | <0.001 |
| 3 mo | 96(94-97)% | 94(91-96)% | 0.08 |
| 6 mo | 90(87-92)% | 86(82-88)% | 0.038 |
| 1 yr | 80(76-83)% | 75(71-79)% | 0.071 |
| 2 yr | 71(67-75)% | 63(59-68)% | 0.007 |
| 3 yr | 67(63-71)% | 55(50-60)% | <0.001 |

I

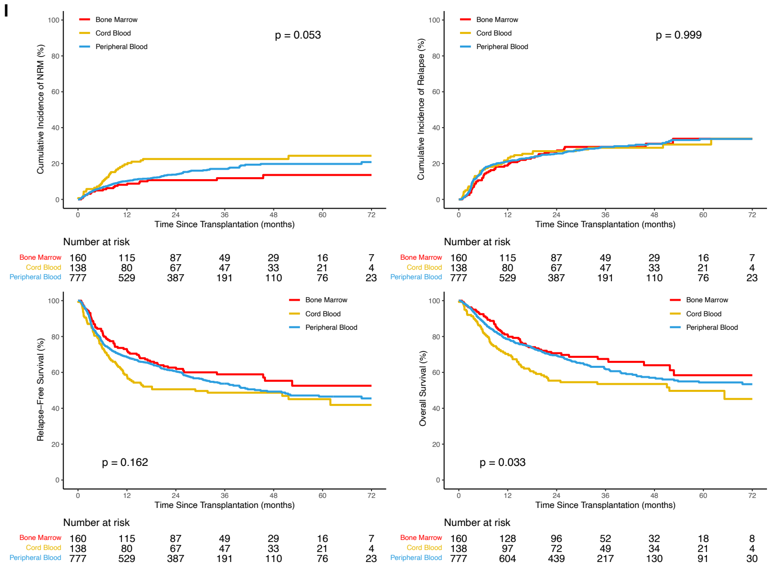


#### Graft Type

| Outcomes | Prob (95% CI) | Prob (95% CI) | Prob (95% CI) | p-value |
|---|---|---|---|---|
| NRM | Bone Marrow n=160 | Cord Blood n=138 | Peripheral Blood n=777 | 0.053 |
| 3 mo | 3(0-6)% | 6(2-10)% | 4(3-5)% | 0.546 |
| 6 mo | 5(2-8)% | 10(5-15)% | 7(5-8)% | 0.255 |
| 1 yr | 8(4-12)% | 20(13-26)% | 10(8-12)% | 0.015 |
| 2 yr | 11(6-16)% | 23(16-30)% | 14(11-16)% | 0.024 |
| 3 yr | 12(7-17)% | 23(16-30)% | 17(14-20)% | 0.05 |
| Relapse | Bone Marrow n=160 | Cord Blood n=138 | Peripheral Blood n=777 | 0.999 |
| 3 mo | 4(1-7)% | 7(3-12)% | 7(5-9)% | 0.17 |
| 6 mo | 13(7-18)% | 17(10-23)% | 16(14-19)% | 0.392 |
| 1 yr | 19(13-25)% | 22(15-29)% | 21(18-24)% | 0.719 |
| 2 yr | 27(20-34)% | 27(19-34)% | 25(22-29)% | 0.91 |
| 3 yr | 29(22-37)% | 29(21-37)% | 29(26-33)% | 0.994 |
| RFS | Bone Marrow n=160 | Cord Blood n=138 | Peripheral Blood n=777 | 0.162 |
| 3 mo | 93(88-96)% | 87(80-92)% | 89(87-91)% | 0.124 |
| 6 mo | 82(76-88)% | 73(65-80)% | 77(74-80)% | 0.122 |
| 1 yr | 73(65-79)% | 58(49-66)% | 69(65-72)% | 0.02 |
| 2 yr | 63(55-70)% | 51(42-59)% | 61(57-64)% | 0.063 |
| 3 yr | 59(50-66)% | 49(40-57)% | 54(50-58)% | 0.225 |
| OS | Bone Marrow n=160 | Cord Blood n=138 | Peripheral Blood n=777 | 0.033 |
| 3 mo | 96(92-98)% | 92(86-96)% | 95(94-97)% | 0.291 |
| 6 mo | 92(87-96)% | 82(74-87)% | 88(86-90)% | 0.02 |
| 1 yr | 81(74-86)% | 70(62-77)% | 79(76-81)% | 0.078 |
| 2 yr | 71(63-77)% | 55(47-63)% | 70(66-73)% | 0.006 |
| 3 yr | 67(59-75)% | 54(45-62)% | 62(58-66)% | 0.056 |

J

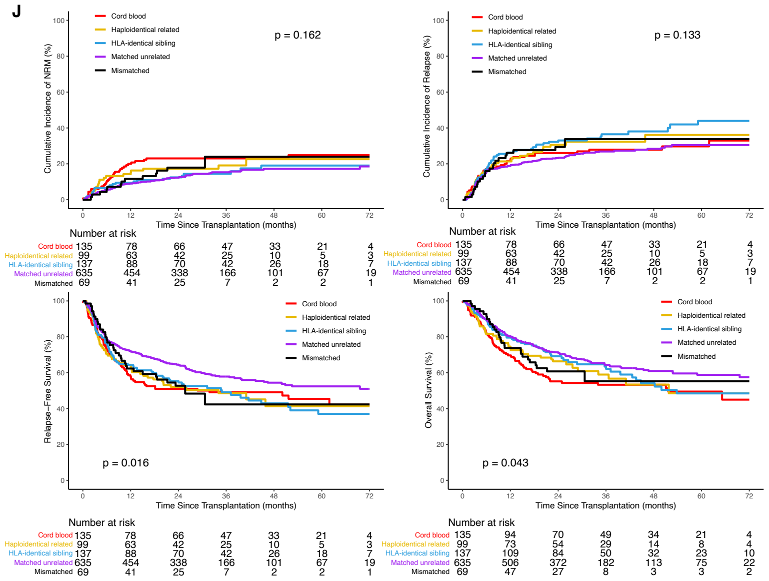


#### Donor Type

| Outcomes | Prob (95% CI) | Prob (95% CI) | Prob (95% CI) | Prob (95% CI) | Prob (95% CI) | p-value |
|---|---|---|---|---|---|---|
| NRM | Cord Blood n=135 | Haploidentical Related n=99 | HLA-identical Sibling n=137 | Matched Unrelated n=635 | Mismatched n=69 | 0.162 |
| 3 mo | 6(2-10)% | 5(1-9)% | 7(2-11)% | 3(2-5)% | 3(0-7)% | 0.384 |
| 6 mo | 10(5-16)% | 13(6-20)% | 7(2-11)% | 5(4-7)% | 6(0-11)% | 0.108 |
| 1 yr | 20(13-27)% | 14(7-21)% | 9(5-14)% | 9(7-11)% | 12(4-19)% | 0.031 |
| 2 yr | 23(16-30)% | 17(10-25)% | 12(7-18)% | 12(10-15)% | 18(9-27)% | 0.05 |
| 3 yr | 23(16-30)% | 19(11-27)% | 14(8-21)% | 15(12-18)% | 24(9-39)% | 0.229 |
| Relapse | Cord Blood n=135 | Haploidentical Related n=99 | HLA-identical Sibling n=137 | Matched Unrelated n=635 | Mismatched n=69 | 0.133 |
| 3 mo | 7(3-12)% | 8(3-13)% | 7(3-12)% | 6(4-8)% | 6(0-11)% | 0.914 |
| 6 mo | 17(11-23)% | 16(9-24)% | 17(11-23)% | 15(13-18)% | 16(7-25)% | 0.989 |
| 1 yr | 22(15-29)% | 21(13-29)% | 26(19-34)% | 19(16-22)% | 26(16-37)% | 0.321 |
| 2 yr | 26(19-33)% | 31(21-40)% | 32(24-40)% | 23(20-27)% | 29(18-40)% | 0.176 |
| 3 yr | 28(20-36)% | 32(23-42)% | 36(28-45)% | 27(23-31)% | 34(20-47)% | 0.259 |
| RFS | Cord Blood n=135 | Haploidentical Related n=99 | HLA-identical Sibling n=137 | Matched Unrelated n=635 | Mismatched n=69 | 0.016 |
| 3 mo | 87(80-91)% | 87(78-92)% | 86(79-91)% | 91(88-93)% | 91(82-96)% | 0.343 |
| 6 mo | 73(64-79)% | 71(60-79)% | 77(69-83)% | 79(76-82)% | 78(67-86)% | 0.277 |
| 1 yr | 58(49-66)% | 64(54-73)% | 64(56-72)% | 72(68-75)% | 62(50-73)% | 0.01 |
| 2 yr | 51(42-59)% | 52(42-61)% | 55(47-63)% | 65(61-68)% | 53(40-64)% | 0.005 |
| 3 yr | 49(40-57)% | 49(38-58)% | 49(40-58)% | 58(54-62)% | 42(26-58)% | 0.07 |
| OS | Cord Blood n=135 | Haploidentical Related n=99 | HLA-identical Sibling n=137 | Matched Unrelated n=635 | Mismatched n=69 | 0.043 |
| 3 mo | 92(86-95)% | 93(86-97)% | 93(88-97)% | 96(95-98)% | 97(89-99)% | 0.178 |
| 6 mo | 81(74-87)% | 82(73-88)% | 89(82-93)% | 90(87-92)% | 91(82-96)% | 0.061 |
| 1 yr | 70(61-77)% | 75(65-82)% | 80(72-85)% | 80(77-83)% | 74(62-83)% | 0.095 |
| 2 yr | 55(46-63)% | 66(56-75)% | 69(61-76)% | 71(68-75)% | 61(48-71)% | 0.008 |
| 3 yr | 53(44-61)% | 59(47-69)% | 63(54-71)% | 64(60-68)% | 55(39-69)% | 0.172 |

K

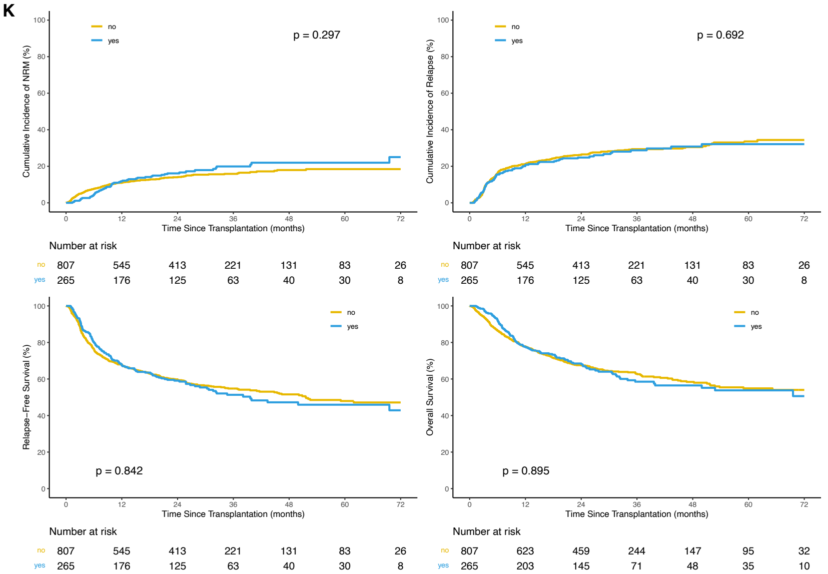


#### ATG

| Outcomes | Prob (95% CI) | Prob (95% CI) | p-value |
|---|---|---|---|
| NRM | no=807 | yes=265 | 0.297 |
| 3 mo | 5(3-6)% | 2(0-3)% | 0.001 |
| 6 mo | 8(6-10)% | 5(2-7)% | 0.047 |
| 1 yr | 11(9-13)% | 12(8-16)% | 0.708 |
| 2 yr | 14(12-16)% | 16(12-21)% | 0.414 |
| 3 yr | 16(13-18)% | 20(15-25)% | 0.174 |
| Relapse | no=807 | yes=265 | 0.692 |
| 3 mo | 6(5-8)% | 7(4-10)% | 0.844 |
| 6 mo | 16(14-19)% | 15(11-20)% | 0.802 |
| 1 yr | 21(18-24)% | 21(16-26)% | 0.922 |
| 2 yr | 26(23-29)% | 25(20-30)% | 0.644 |
| 3 yr | 29(26-33)% | 29(23-35)% | 0.855 |
| RFS | no=807 | yes=265 | 0.842 |
| 3 mo | 89(86-91)% | 92(88-94)% | 0.127 |
| 6 mo | 76(73-79)% | 80(75-84)% | 0.187 |
| 1 yr | 68(65-71)% | 67(61-73)% | 0.864 |
| 2 yr | 60(56-63)% | 59(53-65)% | 0.845 |
| 3 yr | 55(51-58)% | 51(45-58)% | 0.375 |
| OS | no=807 | yes=265 | 0.895 |
| 3 mo | 94(92-96)% | 98(96-99)% | <0.001 |
| 6 mo | 86(84-89)% | 92(88-95)% | 0.006 |
| 1 yr | 78(75-81)% | 78(72-82)% | 0.922 |
| 2 yr | 68(64-71)% | 68(62-74)% | 0.785 |
| 3 yr | 63(59-66)% | 59(52-65)% | 0.26 |

**ii. Site MRD.** Rates of non-relapse mortality (top left), relapse (top right), relapse-free survival (bottom left), and overall survival (bottom right) censored at 72 months are shown for patients with data available for measurable residual disease (MRD) as reported by the treatment site within (A) the full cohort (n=1,001) and (B) patients with baseline *NPM1* and/or *FLT3*-ITD mutations (n=765). Point estimates at different time points are shown in the table (far right). Overall P values: Gray's test for non-relapse mortality (NRM) and relapse; log-rank test for relapse-free survival (RFS) and overall survival (OS). P values for pointwise estimations at different time points: z-test. Confidence interval, CI; Probability, prob; Month, mo; Year, yr; positive, pos; negative, neg.

A

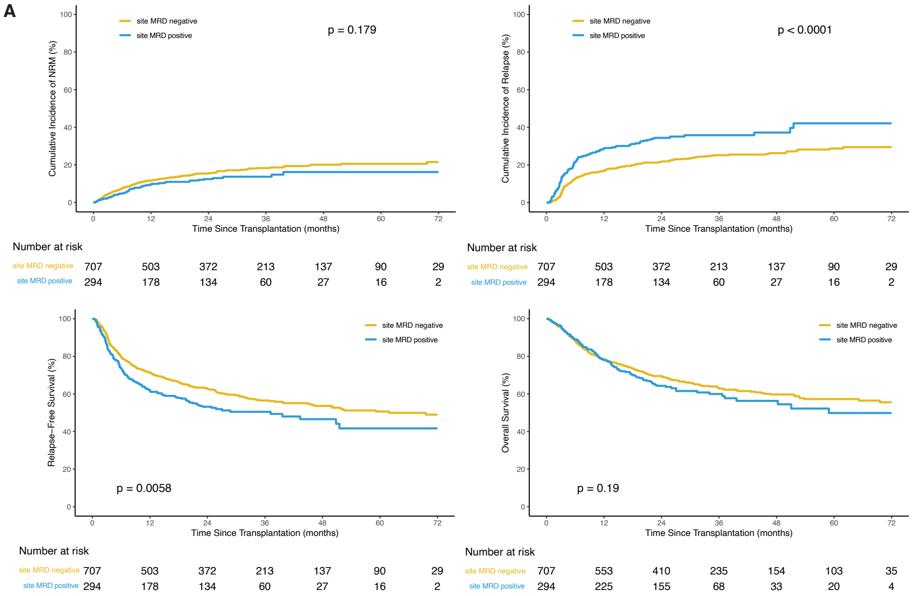


**Site MRD, All Patients**

| Outcomes | Prob (95% CI) | Prob (95% CI) | p-value |
|---|---|---|---|
| NRM | MRDneg=707 | MRDpos=294 | 0.179 |
| 3 mo | 4(3-6)% | 2(1-4)% | 0.089 |
| 6 mo | 8(6-10)% | 5(2-7)% | 0.072 |
| 1 yr | 12(9-14)% | 10(6-13)% | 0.325 |
| 2 yr | 15(13-18)% | 12(9-16)% | 0.211 |
| 3 yr | 18(15-21)% | 14(10-18)% | 0.078 |
| Relapse | MRDneg=707 | MRDpos=294 | <0.001 |
| 3 mo | 4(3-6)% | 12(8-15)% | <0.001 |
| 6 mo | 12(10-15)% | 22(17-27)% | <0.001 |
| 1 yr | 17(14-20)% | 29(23-34)% | <0.001 |
| 2 yr | 22(18-25)% | 34(29-40)% | <0.001 |
| 3 yr | 25(22-29)% | 36(30-42)% | 0.002 |
| RFS | MRDneg=707 | MRDpos=294 | 0.006 |
| 3 mo | 91(89-93)% | 86(82-90)% | 0.02 |
| 6 mo | 80(77-83)% | 73(68-78)% | 0.03 |
| 1 yr | 72(68-75)% | 62(56-67)% | 0.003 |
| 2 yr | 63(59-67)% | 53(47-59)% | 0.004 |
| 3 yr | 57(52-60)% | 50(44-56)% | 0.102 |
| OS | MRDneg=707 | MRDpos=294 | 0.19 |
| 3 mo | 95(93-97)% | 96(93-98)% | 0.676 |
| 6 mo | 88(85-90)% | 89(85-92)% | 0.605 |
| 1 yr | 79(75-81)% | 78(73-82)% | 0.884 |
| 2 yr | 70(66-73)% | 64(58-70)% | 0.119 |
| 3 yr | 63(59-67)% | 60(53-66)% | 0.392 |

B

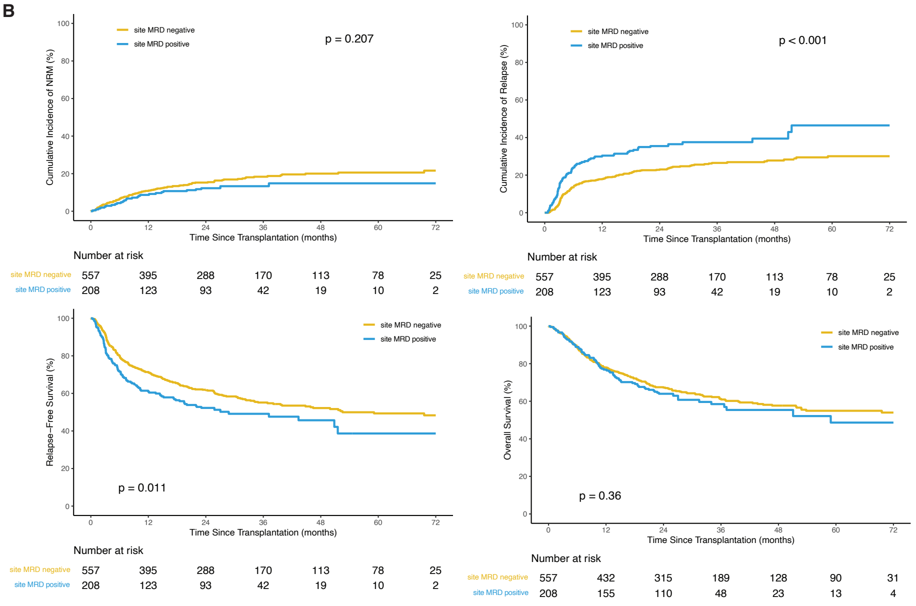


**Site MRD, Baseline *NPM1*/*FLT3*-ITDpos**

| Outcomes | Prob (95% CI) | Prob (95% CI) | p-value |
|---|---|---|---|
| NRM | MRDneg=557 | MRDpos=208 | 0.207 |
| 3 mo | 4(2-5)% | 2(0-4)% | 0.371 |
| 6 mo | 7(5-9)% | 4(2-7)% | 0.19 |
| 1 yr | 11(8-13)% | 9(5-13)% | 0.37 |
| 2 yr | 15(12-18)% | 12(8-17)% | 0.28 |
| 3 yr | 18(15-22)% | 13(8-18)% | 0.1 |
| Relapse | MRDneg=557 | MRDpos=208 | <0.001 |
| 3 mo | 5(3-7)% | 13(9-18)% | <0.001 |
| 6 mo | 13(11-16)% | 24(18-30)% | 0.001 |
| 1 yr | 18(15-21)% | 30(24-37)% | <0.001 |
| 2 yr | 23(19-26)% | 35(29-42)% | <0.001 |
| 3 yr | 26(23-30)% | 38(31-45)% | 0.007 |
| RFS | MRDneg=557 | MRDpos=208 | 0.011 |
| 3 mo | 92(89-94)% | 84(78-88)% | 0.008 |
| 6 mo | 80(76-83)% | 72(65-77)% | 0.02 |
| 1 yr | 71(67-75)% | 61(54-67)% | 0.007 |
| 2 yr | 62(58-66)% | 52(45-59)% | 0.018 |
| 3 yr | 55(51-59)% | 49(42-56)% | 0.169 |
| OS | MRDneg=557 | MRDpos=208 | 0.36 |
| 3 mo | 96(94-97)% | 96(92-98)% | 0.817 |
| 6 mo | 88(86-91)% | 89(84-92)% | 0.869 |
| 1 yr | 78(74-81)% | 77(70-82)% | 0.728 |
| 2 yr | 67(63-71)% | 64(57-70)% | 0.374 |
| 3 yr | 61(57-65)% | 58(50-66)% | 0.534 |

**Figure S3. Detection of mutations in the blood of AML patients during complete remission.**
**i. Variant summary.** The total number (top graph) and variant allele frequency (VAF, bottom graph) of mutations detected per gene during remission prior to transplant in the peripheral blood of AML patients in (A) cohort #1 (n=454 patients) (B) cohort #2 (n=621 patients), and (C) the full cohort (n=1,075 patients). The genes are ordered by decreasing mutation frequency within the cohort of interest.

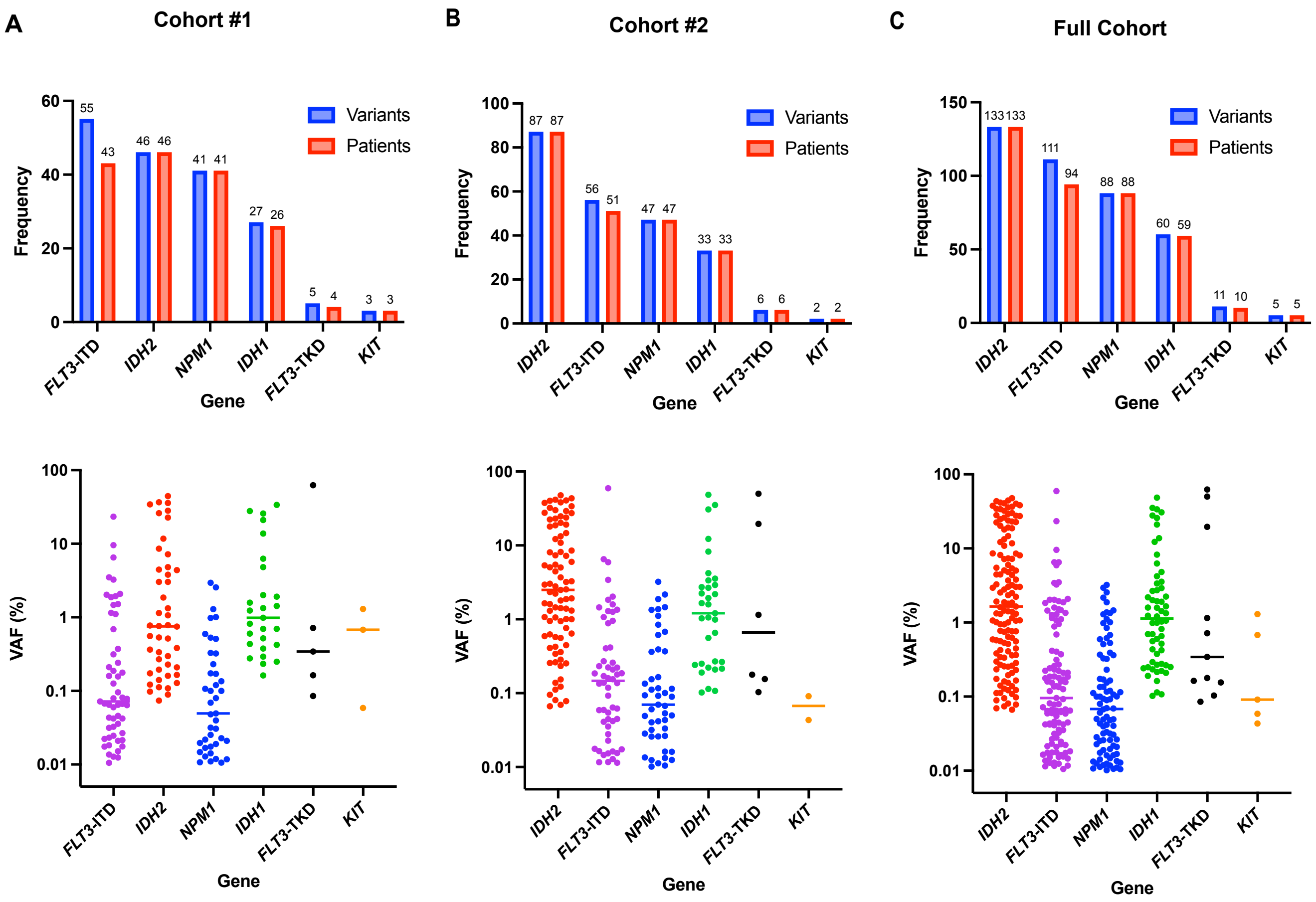

**ii. Orthogonal validation.** A total of 279 variants detected pre-transplant by next-generation sequencing (NGS) were orthogonally validated by digital droplet PCR (ddPCR). The variant allele frequency (VAF) of mutations as determined by NGS were plotted versus ddPCR, with a value of equivalence line displayed as a dotted line. The Pearson correlation coefficient for each comparison is shown on the inset of the graph. A significant correlation was observed in the VAF as detected by NGS and ddPCR with an orthogonal validation rate of (A) 100% in cohort 1, (B) 98.2% in cohort 2, and (C) 99% for the entire cohort.

A

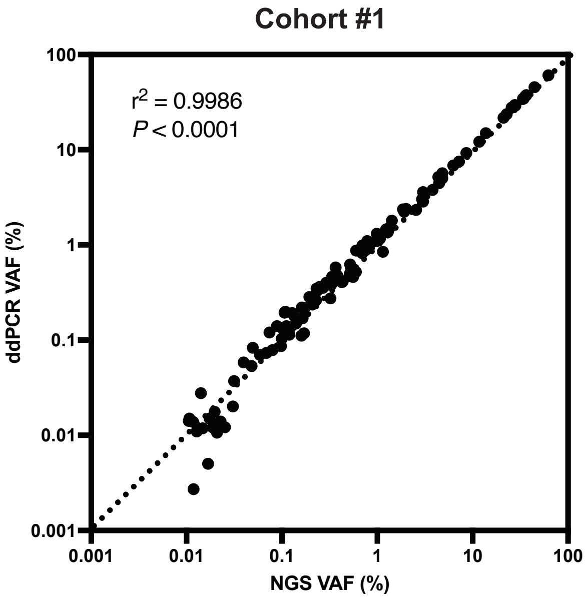


B

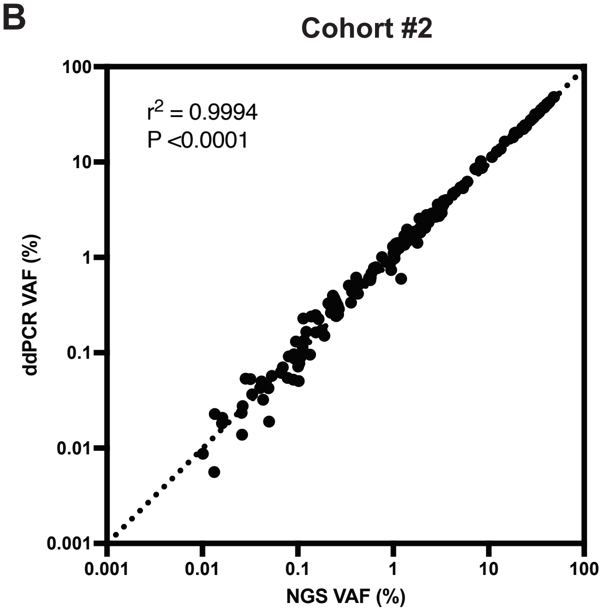


C

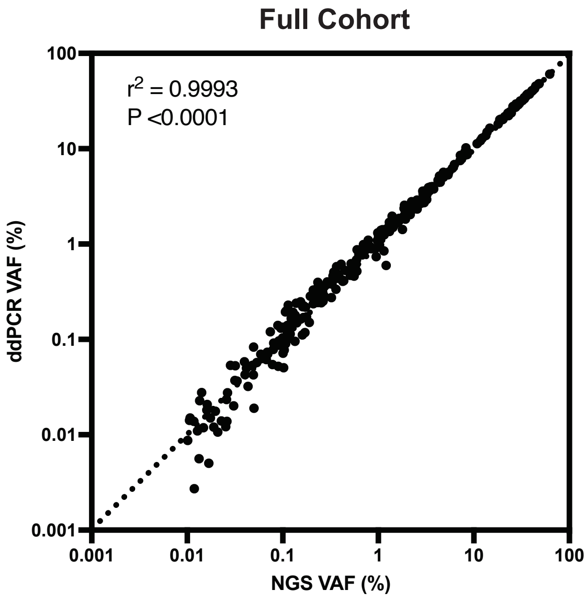

**iii. Mutations by patient.** Heatmap illustrating for each patient (n=1,075 patients) their cohort assignment, conditioning intensity, reported baseline mutation status, and NGS-MRD status. (A) Patients are sorted by NGS-MRD status (positive or negative). (B) Patients are sorted by baseline mutation status (positive, negative, or not tested).

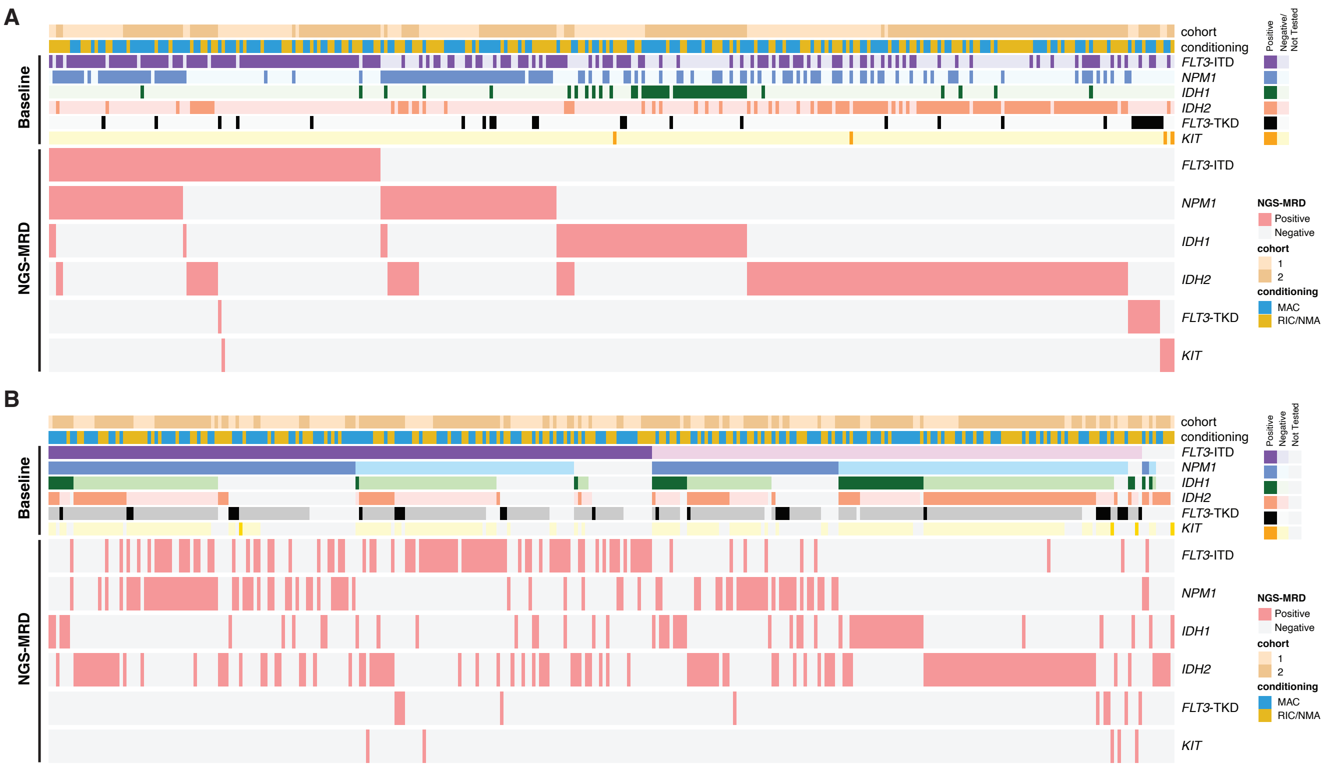

**Figure S4. Association of pre-transplant residual mutations on clinical outcomes.** Rates of non-relapse mortality (NRM, top left), relapse (top right), relapse-free survival (RFS, bottom left), and overall survival (OS, bottom right) censored at 72 months are shown when stratifying by the presence (positive) or absence (negative) of residual mutations detected by NGS pre-transplant for (A) Cohort #1 (n=454), (B) Cohort #2 (n=621), and (C) Combined (n=1,075). Point estimates at different time points are shown in the table (far right). Overall P values: Gray's test for NRM and relapse; log-rank test for RFS and OS. P values for pointwise estimations at different time points: z-test. Confidence interval, CI; Probability, prob; Month, mo; Year, yr.

A

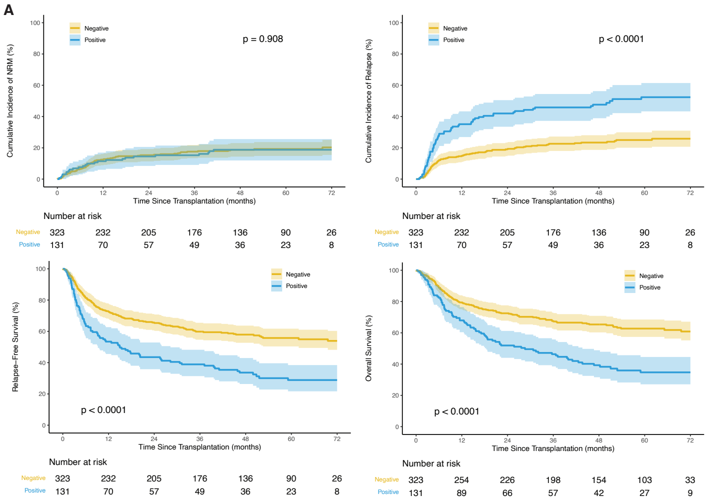


**Cohort #1**

| Outcomes | Prob (95% CI) | Prob (95% CI) | p-value |
|---|---|---|---|
| NRM | Negative=323 | Positive=131 | 0.908 |
| 3 mo | 4(2-6)% | 5(1-9)% | 0.467 |
| 6 mo | 7(4-9)% | 8(3-12)% | 0.681 |
| 1 yr | 12(9-16)% | 11(6-17)% | 0.762 |
| 2 yr | 15(11-19)% | 15(8-21)% | 0.829 |
| 3 yr | 18(13-22)% | 15(9-21)% | 0.546 |
| Relapse | Negative=323 | Positive=131 | <0.001 |
| 3 mo | 4(2-6)% | 11(5-16)% | 0.023 |
| 6 mo | 12(9-16)% | 28(20-36)% | <0.001 |
| 1 yr | 15(11-19)% | 35(27-43)% | <0.001 |
| 2 yr | 19(15-23)% | 42(33-50)% | <0.001 |
| 3 yr | 23(18-27)% | 46(37-54)% | <0.001 |
| RFS | Negative=323 | Positive=131 | <0.001 |
| 3 mo | 92(89-95)% | 84(76-89)% | 0.019 |
| 6 mo | 81(77-85)% | 64(55-72)% | <0.001 |
| 1 yr | 73(67-77)% | 53(45-62)% | <0.001 |
| 2 yr | 66(60-71)% | 44(35-52)% | <0.001 |
| 3 yr | 60(54-65)% | 39(31-47)% | <0.001 |
| OS | Negative=323 | Positive=131 | <0.001 |
| 3 mo | 96(94-98)% | 93(87-96)% | 0.197 |
| 6 mo | 89(85-92)% | 82(75-88)% | 0.09 |
| 1 yr | 79(75-83)% | 68(59-75)% | 0.014 |
| 2 yr | 72(67-77)% | 52(43-60)% | <0.001 |
| 3 yr | 68(62-72)% | 46(38-55)% | <0.001 |

B

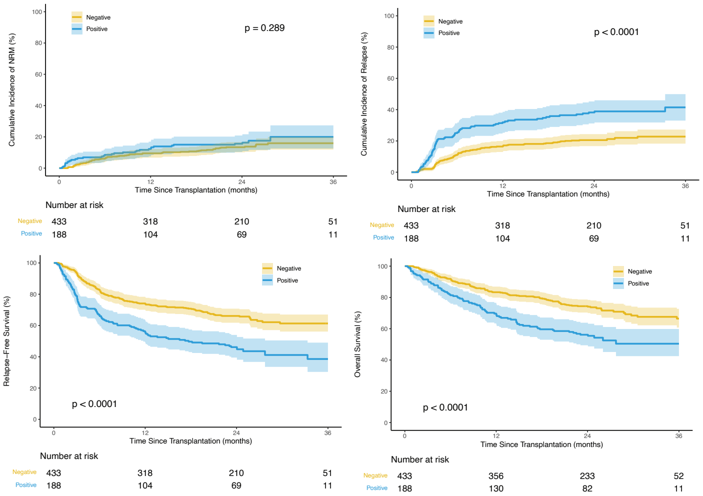


**Cohort #2**

| Outcomes | Prob (95% CI) | Prob (95% CI) | p-value |
|---|---|---|---|
| NRM | Negative=433 | Positive=188 | 0.289 |
| 3 mo | 3(1-5)% | 6(3-10)% | 0.086 |
| 6 mo | 6(4-9)% | 9(5-13)% | 0.333 |
| 1 yr | 9(7-12)% | 12(8-17)% | 0.322 |
| 2 yr | 13(10-17)% | 16(10-21)% | 0.476 |
| 3 yr | 16(12-20)% | 20(13-27)% | 0.347 |
| Relapse | Negative=433 | Positive=188 | <0.001 |
| 3 mo | 3(2-5)% | 15(10-20)% | <0.001 |
| 6 mo | 11(8-14)% | 25(19-31)% | <0.001 |
| 1 yr | 16(13-20)% | 32(25-39)% | <0.001 |
| 2 yr | 21(17-24)% | 38(31-45)% | <0.001 |
| 3 yr | 23(18-27)% | 41(33-50)% | <0.001 |
| RFS | Negative=433 | Positive=188 | <0.001 |
| 3 mo | 94(91-95)% | 79(72-84)% | <0.001 |
| 6 mo | 83(79-86)% | 66(59-73)% | <0.001 |
| 1 yr | 74(70-78)% | 56(48-63)% | <0.001 |
| 2 yr | 66(61-70)% | 46(39-53)% | <0.001 |
| 3 yr | 61(56-66)% | 39(29-48)% | <0.001 |
| OS | Negative=433 | Positive=188 | <0.001 |
| 3 mo | 97(94-98)% | 91(86-95)% | 0.023 |
| 6 mo | 92(89-94)% | 81(75-86)% | 0.001 |
| 1 yr | 83(79-87)% | 70(63-76)% | <0.001 |
| 2 yr | 74(70-78)% | 56(49-63)% | <0.001 |
| 3 yr | 66(60-72)% | 50(41-59)% | 0.003 |

C

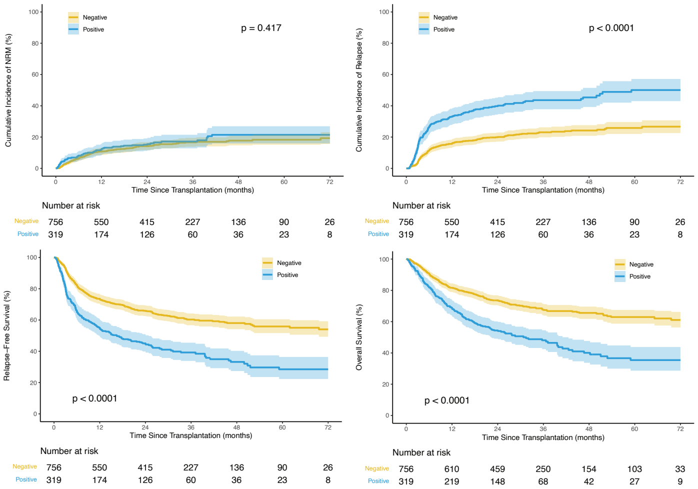


**Combined**

| Outcomes | Prob (95% CI) | Prob (95% CI) | p-value |
|---|---|---|---|
| NRM | Negative=756 | Positive=319 | 0.417 |
| 3 mo | 3(2-5)% | 6(3-9)% | 0.073 |
| 6 mo | 6(5-8)% | 8(5-11)% | 0.312 |
| 1 yr | 11(9-13)% | 12(8-15)% | 0.587 |
| 2 yr | 14(12-17)% | 15(11-19)% | 0.682 |
| 3 yr | 17(14-19)% | 17(13-21)% | 0.85 |
| Relapse | Negative=756 | Positive=319 | <0.001 |
| 3 mo | 4(2-5)% | 13(9-17)% | <0.001 |
| 6 mo | 12(9-14)% | 26(21-31)% | <0.001 |
| 1 yr | 16(13-18)% | 33(28-38)% | <0.001 |
| 2 yr | 20(17-23)% | 40(34-45)% | <0.001 |
| 3 yr | 23(20-26)% | 44(38-49)% | <0.001 |
| RFS | Negative=756 | Positive=319 | <0.001 |
| 3 mo | 93(91-95)% | 81(76-85)% | <0.001 |
| 6 mo | 82(79-85)% | 66(60-70)% | <0.001 |
| 1 yr | 73(70-76)% | 55(49-60)% | <0.001 |
| 2 yr | 66(62-69)% | 45(40-50)% | <0.001 |
| 3 yr | 60(56-64)% | 39(33-45)% | <0.001 |
| OS | Negative=756 | Positive=319 | <0.001 |
| 3 mo | 96(95-98)% | 92(89-95)% | 0.01 |
| 6 mo | 90(88-92)% | 82(77-86)% | <0.001 |
| 1 yr | 82(79-84)% | 69(64-74)% | <0.001 |
| 2 yr | 74(70-77)% | 54(49-60)% | <0.001 |
| 3 yr | 68(64-71)% | 48(42-54)% | <0.001 |

**Figure S5. Univariate cox regression for relapse.** (A) Cohort #1 (n=454), (B) Cohort #2 (n=621), and (C) Combined (n=1,075). Baseline variable by group: Age: < 60 yrs, Sex: female, Conditioning: myeloablative, Graft Type: peripheral blood, Donor Group: HLA-identical sibling, HCT-CI: 0, Karnofsky Score: < 90, ATG: No, Race: Caucasian, ELN: favorable, AML Group: *De novo*, Baseline gene mutation: negative; Site MRD: negative; NGS-MRD by gene: negative. Non-myeloablative, NMA; reduced intensity conditioning, RIC; melphalan, Mel; hematopoietic cell transplantation comorbidity index, HCT-CI; anti-thymocyte globulin ATG; European Leukemia Network, ELN; measurable residual disease, MRD; next-generation sequencing, NGS.

**A**

**Univariate Cox Regression for Relapse**

| | Hazard Ratio | P value |
|---|---|---|
| Age: ≥60 | 1.27 (0.92–1.77) | 0.15 |
| Sex: Male | 0.92 (0.66–1.28) | 0.616 |
| Conditioning: RIC with Mel | 1.36 (0.86–2.13) | 0.185 |
| Conditioning: RIC, Other | 2.33 (1.53–3.56) | <0.001 |
| Conditioning: NMA | 2.19 (1.33–3.59) | 0.002 |
| Graft Type: Cord Blood | 0.96 (0.64–1.43) | 0.833 |
| Graft Type: Bone Marrow | 0.68 (0.4–1.15) | 0.147 |
| Donor Group: Matched Unrelated | 0.64 (0.41–0.99) | 0.044 |
| Donor Group: Haploidentical Related | 0.71 (0.38–1.35) | 0.299 |
| Donor Group: Cord Blood | 0.74 (0.45–1.23) | 0.241 |
| Donor Group: Mismatched | 1.02 (0.31–3.34) | 0.98 |
| HCT–CI: 1–2 | 0.89 (0.54–1.45) | 0.634 |
| HCT–CI: ≥3 | 1 (0.62–1.59) | 0.984 |
| Karnofsky Score: ≥90 | 1.08 (0.77–1.5) | 0.667 |
| ATG: Yes | 1.08 (0.75–1.55) | 0.694 |
| Race: Other | 0.84 (0.53–1.34) | 0.467 |
| ELN: Adverse | 1.15 (0.71–1.86) | 0.576 |
| ELN: Intermediate | 1.17 (0.74–1.85) | 0.489 |
| AML Group: Not *de novo* | 1.4 (0.92–2.13) | 0.115 |
| Baseline *FLT3*–ITD: Positive | 1.05 (0.75–1.47) | 0.79 |
| Baseline *NPM1*: Positive | 1.13 (0.81–1.57) | 0.46 |
| Baseline *FLT3*–TKD: Positive | 0.72 (0.43–1.22) | 0.224 |
| Baseline *KIT*: Positive | 0.98 (0.48–2) | 0.951 |
| Baseline *IDH1*: Positive | 0.88 (0.43–1.79) | 0.715 |
| Baseline *IDH2*: Positive | 1.15 (0.7–1.89) | 0.572 |
| Site MRD: Positive | 1.44 (0.97–2.14) | 0.069 |
| NGS MRD: *FLT3*–ITD Positive | 3.13 (2.06–4.76) | <0.001 |
| NGS MRD: *NPM1* Positive | 4.46 (2.98–6.66) | <0.001 |
| NGS MRD: *FLT3*–TKD Positive | 1.73 (0.43–6.99) | 0.441 |
| NGS MRD: *KIT* Positive | 0 (0–Inf) | 0.994 |
| NGS MRD: *IDH1* Positive | 1.29 (0.68–2.45) | 0.439 |
| NGS MRD: *IDH2* Positive | 1.36 (0.83–2.23) | 0.22 |

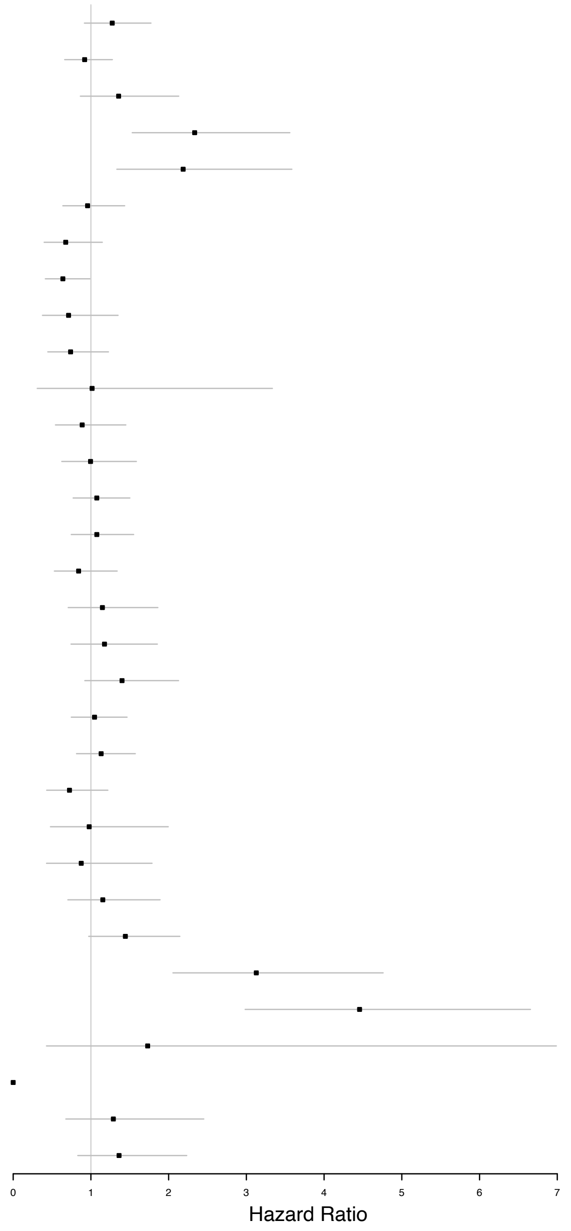

**B**

#### Univariate Cox Regression for Relapse

| | Hazard Ratio | P value |
|---|---|---|
| Age: ≥60 | 1.03 (0.76–1.4) | 0.841 |
| Sex: Male | 1.12 (0.82–1.52) | 0.469 |
| Conditioning: RIC with Mel | 0.83 (0.54–1.29) | 0.413 |
| Conditioning: RIC, Other | 1.08 (0.7–1.65) | 0.732 |
| Conditioning: NMA | 1.49 (0.9–2.46) | 0.124 |
| Graft Type: Cord Blood | 1.26 (0.66–2.41) | 0.476 |
| Graft Type: Bone Marrow | 1.27 (0.86–1.89) | 0.231 |
| Donor Group: Matched Unrelated | 0.68 (0.43–1.08) | 0.101 |
| Donor Group: Haploidentical Related | 1.14 (0.62–2.12) | 0.67 |
| Donor Group: Cord Blood | 0.82 (0.37–1.84) | 0.636 |
| Donor Group: Mismatched | 0.87 (0.47–1.61) | 0.658 |
| HCT–CI: 1–2 | 1.26 (0.81–1.97) | 0.302 |
| HCT–CI: ≥3 | 1.17 (0.78–1.76) | 0.451 |
| Karnofsky Score: ≥90 | 1.11 (0.81–1.53) | 0.506 |
| ATG: Yes | 0.81 (0.56–1.18) | 0.279 |
| Race: Other | 1.51 (0.98–2.34) | 0.061 |
| ELN: Adverse | 1.95 (1.24–3.08) | 0.004 |
| ELN: Intermediate | 1.82 (1.16–2.84) | 0.009 |
| AML Group: Not *de novo* | 1.52 (0.98–2.34) | 0.059 |
| Baseline *FLT3*–ITD: Positive | 1.59 (1.16–2.18) | 0.004 |
| Baseline *NPM1*: Positive | 0.82 (0.6–1.12) | 0.213 |
| Baseline *FLT3*–TKD: Positive | 0.88 (0.54–1.42) | 0.595 |
| Baseline *KIT*: Positive | 0.8 (0.39–1.62) | 0.528 |
| Baseline *IDH1*: Positive | 1.09 (0.7–1.69) | 0.701 |
| Baseline *IDH2*: Positive | 0.68 (0.46–1.02) | 0.059 |
| Site MRD: Positive | 1.78 (1.29–2.45) | <0.001 |
| NGS MRD: *FLT3*–ITD Positive | 5.7 (3.94–8.24) | <0.001 |
| NGS MRD: *NPM1* Positive | 3.41 (2.24–5.19) | <0.001 |
| NGS MRD: *FLT3*–TKD Positive | 2.77 (0.88–8.7) | 0.08 |
| NGS MRD: *KIT* Positive | 4.38 (1.08–17.7) | 0.038 |
| NGS MRD: *IDH1* Positive | 1.14 (0.6–2.16) | 0.688 |
| NGS MRD: *IDH2* Positive | 0.97 (0.61–1.53) | 0.888 |

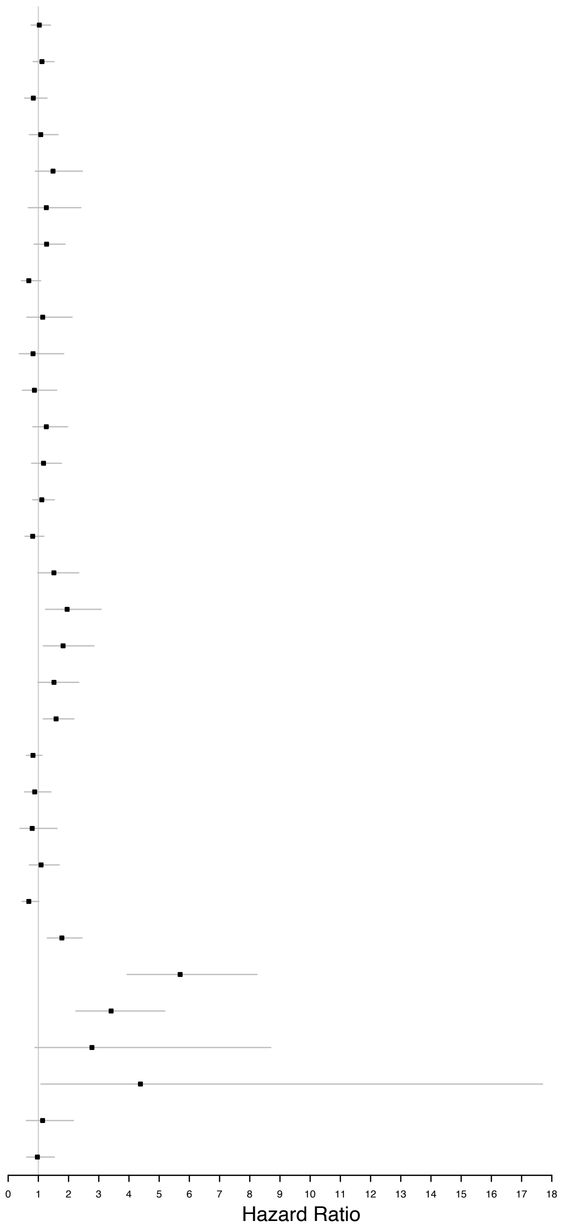

**C**

##### Univariate Cox Regression for Relapse

| | Hazard Ratio | P value |
|---|---|---|
| Age: ≥60 | 1.14 (0.91–1.42) | 0.262 |
| Sex: Male | 1.02 (0.81–1.27) | 0.871 |
| Conditioning: RIC with Mel | 1.05 (0.77–1.43) | 0.771 |
| Conditioning: RIC, Other | 1.55 (1.15–2.09) | 0.004 |
| Conditioning: NMA | 1.77 (1.25–2.51) | 0.001 |
| Graft Type: Cord Blood | 1.06 (0.76–1.48) | 0.723 |
| Graft Type: Bone Marrow | 0.98 (0.72–1.34) | 0.899 |
| Donor Group: Matched Unrelated | 0.66 (0.48–0.9) | 0.009 |
| Donor Group: Haploidentical Related | 0.91 (0.59–1.42) | 0.69 |
| Donor Group: Cord Blood | 0.77 (0.51–1.17) | 0.22 |
| Donor Group: Mismatched | 0.86 (0.52–1.43) | 0.563 |
| HCT–CI: 1–2 | 1.08 (0.77–1.49) | 0.661 |
| HCT–CI: ≥3 | 1.1 (0.81–1.49) | 0.557 |
| Karnofsky Score: ≥90 | 1.09 (0.87–1.38) | 0.443 |
| ATG: Yes | 0.94 (0.72–1.21) | 0.617 |
| Race: Other | 1.11 (0.81–1.53) | 0.508 |
| ELN: Adverse | 1.54 (1.11–2.15) | 0.011 |
| ELN: Intermediate | 1.49 (1.09–2.06) | 0.014 |
| AML Group: Not *de novo* | 1.46 (1.08–1.97) | 0.014 |
| Baseline *FLT3*–ITD: Positive | 1.31 (1.04–1.65) | 0.021 |
| Baseline *NPM1*: Positive | 0.96 (0.76–1.2) | 0.693 |
| Baseline *FLT3*–TKD: Positive | 0.81 (0.57–1.15) | 0.231 |
| Baseline *KIT*: Positive | 0.89 (0.54–1.46) | 0.636 |
| Baseline *IDH1*: Positive | 1.02 (0.7–1.47) | 0.927 |
| Baseline *IDH2*: Positive | 0.83 (0.61–1.12) | 0.224 |
| Site MRD: Positive | 1.63 (1.28–2.08) | <0.001 |
| NGS MRD: *FLT3*–ITD Positive | 4.25 (3.22–5.6) | <0.001 |
| NGS MRD: *NPM1* Positive | 3.9 (2.92–5.21) | <0.001 |
| NGS MRD: *FLT3*–TKD Positive | 2.26 (0.93–5.47) | 0.071 |
| NGS MRD: *KIT* Positive | 1.66 (0.41–6.67) | 0.476 |
| NGS MRD: *IDH1* Positive | 1.21 (0.77–1.9) | 0.413 |
| NGS MRD: *IDH2* Positive | 1.12 (0.8–1.57) | 0.496 |

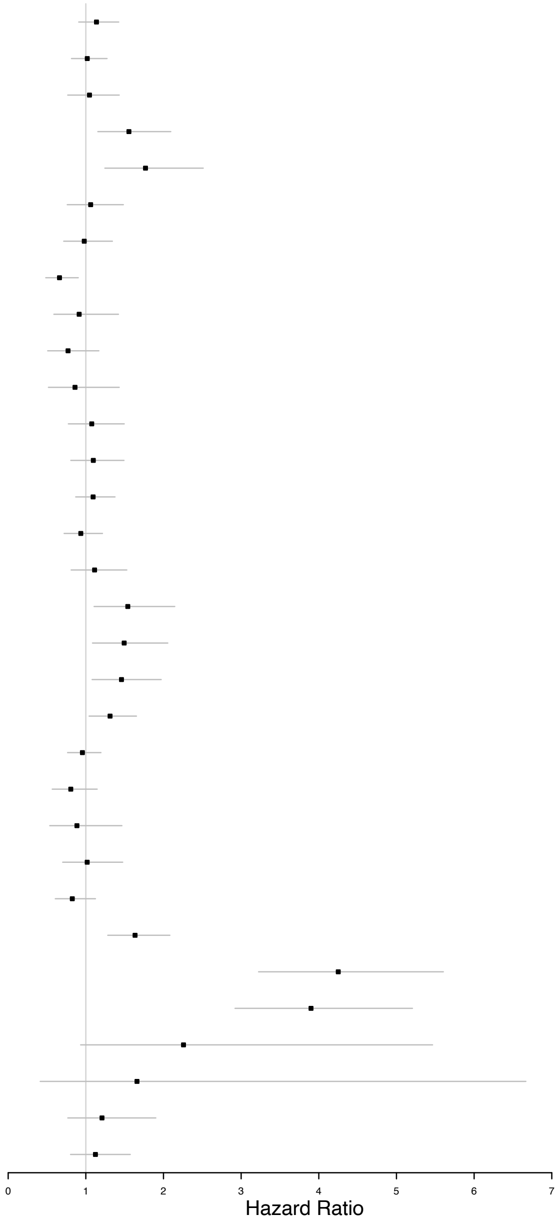

**Figure S6. Association of NGS-MRD status for *NPM1* and/or *FLT3*-ITD on clinical outcomes.**
**i. Agnostic to baseline.** Rates of non-relapse mortality (NRM, top left), relapse (top right), relapse-free survival (RFS, bottom left), and overall survival (OS, bottom right) censored at 72 months are shown by pre-transplant NGS-MRD status for *NPM1* and/or *FLT3*-ITD regardless of baseline genetics for (A) Cohort #1 (n=454), (B) Cohort #2 (n=621), and (C) Combined (n=1,075). Point estimates at different time points are shown in the table (far right). Overall P values: Gray's test for NRM and relapse; log-rank test for RFS and OS. P values for pointwise estimations at different time points: z-test. Confidence interval, CI; Probability, prob; Month, mo; Year, yr; positive, pos; negative, neg.

A

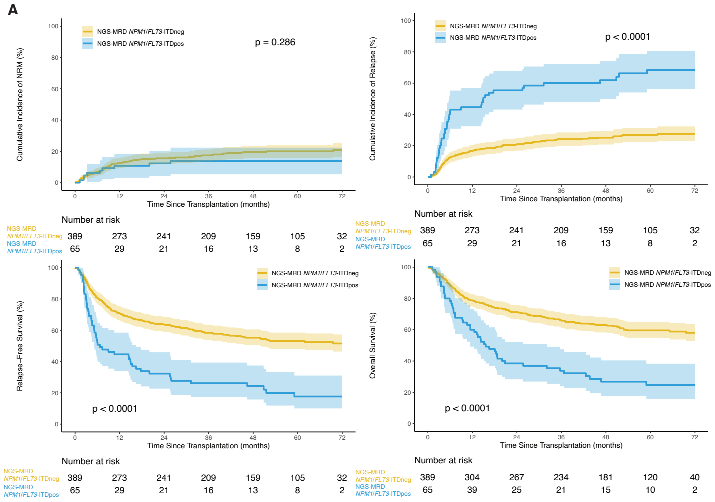


**Cohort #1**

| Outcomes | Prob (95% CI) | Prob (95% CI) | p-value |
|---|---|---|---|
| NRM | *NPM1*/*FLT3*-ITDneg=389 | *NPM1*/*FLT3*-ITDpos=65 | 0.286 |
| 3 mo | 4(2-6)% | 5(0-10)% | 0.858 |
| 6 mo | 7(4-9)% | 6(0-12)% | 0.806 |
| 1 yr | 12(9-16)% | 11(3-18)% | 0.7 |
| 2 yr | 16(12-19)% | 12(4-20)% | 0.476 |
| 3 yr | 17(14-21)% | 14(5-22)% | 0.454 |
| Relapse | *NPM1*/*FLT3*-ITDneg=389 | *NPM1*/*FLT3*-ITDpos=65 | <0.001 |
| 3 mo | 4(2-6)% | 15(7-24)% | 0.017 |
| 6 mo | 13(9-16)% | 42(29-54)% | <0.001 |
| 1 yr | 17(13-21)% | 45(32-57)% | <0.001 |
| 2 yr | 21(17-25)% | 55(43-68)% | <0.001 |
| 3 yr | 24(20-28)% | 60(48-72)% | <0.001 |
| RFS | *NPM1*/*FLT3*-ITDneg=389 | *NPM1*/*FLT3*-ITDpos=65 | <0.001 |
| 3 mo | 92(88-94)% | 80(68-88)% | 0.026 |
| 6 mo | 80(76-84)% | 52(40-64)% | <0.001 |
| 1 yr | 71(66-75)% | 45(32-56)% | <0.001 |
| 2 yr | 64(59-68)% | 32(21-44)% | <0.001 |
| 3 yr | 58(53-63)% | 26(16-37)% | <0.001 |
| OS | *NPM1*/*FLT3*-ITDneg=389 | *NPM1*/*FLT3*-ITDpos=65 | <0.001 |
| 3 mo | 96(93-97)% | 94(84-98)% | 0.572 |
| 6 mo | 88(85-91)% | 78(66-87)% | 0.063 |
| 1 yr | 79(74-83)% | 60(47-71)% | 0.003 |
| 2 yr | 71(66-75)% | 38(27-50)% | <0.001 |
| 3 yr | 66(61-71)% | 34(23-45)% | <0.001 |

B

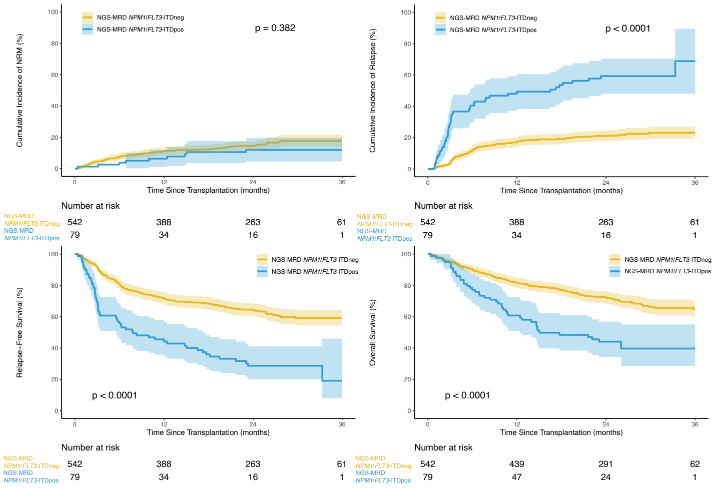


**Cohort #2**

| Outcomes | Prob (95% CI) | Prob (95% CI) | p-value |
|---|---|---|---|
| NRM | *NPM1*/*FLT3*-ITDneg=542 | *NPM1*/*FLT3*-ITDpos=79 | 0.382 |
| 3 mo | 4(3-6)% | 1(0-4)% | 0.041 |
| 6 mo | 7(5-10)% | 4(0-8)% | 0.142 |
| 1 yr | 11(8-14)% | 6(1-12)% | 0.138 |
| 2 yr | 14(11-17)% | 12(5-19)% | 0.56 |
| 3 yr | 18(14-22)% | 12(5-19)% | 0.18 |
| Relapse | *NPM1*/*FLT3*-ITDneg=542 | *NPM1*/*FLT3*-ITDpos=79 | <0.001 |
| 3 mo | 4(2-5)% | 29(19-39)% | <0.001 |
| 6 mo | 12(9-14)% | 41(30-51)% | <0.001 |
| 1 yr | 17(14-20)% | 49(38-61)% | <0.001 |
| 2 yr | 21(18-24)% | 59(48-70)% | <0.001 |
| 3 yr | 23(19-27)% | 69(48-90)% | <0.001 |
| RFS | *NPM1*/*FLT3*-ITDneg=542 | *NPM1*/*FLT3*-ITDpos=79 | <0.001 |
| 3 mo | 92(89-94)% | 70(58-78)% | <0.001 |
| 6 mo | 81(77-84)% | 56(44-66)% | <0.001 |
| 1 yr | 72(68-76)% | 44(33-55)% | <0.001 |
| 2 yr | 65(60-68)% | 29(19-39)% | <0.001 |
| 3 yr | 59(54-64)% | 19(6-38)% | <0.001 |
| OS | *NPM1*/*FLT3*-ITDneg=542 | *NPM1*/*FLT3*-ITDpos=79 | <0.001 |
| 3 mo | 95(93-97)% | 95(87-98)% | 0.975 |
| 6 mo | 90(87-92)% | 77(66-85)% | 0.008 |
| 1 yr | 82(78-85)% | 61(49-71)% | <0.001 |
| 2 yr | 72(68-76)% | 44(33-55)% | <0.001 |
| 3 yr | 65(59-70)% | 40(27-52)% | <0.001 |

C

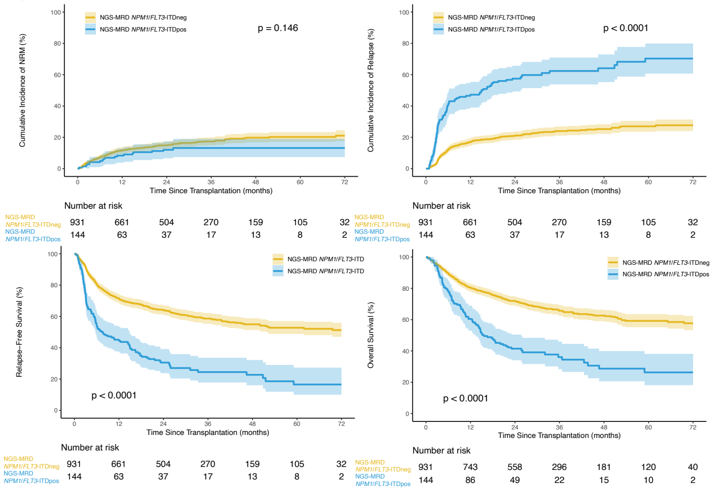


**Combined**

| Outcomes | Prob (95% CI) | Prob (95% CI) | p-value |
|---|---|---|---|
| NRM | *NPM1*/*FLT3*-ITDneg=931 | NPM1/*FLT3*-ITDpos=144 | 0.146 |
| 3 mo | 4(3-6)% | 3(0-5)% | 0.32 |
| 6 mo | 7(6-9)% | 5(1-8)% | 0.239 |
| 1 yr | 12(9-14)% | 8(4-13)% | 0.21 |
| 2 yr | 15(13-17)% | 12(7-17)% | 0.342 |
| 3 yr | 17(15-20)% | 13(7-19)% | 0.198 |
| Relapse | *NPM1*/*FLT3*-ITDneg=931 | NPM1/*FLT3*-ITDpos=144 | <0.001 |
| 3 mo | 4(3-5)% | 23(16-30)% | <0.001 |
| 6 mo | 12(10-14)% | 41(33-49)% | <0.001 |
| 1 yr | 17(14-19)% | 47(39-55)% | <0.001 |
| 2 yr | 21(18-24)% | 57(49-66)% | <0.001 |
| 3 yr | 24(21-27)% | 62(54-71)% | <0.001 |
| RFS | *NPM1*/*FLT3*-ITDneg=931 | NPM1/*FLT3*-ITDpos=144 | <0.001 |
| 3 mo | 92(90-93)% | 74(66-81)% | <0.001 |
| 6 mo | 81(78-83)% | 54(46-62)% | <0.001 |
| 1 yr | 72(69-74)% | 44(36-52)% | <0.001 |
| 2 yr | 64(61-67)% | 31(23-38)% | <0.001 |
| 3 yr | 59(55-62)% | 24(17-32)% | <0.001 |
| OS | *NPM1*/*FLT3*-ITDneg=931 | NPM1/*FLT3*-ITDpos=144 | <0.001 |
| 3 mo | 95(94-96)% | 94(89-97)% | 0.683 |
| 6 mo | 89(87-91)% | 78(70-84)% | 0.001 |
| 1 yr | 81(78-83)% | 60(52-68)% | <0.001 |
| 2 yr | 72(69-75)% | 41(33-49)% | <0.001 |
| 3 yr | 66(62-69)% | 36(28-45)% | <0.001 |

**ii. Baseline informed.** Rates of non-relapse mortality (NRM, top left), relapse (top right), relapse-free survival (RFS, bottom left), and overall survival (OS, bottom right) censored at 72 months are shown by pre-transplant NGS-MRD status for *NPM1* and/or *FLT3*-ITD based on baseline genetics for (A) Cohort #1 (n=371), (B) Cohort #2 (n=451), and (C) Combined (n=822). Point estimates at different time points are shown in the table (far right). Overall P values: Gray's test for NRM and relapse; log-rank test for RFS and OS. P values for pointwise estimations at different time points: z-test. Confidence interval, CI; Probability, prob; Month, mo; Year, yr; positive, pos; negative, neg.

A

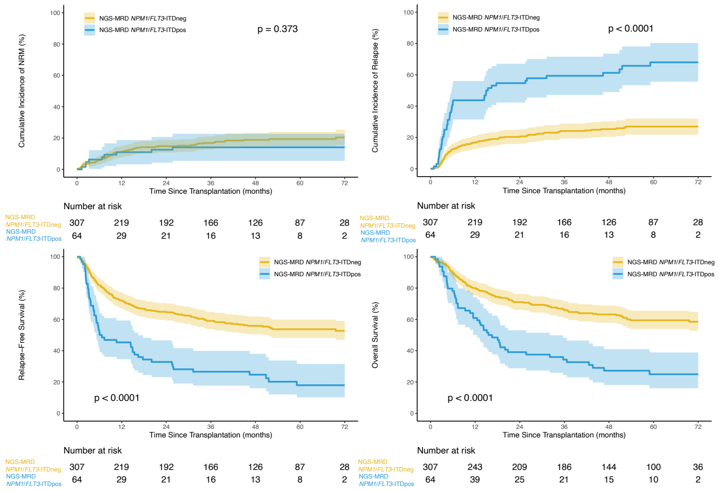


**Cohort #1**

| Outcomes | Prob (95% CI) | Prob (95% CI) | p-value |
|---|---|---|---|
| NRM | *NPM1/FLT3*-ITDneg=307 | *NPM1/FLT3*-ITDpos=64 | 0.373 |
| 3 mo | 4(2-6)% | 5(0-10)% | 0.7 |
| 6 mo | 6(3-9)% | 6(0-12)% | 0.911 |
| 1 yr | 11(8-15)% | 11(3-19)% | 0.961 |
| 2 yr | 15(11-19)% | 12(4-21)% | 0.622 |
| 3 yr | 17(13-21)% | 14(5-23)% | 0.569 |
| Relapse | *NPM1/FLT3*-ITDneg=307 | *NPM1/FLT3*-ITDpos=64 | <0.001 |
| 3 mo | 4(2-6)% | 16(7-25)% | 0.016 |
| 6 mo | 12(9-16)% | 42(30-54)% | <0.001 |
| 1 yr | 17(13-21)% | 44(31-56)% | <0.001 |
| 2 yr | 20(16-25)% | 55(42-67)% | <0.001 |
| 3 yr | 24(19-29)% | 59(47-72)% | <0.001 |
| RFS | *NPM1/FLT3*-ITDneg=307 | *NPM1/FLT3*-ITDpos=64 | <0.001 |
| 3 mo | 92(89-95)% | 80(68-88)% | 0.017 |
| 6 mo | 82(77-86)% | 52(39-63)% | <0.001 |
| 1 yr | 72(67-77)% | 45(33-57)% | <0.001 |
| 2 yr | 65(59-70)% | 33(22-44)% | <0.001 |
| 3 yr | 59(53-64)% | 27(16-38)% | <0.001 |
| OS | *NPM1/FLT3*-ITDneg=307 | *NPM1/FLT3*-ITDpos=64 | <0.001 |
| 3 mo | 96(94-98)% | 94(84-98)% | 0.406 |
| 6 mo | 90(86-92)% | 78(66-86)% | 0.037 |
| 1 yr | 80(75-84)% | 61(48-72)% | 0.003 |
| 2 yr | 71(66-76)% | 39(27-51)% | <0.001 |
| 3 yr | 66(61-71)% | 34(23-46)% | <0.001 |

B

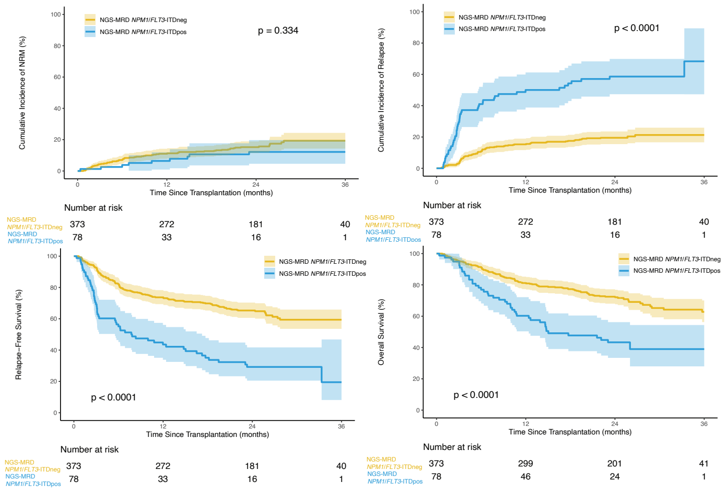


**Cohort #2**

| Outcomes | Prob (95% CI) | Prob (95% CI) | p-value |
|---|---|---|---|
| NRM | *NPM1/FLT3*-ITDneg=373 | *NPM1/FLT3*-ITDpos=78 | 0.334 |
| 3 mo | 4(2-6)% | 1(0-4)% | 0.07 |
| 6 mo | 7(5-10)% | 4(0-8)% | 0.187 |
| 1 yr | 11(8-14)% | 6(1-12)% | 0.156 |
| 2 yr | 15(12-19)% | 12(5-20)% | 0.477 |
| 3 yr | 19(14-24)% | 12(5-20)% | 0.123 |
| Relapse | *NPM1/FLT3*-ITDneg=373 | *NPM1/FLT3*-ITDpos=78 | <0.001 |
| 3 mo | 3(1-5)% | 29(19-40)% | <0.001 |
| 6 mo | 11(8-14)% | 41(30-52)% | <0.001 |
| 1 yr | 15(12-19)% | 50(39-61)% | <0.001 |
| 2 yr | 19(15-24)% | 59(47-70)% | <0.001 |
| 3 yr | 21(17-26)% | 68(47-89)% | <0.001 |
| RFS | *NPM1/FLT3*-ITDneg=373 | *NPM1/FLT3*-ITDpos=78 | <0.001 |
| 3 mo | 92(89-95)% | 69(58-78)% | <0.001 |
| 6 mo | 82(78-86)% | 55(43-65)% | <0.001 |
| 1 yr | 74(69-78)% | 44(32-54)% | <0.001 |
| 2 yr | 65(60-70)% | 29(19-40)% | <0.001 |
| 3 yr | 59(53-65)% | 19(6-38)% | <0.001 |
| OS | *NPM1/FLT3*-ITDneg=373 | *NPM1/FLT3*-ITDpos=78 | <0.001 |
| 3 mo | 95(92-97)% | 95(87-98)% | 0.912 |
| 6 mo | 91(88-94)% | 77(66-85)% | 0.004 |
| 1 yr | 81(77-85)% | 60(49-70)% | <0.001 |
| 2 yr | 72(68-77)% | 43(32-54)% | <0.001 |
| 3 yr | 63(55-69)% | 39(26-52)% | 0.001 |

C

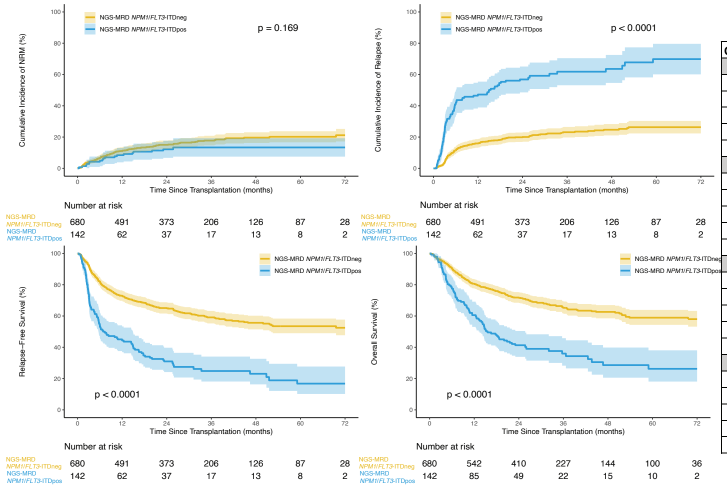


**Combined**

| Outcomes | Prob (95% CI) | Prob (95% CI) | p-value |
|---|---|---|---|
| NRM | *NPM1/FLT3*-ITDneg=680 | *NPM1/FLT3*-ITDpos=142 | 0.169 |
| 3 mo | 4(3-5)% | 3(0-6)% | 0.466 |
| 6 mo | 7(5-8)% | 5(1-9)% | 0.41 |
| 1 yr | 11(9-13)% | 8(4-13)% | 0.321 |
| 2 yr | 15(12-18)% | 12(7-18)% | 0.361 |
| 3 yr | 18(15-21)% | 13(7-19)% | 0.195 |
| Relapse | *NPM1/FLT3*-ITDneg=680 | *NPM1/FLT3*-ITDpos=142 | <0.001 |
| 3 mo | 4(2-5)% | 23(16-30)% | <0.001 |
| 6 mo | 11(9-14)% | 42(33-50)% | <0.001 |
| 1 yr | 16(13-19)% | 47(39-55)% | <0.001 |
| 2 yr | 20(17-23)% | 57(49-65)% | <0.001 |
| 3 yr | 23(20-26)% | 62(53-70)% | <0.001 |
| RFS | *NPM1/FLT3*-ITDneg=680 | *NPM1/FLT3*-ITDpos=142 | <0.001 |
| 3 mo | 92(90-94)% | 74(66-80)% | <0.001 |
| 6 mo | 82(79-85)% | 54(45-61)% | <0.001 |
| 1 yr | 73(69-76)% | 44(36-52)% | <0.001 |
| 2 yr | 65(61-69)% | 31(23-39)% | <0.001 |
| 3 yr | 59(55-63)% | 25(17-33)% | <0.001 |
| OS | *NPM1/FLT3*-ITDneg=680 | *NPM1/FLT3*-ITDpos=142 | <0.001 |
| 3 mo | 96(94-97)% | 94(89-97)% | 0.511 |
| 6 mo | 90(88-92)% | 77(70-83)% | <0.001 |
| 1 yr | 81(77-83)% | 61(52-68)% | <0.001 |
| 2 yr | 72(68-75)% | 41(33-49)% | <0.001 |
| 3 yr | 66(61-69)% | 36(27-45)% | <0.001 |

**Figure S7. Association of site MRD and NGS-MRD status on clinical outcomes for patients with baseline *FLT3*-ITD and/or *NPM1* mutations.**

**i. *FLT3*-ITD mutated, *NPM1* mutated at baseline.** Relapse (left) and overall survival (right) censored at 72 months are shown for patients (A) in the defined cohort, (B) with site MRD status, and (C) with NGS-MRD status. Overall P values: Gray's test for relapse; log-rank test for overall survival (OS). Positive, pos; negative, neg

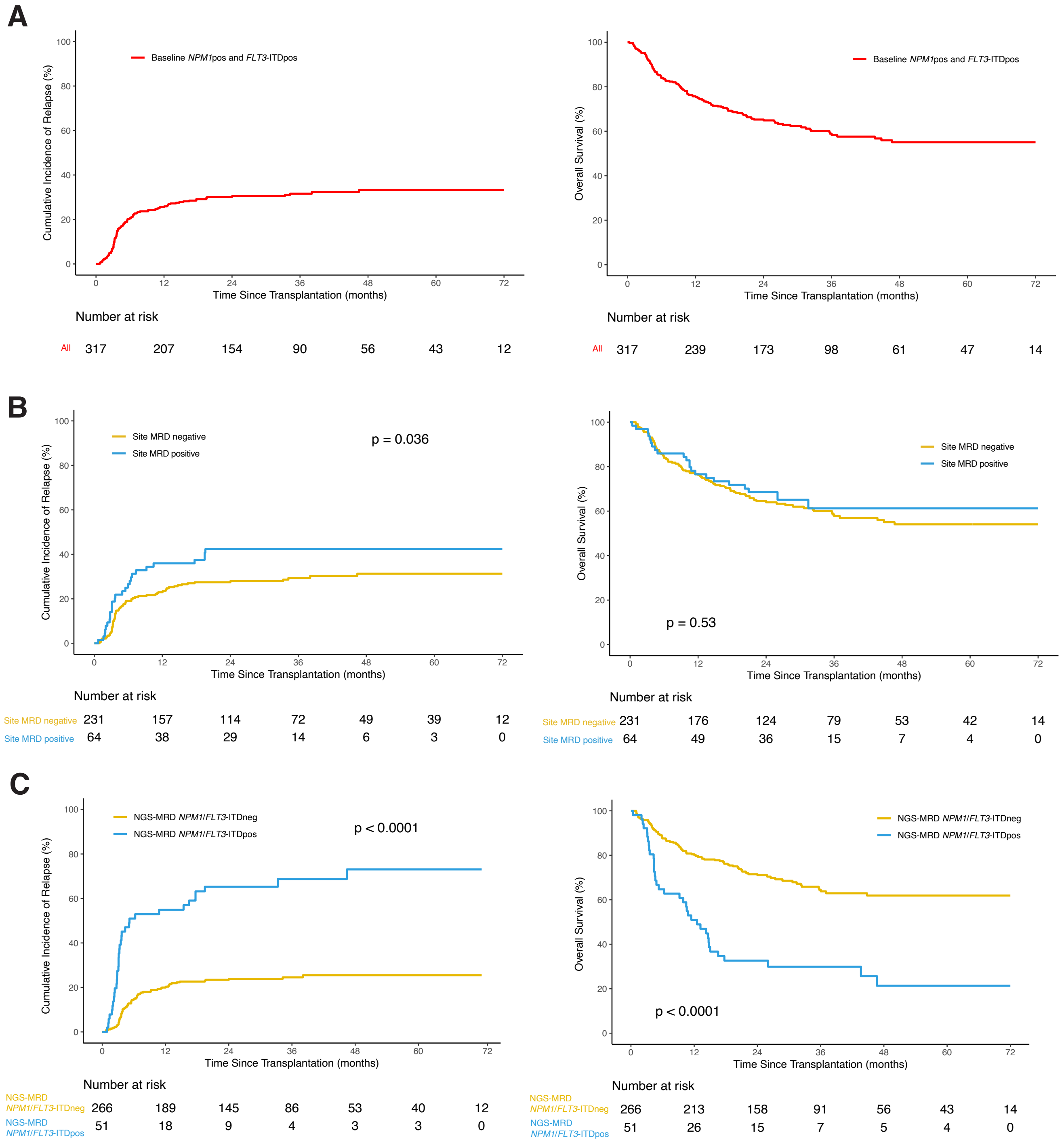

**ii. *FLT3*-ITD mutated, *NPM1* wild-type at baseline.** Relapse (left) and overall survival (right) censored at 72 months are shown for patients (A) in the defined cohort, (B) with site MRD status, and (C) with NGS-MRD status. Overall P values: Gray's test for relapse; log-rank test for overall survival (OS). Positive, pos; negative, neg.

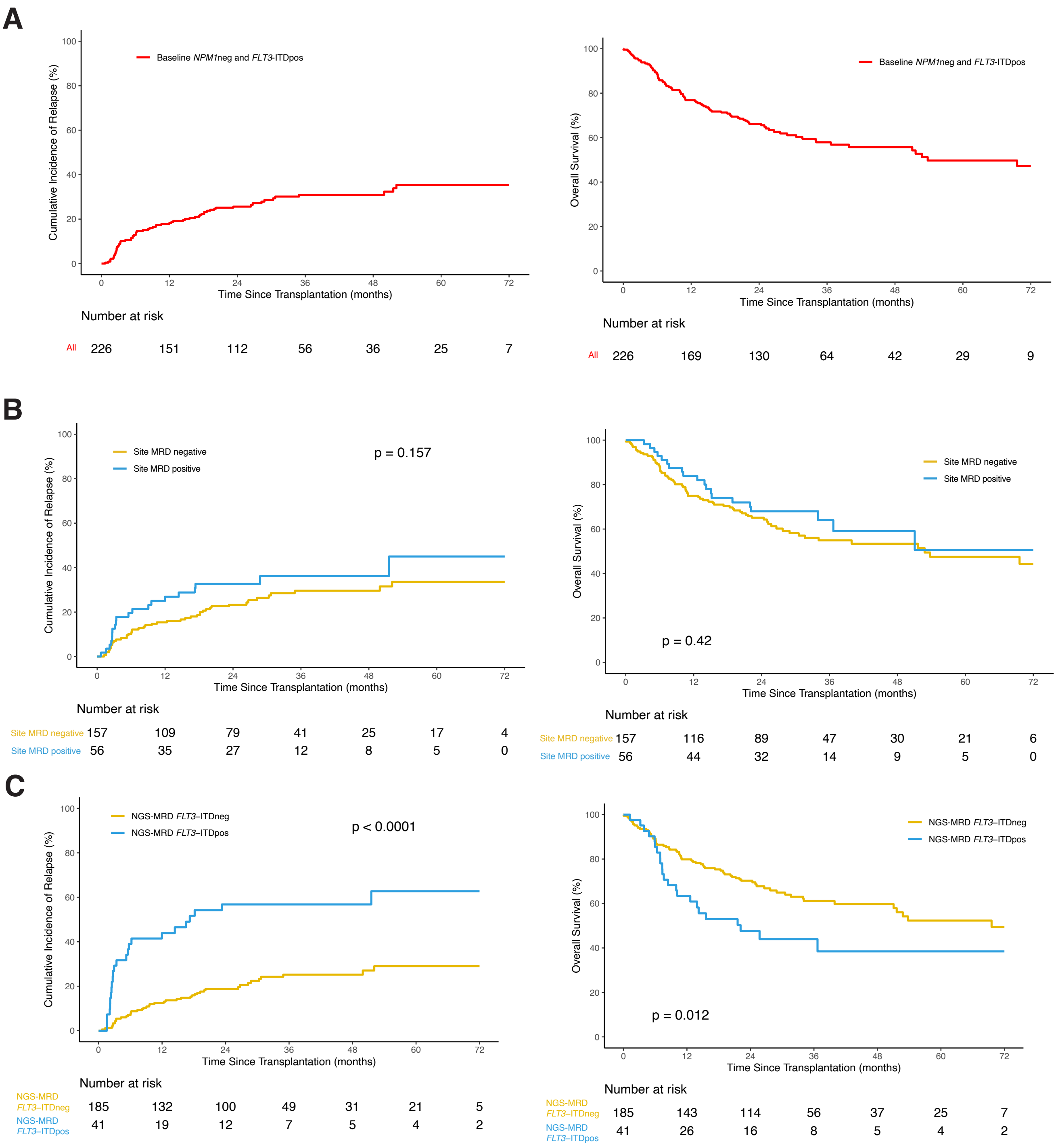

**iii. *FLT3*-ITD wild-type, *NPM1* mutated at baseline.** Relapse (left) and overall survival (right) censored at 72 months are shown for patients (A) in the defined cohort, (B) with site MRD status, and (C) with NGS-MRD status. Overall P values: Gray's test for relapse; log-rank test for overall survival (OS). Positive, pos; negative, neg.

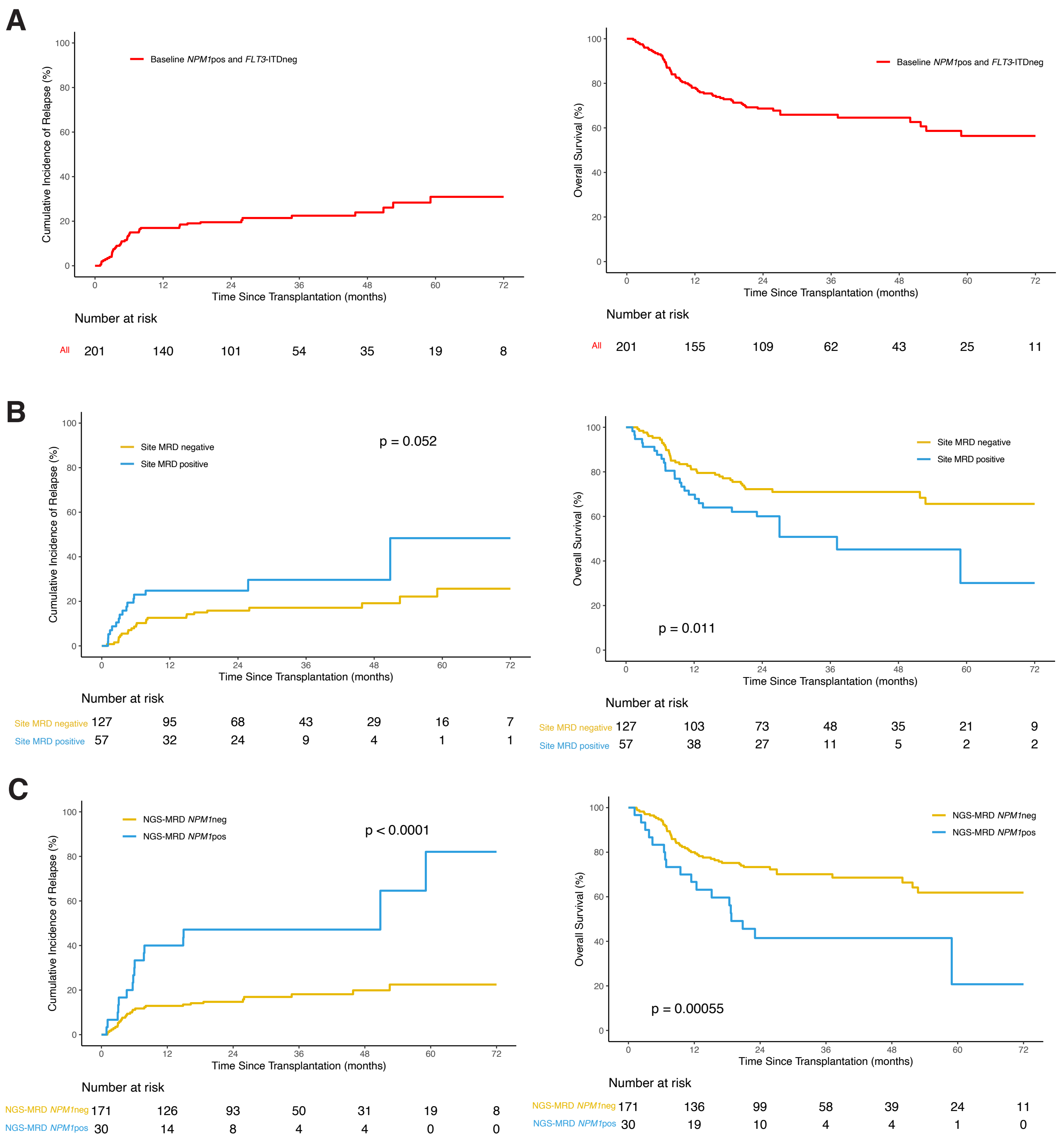

**Figure S8. Association of age and NGS-MRD status for *NPM1* and/or *FLT3*-ITD on clinical outcomes.** Rates of non-relapse mortality (NRM, top left), relapse (top right), relapse-free survival (RFS, bottom left), and overall survival (OS, bottom right) censored at 72 months are shown by pre-transplant NGS-MRD status for *NPM1* and/or *FLT3*-ITD based on baseline genetics for patients aged (A) <60 years and (B) ≥60 years. Point estimates at different time points are shown in the table (far right). Overall P values: Gray's test for NRM and relapse; log-rank test for RFS and OS. P values for pointwise estimations at different time points: z-test. Confidence interval, CI; Probability, prob; Month, mo; Year, yr; positive, pos; negative, neg.

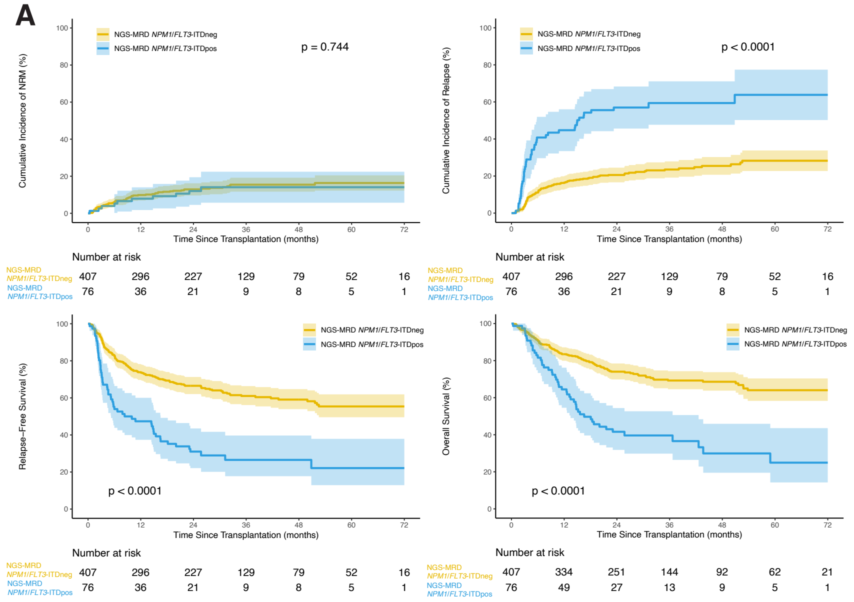


**Patients < 60yrs**

| Outcomes | Prob (95% CI) | Prob (95% CI) | p-value |
|---|---|---|---|
| NRM | *NPM1*/*FLT3*-ITDneg=407 | *NPM1*/*FLT3*-ITDpos=76 | 0.744 |
| 3 mo | 4(2-6)% | 3(0-6)% | 0.611 |
| 6 mo | 6(4-8)% | 5(0-10)% | 0.755 |
| 1 yr | 10(7-13)% | 8(2-14)% | 0.568 |
| 2 yr | 13(10-16)% | 12(5-19)% | 0.828 |
| 3 yr | 15(12-19)% | 14(6-22)% | 0.77 |
| Relapse | *NPM1*/*FLT3*-ITDneg=407 | *NPM1*/*FLT3*-ITDpos=76 | <0.001 |
| 3 mo | 4(2-6)% | 24(14-33)% | <0.001 |
| 6 mo | 11(8-14)% | 41(30-52)% | <0.001 |
| 1 yr | 16(13-20)% | 45(33-56)% | <0.001 |
| 2 yr | 21(17-25)% | 57(46-68)% | <0.001 |
| 3 yr | 24(19-28)% | 59(48-71)% | <0.001 |
| RFS | *NPM1*/*FLT3*-ITDneg=407 | *NPM1*/*FLT3*-ITDpos=76 | <0.001 |
| 3 mo | 92(89-95)% | 74(62-82)% | <0.001 |
| 6 mo | 83(79-86)% | 54(42-64)% | <0.001 |
| 1 yr | 74(69-78)% | 47(36-58)% | <0.001 |
| 2 yr | 67(62-71)% | 31(21-42)% | <0.001 |
| 3 yr | 61(56-66)% | 27(17-38)% | <0.001 |
| OS | *NPM1*/*FLT3*-ITDneg=407 | *NPM1*/*FLT3*-ITDpos=76 | <0.001 |
| 3 mo | 96(94-98)% | 97(90-99)% | 0.609 |
| 6 mo | 91(88-93)% | 82(71-89)% | 0.046 |
| 1 yr | 83(79-87)% | 64(53-74)% | 0.001 |
| 2 yr | 74(69-78)% | 42(30-52)% | <0.001 |
| 3 yr | 69(64-74)% | 40(28-51)% | <0.001 |

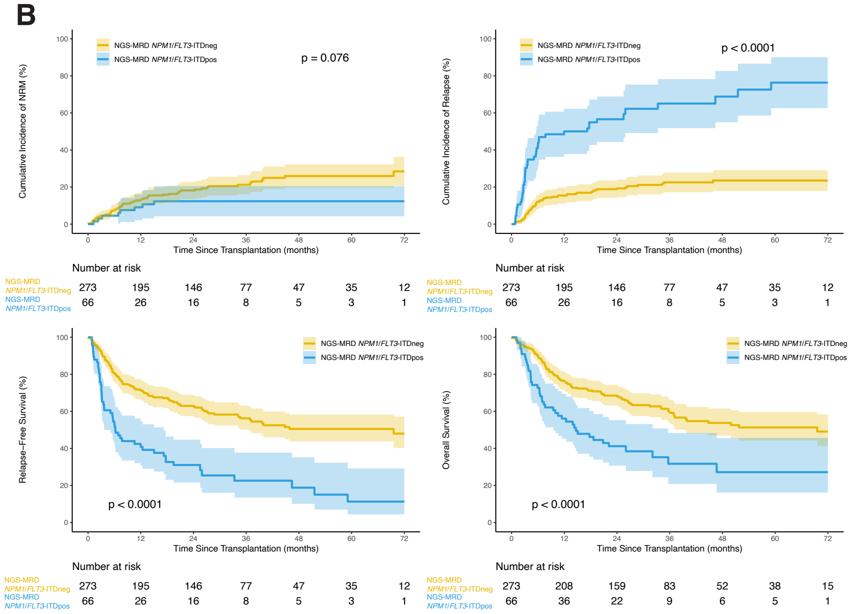


**Patients ≥ 60yrs**

| Outcomes | Prob (95% CI) | Prob (95% CI) | p-value |
|---|---|---|---|
| NRM | *NPM1*/*FLT3*-ITDneg=273 | *NPM1*/*FLT3*-ITDpos=66 | 0.076 |
| 3 mo | 4(2-7)% | 3(0-7)% | 0.58 |
| 6 mo | 7(4-10)% | 5(0-10)% | 0.358 |
| 1 yr | 13(9-17)% | 9(2-16)% | 0.361 |
| 2 yr | 18(14-23)% | 12(4-21)% | 0.223 |
| 3 yr | 21(16-26)% | 12(4-21)% | 0.071 |
| Relapse | *NPM1*/*FLT3*-ITDneg=273 | *NPM1*/*FLT3*-ITDpos=66 | <0.001 |
| 3 mo | 3(1-5)% | 23(13-33)% | <0.001 |
| 6 mo | 12(8-16)% | 42(30-54)% | <0.001 |
| 1 yr | 15(11-20)% | 50(38-62)% | <0.001 |
| 2 yr | 19(14-24)% | 57(44-69)% | <0.001 |
| 3 yr | 23(17-28)% | 65(52-78)% | <0.001 |
| RFS | *NPM1*/*FLT3*-ITDneg=273 | *NPM1*/*FLT3*-ITDpos=66 | <0.001 |
| 3 mo | 92(88-95)% | 74(62-83)% | 0.001 |
| 6 mo | 81(75-85)% | 53(40-64)% | <0.001 |
| 1 yr | 72(66-77)% | 41(29-52)% | <0.001 |
| 2 yr | 63(57-68)% | 31(20-42)% | <0.001 |
| 3 yr | 56(50-62)% | 23(12-35)% | <0.001 |
| OS | *NPM1*/*FLT3*-ITDneg=273 | *NPM1*/*FLT3*-ITDpos=66 | <0.001 |
| 3 mo | 95(91-97)% | 91(81-96)% | 0.295 |
| 6 mo | 90(85-93)% | 73(60-82)% | 0.003 |
| 1 yr | 76(71-81)% | 56(43-67)% | 0.002 |
| 2 yr | 68(63-74)% | 41(29-53)% | <0.001 |
| 3 yr | 60(53-66)% | 32(19-45)% | <0.001 |

**Figure S9. Association of persistent mutations by gene on clinical outcomes.** Rates of non-relapse mortality (NRM, top left), relapse (top right), relapse-free survival (RFS, bottom left), and overall survival (OS, bottom right) censored at 72 months are shown by gene based on baseline genetics and the detection of persistent mutations by NGS pre-transplant for (A) *FLT3*-ITD, (B) *NPM1*, (C) *NPM1* and/or *FLT3*-ITD, and (D) *NPM1* and *FLT3*-ITD. Point estimates at different time points are shown in the table (far right). Overall P values: Gray's test for NRM and relapse; log-rank test for DFS and OS. P values for pointwise estimations at different time points: z-test. Confidence interval, CI; Probability, prob; Month, mo; Year, yr; positive, pos; negative, neg.

A

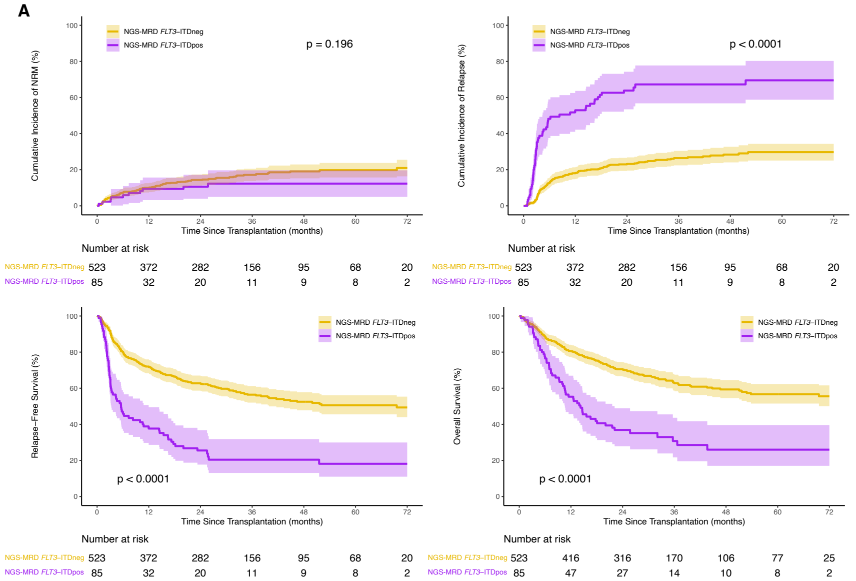


##### *FLT3*-ITD

| Outcomes | Prob (95% CI) | Prob (95% CI) | p-value |
|---|---|---|---|
| NRM | *FLT3*-ITDneg=523 | *FLT3*-ITDpos=85 | 0.196 |
| 3 mo | 5(3-6)% | 2(0-6)% | 0.237 |
| 6 mo | 7(5-10)% | 6(1-11)% | 0.621 |
| 1 yr | 10(7-13)% | 9(3-16)% | 0.872 |
| 2 yr | 14(11-17)% | 11(4-17)% | 0.32 |
| 3 yr | 17(14-21)% | 12(5-20)% | 0.244 |
| Relapse | *FLT3*-ITDneg=523 | *FLT3*-ITDpos=85 | <0.001 |
| 3 mo | 3(2-5)% | 32(22-42)% | <0.001 |
| 6 mo | 13(10-15)% | 48(38-59)% | <0.001 |
| 1 yr | 18(15-21)% | 53(42-64)% | <0.001 |
| 2 yr | 23(19-27)% | 64(53-74)% | <0.001 |
| 3 yr | 26(22-30)% | 67(57-78)% | <0.001 |
| DFS | *FLT3*-ITDneg=523 | *FLT3*-ITDpos=85 | <0.001 |
| 3 mo | 92(89-94)% | 66(55-75)% | <0.001 |
| 6 mo | 80(76-83)% | 46(35-56)% | <0.001 |
| 1 yr | 72(68-76)% | 38(27-48)% | <0.001 |
| 2 yr | 63(58-67)% | 26(17-35)% | <0.001 |
| 3 yr | 56(52-61)% | 20(12-30)% | <0.001 |
| OS | *FLT3*-ITDneg=523 | *FLT3*-ITDpos=85 | <0.001 |
| 3 mo | 95(93-97)% | 94(86-98)% | 0.633 |
| 6 mo | 89(85-91)% | 76(66-84)% | 0.012 |
| 1 yr | 81(77-84)% | 55(44-65)% | <0.001 |
| 2 yr | 70(66-74)% | 37(27-47)% | <0.001 |
| 3 yr | 63(58-67)% | 31(20-42)% | <0.001 |

B

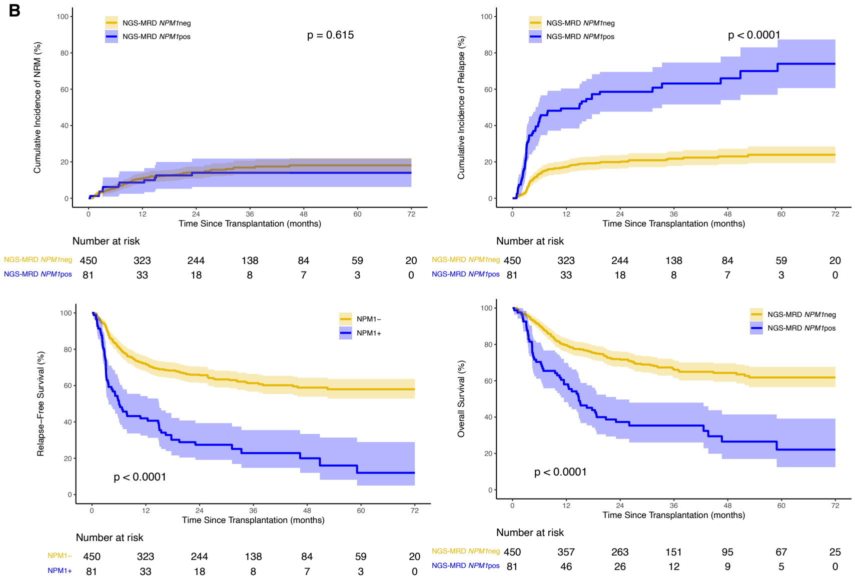


##### *NPM1*

| Outcomes | Prob (95% CI) | Prob (95% CI) | p-value |
|---|---|---|---|
| NRM | *NPM1*neg=450 | *NPM1*pos=81 | 0.615 |
| 3 mo | 3(2-5)% | 4(0-8)% | 0.871 |
| 6 mo | 6(4-8)% | 6(1-11)% | 0.894 |
| 1 yr | 11(8-14)% | 9(2-15)% | 0.554 |
| 2 yr | 14(11-17)% | 14(6-22)% | 0.979 |
| 3 yr | 17(13-21)% | 14(6-22)% | 0.515 |
| Relapse | *NPM1*neg=450 | *NPM1*pos=81 | <0.001 |
| 3 mo | 4(2-6)% | 22(13-31)% | <0.001 |
| 6 mo | 13(10-17)% | 43(32-54)% | <0.001 |
| 1 yr | 17(14-21)% | 49(38-60)% | <0.001 |
| 2 yr | 20(16-24)% | 59(48-70)% | <0.001 |
| 3 yr | 22(18-26)% | 63(52-75)% | <0.001 |
| DFS | *NPM1*neg=450 | *NPM1*pos=81 | <0.001 |
| 3 mo | 93(90-95)% | 74(63-82)% | <0.001 |
| 6 mo | 81(77-84)% | 51(39-61)% | <0.001 |
| 1 yr | 72(68-76)% | 42(31-52)% | <0.001 |
| 2 yr | 66(61-70)% | 27(18-38)% | <0.001 |
| 3 yr | 61(56-66)% | 23(14-33)% | <0.001 |
| OS | *NPM1*neg=450 | *NPM1*pos=81 | <0.001 |
| 3 mo | 96(94-98)% | 93(84-97)% | 0.233 |
| 6 mo | 90(87-93)% | 70(59-79)% | <0.001 |
| 1 yr | 80(76-83)% | 58(47-68)% | <0.001 |
| 2 yr | 72(67-76)% | 37(27-48)% | <0.001 |
| 3 yr | 66(61-71)% | 35(25-46)% | <0.001 |

**C**

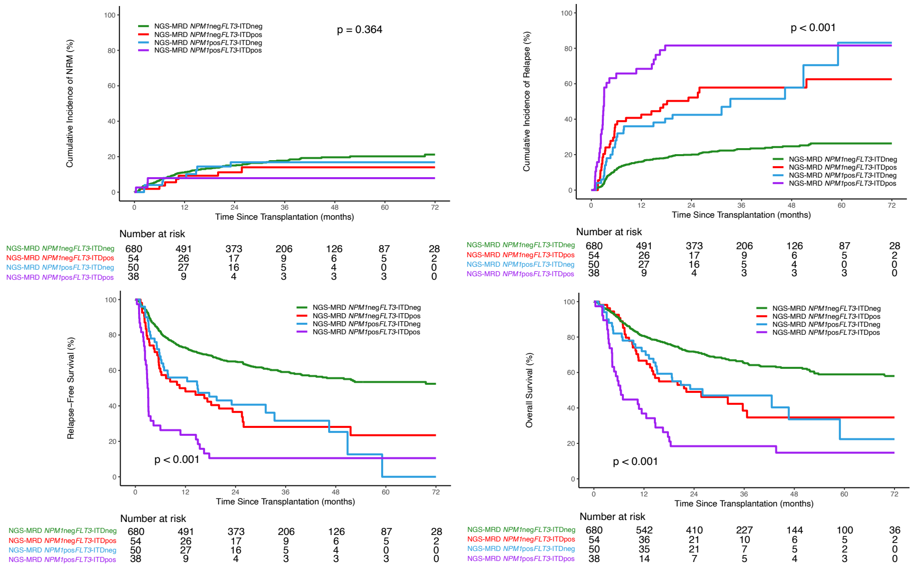


##### *NPM1* and/or *FLT3*-ITD

| Outcomes | Prob (95% CI) | Prob (95% CI) | Prob (95% CI) | Prob (95% CI) | p-value |
|---|---|---|---|---|---|
| NRM | *FLT3*-ITD*neg* *NPM1neg* *n*=680 | *FLT3*-ITDpos NPM1neg n=54 | *FLT3*-ITDneg *NPM1*pos n=50 | *FLT3*-ITD*pos* *NPM1pos* *n*=38 | 0.364 |
| 3 mo | 4(3-5)% | 2(0-5)% | 4(0-9)% | 3(0-8)% | 0.732 |
| 6 mo | 7(5-8)% | 4(0-9)% | 4(0-9)% | 8(0-17)% | 0.599 |
| 1 yr | 11(9-13)% | 9(1-17)% | 8(0-16)% | 8(0-17)% | 0.785 |
| 2 yr | 15(12-18)% | 11(3-20)% | 17(6-28)% | 8(0-17)% | 0.389 |
| 3 yr | 18(15-21)% | 14(4-24)% | 17(6-28)% | 8(0-17)% | 0.208 |
| Relapse | *FLT3*-ITD*neg* *NPM1neg* *n*=680 | *FLT3*-ITDpos NPM1neg n=54 | *FLT3*-ITDneg *NPM1*pos n=50 | *FLT3*-ITD*pos* *NPM1pos* *n*=38 | <0.001 |
| 3 mo | 4(2-5)% | 20(10-31)% | 8(0-16)% | 47(31-64)% | <0.001 |
| 6 mo | 11(9-14)% | 37(24-50)% | 28(15-41)% | 66(50-81)% | <0.001 |
| 1 yr | 16(13-19)% | 43(29-56)% | 36(23-49)% | 68(53-84)% | <0.001 |
| 2 yr | 20(17-23)% | 52(39-66)% | 42(28-57)% | 82(69-95)% | <0.001 |
| 3 yr | 23(20-26)% | 58(44-72)% | 52(35-68)% | 82(69-95)% | <0.001 |
| RFS | *FLT3*-ITD*neg* *NPM1neg* *n*=680 | *FLT3*-ITDpos NPM1neg n=54 | *FLT3*-ITDneg *NPM1*pos n=50 | *FLT3*-ITD*pos* *NPM1pos* *n*=38 | <0.001 |
| 3 mo | 92(90-94)% | 78(64-87)% | 88(75-94)% | 50(33-65)% | <0.001 |
| 6 mo | 82(79-85)% | 59(45-71)% | 68(53-79)% | 26(14-41)% | <0.001 |
| 1 yr | 73(69-76)% | 48(34-61)% | 56(41-68)% | 24(12-38)% | <0.001 |
| 2 yr | 65(61-69)% | 37(24-49)% | 41(27-54)% | 11(3-23)% | <0.001 |
| 3 yr | 59(55-63)% | 28(16-41)% | 32(17-47)% | 11(3-23)% | <0.001 |
| OS | *FLT3*-ITD*neg* *NPM1neg* *n*=680 | *FLT3*-ITDpos NPM1neg n=54 | *FLT3*-ITDneg *NPM1*pos n=50 | *FLT3*-ITD*pos* *NPM1pos* *n*=38 | <0.001 |
| 3 mo | 96(94-97)% | 98(88-100)% | 94(83-98)% | 89(74-96)% | 0.317 |
| 6 mo | 90(88-92)% | 91(79-96)% | 82(68-90)% | 53(36-67)% | <0.001 |
| 1 yr | 81(77-83)% | 67(52-78)% | 72(57-82)% | 37(22-52)% | <0.001 |
| 2 yr | 72(68-75)% | 49(35-62)% | 51(36-64)% | 18(8-32)% | <0.001 |
| 3 yr | 66(61-69)% | 38(24-53)% | 47(32-61)% | 18(8-32)% | <0.001 |

**D**

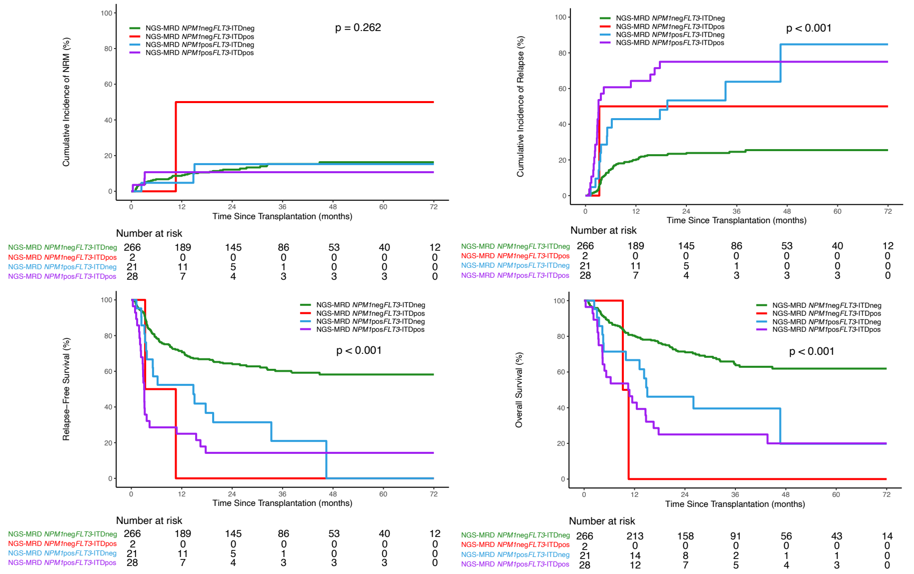


##### *NPM1* and *FLT3*-ITD

| Outcomes | Prob (95% CI) | Prob (95% CI) | Prob (95% CI) | Prob (95% CI) | p-value |
|---|---|---|---|---|---|
| NRM | *FLT3*-ITDneg *NPM1* neg n=266 | *FLT3*-ITDpos *NPM1* neg n=2 | *FLT3*-ITDneg *NPM1* pos n=21 | *FLT3*-ITDpos *NPM1* pos n=28 | 0.262 |
| 3 mo | 4(2-7)% | 0(0-0)% | 5(0-14)% | 4(0-11)% | 0.004 |
| 6 mo | 7(4-10)% | 0(0-0)% | 5(0-14)% | 11(0-23)% | <0.001 |
| 1 yr | 9(5-12)% | 50(0-100)% | 5(0-14)% | 11(0-23)% | 0.778 |
| 2 yr | 12(8-16)% | 50(0-100)% | 15(0-32)% | 11(0-23)% | 0.924 |
| 3 yr | 15(11-20)% | 50(0-100)% | 15(0-32)% | 11(0-23)% | 0.861 |
| Relapse | *FLT3*-ITDneg *NPM1* neg n=266 | *FLT3*-ITDpos *NPM1* neg n=2 | *FLT3*-ITDneg *NPM1* pos n=21 | *FLT3*-ITDpos *NPM1* pos n=28 | <0.001 |
| 3 mo | 3(1-5)% | 0(0-0)% | 10(0-22)% | 43(24-62)% | <0.001 |
| 6 mo | 15(10-19)% | 50(0-100)% | 38(17-60)% | 61(42-79)% | <0.001 |
| 1 yr | 20(15-25)% | 50(0-100)% | 43(21-65)% | 64(46-83)% | <0.001 |
| 2 yr | 23(18-29)% | 50(0-100)% | 53(31-76)% | 75(58-92)% | <0.001 |
| 3 yr | 25(19-30)% | 50(0-100)% | 64(36-91)% | 75(58-92)% | <0.001 |
| RFS | *FLT3*-ITDneg *NPM1* neg n=266 | *FLT3*-ITDpos *NPM1* neg n=2 | *FLT3*-ITDneg *NPM1* pos n=21 | *FLT3*-ITDpos *NPM1* pos n=28 | <0.001 |
| 3 mo | 93(89-95)% | 1 | 86(62-95)% | 54(34-70)% | <0.001 |
| 6 mo | 79(73-83)% | 50(1-91)% | 57(34-75)% | 29(14-46)% | <0.001 |
| 1 yr | 71(66-76)% | 0(NA-NA)% | 52(30-71)% | 25(11-42)% | <0.001 |
| 2 yr | 64(58-70)% | 0(NA-NA)% | 31(13-52)% | 14(4-30)% | <0.001 |
| 3 yr | 60(54-66)% | 0(NA-NA)% | 21(5-45)% | 14(4-30)% | <0.001 |
| OS | *FLT3*-ITDneg *NPM1* neg n=266 | *FLT3*-ITDpos *NPM1* neg n=2 | *FLT3*-ITDneg *NPM1* pos n=21 | *FLT3*-ITDpos *NPM1* pos n=28 | <0.001 |
| 3 mo | 96(93-98)% | 1 | 95(71-99)% | 89(70-96)% | 0.748 |
| 6 mo | 88(83-91)% | 1 | 71(47-86)% | 57(37-73)% | 0.006 |
| 1 yr | 80(75-85)% | 0(NA-NA)% | 67(43-83)% | 43(25-60)% | 0.001 |
| 2 yr | 71(66-77)% | 0(NA-NA)% | 46(24-66)% | 25(11-42)% | <0.001 |
| 3 yr | 64(58-70)% | 0(NA-NA)% | 40(18-60)% | 25(11-42)% | <0.001 |

**Figure S10. Association of conditioning intensity, age, and NGS-MRD status for *NPM1* and/or *FLT3*-ITD on clinical outcomes.** Rates of non-relapse mortality (NRM), relapse, relapse-free survival (RFS), and overall survival (OS) censored at 72 months are shown by conditioning intensity and pre-transplant NGS-MRD status for *NPM1* and/or *FLT3*-ITD based on baseline genetics for (A) All patients, (B) Patients aged <60 years, and (C) Patients aged ≥60 years. Point estimates at different time points are shown in the table (bottom). Overall P values: Gray's test for NRM and relapse; log-rank test for RFS and OS. P values for pointwise estimations at different time points: z-test. Confidence interval, CI; Probability, prob; Month, mo; Year, yr; positive, pos; negative, neg; myeloablative conditioning, MAC; reduced intensity conditioning, RIC; nonmyeloablative conditioning, NMA.

**A**

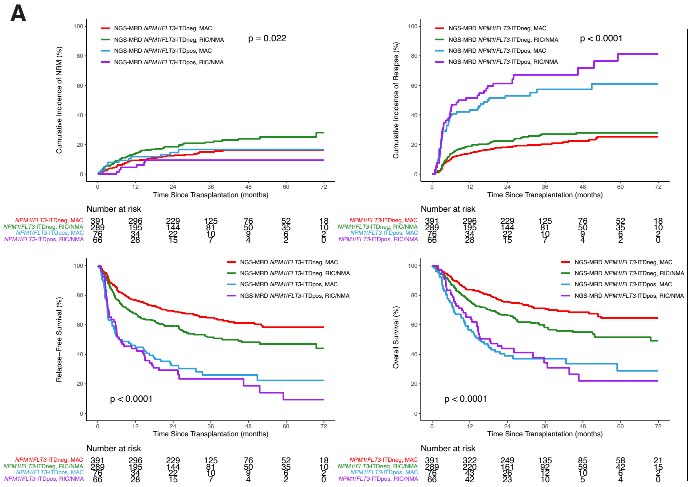


**All Patients**

| Outcomes | Prob (95% CI) | Prob (95% CI) | Prob (95% CI) | Prob (95% CI) | p-value |
|---|---|---|---|---|---|
| NRM | *NPM1*/*FLT3*-ITDneg, MAC=391 | *NPM1*/*FLT3*-ITDneg, RIC/NMA=289 | *NPM1*/*FLT3*-ITDpos, MAC=76 | *NPM1*/*FLT3*-ITDpos, RIC/NMA=66 | 0.022 |
| 3 mo | 3(1-4)% | 6(3-8)% | 5(0-10)% | 0(0-0)% | <0.001 |
| 6 mo | 5(3-7)% | 9(6-12)% | 8(2-14)% | 2(0-5)% | 0.008 |
| 1 yr | 9(6-12)% | 14(10-17)% | 12(5-19)% | 5(0-10)% | 0.048 |
| 2 yr | 12(9-16)% | 19(14-23)% | 15(7-23)% | 9(2-17)% | 0.099 |
| 3 yr | 15(11-19)% | 21(16-26)% | 17(8-25)% | 9(2-17)% | 0.049 |
| Relapse | *NPM1*/*FLT3*-ITDneg, MAC=391 | *NPM1*/*FLT3*-ITDneg, RIC/NMA=289 | *NPM1*/*FLT3*-ITDpos, MAC=76 | *NPM1*/*FLT3*-ITDpos, RIC/NMA=66 | <0.001 |
| 3 mo | 3(2-5)% | 4(2-6)% | 22(13-32)% | 24(14-35)% | <0.001 |
| 6 mo | 10(7-13)% | 14(10-18)% | 39(28-51)% | 44(32-56)% | <0.001 |
| 1 yr | 14(10-17)% | 19(14-23)% | 43(32-55)% | 52(39-64)% | <0.001 |
| 2 yr | 18(14-22)% | 22(17-27)% | 53(42-64)% | 61(49-73)% | <0.001 |
| 3 yr | 20(16-24)% | 27(21-33)% | 57(45-69)% | 67(54-80)% | <0.001 |
| RFS | *NPM1*/*FLT3*-ITDneg, MAC=391 | *NPM1*/*FLT3*-ITDneg, RIC/NMA=289 | *NPM1*/*FLT3*-ITDpos, MAC=76 | *NPM1*/*FLT3*-ITDpos, RIC/NMA=66 | <0.001 |
| 3 mo | 94(91-96)% | 90(86-93)% | 72(61-81)% | 76(64-84)% | <0.001 |
| 6 mo | 85(81-89)% | 77(72-82)% | 53(41-63)% | 55(42-66)% | <0.001 |
| 1 yr | 77(72-81)% | 68(62-73)% | 45(33-55)% | 44(32-55)% | <0.001 |
| 2 yr | 69(65-74)% | 59(53-65)% | 32(22-43)% | 29(19-41)% | <0.001 |
| 3 yr | 65(59-70)% | 52(45-58)% | 26(16-37)% | 23(13-36)% | <0.001 |
| OS | *NPM1*/*FLT3*-ITDneg, MAC=391 | *NPM1*/*FLT3*-ITDneg, RIC/NMA=289 | *NPM1*/*FLT3*-ITDpos, MAC=76 | *NPM1*/*FLT3*-ITDpos, RIC/NMA=66 | <0.001 |
| 3 mo | 97(95-98)% | 94(91-96)% | 92(83-96)% | 97(88-99)% | 0.188 |
| 6 mo | 93(89-95)% | 88(83-91)% | 75(64-83)% | 80(69-88)% | <0.001 |
| 1 yr | 84(80-87)% | 76(71-81)% | 57(45-67)% | 65(52-75)% | <0.001 |
| 2 yr | 76(71-80)% | 67(61-72)% | 39(28-50)% | 44(32-56)% | <0.001 |
| 3 yr | 71(66-76)% | 58(52-64)% | 37(26-48)% | 34(21-48)% | <0.001 |

**B**

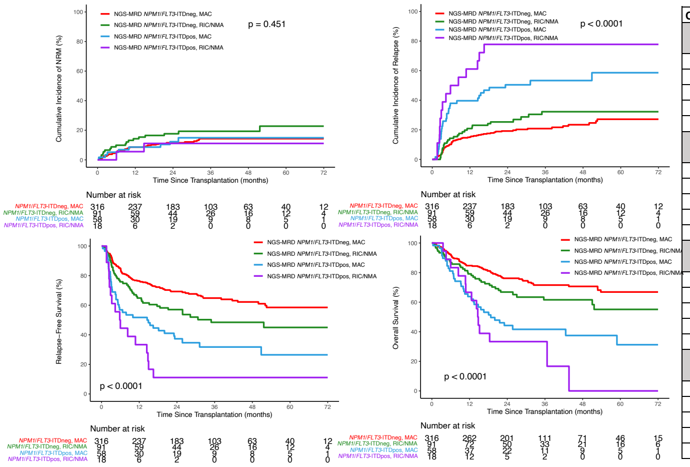


**Patients < 60yrs**

| Outcomes | Prob (95% CI) | Prob (95% CI) | Prob (95% CI) | Prob (95% CI) | p-value |
|---|---|---|---|---|---|
| NRM | *NPM1*/*FLT3*-ITDneg, MAC=316 | *NPM1*/*FLT3*-ITDneg, RIC/NMA=91 | *NPM1*/*FLT3*-ITDpos, MAC=58 | *NPM1*/*FLT3*-ITDpos, RIC/NMA=18 | 0.451 |
| 3 mo | 3(1-5)% | 7(1-12)% | 3(0-8)% | 0(0-0)% | <0.001 |
| 6 mo | 5(3-7)% | 10(4-16)% | 5(0-11)% | 6(0-17)% | 0.561 |
| 1 yr | 9(5-12)% | 14(7-22)% | 9(1-16)% | 6(0-17)% | 0.48 |
| 2 yr | 12(8-15)% | 18(10-26)% | 12(4-21)% | 11(0-27)% | 0.586 |
| 3 yr | 14(10-19)% | 19(11-28)% | 15(5-25)% | 11(0-27)% | 0.72 |
| Relapse | *NPM1*/*FLT3*-ITDneg, MAC=316 | *NPM1*/*FLT3*-ITDneg, RIC/NMA=91 | *NPM1*/*FLT3*-ITDpos, MAC=58 | *NPM1*/*FLT3*-ITDpos, RIC/NMA=18 | <0.001 |
| 3 mo | 4(2-6)% | 4(0-9)% | 21(10-31)% | 33(11-56)% | 0.001 |
| 6 mo | 10(7-13)% | 14(7-22)% | 38(25-51)% | 50(26-74)% | <0.001 |
| 1 yr | 15(11-19)% | 21(12-29)% | 40(27-52)% | 61(37-85)% | <0.001 |
| 2 yr | 19(15-24)% | 25(16-34)% | 50(37-64)% | 78(56-99)% | <0.001 |
| 3 yr | 21(16-26)% | 32(22-43)% | 53(40-67)% | 78(56-99)% | <0.001 |
| RFS | *NPM1*/*FLT3*-ITDneg, MAC=316 | *NPM1*/*FLT3*-ITDneg, RIC/NMA=91 | *NPM1*/*FLT3*-ITDpos, MAC=58 | *NPM1*/*FLT3*-ITDpos, RIC/NMA=18 | <0.001 |
| 3 mo | 93(90-96)% | 89(81-94)% | 76(63-85)% | 67(40-83)% | 0.002 |
| 6 mo | 85(80-88)% | 76(66-83)% | 57(43-68)% | 44(22-65)% | <0.001 |
| 1 yr | 76(71-81)% | 65(54-74)% | 52(38-64)% | 33(14-55)% | <0.001 |
| 2 yr | 69(64-74)% | 57(46-66)% | 37(25-50)% | 11(2-30)% | <0.001 |
| 3 yr | 65(59-70)% | 48(37-59)% | 32(20-45)% | 11(2-30)% | <0.001 |
| OS | *NPM1*/*FLT3*-ITDneg, MAC=316 | *NPM1*/*FLT3*-ITDneg, RIC/NMA=91 | *NPM1*/*FLT3*-ITDpos, MAC=58 | *NPM1*/*FLT3*-ITDpos, RIC/NMA=18 | <0.001 |
| 3 mo | 97(95-99)% | 93(86-97)% | 97(87-99)% | 1 | 0.606 |
| 6 mo | 92(88-95)% | 87(78-92)% | 81(68-89)% | 83(57-94)% | 0.103 |
| 1 yr | 85(80-88)% | 79(69-86)% | 64(50-75)% | 67(40-83)% | 0.006 |
| 2 yr | 76(71-81)% | 67(56-76)% | 44(31-57)% | 33(14-55)% | <0.001 |
| 3 yr | 72(66-77)% | 62(50-71)% | 42(28-54)% | 33(14-55)% | <0.001 |

**C**

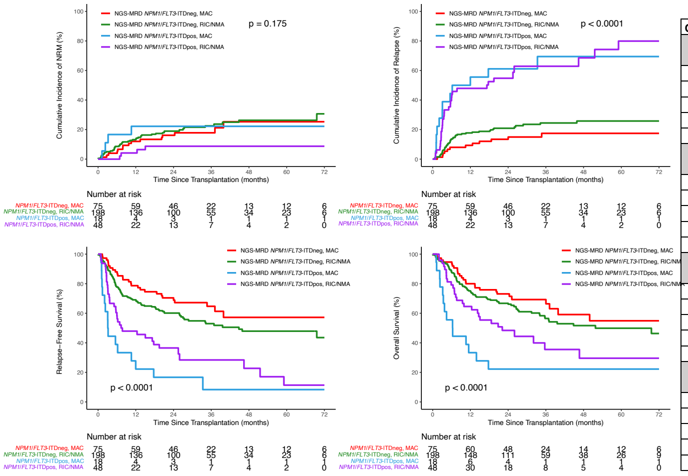


**Patients ≥ 60yrs**

| Outcomes | Prob (95% CI) | Prob (95% CI) | Prob (95% CI) | Prob (95% CI) | p-value |
|---|---|---|---|---|---|
| NRM | *NPM1*/*FLT3*-ITDneg, MAC=75 | *NPM1*/*FLT3*-ITDneg, RIC/NMA=198 | *NPM1*/*FLT3*-ITDpos, MAC=18 | *NPM1*/*FLT3*-ITDpos, RIC/NMA=48 | 0.175 |
| 3 mo | 3(0-6)% | 5(2-8)% | 11(0-26)% | 0(0-0)% | 0.002 |
| 6 mo | 4(0-8)% | 9(5-13)% | 17(0-35)% | 0(0-0)% | <0.001 |
| 1 yr | 12(5-19)% | 13(8-18)% | 22(2-43)% | 4(0-10)% | 0.063 |
| 2 yr | 16(8-25)% | 19(13-24)% | 22(2-43)% | 9(0-17)% | 0.225 |
| 3 yr | 18(9-27)% | 23(16-29)% | 22(2-43)% | 9(0-17)% | 0.076 |
| Relapse | *NPM1*/*FLT3*-ITDneg, MAC=75 | *NPM1*/*FLT3*-ITDneg, RIC/NMA=198 | *NPM1*/*FLT3*-ITDpos, MAC=18 | *NPM1*/*FLT3*-ITDpos, RIC/NMA=48 | <0.001 |
| 3 mo | 1(0-4)% | 4(1-7)% | 28(6-49)% | 21(9-32)% | 0.001 |
| 6 mo | 8(2-14)% | 14(9-18)% | 44(20-68)% | 42(28-56)% | <0.001 |
| 1 yr | 9(3-16)% | 18(12-23)% | 56(31-80)% | 48(34-62)% | <0.001 |
| 2 yr | 13(6-21)% | 21(15-27)% | 61(37-85)% | 55(40-69)% | <0.001 |
| 3 yr | 17(8-27)% | 24(18-31)% | 69(43-95)% | 63(47-79)% | <0.001 |
| RFS | *NPM1*/*FLT3*-ITDneg, MAC=75 | *NPM1*/*FLT3*-ITDneg, RIC/NMA=198 | *NPM1*/*FLT3*-ITDpos, MAC=18 | *NPM1*/*FLT3*-ITDpos, RIC/NMA=48 | <0.001 |
| 3 mo | 96(88-99)% | 91(86-94)% | 61(35-79)% | 79(65-88)% | 0.001 |
| 6 mo | 88(78-94)% | 78(71-83)% | 39(17-60)% | 58(43-71)% | <0.001 |
| 1 yr | 79(68-86)% | 69(62-75)% | 22(7-43)% | 48(33-61)% | <0.001 |
| 2 yr | 70(59-79)% | 60(53-67)% | 17(4-37)% | 37(23-50)% | <0.001 |
| 3 yr | 65(52-75)% | 53(45-60)% | 8(1-29)% | 28(15-43)% | <0.001 |
| OS | *NPM1*/*FLT3*-ITDneg, MAC=75 | *NPM1*/*FLT3*-ITDneg, RIC/NMA=198 | *NPM1*/*FLT3*-ITDpos, MAC=18 | *NPM1*/*FLT3*-ITDpos, RIC/NMA=48 | <0.001 |
| 3 mo | 96(88-99)% | 94(90-97)% | 78(51-91)% | 96(84-99)% | 0.324 |
| 6 mo | 95(86-98)% | 88(82-92)% | 56(31-75)% | 79(65-88)% | 0.001 |
| 1 yr | 80(69-87)% | 75(68-81)% | 33(14-55)% | 65(49-76)% | <0.001 |
| 2 yr | 73(61-82)% | 67(60-73)% | 22(7-43)% | 48(33-62)% | <0.001 |
| 3 yr | 69(57-79)% | 56(48-64)% | 22(7-43)% | 35(20-52)% | <0.001 |

**Figure S11. Association of *NPM1* and/or *FLT3*-ITD mutational status and NGS-MRD VAF on clinical outcomes.**

**i. *NPM1* and/or *FLT3*-ITD mutational status.** Multivariate Cox regression analysis shown for relapse (left) and overall survival (right) for patients with baseline mutation profiles (A) *NPM1* positive and *FLT3*-ITD positive (n=317), (B) *NPM1* positive and *FLT3*-ITD negative (n=201), and (C) *NPM1* negative and *FLT3*-ITD positive (n=226). Positive, pos; negative, neg; reduced intensity conditioning, RIC; nonmyeloablative conditioning, NMA; Melphalan, Mel; Next-generation sequencing, NGS; measurable residual disease, MRD.

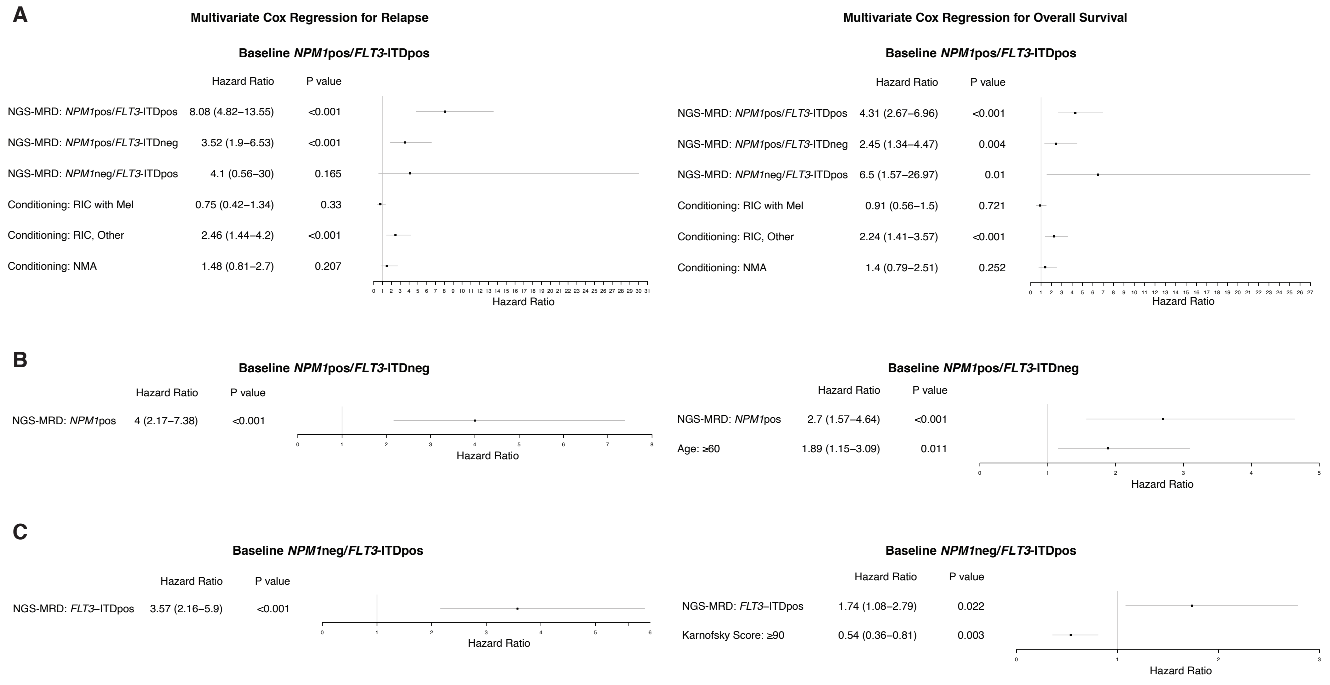

**ii. VAF cutoff, outcomes.** Rates of non-relapse mortality (NRM), relapse, relapse-free survival (RFS), and overall survival (OS) censored at 72 months are shown for patients with *NPM1* and/or *FLT3*-ITD baseline genetics (n=822) by NGS-MRD status pre-transplant using variant allele frequency (VAF) cutoffs of (A) ≥ 0.01% (B) ≥ 0.1%, and (C) ≥ 1%. Point estimates at different time points are shown in the table (bottom). Overall P values: Gray's test for NRM and relapse; log-rank test for RFS and OS. P values for pointwise estimations at different time points: z-test. Confidence interval, CI; Probability, prob; Month, mo; Year, yr; positive, pos; negative, neg.

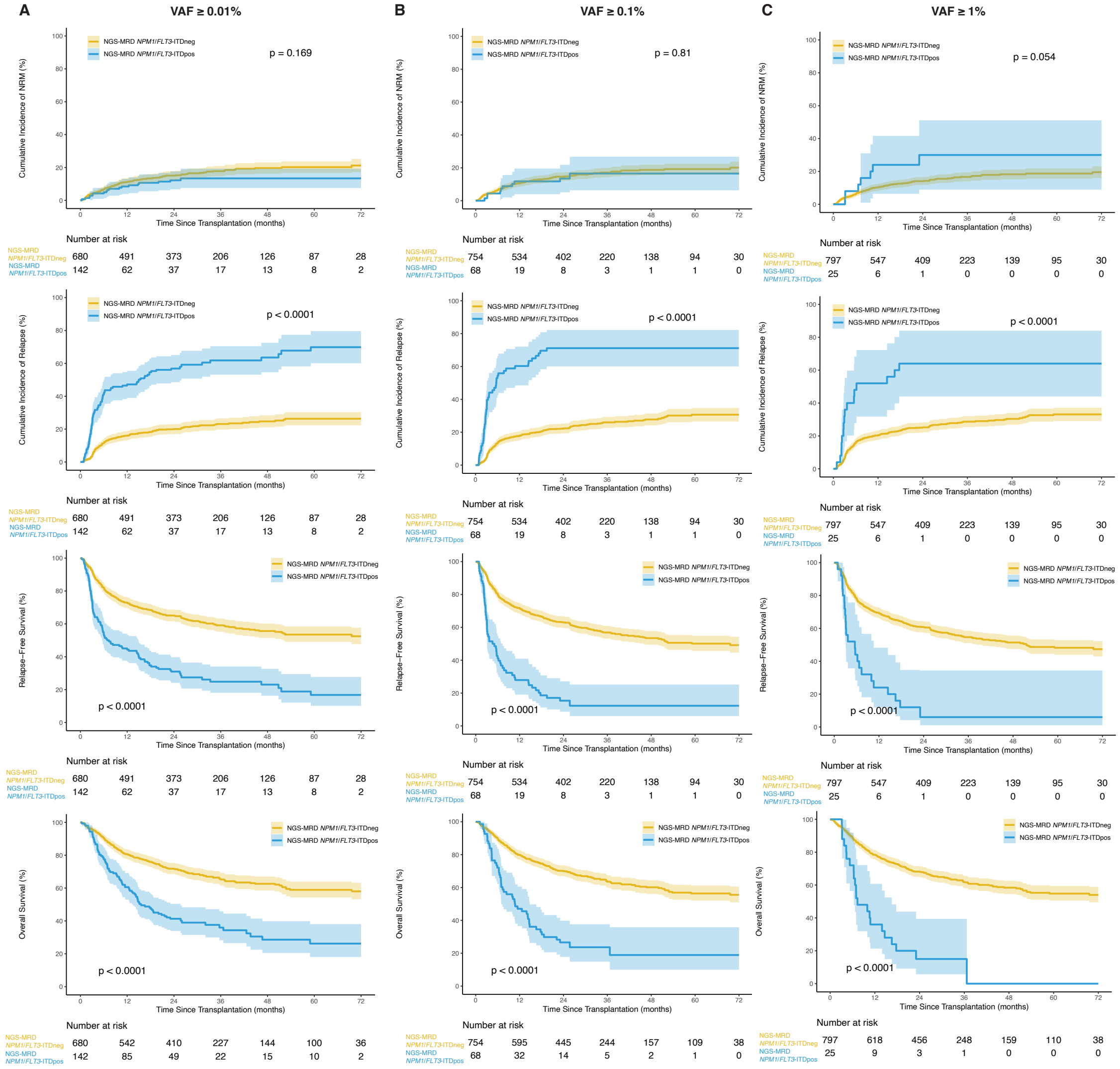


| Outcomes | Prob (95% CI) | Prob (95% CI) | p-value |
|---|---|---|---|
| NRM | *NPM1*/*FLT3*-ITDneg (VAF≤0.01%)=680 | *NPM1*/*FLT3*-ITDpos (VAF≤0.01%)=142 | 0.169 |
| 3 mo | 4(3-5)% | 3(0-6)% | 0.466 |
| 6 mo | 7(5-8)% | 5(1-9)% | 0.41 |
| 1 yr | 11(9-13)% | 8(4-13)% | 0.321 |
| 2 yr | 15(12-18)% | 12(7-18)% | 0.361 |
| 3 yr | 18(15-21)% | 13(7-19)% | 0.195 |
| Relapse | *NPM1*/*FLT3*-ITDneg (VAF≤0.01%)=680 | *NPM1*/*FLT3*-ITDpos (VAF≤0.01%)=142 | <0.001 |
| 3 mo | 4(2-5)% | 23(16-30)% | <0.001 |
| 6 mo | 11(9-14)% | 42(33-50)% | <0.001 |
| 1 yr | 16(13-19)% | 47(39-55)% | <0.001 |
| 2 yr | 20(17-23)% | 57(49-65)% | <0.001 |
| 3 yr | 23(20-26)% | 62(53-70)% | <0.001 |
| RFS | *NPM1*/*FLT3*-ITDneg (VAF≤0.01%)=680 | *NPM1*/*FLT3*-ITDpos (VAF≤0.01%)=142 | <0.001 |
| 3 mo | 92(90-94)% | 74(66-80)% | <0.001 |
| 6 mo | 82(79-85)% | 54(45-61)% | <0.001 |
| 1 yr | 73(69-76)% | 44(36-52)% | <0.001 |
| 2 yr | 65(61-69)% | 31(23-39)% | <0.001 |
| 3 yr | 59(55-63)% | 25(17-33)% | <0.001 |
| OS | *NPM1*/*FLT3*-ITDneg (VAF≤0.01%)=680 | *NPM1*/*FLT3*-ITDpos (VAF≤0.01%)=142 | <0.001 |
| 3 mo | 96(94-97)% | 94(89-97)% | 0.511 |
| 6 mo | 90(88-92)% | 77(70-83)% | <0.001 |
| 1 yr | 81(77-83)% | 61(52-68)% | <0.001 |
| 2 yr | 72(68-75)% | 41(33-49)% | <0.001 |
| 3 yr | 66(61-69)% | 36(27-45)% | <0.001 |

| Outcomes | Prob (95% CI) | Prob (95% CI) | p-value |
|---|---|---|---|
| NRM | *NPM1*/*FLT3*-ITDneg (VAF≤0.1%)=754 | *NPM1*/*FLT3*-ITDpos (VAF≤0.01%)=68 | 0.81 |
| 3 mo | 4(3-5)% | 1(0-4)% | 0.125 |
| 6 mo | 7(5-8)% | 4(0-9)% | 0.433 |
| 1 yr | 11(8-13)% | 12(4-20)% | 0.762 |
| 2 yr | 15(12-17)% | 13(5-22)% | 0.796 |
| 3 yr | 17(14-20)% | 17(6-27)% | 0.92 |
| Relapse | *NPM1*/*FLT3*-ITDneg (VAF≤0.1%)=754 | *NPM1*/*FLT3*-ITDpos (VAF≤0.01%)=68 | <0.001 |
| 3 mo | 5(3-6)% | 34(22-45)% | <0.001 |
| 6 mo | 13(11-16)% | 54(42-66)% | <0.001 |
| 1 yr | 18(15-21)% | 60(48-72)% | <0.001 |
| 2 yr | 22(19-25)% | 71(60-82)% | <0.001 |
| 3 yr | 26(23-29)% | 71(60-82)% | <0.001 |
| RFS | *NPM1*/*FLT3*-ITDneg (VAF≤0.1%)=754 | *NPM1*/*FLT3*-ITDpos (VAF≤0.01%)=68 | <0.001 |
| 3 mo | 91(89-93)% | 65(52-75)% | <0.001 |
| 6 mo | 80(77-83)% | 41(29-53)% | <0.001 |
| 1 yr | 72(68-75)% | 28(18-39)% | <0.001 |
| 2 yr | 63(60-67)% | 15(8-25)% | <0.001 |
| 3 yr | 57(53-61)% | 12(5-23)% | <0.001 |
| OS | *NPM1*/*FLT3*-ITDneg (VAF≤0.1%)=754 | *NPM1*/*FLT3*-ITDpos (VAF≤0.01%)=68 | <0.001 |
| 3 mo | 96(94-97)% | 93(83-97)% | 0.339 |
| 6 mo | 90(87-92)% | 72(60-81)% | 0.002 |
| 1 yr | 80(77-83)% | 47(35-58)% | <0.001 |
| 2 yr | 70(67-73)% | 27(17-38)% | <0.001 |
| 3 yr | 64(60-67)% | 24(14-35)% | <0.001 |

| Outcomes | Prob (95% CI) | Prob (95% CI) | p-value |
|---|---|---|---|
| NRM | *NPM1*/*FLT3*-ITDneg (VAF≤1%)=797 | *NPM1*/*FLT3*-ITDpos (VAF≤1%)=25 | 0.054 |
| 3 mo | 4(3-5)% | 0(0-0)% | <0.001 |
| 6 mo | 6(5-8)% | 8(0-19)% | 0.762 |
| 1 yr | 10(8-12)% | 24(6-42)% | 0.127 |
| 2 yr | 14(12-17)% | 30(9-51)% | 0.142 |
| 3 yr | 17(14-19)% | 30(9-51)% | 0.218 |
| Relapse | *NPM1*/*FLT3*-ITDneg (VAF≤1%)=797 | *NPM1*/*FLT3*-ITDpos (VAF≤1%)=25 | <0.001 |
| 3 mo | 6(5-8)% | 32(13-51)% | 0.007 |
| 6 mo | 16(13-18)% | 48(28-68)% | 0.002 |
| 1 yr | 20(18-23)% | 52(32-72)% | 0.002 |
| 2 yr | 25(22-28)% | 64(44-84)% | <0.001 |
| 3 yr | 29(25-32)% | 64(44-84)% | <0.001 |
| RFS | *NPM1*/*FLT3*-ITDneg (VAF≤1%)=797 | *NPM1*/*FLT3*-ITDpos (VAF≤1%)=25 | <0.001 |
| 3 mo | 90(88-92)% | 68(46-83)% | 0.02 |
| 6 mo | 78(75-81)% | 44(24-62)% | <0.001 |
| 1 yr | 69(66-73)% | 24(10-42)% | <0.001 |
| 2 yr | 61(57-64)% | 6(1-22)% | <0.001 |
| 3 yr | 55(51-58)% | 6(1-22)% | <0.001 |
| OS | *NPM1*/*FLT3*-ITDneg (VAF≤1%)=797 | *NPM1*/*FLT3*-ITDpos (VAF≤1%)=25 | <0.001 |
| 3 mo | 95(94-97)% | 1 | NA |
| 6 mo | 89(86-91)% | 72(50-86)% | 0.065 |
| 1 yr | 78(75-81)% | 36(18-54)% | <0.001 |
| 2 yr | 68(65-71)% | 15(4-32)% | <0.001 |
| 3 yr | 62(58-65)% | 15(4-32)% | <0.001 |

**iii. VAF cutoff, univariate analysis.** Univariate Cox regression analysis shown for (A) relapse and (B) overall survival for patients with baseline *NPM1* and/or *FLT3*-ITD mutations (n=822) where NGS-MRD status for *NPM1* and/or *FLT3*-ITD was binned by variant allele fraction (VAF) into 3 groups: 0.01%≤VAF<0.1%, 0.1%≤VAF<1%, and VAF≥1%. If a patient was positive for more than one mutation, the highest VAF was used. Baseline variable for all groups is NGS-MRD *NPM1*/*FLT3*-ITDneg. Positive, pos; negative, neg; Next-generation sequencing, NGS; measurable residual disease, MRD.

**A**

**Univariate Cox Regression for Relapse**

| | | |
|---|---|---|
| NGS-MRD *NPM1*/*FLT3*-ITDpos (0.01≤VAF<0.1%) | 2.87 (2.02–4.06) | <0.001 |
| NGS-MRD *NPM1*/*FLT3*-ITDpos (0.1≤VAF<1%) | 6.31 (4.3–9.27) | <0.001 |
| NGS-MRD *NPM1*/*FLT3*-ITDpos (VAF≥1%) | 5.66 (3.36–9.52) | <0.001 |

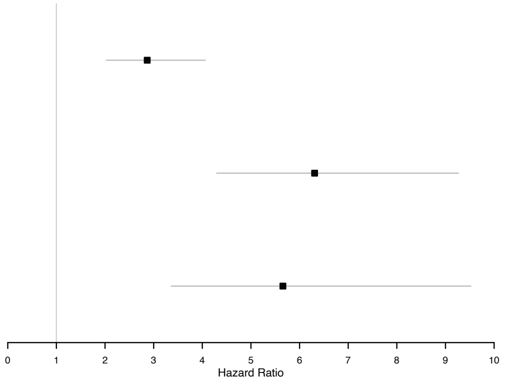


**Univariate Cox Regression for Overall Survival**

**B**

| | Hazard Ratio | P value |
|---|---|---|
| NGS-MRD *NPM1*/*FLT3*-ITDpos (0.01≤VAF<0.1%) | 1.8 (1.29–2.52) | <0.001 |
| NGS-MRD *NPM1*/*FLT3*-ITDpos (0.1≤VAF<1%) | 3.12 (2.11–4.6) | <0.001 |
| NGS-MRD *NPM1*/*FLT3*-ITDpos (VAF≥1%) | 4.97 (3.19–7.74) | <0.001 |

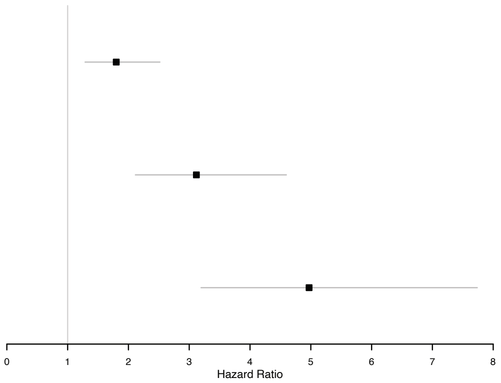

**iv. VAF cutoff, multivariate analysis.** Multivariate Cox regression analysis shown for (A) relapse and (B) overall survival for patients with baseline *NPM1* and/or *FLT3*-ITD mutations (n=822) where NGS-MRD status for *NPM1* and/or *FLT3*-ITD was binned by variant allele fraction (VAF) into 3 groups: 0.01%≤VAF<0.1%, 0.1%≤VAF<1%, and VAF≥1%. If a patient was positive for more than one mutation, the highest VAF was used. Positive, pos; negative, neg; reduced intensity conditioning, RIC; nonmyeloablative conditioning, NMA; Melphalan, Mel; Next-generation sequencing, NGS; measurable residual disease, MRD.

**A**

**Multivariate Cox Regression for Relapse**

| | Hazard Ratio | P value |
|---|---|---|
| NGS-MRD *NPM1*/*FLT3*-ITDpos (0.01≤VAF<0.1%) | 3.08 (2.17–4.37) | <0.001 |
| NGS-MRD *NPM1*/*FLT3*-ITDpos (0.1≤VAF<1%) | 6.19 (4.18–9.17) | <0.001 |
| NGS-MRD *NPM1*/*FLT3*-ITDpos (VAF≥1%) | 5.62 (3.33–9.49) | <0.001 |
| Conditioning: RIC with Mel | 0.9 (0.63–1.29) | 0.57 |
| Conditioning: RIC, Other | 1.8 (1.28–2.53) | <0.001 |
| Conditioning: NMA | 1.49 (1–2.23) | 0.052 |
| Baseline *FLT3*–ITD: Positive | 1.51 (1.1–2.08) | 0.011 |

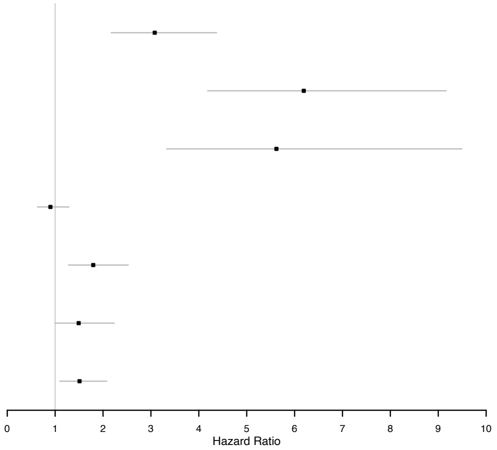


**B**

**Multivariate Cox Regression for Overall Survival**

| | Hazard Ratio | P value |
|---|---|---|
| NGS-MRD *NPM1*/*FLT3*-ITDpos (0.01≤VAF<0.1%) | 1.8 (1.29–2.52) | <0.001 |
| NGS-MRD *NPM1*/*FLT3*-ITDpos (0.1≤VAF<1%) | 2.94 (1.99–4.35) | <0.001 |
| NGS-MRD *NPM1*/*FLT3*-ITDpos (VAF≥1%) | 5.86 (3.74–9.2) | <0.001 |
| Conditioning: RIC/NMA | 1.14 (0.88–1.48) | 0.316 |
| Graft Type: Cord Blood | 1.67 (1.24–2.25) | <0.001 |
| Graft Type: Bone Marrow | 0.9 (0.64–1.28) | 0.567 |
| Karnofsky Score: ≥90 | 0.76 (0.61–0.95) | 0.016 |
| Baseline *IDH2*: Positive | 0.65 (0.43–0.99) | 0.044 |
| Age: ≥60 | 1.31 (1.02–1.7) | 0.038 |

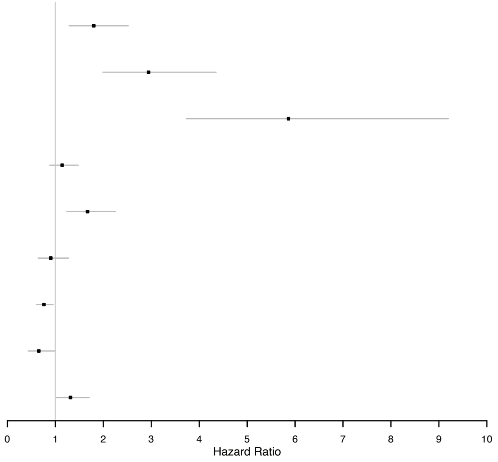

**Figure S12. Association of post-transplant maintenance therapy and NGS-MRD status on clinical outcomes.**

**i. FLT3 inhibitor.** Rates of non-relapse mortality (NRM), relapse, relapse-free survival (RFS), and overall survival (OS) censored at 72 months are shown for patients with *FLT3*-ITD positive baseline genetics based on (A) post-transplant FLT3 inhibitor (FLT3i) maintenance therapy usage alone and (B) in combination with pre-transplant NGS-MRD status. Point estimates at different time points are shown in the table (bottom). Overall P values: Gray's test for NRM and relapse; log-rank test for RFS and OS. P values for pointwise estimations at different time points: z-test. Confidence interval, CI; Probability, prob; Month, mo; Year, yr; positive, pos; negative, neg.

A

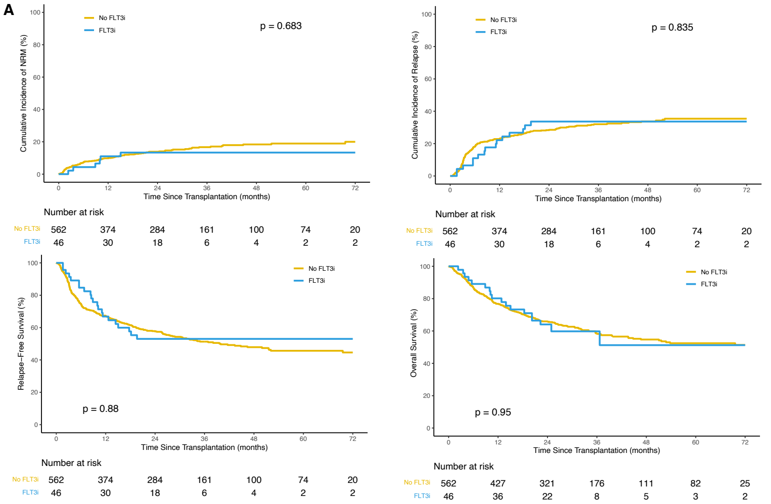


**Baseline *FLT3*-ITD, FLT3i**

| Outcomes | Prob (95% CI) | Prob (95% CI) | p-value |
|---|---|---|---|
| NRM | No FLT3i=562 | FLT3i=46 | 0.683 |
| 3 mo | 4(3-6)% | 2(0-6)% | 0.332 |
| 6 mo | 7(5-9)% | 4(0-10)% | 0.361 |
| 1 yr | 10(7-12)% | 11(2-20)% | 0.801 |
| 2 yr | 14(11-17)% | 13(3-23)% | 0.924 |
| 3 yr | 17(13-20)% | 13(3-23)% | 0.537 |
| Relapse | No FLT3i=562 | FLT3i=46 | 0.835 |
| 3 mo | 8(5-10)% | 4(0-10)% | 0.308 |
| 6 mo | 18(15-21)% | 11(2-20)% | 0.148 |
| 1 yr | 23(20-26)% | 22(10-34)% | 0.893 |
| 2 yr | 28(25-32)% | 34(20-48)% | 0.476 |
| 3 yr | 32(28-36)% | 34(20-48)% | 0.826 |
| RFS | No FLT3i=562 | FLT3i=46 | 0.88 |
| 3 mo | 88(85-90)% | 93(81-98)% | 0.152 |
| 6 mo | 75(71-78)% | 85(71-92)% | 0.072 |
| 1 yr | 67(63-71)% | 67(51-79)% | 0.961 |
| 2 yr | 58(54-62)% | 53(37-66)% | 0.537 |
| 3 yr | 51(47-56)% | 53(37-66)% | 0.828 |
| OS | No FLT3i=562 | FLT3i=46 | 0.95 |
| 3 mo | 95(93-97)% | 98(86-100)% | 0.23 |
| 6 mo | 87(84-89)% | 89(76-95)% | 0.614 |
| 1 yr | 77(73-80)% | 80(65-89)% | 0.587 |
| 2 yr | 66(62-70)% | 64(48-76)% | 0.802 |
| 3 yr | 59(54-63)% | 60(43-73)% | 0.88 |

B

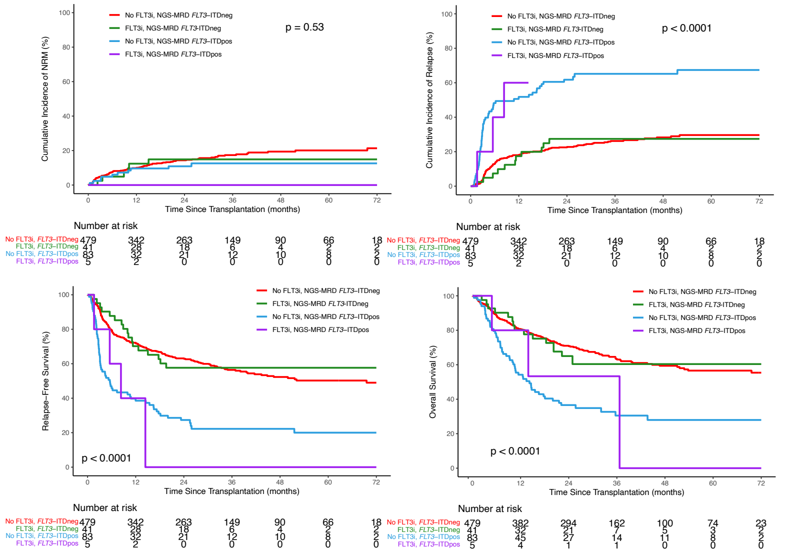


**Baseline *FLT3*-ITD, FLT3i & NGS-MRD**

| Outcomes | Prob (95% CI) | Prob (95% CI) | Prob (95% CI) | Prob (95% CI) | p-value |
|---|---|---|---|---|---|
| NRM | No FLT3i, *FLT3*-ITDneg=479 | FLT3i, *FLT3*-ITDneg=41 | No FLT3i, *FLT3*-ITDpos=83 | FLT3i, *FLT3*-ITDpos=5 | 0.53 |
| 3 mo | 5(3-7)% | 2(0-7)% | 2(0-6)% | 0(0-0)% | <0.001 |
| 6 mo | 8(5-10)% | 5(0-12)% | 6(1-11)% | 0(0-0)% | <0.001 |
| 1 yr | 10(7-13)% | 12(2-23)% | 10(3-16)% | 0(0-0)% | <0.001 |
| 2 yr | 14(11-18)% | 15(4-26)% | 11(4-18)% | 0(0-0)% | <0.001 |
| 3 yr | 17(14-21)% | 15(4-26)% | 13(5-20)% | 0(0-0)% | <0.001 |
| Relapse | No FLT3i, *FLT3*-ITDneg=479 | FLT3i, *FLT3*-ITDneg=41 | No FLT3i, *FLT3*-ITDpos=83 | FLT3i, *FLT3*-ITDpos=5 | <0.001 |
| 3 mo | 3(2-5)% | 2(0-7)% | 33(22-43)% | 20(0-59)% | <0.001 |
| 6 mo | 13(10-16)% | 7(0-16)% | 48(37-59)% | 40(0-89)% | <0.001 |
| 1 yr | 18(15-21)% | 17(6-29)% | 52(41-63)% | 60(9-100)% | <0.001 |
| 2 yr | 23(19-26)% | 27(13-41)% | 62(51-72)% | 100(22-100)% | <0.001 |
| 3 yr | 26(22-30)% | 27(13-41)% | 65(54-76)% | 100(22-100)% | <0.001 |
| RFS | No FLT3i, *FLT3*-ITDneg=479 | FLT3i, *FLT3*-ITDneg=41 | No FLT3i, *FLT3*-ITDpos=83 | FLT3i, *FLT3*-ITDpos=5 | <0.001 |
| 3 mo | 92(89-94)% | 95(82-99)% | 65(54-74)% | 80(20-97)% | NA |
| 6 mo | 80(76-83)% | 88(73-95)% | 46(35-56)% | 60(13-88)% | NA |
| 1 yr | 72(68-76)% | 70(53-82)% | 39(28-49)% | 40(5-75)% | NA |
| 2 yr | 63(59-67)% | 58(41-71)% | 27(18-37)% | 0(NA-NA)% | NA |
| 3 yr | 56(51-61)% | 58(41-71)% | 22(14-32)% | 0(NA-NA)% | NA |
| OS | No FLT3i, *FLT3*-ITDneg=479 | FLT3i, *FLT3*-ITDneg=41 | No FLT3i, *FLT3*-ITDpos=83 | FLT3i, *FLT3*-ITDpos=5 | <0.001 |
| 3 mo | 95(93-97)% | 98(84-100)% | 94(86-97)% | 1 | 0.767 |
| 6 mo | 89(85-91)% | 90(76-96)% | 76(65-84)% | 80(20-97)% | 0.069 |
| 1 yr | 81(77-84)% | 80(64-90)% | 54(43-64)% | 80(20-97)% | <0.001 |
| 2 yr | 71(67-75)% | 65(48-78)% | 37(26-47)% | 53(7-86)% | <0.001 |
| 3 yr | 63(58-68)% | 60(42-74)% | 31(20-41)% | 53(7-86)% | <0.001 |

**ii. FLT3 inhibitor and/or hypomethylating agents.** Rates of non-relapse mortality (NRM), relapse, relapse-free survival (RFS), and overall survival (OS) censored at 72 months are shown for patients with *NPM1* and/or *FLT3*-ITD positive baseline genetics based on (A) post-transplant maintenance therapy (including FLT3 inhibitors, or hypomethylating agents) usage alone and (B) in combination with pre-transplant *NPM1* and/or *FLT3*-ITD NGS-MRD status. Point estimates at different time points are shown in the table (bottom). Overall P values: Gray's test for NRM and relapse; log-rank test for RFS and OS. P values for pointwise estimations at different time points: z-test. Confidence interval, CI; Probability, prob; Month, mo; Year, yr; positive, pos; negative, neg

A

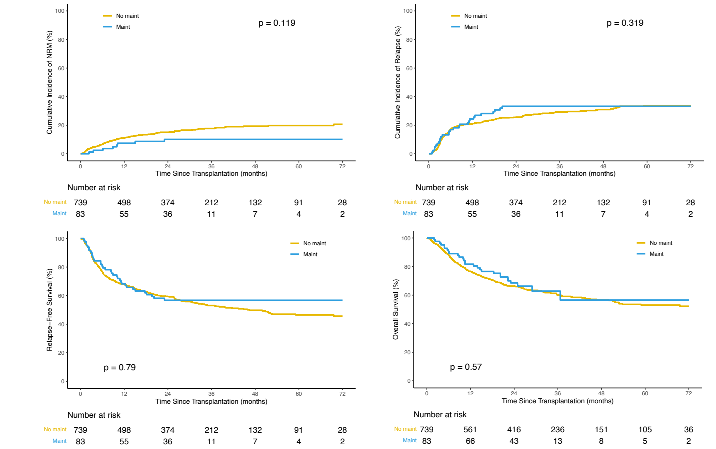


**Baseline *NPM1/FLT3*-ITD, Post-transplant Maintanance**

| Outcomes | Prob (95% CI) | Prob (95% CI) | p-value |
|---|---|---|---|
| NRM | No maint=739 | Maint=83 | 0.119 |
| 3 mo | 4(3-5)% | 1(0-4)% | 0.043 |
| 6 mo | 7(5-9)% | 2(0-6)% | 0.024 |
| 1 yr | 11(9-13)% | 7(2-13)% | 0.25 |
| 2 yr | 15(12-18)% | 10(3-17)% | 0.173 |
| 3 yr | 18(15-21)% | 10(3-17)% | 0.04 |
| Relapse | No maint=739 | Maint=83 | 0.319 |
| 3 mo | 7(5-9)% | 8(2-14)% | 0.633 |
| 6 mo | 17(14-19)% | 17(9-25)% | 0.95 |
| 1 yr | 21(18-24)% | 24(15-34)% | 0.501 |
| 2 yr | 25(22-29)% | 33(23-44)% | 0.161 |
| 3 yr | 29(26-33)% | 33(23-44)% | 0.475 |
| RFS | No maint=739 | Maint=83 | 0.79 |
| 3 mo | 89(87-91)% | 90(82-95)% | 0.7 |
| 6 mo | 77(73-79)% | 81(70-88)% | 0.376 |
| 1 yr | 68(65-71)% | 68(57-77)% | 0.965 |
| 2 yr | 59(56-63)% | 57(45-67)% | 0.636 |
| 3 yr | 53(49-57)% | 57(45-67)% | 0.535 |
| OS | No maint=739 | Maint=83 | 0.57 |
| 3 mo | 95(93-97)% | 98(91-99)% | 0.21 |
| 6 mo | 88(85-90)% | 90(82-95)% | 0.494 |
| 1 yr | 77(73-80)% | 82(71-89)% | 0.275 |
| 2 yr | 66(63-70)% | 69(57-78)% | 0.684 |
| 3 yr | 60(56-64)% | 63(49-74)% | 0.697 |

B

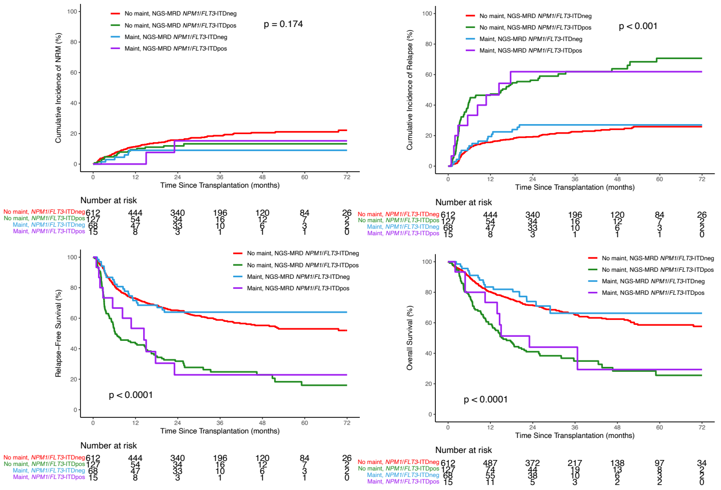


**Baseline *NPM1/FLT3*-ITD, Post-transplant Maintanance & NGS-MRD**

| Outcomes | Prob (95% CI) | Prob (95% CI) | Prob (95% CI) | Prob (95% CI) | p-value |
|---|---|---|---|---|---|
| NRM | No maint, *NPM1*/*FLT3*-ITDneg=612 | No maint, *NPM1*/FLT3-ITDpos=127 | Maint, *NPM1*/*FLT3*-ITDneg=68 | Maint, *NPM1*/*FLT3*-ITDpos=15 | 0.174 |
| 3 mo | 4(3-6)% | 3(0-6)% | 1(0-4)% | 0(0-0)% | <0.001 |
| 6 mo | 7(5-9)% | 6(2-10)% | 3(0-7)% | 0(0-0)% | <0.001 |
| 1 yr | 11(9-14)% | 9(4-15)% | 9(2-16)% | 0(0-0)% | <0.001 |
| 2 yr | 16(13-19)% | 12(6-18)% | 9(2-16)% | 15(0-37)% | 0.287 |
| 3 yr | 19(15-22)% | 13(7-20)% | 9(2-16)% | 15(0-37)% | 0.072 |
| Relapse | No maint, *NPM1*/*FLT3*-ITDneg=612 | No maint, *NPM1*/FLT3-ITDpos=127 | Maint, *NPM1*/*FLT3*-ITDneg=68 | Maint, *NPM1*/*FLT3*-ITDpos=15 | <0.001 |
| 3 mo | 4(2-5)% | 23(16-30)% | 4(0-9)% | 27(3-50)% | <0.001 |
| 6 mo | 11(9-14)% | 43(34-51)% | 13(5-21)% | 33(8-58)% | <0.001 |
| 1 yr | 16(13-18)% | 47(39-56)% | 19(10-29)% | 47(20-73)% | <0.001 |
| 2 yr | 19(16-22)% | 56(48-65)% | 27(16-38)% | 62(35-89)% | <0.001 |
| 3 yr | 23(19-26)% | 62(53-71)% | 27(16-38)% | 62(35-89)% | <0.001 |
| RFS | No maint, *NPM1*/*FLT3*-ITDneg=612 | No maint, *NPM1*/FLT3-ITDpos=127 | Maint, *NPM1*/*FLT3*-ITDneg=68 | Maint, *NPM1*/*FLT3*-ITDpos=15 | <0.001 |
| 3 mo | 92(90-94)% | 74(65-81)% | 94(85-98)% | 73(44-89)% | <0.001 |
| 6 mo | 82(78-85)% | 52(43-60)% | 84(73-91)% | 67(38-85)% | <0.001 |
| 1 yr | 73(69-76)% | 43(35-52)% | 72(59-81)% | 53(26-74)% | <0.001 |
| 2 yr | 65(61-69)% | 32(24-40)% | 64(51-74)% | 23(6-47)% | <0.001 |
| 3 yr | 59(55-63)% | 25(17-33)% | 64(51-74)% | 23(6-47)% | <0.001 |
| OS | No maint, *NPM1*/*FLT3*-ITDneg=612 | No maint, *NPM1*/FLT3-ITDpos=127 | Maint, *NPM1*/*FLT3*-ITDneg=68 | Maint, *NPM1*/*FLT3*-ITDpos=15 | <0.001 |
| 3 mo | 95(93-97)% | 94(89-97)% | 99(90-100)% | 93(61-99)% | 0.241 |
| 6 mo | 90(88-92)% | 77(69-84)% | 93(83-97)% | 80(50-93)% | 0.005 |
| 1 yr | 80(77-83)% | 59(50-67)% | 83(72-90)% | 73(44-89)% | <0.001 |
| 2 yr | 72(68-75)% | 41(32-49)% | 74(61-83)% | 44(19-67)% | <0.001 |
| 3 yr | 65(61-69)% | 35(26-44)% | 66(50-78)% | 44(19-67)% | <0.001 |

**Table S1. Regions of interest in targeted DNA-sequencing panel**

| Gene | Transcript | Exons | ROI Genomic Coordinates (hg19) |
|---|---|---|---|
| **FLT3** | NM_004119 | 14,15,20 | chr13:28592583-28592746<br>chr13:28608003-28608148<br>chr13:28608198-28608371 |
| **IDH1** | NM_005896 | 4 | chr2:209113072-209113404 |
| **IDH2** | NM_002168 | 4,6 | chr15:90630650-90630827<br>chr15:90631798-90631999 |
| **KIT** | NM_000222 | 8,17 | chr4:55589729-55589884<br>chr4:55599215-55599378 |
| **NPM1** | NM_002520 | 11 | chr5:170837510-170837586 |

**Table S2. Sample sequencing summary (n = 1,075)**

| | |
|---|---|
| **Total read pairs** | 15,679,206,759 |
| **Average read pairs per sample** | 12,377,916 ± 4,573,374 |
| **Total mapped read pairs** | 13,280,894,358 |
| **Average mapped read pairs per sample** | 12,354,320 ± 4,602,929 |
| **Average on-target deduplication ratio** | 6.63 ± 2.97 |
| **Average UMI depth per GSP2** | 22,253 ± 5,697 |

### Table S3. Variants detected by next-generation sequencing in the blood of AML patients prior to conditioning

| Subject ID | Cohort | Gene | HGVSc | HGVSp | VAF | Alternate Observations | Deep Alternate Observations | Unique Alternate Observations | Depth | Deep Depth | Variant Consequence | Chromosome | Position | Reference | Alternate | ddPCR Validated | ddPCR VAF |
|---|---|---|---|---|---|---|---|---|---|---|---|---|---|---|---|---|---|
| CH 012119 | 1 | NPM1 | NM_002520.6:c.860_863dup | NP_002511.1:p.Trp288CysfsTer12 | 0.00011 | 4 | 1 | 1 | 37243 | 10054 | frameshift | 5 | 170837543 | C | CTCTG | Yes | 0.00015 |
| CH 012230 | 1 | IDH2 | NM_002168.3:c.419G>A | NP_002159.2:p.Arg140Gln | 0.00747 | 159 | 89 | 72 | 18648 | 11911 | missense | 15 | 90631934 | C | T | Yes | 0.00875 |
| CH 012243 | 1 | IDH1 | NM_005896.3:c.394C>G | NP_005887.2:p.Arg132Gly | 0.25693 | 11012 | 4653 | 700 | 41735 | 18110 | missense | 2 | 209113113 | G | C | Yes | 0.27682 |
| CH 012231 | 1 | FLT3 | NM_004119.2:c.1744_1780dup | NP_004110.2:p.Phe594TyrfsTer6 | 0.00015 | 14 | NA | 12 | 92252 | NA | FLT3-ITD | 13 | 28608275 | A | AAATCAACGTAGAAGTACTCATTATCTGAGGAGCCGGT | Not Done | Not Done |
| CH 012231 | 1 | NPM1 | NM_002520.6:c.860_863dup | NP_002511.1:p.Trp288CysfsTer12 | 0.00015 | 6 | 0 | 4 | 40611 | 7680 | frameshift | 5 | 170837543 | C | CTCTG | Yes | 0.00012 |
| CH 012114 | 1 | IDH2 | NM_002168.3:c.419G>A | NP_002159.2:p.Arg140Gln | 0.00074 | 27 | 11 | 23 | 22547 | 14865 | missense | 15 | 90631934 | C | T | Yes | 0.00120 |
| CH 012020 | 1 | IDH1 | NM_005896.3:c.394C>T | NP_005887.2:p.Arg132Cys | 0.00601 | 280 | 118 | 117 | 36952 | 19630 | missense | 2 | 209113113 | G | A | Yes | 0.00872 |
| CH 012028 | 1 | NPM1 | NM_002520.6:c.860_863dup | NP_002511.1:p.Trp288CysfsTer12 | 0.00107 | 28 | 3 | 19 | 26256 | 7397 | frameshift | 5 | 170837543 | C | CTCTG | Yes | 0.00194 |
| CH 012028 | 1 | FLT3 | NM_004119.2:c.1837+2_1837+3insTACCAGGTGACCGGCTCCTCAGATAATGAGTACTTCTACGTTGATTTCAGAGAATATGAATATGATCTCAAATGGGAGTTTCCAAGAGAAAATTTAGAGTTTGGT | p.? | 0.00021 | 17 | NA | 14 | 80958 | NA | FLT3-ITD | 13 | 28608216 | T | TACCAAACTCTAAATTTTCTCTTGGAAACTCCCATTTGAGATCATATTCATATTCTCTGAAATCAACGTAGAAGTACTCATTATCTGAGGAGCCGGTCACCTGGTA | Not Done | Not Done |
| CH 012019 | 1 | IDH2 | NM_002168.3:c.515G>A | NP_002159.2:p.Arg172Lys | 0.00784 | 185 | 101 | 88 | 20376 | 12878 | missense | 15 | 90631838 | C | T | Yes | 0.01096 |
| CH 012012 | 1 | FLT3 | NM_004119.2:c.1779_1811dup | NP_004110.2:p.Asp593_Trp603dup | 0.00157 | 131 | NA | 70 | 83574 | NA | FLT3-ITD | 13 | 28608246 | C | CCCATTTGAGATCATATTCATATTCTCTGAAATC | Not Done | Not Done |
| CH 012013 | 1 | KIT | NM_000222.2:c.2466T>A | NP_000213.1:p.Asn822Lys | 0.00678 | 413 | 143 | 186 | 47724 | 21107 | missense | 4 | 55599340 | T | A | Not Done | Not Done |
| CH 012061 | 1 | NPM1 | NM_002520.6:c.860_863dup | NP_002511.1:p.Trp288CysfsTer12 | 0.00328 | 121 | 7 | 61 | 36852 | 8075 | frameshift | 5 | 170837543 | C | CTCTG | Yes | 0.00381 |
| CH 012042 | 1 | NPM1 | NM_002520.6:c.860_863dup | NP_002511.1:p.Trp288CysfsTer12 | 0.01009 | 319 | 39 | 125 | 31628 | 9001 | frameshift | 5 | 170837543 | C | CTCTG | Yes | 0.01198 |
| CH 012042 | 1 | IDH1 | NM_005896.3:c.394C>T | NP_005887.2:p.Arg132Cys | 0.00987 | 405 | 193 | 153 | 37312 | 19550 | missense | 2 | 209113113 | G | A | Yes | 0.01311 |
| CH 012043 | 1 | IDH1 | NM_005896.3:c.394C>T | NP_005887.2:p.Arg132Cys | 0.00519 | 182 | 93 | 93 | 32538 | 17930 | missense | 2 | 209113113 | G | A | Yes | 0.00624 |
| CH 012057 | 1 | NPM1 | NM_002520.6:c.863_864insCATG | NP_002511.1:p.Trp288CysfsTer12 | 0.00019 | 6 | 2 | 4 | 31565 | 10884 | frameshift | 5 | 170837545 | C | CTGCA | Yes | 0.00012 |
| CH 012057 | 1 | FLT3 | NM_004119.2:c.1789_1821dup | NP_004110.2:p.Tyr597_Arg607dup | 0.00011 | 8 | NA | 7 | 75896 | NA | FLT3-ITD | 13 | 28608235 | T | TCTTGGAAACTCCCATTTGAGATCATATTCATAT | Not Done | Not Done |
| CH 012062 | 1 | NPM1 | NM_002520.6:c.860_863dup | NP_002511.1:p.Trp288CysfsTer12 | 0.00040 | 10 | 2 | 8 | 25198 | 9902 | frameshift | 5 | 170837543 | C | CTCTG | Yes | 0.00058 |
| CH 012062 | 1 | FLT3 | NM_004119.2:c.1747_1782dup | NP_004110.2:p.Gly583_Phe594dup | 0.00066 | 35 | NA | 20 | 52708 | NA | FLT3-ITD | 13 | 28608274 | G | GAAATCAACGTAGAAGTACTCATTATCTGAGGAGCCG | Not Done | Not Done |
| CH 012078 | 1 | FLT3 | NM_004119.2:c.1752_1817dup | NP_004110.2:p.Ser585_Pro606dup | 0.00078 | 48 | NA | 38 | 61563 | NA | FLT3-ITD | 13 | 28608239 | G | GGAAACTCCCATTTGAGATCATATTCATATTCTCTGAAATCAACGTAGAAGTACTCATTATCTGAGG | Not Done | Not Done |
| CH 012555 | 1 | IDH1 | NM_005896.3:c.394C>G | NP_005887.2:p.Arg132Gly | 0.04814 | 1426 | 682 | 354 | 29094 | 14168 | missense | 2 | 209113113 | G | C | Yes | 0.05669 |
| CH 012493 | 1 | NPM1 | NM_002520.6:c.860_863dup | NP_002511.1:p.Trp288CysfsTer12 | 0.00100 | 22 | 0 | 16 | 21997 | 7524 | frameshift | 5 | 170837543 | C | CTCTG | Yes | 0.00134 |
| CH 012493 | 1 | FLT3 | NM_004119.2:c.1754_1795dup | NP_004110.2:p.Ser585_Glu598dup | 0.00081 | 44 | NA | 30 | 54189 | NA | FLT3-ITD | 13 | 28608261 | A | ATTCATATTCTCTGAAATCAACGTAGAAGTACTCATTATCTGA | Not Done | Not Done |
| CH 012512 | 1 | IDH2 | NM_002168.3:c.418C>T | NP_002159.2:p.Arg140Trp | 0.00336 | 68 | 33 | 41 | 16062 | 9811 | missense | 15 | 90631935 | G | A | Yes | 0.00463 |
| CH 012505 | 1 | IDH1 | NM_005896.3:c.394C>A | NP_005887.2:p.Arg132Ser | 0.21037 | 6529 | 3600 | 562 | 30474 | 17113 | missense | 2 | 209113113 | G | T | Yes | 0.21690 |
| CH 012515 | 1 | IDH2 | NM_002168.3:c.419G>A | NP_002159.2:p.Arg140Gln | 0.28177 | 4924 | 3290 | 432 | 17187 | 11676 | missense | 15 | 90631934 | C | T | Yes | 0.29130 |
| CH 012541 | 1 | IDH2 | NM_002168.3:c.419G>A | NP_002159.2:p.Arg140Gln | 0.00557 | 131 | 55 | 64 | 18810 | 9873 | missense | 15 | 90631934 | C | T | Yes | 0.00460 |
| CH 012209 | 1 | FLT3 | NM_004119.2:c.1794_1795insCCTCAT | NP_004110.2:p.Glu598_Tyr599insProHis | 0.00060 | 27 | 6 | 23 | 44942 | 13378 | FLT3-ITD | 13 | 28608261 | A | AATGAGG | Not Done | Not Done |
| CH 012215 | 1 | NPM1 | NM_002520.6:c.860_863dup | NP_002511.1:p.Trp288CysfsTer12 | 0.01295 | 398 | 49 | 156 | 30732 | 9124 | frameshift | 5 | 170837543 | C | CTCTG | Yes | 0.01365 |
| CH 012215 | 1 | FLT3 | NM_004119.2:c.1770_1793dup | NP_004110.2:p.Tyr597_Glu598insAspTyrValAspPheArgGluTyr | 0.03500 | 2547 | NA | 258 | 72769 | NA | FLT3-ITD | 13 | 28608262 | T | TTCATATTCTCTGAAATCAACGTAG | Not Done | Not Done |
| CH 012215 | 1 | FLT3 | NM_004119.2:c.1721_1825dup | NP_004110.2:p.Ser574_Glu608dup | 0.00314 | 248 | NA | 76 | 79104 | NA | FLT3-ITD | 13 | 28608234 | C | CTCTTGGAAACTCCCATTTGAGATCATATTCATATTCTCTGAAATCAACGTAGAAGTACTCATTATCTGAGGAGCCGGTCACCTGTACCATCTGTAGCTGGCTTTC | Not Done | Not Done |
| CH 012232 | 1 | NPM1 | NM_002520.6:c.863_864insCATG | NP_002511.1:p.Trp288CysfsTer12 | 0.00020 | 6 | 0 | 5 | 30498 | 8184 | frameshift | 5 | 170837545 | C | CTGCA | Yes | 0.00018 |
| CH 012232 | 1 | FLT3 | NM_004119.2:c.1759_1791dup | NP_004110.2:p.Asn587_Tyr597dup | 0.00018 | 13 | NA | 8 | 74031 | NA | FLT3-ITD | 13 | 28608266 | T | TATTCTCTGAAATCAACGTAGAAGTACTCATTAT | Not Done | Not Done |
| CH 012238 | 1 | IDH2 | NM_002168.3:c.419G>A | NP_002159.2:p.Arg140Gln | 0.01328 | 344 | 198 | 126 | 23449 | 14907 | missense | 15 | 90631934 | C | T | Yes | 0.01490 |
| CH 012247 | 1 | IDH1 | NM_005896.3:c.395G>A | NP_005887.2:p.Arg132His | 0.01237 | 572 | 170 | 207 | 40301 | 13742 | missense | 2 | 209113112 | C | T | Yes | 0.01460 |
| CH 012183 | 1 | NPM1 | NM_002520.6:c.860_863dup | NP_002511.1:p.Trp288CysfsTer12 | 0.00023 | 7 | 1 | 4 | 30677 | 11099 | frameshift | 5 | 170837543 | C | CTCTG | Yes | 0.00014 |
| CH 012185 | 1 | IDH1 | NM_005896.3:c.394C>T | NP_005887.2:p.Arg132Cys | 0.00696 | 253 | 121 | 108 | 32807 | 17390 | missense | 2 | 209113113 | G | A | Yes | 0.00821 |
| CH 012189 | 1 | FLT3 | NM_004119.2:c.1787_1788insGGCCCCTGATTTCAGAGA | NP_004110.2:p.Glu596_Tyr597insAlaProAspPheArgGlu | 0.00022 | 13 | NA | 7 | 58424 | NA | FLT3-ITD | 13 | 28608268 | T | TTCTCTGAAATCAGGGGCC | Not Done | Not Done |
| CH 012210 | 1 | FLT3 | NM_004119.2:c.2503G>T | NP_004110.2:p.Asp835Tyr | 0.62383 | 13479 | 7441 | 767 | 21788 | 11928 | missense | 13 | 28592642 | C | A | Yes | 0.60500 |
| CH 012301 | 1 | NPM1 | NM_002520.6:c.863_864insCATG | NP_002511.1:p.Trp288CysfsTer12 | 0.00135 | 41 | 7 | 30 | 30341 | 6684 | frameshift | 5 | 170837545 | C | CTGCA | Yes | 0.00179 |
| CH 012301 | 1 | FLT3 | NM_004119.2:c.1833_1834insCCCACTTTCAGAGAATATGAATATGATCTCAAATGGGAGTTTCCAAGAGAAAATTTAGAG | NP_004110.2:p.Glu611_Phe612insProThrPheArgGluTyrGluTyrAspLeuLysTrpGluPheProArgGluAsnLeuGlu | 0.00079 | 80 | NA | 57 | 101062 | NA | FLT3-ITD | 13 | 28608222 | A | ACTCTAAATTTTCTCTTGGAAACTCCCATTTGAGATCATATTCATATTCTCTGAAAGTGGG | Not Done | Not Done |
| CH 012237 | 1 | IDH1 | NM_005896.3:c.394C>G | NP_005887.2:p.Arg132Gly | 0.01911 | 1013 | 386 | 298 | 47201 | 20194 | missense | 2 | 209113113 | G | C | Yes | 0.02237 |
| CH 012284 | 1 | IDH1 | NM_005896.3:c.395G>A | NP_005887.2:p.Arg132His | 0.00292 | 152 | 47 | 78 | 40470 | 16071 | missense | 2 | 209113112 | C | T | Yes | 0.00400 |
| CH 012254 | 1 | FLT3 | NM_004119.2:c.2508_2510del | NP_004110.2:p.Ile836del | 0.00343 | 105 | 45 | 64 | 30592 | 13116 | deletion | 13 | 28592634 | CATG | C | Not Done | Not Done |
| CH 012555 | 1 | FLT3 | NM_004119.2:c.1838-5_1838-4insCCTTCGGCTCCTCAGATAATGAGTACTTCTACGTTGATTTCAGAGAATATGAATATGATCTCAAATGGGAGTTTCCAAGAGAAAATTTAGAGTTTGGTAAGAATGGAATGTGCCAAATGTTTCTGCAGCATTTCTTTTCCATTGGAAAATCTTTAAAATGCACGTACTCACCATTTGTCTTT | p.? | 0.00210 | 169 | NA | 47 | 80376 | NA | FLT3-ITD | 13 | 28608132 | C | CAAAGACAAATGGTGAGTACGTGCATTTTAAAGATTTTCCAATGGAAAAGAAATGCTGCAGAAACATTTGGCACATTCCATTCTTACCAAACTCTAAATTTTCTCTTGGAAACTCCCATTTGAGATCATATTCATATTCTCTGAAATCAACGTAGAAGTACTCATTATCTGAGGAGCCGAAGG | Not Done | Not Done |
| CH 012100 | 1 | IDH2 | NM_002168.3:c.515G>A | NP_002159.2:p.Arg172Lys | 0.00141 | 31 | 18 | 24 | 19024 | 12810 | missense | 15 | 90631838 | C | T | Yes | 0.00149 |
| CH 012123 | 1 | IDH1 | NM_005896.3:c.394C>A | NP_005887.2:p.Arg132Ser | 0.27757 | 11133 | 4568 | 656 | 38456 | 16457 | missense | 2 | 209113113 | G | T | Yes | 0.29504 |
| CH 012139 | 1 | IDH1 | NM_005896.3:c.394C>T | NP_005887.2:p.Arg132Cys | 0.01425 | 496 | 203 | 179 | 28504 | 14249 | missense | 2 | 209113113 | G | A | Yes | 0.01799 |
| CH 012288 | 1 | NPM1 | NM_002520.6:c.860_863dup | NP_002511.1:p.Trp288CysfsTer12 | 0.00032 | 9 | 4 | 9 | 28465 | 7837 | frameshift | 5 | 170837543 | C | CTCTG | Yes | 0.00037 |
| CH 012288 | 1 | FLT3 | NM_004119.2:c.1807_1808insCCGAATATGAATATGATCTCAAAT | NP_004110.2:p.Lys602_Trp603insSerGluTyrGluTyrAspLeuLys | 0.06501 | 3673 | NA | 224 | 56496 | NA | FLT3-ITD | 13 | 28608248 | C | CATTTGAGATCATATTCATATTCGG | Not Done | Not Done |
| CH 012289 | 1 | NPM1 | NM_002520.6:c.860_863dup | NP_002511.1:p.Trp288CysfsTer12 | 0.00511 | 149 | 17 | 75 | 29176 | 7465 | frameshift | 5 | 170837543 | C | CTCTG | Yes | 0.00469 |
| CH 012323 | 1 | IDH2 | NM_002168.3:c.419G>A | NP_002159.2:p.Arg140Gln | 0.00128 | 51 | 22 | 32 | 26902 | 17214 | missense | 15 | 90631934 | C | T | Yes | 0.00192 |
| CH 012326 | 1 | IDH2 | NM_002168.3:c.419G>A | NP_002159.2:p.Arg140Gln | 0.22757 | 3460 | 2268 | 389 | 15088 | 9966 | missense | 15 | 90631934 | C | T | Yes | 0.23657 |
| CH 012337 | 1 | FLT3 | NM_004119.2:c.1837+4_1837+5insCCTTCGGCTCCTCAGATAATGAGTACTTCTACGTTGATTTCAGAGAATATGAATATGATCTCAAATGGGAGTTTCCAAGAGAAAATTTAGAGTTTGGTAAGAATGGAATGTGCCAAATGTTTCTGCAGCATTTCTTTTCCATTGGAAAATCTTTAAAATGCACGTACTCACCATTTGTCTTT | p.? | 0.00036 | 37 | NA | 18 | 103582 | NA | FLT3-ITD | 13 | 28608214 | C | CAAAGACAAATGGTGAGTACGTGCATTTTAAAGATTTTCCAATGGAAAAGAAATGCTGCAGAAACATTTGGCACATTCCATTCTTACCAAACTCTAAATTTTCTCTTGGAAACTCCCATTTGAGATCATATTCATATTCTCTGAAATCAACGTAGAAGTACTCATTATCTGAGGAGCCGAAGG | Not Done | Not Done |
| CH 012329 | 1 | IDH2 | NM_002168.3:c.419G>A | NP_002159.2:p.Arg140Gln | 0.35750 | 5144 | 3368 | 469 | 14452 | 9421 | missense | 15 | 90631934 | C | T | Yes | 0.36602 |
| CH 012327 | 1 | IDH2 | NM_002168.3:c.419G>A | NP_002159.2:p.Arg140Gln | 0.00194 | 42 | 22 | 26 | 17620 | 11313 | missense | 15 | 90631934 | C | T | Yes | 0.00284 |
| CH 012346 | 1 | FLT3 | NM_004119.2:c.1775_1837+3dup | p.? | 0.00025 | 29 | NA | 23 | 118076 | NA | FLT3-ITD | 13 | 28608215 | T | TTACCAAACTCTAAATTTTCTCTTGGAAACTCCCATTTGAGATCATATTCATATTCTCTGAAATCAA | Not Done | Not Done |
| CH 012366 | 1 | FLT3 | NM_004119.2:c.1813_1814insGGGGCTATGATCTCAAATGGGAGT | NP_004110.2:p.Glu604_Phe605insTrpGlyTyrAspLeuLysTrpGlu | 0.00690 | 440 | NA | 67 | 63794 | NA | FLT3-ITD | 13 | 28608242 | A | AACTCCCATTTGAGATCATAGCCCC | Not Done | Not Done |
| CH 012367 | 1 | NPM1 | NM_002520.6:c.860_863dup | NP_002511.1:p.Trp288CysfsTer12 | 0.00530 | 185 | 14 | 95 | 34909 | 7295 | frameshift | 5 | 170837543 | C | CTCTG | Yes | 0.00472 |
| CH 012367 | 1 | IDH2 | NM_002168.3:c.419G>A | NP_002159.2:p.Arg140Gln | 0.00531 | 137 | 85 | 74 | 24740 | 15993 | missense | 15 | 90631934 | C | T | Yes | 0.00570 |

| Subject ID | Cohort | Gene | HGVSc | HGVSp | VAF | Alternate Observations | Deep Alternate Observations | Unique Alternate Observations | Depth | Deep Depth | Variant Consequence | Chromosome | Position | Reference | Alternate | ddPCR Validated | ddPCR VAF |
|---|---|---|---|---|---|---|---|---|---|---|---|---|---|---|---|---|---|
| CH 012372 | 1 | NPM1 | NM_002520.6:c.860_863dup | NP_002511.1:p.Trp288CysfsTer12 | 0.00013 | 3 | 0 | 1 | 23378 | 4515 | frameshift | 5 | 170837543 | C | CTCTG | Yes | 0.00011 |
| CH 012379 | 1 | NPM1 | NM_002520.6:c.863_864insCCTG | NP_002511.1:p.Trp288CysfsTer12 | 0.00017 | 6 | 0 | 4 | 35578 | 5091 | frameshift | 5 | 170837544 | T | TCTGC | Yes | 0.00005 |
| CH 012397 | 1 | NPM1 | NM_002520.6:c.860_863dup | NP_002511.1:p.Trp288CysfsTer12 | 0.00171 | 65 | 9 | 42 | 38027 | 11794 | frameshift | 5 | 170837543 | C | CTCTG | Yes | 0.00118 |
| CH 012397 | 1 | FLT3 | NM_004119.2:c.1782_1783insCCAGATAATGAGTACTTCTACGTTGATTTC | NP_004110.2:p.Phe594_Arg595insProAspAsnGluTyrPheTyrValAspPhe | 0.00126 | 110 | NA | 66 | 87194 | NA | FLT3-ITD | 13 | 28608273 | T | TGAAATCAACGTAGAAGTACTCATTATCTGG | Not Done | Not Done |
| CH 012397 | 1 | FLT3 | NM_004119.2:c.1753_1782dup | NP_004110.2:p.Ser585_Phe594dup | 0.00067 | 61 | NA | 38 | 90587 | NA | FLT3-ITD | 13 | 28608274 | G | GAAATCAACGTAGAAGTACTCATTATCTGAG | Not Done | Not Done |
| CH 012444 | 1 | NPM1 | NM_002520.6:c.860_863dup | NP_002511.1:p.Trp288CysfsTer12 | 0.00014 | 4 | 0 | 4 | 28192 | 4048 | frameshift | 5 | 170837543 | C | CTCTG | Yes | 0.00028 |
| CH 012416 | 1 | IDH2 | NM_002168.3:c.419G>A | NP_002159.2:p.Arg140Gln | 0.36639 | 8997 | 5320 | 622 | 24225 | 14520 | missense | 15 | 90631934 | C | T | Yes | 0.37451 |
| CH 012478 | 1 | FLT3 | NM_004119.2:c.1781_1807dup | NP_004110.2:p.Phe594_Lys602dup | 0.00012 | 12 | NA | 9 | 96369 | NA | FLT3-ITD | 13 | 28608249 | A | ATTTGAGATCATATTCATATTCTCTGAA | Not Done | Not Done |
| CH 012434 | 1 | IDH1 | NM_005896.3:c.395G>A | NP_005887.2:p.Arg132His | 0.00233 | 119 | 33 | 71 | 36516 | 14175 | missense | 2 | 209113112 | C | T | Yes | 0.00330 |
| CH 012420 | 1 | NPM1 | NM_002520.6:c.860_863dup | NP_002511.1:p.Trp288CysfsTer12 | 0.02549 | 863 | 51 | 256 | 33863 | 5492 | frameshift | 5 | 170837543 | C | CTCTG | Yes | 0.02333 |
| CH 012420 | 1 | FLT3 | NM_004119.2:c.1779_1805dup | NP_004110.2:p.Leu601_Lys602insAsnPheArgGluTyrGluTyrAspLeu | 0.01514 | 1544 | NA | 388 | 101970 | NA | FLT3-ITD | 13 | 28608251 | T | TTGAGATCATATTCATATTCTCTGAAAT | Not Done | Not Done |
| CH 012420 | 1 | FLT3 | NM_004119.2:c.1779_1805dup | NP_004110.2:p.Leu601_Lys602insAsnPheArgGluTyrGluTyrAspLeu | 0.00014 | 14 | NA | 8 | 102323 | NA | FLT3-ITD | 13 | 28608250 | T | TTTGAGATCATATTCATATTCTCTGAAA | Not Done | Not Done |
| CH 012422 | 1 | IDH1 | NM_005896.3:c.395G>A | NP_005887.2:p.Arg132His | 0.00162 | 89 | 30 | 60 | 43645 | 18519 | missense | 2 | 209113112 | C | T | Yes | 0.00220 |
| CH 012422 | 1 | IDH2 | NM_002168.3:c.419G>A | NP_002159.2:p.Arg140Gln | 0.00298 | 87 | 47 | 50 | 23494 | 15776 | missense | 15 | 90631934 | C | T | Yes | 0.00382 |
| CH 012422 | 1 | IDH1 | NM_005896.3:c.394C>T | NP_005887.2:p.Arg132Cys | 0.00276 | 159 | 51 | 88 | 43634 | 18458 | missense | 2 | 209113113 | G | A | Yes | 0.00364 |
| CH 012418 | 1 | NPM1 | NM_002520.6:c.863_864insCCAG | NP_002511.1:p.Trp288CysfsTer12 | 0.00022 | 7 | 1 | 5 | 32176 | 4816 | frameshift | 5 | 170837546 | T | TGCCA | Not Done | Not Done |
| CH 012418 | 1 | FLT3 | NM_004119.2:c.1782_1829dup | NP_004110.2:p.Phe594_Asn609dup | 0.00013 | 13 | NA | 8 | 101879 | NA | FLT3-ITD | 13 | 28608230 | T | TTTTCTCTTGGAAACTCCCATTTGAGATCATATTCATATTCTCTGAAAT | Not Done | Not Done |
| CH 012442 | 1 | IDH1 | NM_005896.3:c.395G>A | NP_005887.2:p.Arg132His | 0.00818 | 438 | 185 | 168 | 50814 | 22624 | missense | 2 | 209113112 | C | T | Yes | 0.00942 |
| CH 012449 | 1 | FLT3 | NM_004119.2:c.2503G>T | NP_004110.2:p.Asp835Tyr | 0.00717 | 212 | 106 | 110 | 28687 | 14782 | missense | 13 | 28592642 | C | A | Yes | 0.00848 |
| CH 012449 | 1 | FLT3 | NM_004119.2:c.2524T>C | NP_004110.2:p.Tyr842His | 0.00085 | 55 | 13 | 44 | 29414 | 15220 | missense | 13 | 28592621 | A | G | Not Done | Not Done |
| CH 012449 | 1 | FLT3 | NM_004119.2:c.1837_1837+1insTCCAGGACTTCTACGTTGATTTCAGAGAATATGAATATGATCTCAAATGGGAGTTTCCAAGAGAAAATTTAGAGTTTG | NP_004110.2:p.Phe612_Gly613insValGlnAspPheTyrValAspPheArgGluTyrGluTyrAspLeuLysTrpGluPheProArgGluAsnLeuGluPhe | 0.09582 | 10214 | NA | 467 | 106598 | NA | FLT3-ITD | 13 | 28608218 | C | CCAAACTCTAAATTTTCTCTTGGAAACTCCCATTTGAGATCATATTCATATTCTCTGAAATCAACGTAGAAGTCCTGGA | Not Done | Not Done |
| CH 012449 | 1 | FLT3 | NM_004119.2:c.1816_1817insAAAGAGTTCAGGTGACCGGCTCCTCAGATAATGAGTACTTCTACGTTGATTTCAGAGAATATGAATATGATCTCAAATGGGAGTTTC | NP_004110.2:p.Phe605_Pro606insGlnArgValGlnValThrGlySerSerAspAsnGluTyrPheTyrValAspPheArgGluTyrGluTyrAspLeuLysTrpGluPhe | 0.02087 | 1649 | NA | 249 | 78997 | NA | FLT3-ITD | 13 | 28608239 | G | GGAAACTCCCATTTGAGATCATATTCATATTCTCTGAAATCAACGTAGAAGTACTCATTATCTGAGGAGCCGGTCACCTGAACTCTTT | Not Done | Not Done |
| CH 012446 | 1 | NPM1 | NM_002520.6:c.860_863dup | NP_002511.1:p.Trp288CysfsTer12 | 0.00031 | 9 | 0 | 4 | 29363 | 5192 | frameshift | 5 | 170837543 | C | CTCTG | Yes | 0.00020 |
| CH 012485 | 1 | FLT3 | NM_004119.2:c.1802_1816dup | NP_004110.2:p.Leu601_Phe605dup | 0.00177 | 93 | 25 | 59 | 52520 | 13058 | FLT3-ITD | 13 | 28608239 | G | GGAAACTCCCATTTGA | Not Done | Not Done |
| CH 012485 | 1 | NPM1 | NM_002520.6:c.860_863dup | NP_002511.1:p.Trp288CysfsTer12 | 0.02950 | 801 | 33 | 208 | 27151 | 3580 | frameshift | 5 | 170837543 | C | CTCTG | Yes | 0.03020 |
| CH 012485 | 1 | FLT3 | NM_004119.2:c.1738_1837+20dup | p.? | 0.01107 | 1403 | NA | 259 | 126683 | NA | FLT3-ITD | 13 | 28608199 | T | TTGGCACATTCCATTCTTACCAAACTCTAAATTTTCTCTTGGAAACTCCCATTTGAGATCATATTCATATTCTCTGAAATCAACGTAGAAGTACTCATTATCTGAGGAGCCGGTCACCTGT | Not Done | Not Done |
| CH 012485 | 1 | FLT3 | NM_004119.2:c.1738_1837+20dup | p.? | 0.00242 | 314 | NA | 73 | 129848 | NA | FLT3-ITD | 13 | 28608198 | T | TTTGGCACATTCCATTCTTACCAAACTCTAAATTTTCTCTTGGAAACTCCCATTTGAGATCATATTCATATTCTCTGAAATCAACGTAGAAGTACTCATTATCTGAGGAGCCGGTCACCTG | Not Done | Not Done |
| CH 012490 | 1 | IDH2 | NM_002168.3:c.419G>A | NP_002159.2:p.Arg140Gln | 0.03804 | 973 | 497 | 269 | 23680 | 13065 | missense | 15 | 90631934 | C | T | Yes | 0.03784 |
| CH 012568 | 1 | IDH2 | NM_002168.3:c.419G>A | NP_002159.2:p.Arg140Gln | 0.01146 | 163 | 86 | 63 | 14227 | 7506 | missense | 15 | 90631934 | C | T | Yes | 0.00849 |
| CH 012500 | 1 | FLT3 | NM_004119.2:c.1784_1804dup | NP_004110.2:p.Arg595_Leu601dup | 0.23355 | 7809 | 2273 | 542 | 33436 | 10098 | FLT3-ITD | 13 | 28608251 | T | TTGAGATCATATTCATATTCTC | Not Done | Not Done |
| CH 012500 | 1 | KIT | NM_000222.2:c.2447A>T | NP_000213.1:p.Asp816Val | 0.01298 | 473 | 127 | 185 | 32065 | 9785 | missense | 4 | 55599321 | A | T | Yes | 0.01419 |
| CH 012476 | 1 | IDH1 | NM_005896.3:c.394C>G | NP_005887.2:p.Arg132Gly | 0.33789 | 12299 | 3240 | 700 | 35090 | 9589 | missense | 2 | 209113113 | G | C | Yes | 0.34389 |
| CH 012169 | 1 | FLT3 | NM_004119.2:c.2482T>G | NP_004110.2:p.Cys828Gly | 0.00163 | 30 | 15 | 23 | 19791 | 9212 | missense | 13 | 28592663 | A | C | Not Done | Not Done |
| CH 012145 | 1 | IDH2 | NM_002168.3:c.419G>A | NP_002159.2:p.Arg140Gln | 0.00270 | 73 | 32 | 38 | 22994 | 11868 | missense | 15 | 90631934 | C | T | Yes | 0.00354 |
| CH 012173 | 1 | IDH2 | NM_002168.3:c.419G>A | NP_002159.2:p.Arg140Gln | 0.00228 | 52 | 24 | 35 | 20368 | 10532 | missense | 15 | 90631934 | C | T | Yes | 0.00262 |
| CH 012159 | 1 | FLT3 | NM_004119.2:c.1777_1824dup | NP_004110.2:p.Asp593_Glu608dup | 0.00042 | 37 | NA | 24 | 87849 | NA | FLT3-ITD | 13 | 28608231 | T | TTTCTCTTGGAAACTCCCATTTGAGATCATATTCATATTCTCTGAAATC | Not Done | Not Done |
| CH 012050 | 1 | FLT3 | NM_004119.2:c.1746_1801dup | NP_004110.2:p.Leu601ProfsTer29 | 0.00064 | 36 | NA | 19 | 56304 | NA | FLT3-ITD | 13 | 28608255 | G | GATCATATTCATATTCTCTGAAATCAACGTAGAAGTACTCATTATCTGAGGAGCCGG | Not Done | Not Done |
| CH 012029 | 1 | IDH2 | NM_002168.3:c.419G>A | NP_002159.2:p.Arg140Gln | 0.00112 | 28 | 9 | 19 | 13583 | 8026 | missense | 15 | 90631934 | C | T | Yes | 0.00140 |
| CH 012044 | 1 | IDH2 | NM_002168.3:c.419G>A | NP_002159.2:p.Arg140Gln | 0.00122 | 28 | 14 | 21 | 17808 | 11476 | missense | 15 | 90631934 | C | T | Yes | 0.00135 |
| CH 012046 | 1 | IDH2 | NM_002168.3:c.419G>A | NP_002159.2:p.Arg140Gln | 0.04451 | 528 | 348 | 169 | 12086 | 7818 | missense | 15 | 90631934 | C | T | Yes | 0.04464 |
| CH 012084 | 1 | FLT3 | NM_004119.2:c.1760_1837+21dup | p.? | 0.01877 | 1761 | NA | 236 | 93797 | NA | FLT3-ITD | 13 | 28608198 | T | TTTGGCACATTCCATTCTTACCAAACTCTAAATTTTCTCTTGGAAACTCCCATTTGAGATCATATTCATATTCTCTGAAATCAACGTAGAAGTACTCATT | Not Done | Not Done |
| CH 012084 | 1 | FLT3 | NM_004119.2:c.1759_1797dup | NP_004110.2:p.Asn587_Tyr599dup | 0.02029 | 1141 | NA | 333 | 56241 | NA | FLT3-ITD | 13 | 28608260 | T | TATTCATATTCTCTGAAATCAACGTAGAAGTACTCATTAT | Not Done | Not Done |
| CH 012084 | 1 | FLT3 | NM_004119.2:c.1734_1805dup | NP_004110.2:p.Val579_Lys602dup | 0.01474 | 885 | NA | 136 | 60047 | NA | FLT3-ITD | 13 | 28608250 | T | TTTGAGATCATATTCATATTCTCTGAAATCAACGTAGAAGTACTCATTATCTGAGGAGCCGGTCACCTGTACC | Not Done | Not Done |
| CH 012084 | 1 | FLT3 | NM_004119.2:c.1790_1834dup | NP_004110.2:p.Tyr597_Glu611dup | 0.00188 | 169 | NA | 73 | 90081 | NA | FLT3-ITD | 13 | 28608221 | A | AACTCTAAATTTTCTCTTGGAAACTCCCATTTGAGATCATATTCAT | Not Done | Not Done |
| CH 012090 | 1 | IDH2 | NM_002168.3:c.419G>A | NP_002159.2:p.Arg140Gln | 0.00160 | 30 | 16 | 22 | 15073 | 9969 | missense | 15 | 90631934 | C | T | Yes | 0.00111 |
| CH 012560 | 1 | FLT3 | NM_004119.2:c.1747_1788dup | NP_004110.2:p.Gly583_Glu596dup | 0.00033 | 21 | NA | 13 | 64351 | NA | FLT3-ITD | 13 | 28608267 | A | ATTCTCTGAAATCAACGTAGAAGTACTCATTATCTGAGGAGCC | Not Done | Not Done |
| CH 012554 | 1 | IDH2 | NM_002168.3:c.419G>A | NP_002159.2:p.Arg140Gln | 0.44521 | 7951 | 4908 | 472 | 17801 | 11024 | missense | 15 | 90631934 | C | T | Yes | 0.45490 |
| CH 012517 | 1 | IDH2 | NM_002168.3:c.419G>A | NP_002159.2:p.Arg140Gln | 0.00365 | 50 | 35 | 34 | 14374 | 9584 | missense | 15 | 90631934 | C | T | Yes | 0.00580 |
| CH 012517 | 1 | FLT3 | NM_004119.2:c.1794_1795insCCATACGTTGATTTCAGAGAATATGAA | NP_004110.2:p.Glu598_Tyr599insProTyrValAspPheArgGluTyrGlu | 0.00032 | 16 | NA | 9 | 50709 | NA | FLT3-ITD | 13 | 28608261 | A | ATTCATATTCTCTGAAATCAACGTATGG | Not Done | Not Done |
| CH 012497 | 1 | IDH1 | NM_005896.3:c.394C>A | NP_005887.2:p.Arg132Ser | 0.01335 | 463 | 176 | 177 | 31803 | 13184 | missense | 2 | 209113113 | G | T | Yes | 0.01509 |
| CH 012520 | 1 | IDH2 | NM_002168.3:c.515G>A | NP_002159.2:p.Arg172Lys | 0.00098 | 17 | 10 | 14 | 15808 | 10222 | missense | 15 | 90631838 | C | T | Yes | 0.00086 |
| CH 012529 | 1 | IDH2 | NM_002168.3:c.419G>A | NP_002159.2:p.Arg140Gln | 0.11759 | 1717 | 1062 | 315 | 14434 | 9031 | missense | 15 | 90631934 | C | T | Yes | 0.12136 |
| CH 012565 | 1 | FLT3 | NM_004119.2:c.1779_1793dup | NP_004110.2:p.Asp593_Tyr597dup | 0.00072 | 28 | 12 | 24 | 38936 | 13396 | FLT3-ITD | 13 | 28608262 | T | TTCATATTCTCTGAAA | Not Done | Not Done |
| CH 012565 | 1 | FLT3 | NM_004119.2:c.1714_1803dup | NP_004110.2:p.Tyr572_Leu601dup | 0.00018 | 11 | NA | 8 | 60283 | NA | FLT3-ITD | 13 | 28608252 | T | TGAGATCATATTCATATTCTCTGAAATCAACGTAGAAGTACTCATTATCTGAGGAGCCGGTCACCTGTACCATCTGTAGCTGGCTTTCATA | Not Done | Not Done |
| CH 012528 | 1 | NPM1 | NM_002520.6:c.860_863dup | NP_002511.1:p.Trp288CysfsTer12 | 0.00980 | 235 | 16 | 91 | 23979 | 5650 | frameshift | 5 | 170837543 | C | CTCTG | Yes | 0.01095 |
| CH 012528 | 1 | FLT3 | NM_004119.2:c.1758_1811dup | NP_004110.2:p.Asp586_Trp603dup | 0.03263 | 1929 | NA | 353 | 59117 | NA | FLT3-ITD | 13 | 28608246 | C | CCCATTTGAGATCATATTCATATTCTCTGAAATCAACGTAGAAGTACTCATTATC | Not Done | Not Done |
| CH 012546 | 1 | NPM1 | NM_002520.6:c.860_863dup | NP_002511.1:p.Trp288CysfsTer12 | 0.00021 | 5 | 0 | 3 | 23886 | 7340 | frameshift | 5 | 170837543 | C | CTCTG | Yes | 0.00011 |
| CH 012531 | 1 | NPM1 | NM_002520.6:c.860_863dup | NP_002511.1:p.Trp288CysfsTer12 | 0.00012 | 3 | 0 | 2 | 25523 | 6894 | frameshift | 5 | 170837543 | C | CTCTG | Yes | 0.00014 |
| CH 012551 | 1 | IDH2 | NM_002168.3:c.419G>A | NP_002159.2:p.Arg140Gln | 0.08580 | 1681 | 1017 | 313 | 18724 | 11853 | missense | 15 | 90631934 | C | T | Yes | 0.09260 |
| CH 012551 | 1 | NPM1 | NM_002520.6:c.860_863dup | NP_002511.1:p.Trp288CysfsTer12 | 0.00049 | 14 | 0 | 9 | 28293 | 5606 | frameshift | 5 | 170837543 | C | CTCTG | Yes | 0.00083 |
| CH 012542 | 1 | FLT3 | NM_004119.2:c.1813_1814insCCCCTTTCAGAGAATATGAATATGATCTCAAATGGGAGT | NP_004110.2:p.Glu604_Phe605insSerProPheArgGluTyrGluTyrAspLeuLysTrpGlu | 0.00160 | 90 | NA | 66 | 56344 | NA | FLT3-ITD | 13 | 28608242 | A | AACTCCCATTTGAGATCATATTCATATTCTCTGAAAGGGG | Not Done | Not Done |

| Subject ID | Cohort | Gene | HGVSc | HGVSp | VAF | Alternate Observations | Deep Alternate Observations | Unique Alternate Observations | Depth | Deep Depth | Variant Consequence | Chromosome | Position | Reference | Alternate | ddPCR Validated | ddPCR VAF |
|---|---|---|---|---|---|---|---|---|---|---|---|---|---|---|---|---|---|
| CH 012182 | 1 | NPM1 | NM_002520.6:c.860_863dup | NP_002511.1:p.Trp288CysfsTer12 | 0.00080 | 18 | 2 | 14 | 22605 | 6305 | frameshift | 5 | 170837543 | C | CTCTG | Yes | 0.00079 |
| CH 012221 | 1 | NPM1 | NM_002520.6:c.860_863dup | NP_002511.1:p.Trp288CysfsTer12 | 0.00025 | 5 | 1 | 2 | 19741 | 6487 | frameshift | 5 | 170837543 | C | CTCTG | Yes | 0.00012 |
| CH 012235 | 1 | IDH1 | NM_005896.3:c.395G>A | NP_005887.2:p.Arg132His | 0.00250 | 118 | 36 | 73 | 33189 | 14427 | missense | 2 | 209113112 | C | T | Yes | 0.00362 |
| CH 012235 | 1 | NPM1 | NM_002520.6:c.860_863dup | NP_002511.1:p.Trp288CysfsTer12 | 0.00323 | 92 | 7 | 54 | 28496 | 7436 | frameshift | 5 | 170837543 | C | CTCTG | Yes | 0.00275 |
| CH 012235 | 1 | FLT3 | NM_004119.2:c.1779_1837+4dup | p.? | 0.00096 | 97 | NA | 48 | 101165 | NA | FLT3-ITD | 13 | 28608215 | T | TTACCAAACTCTAAATTTTCTCTTGGAAACTCCCATTTGAGATCATATTCATATTCTCTGAAAT | Not Done | Not Done |
| CH 012241 | 1 | NPM1 | NM_002520.6:c.860_863dup | NP_002511.1:p.Trp288CysfsTer12 | 0.00011 | 3 | 1 | 2 | 27196 | 8911 | frameshift | 5 | 170837543 | C | CTCTG | Not Done | Not Done |
| CH 012252 | 1 | IDH2 | NM_002168.3:c.419G>A | NP_002159.2:p.Arg140Gln | 0.04801 | 913 | 592 | 245 | 19086 | 12332 | missense | 15 | 90631934 | C | T | Yes | 0.05022 |
| CH 012265 | 1 | IDH2 | NM_002168.3:c.419G>A | NP_002159.2:p.Arg140Gln | 0.00089 | 23 | 11 | 15 | 18296 | 12348 | missense | 15 | 90631934 | C | T | Yes | 0.00140 |
| CH 012242 | 1 | FLT3 | NM_004119.2:c.1784_1837+12dup | p.? | 0.01940 | 2450 | NA | 319 | 126275 | NA | FLT3-ITD | 13 | 28608207 | T | TTCCATTCTTACCAAACTCTAAATTTTCTCTTGGAAACTCCCATTTGAGATCATATTCATATTCTCT | Not Done | Not Done |
| CH 012242 | 1 | FLT3 | NM_004119.2:c.1837+20_1837+21insGAATATGAATATGATCTCAAATGGGAGTTTCCAAGAGAAAATTTAGAGTTTGGTAAGAATGGAATGTGCCAAA | p.? | 0.00065 | 90 | NA | 45 | 139234 | NA | FLT3-ITD | 13 | 28608199 | T | TTTGGCACATTCCATTCTTACCAAACTCTAAATTTTCTCTTGGAAACTCCCATTTGAGATCATATTCATATTCT | Not Done | Not Done |
| CH 012242 | 1 | FLT3 | NM_004119.2:c.1778_1779insCATGGTACAGGTGACCGGCTCCTCAGATAATGAGTACTTCTACGTTGA | NP_004110.2:p.Met578_Asp593dup | 0.00021 | 17 | NA | 15 | 79304 | NA | FLT3-ITD | 13 | 28608277 | A | ATCAACGTAGAAGTACTCATTATCTGAGGAGCCGGTCACCTGTACCATG | Not Done | Not Done |
| CH 012258 | 1 | NPM1 | NM_002520.6:c.860_863dup | NP_002511.1:p.Trp288CysfsTer12 | 0.00012 | 4 | 1 | 3 | 33661 | 6922 | frameshift | 5 | 170837543 | C | CTCTG | Yes | 0.00003 |
| CH 012273 | 1 | FLT3 | NM_004119.2:c.1770_1793dup | NP_004110.2:p.Tyr597_Glu598insAspTyrValAspPheArgGluTyr | 0.01151 | 700 | NA | 44 | 60829 | NA | FLT3-ITD | 13 | 28608262 | T | TTCATATTCTCTGAAATCAACGTAG | Not Done | Not Done |
| CH 012095 | 1 | IDH2 | NM_002168.3:c.514A>T | NP_002159.2:p.Arg172Trp | 0.00516 | 107 | 59 | 59 | 20838 | 11426 | missense | 15 | 90631839 | T | A | Yes | 0.00501 |
| CH 012141 | 1 | IDH2 | NM_002168.3:c.515G>A | NP_002159.2:p.Arg172Lys | 0.00382 | 61 | 32 | 36 | 13901 | 8377 | missense | 15 | 90631838 | C | T | Yes | 0.00474 |
| CH 012131 | 1 | NPM1 | NM_002520.6:c.860_863dup | NP_002511.1:p.Trp288CysfsTer12 | 0.00017 | 6 | 0 | 4 | 34325 | 5707 | frameshift | 5 | 170837543 | C | CTCTG | Yes | 0.00015 |
| CH 012161 | 1 | IDH2 | NM_002168.3:c.419G>A | NP_002159.2:p.Arg140Gln | 0.26084 | 6195 | 3187 | 480 | 23059 | 12218 | missense | 15 | 90631934 | C | T | Yes | 0.27842 |
| CH 012296 | 1 | IDH1 | NM_005896.3:c.395G>A | NP_005887.2:p.Arg132His | 0.00434 | 179 | 74 | 86 | 39422 | 17049 | missense | 2 | 209113112 | C | T | Yes | 0.00413 |
| CH 012296 | 1 | IDH2 | NM_002168.3:c.419G>A | NP_002159.2:p.Arg140Gln | 0.00165 | 39 | 22 | 30 | 21626 | 13336 | missense | 15 | 90631934 | C | T | Yes | 0.00170 |
| CH 012177 | 1 | IDH1 | NM_005896.3:c.394C>T | NP_005887.2:p.Arg132Cys | 0.06265 | 2896 | 1236 | 429 | 40875 | 19728 | missense | 2 | 209113113 | G | A | Yes | 0.06833 |
| CH 012280 | 1 | IDH1 | NM_005896.3:c.395G>A | NP_005887.2:p.Arg132His | 0.13771 | 5041 | 2457 | 513 | 34920 | 17842 | missense | 2 | 209113112 | C | T | Yes | 0.14898 |
| CH 012310 | 1 | NPM1 | NM_002520.6:c.860_863dup | NP_002511.1:p.Trp288CysfsTer12 | 0.00596 | 193 | 15 | 83 | 32394 | 7788 | frameshift | 5 | 170837543 | C | CTCTG | Yes | 0.00520 |
| CH 012310 | 1 | FLT3 | NM_004119.2:c.1837_1837+1insAAGAATGCGGCTCCTCAGATAATGAGTACTTCTACGTTGATTTCAGAGAATATGAATATGATCTCAAATGGGAGTTTCCAAGAGAAAATTTAGAGTTTG | NP_004110.2:p.Phe612_Gly613insGluGluCysGlySerSerAspAsnGluTyrPheTyrValAspPheArgGluTyrGluTyrAspLeuLysTrpGluPheProArgGluAsnLeuGluPh | 0.00373 | 396 | NA | 95 | 106289 | NA | FLT3-ITD | 13 | 28608218 | C | CCAAACTCTAAATTTTCTCTTGGAAACTCCCATTTGAGATCATATTCATATTCTCTGAAATCAACGTAGAAGTACTCATTATCTGAGGAGCCGCATTCTT | Not Done | Not Done |
| CH 012313 | 1 | NPM1 | NM_002520.6:c.860_863dup | NP_002511.1:p.Trp288CysfsTer12 | 0.00048 | 17 | 0 | 12 | 35344 | 6969 | frameshift | 5 | 170837543 | C | CTCTG | Yes | 0.00053 |
| CH 012313 | 1 | FLT3 | NM_004119.2:c.1742_1822dup | NP_004110.2:p.Val581_Arg607dup | 0.00023 | 23 | NA | 13 | 99496 | NA | FLT3-ITD | 13 | 28608234 | C | CTCTTGGAAACTCCCATTTGAGATCATATTCATATTCTCTGAAATCAACGTAGAAGTACTCATTATCTGAGGAGCCGGTCAC | Not Done | Not Done |
| CH 012313 | 1 | IDH1 | NM_005896.3:c.394C>T | NP_005887.2:p.Arg132Cys | 0.00698 | 403 | 118 | 141 | 43823 | 16898 | missense | 2 | 209113113 | G | A | Yes | 0.00988 |
| CH 012347 | 1 | IDH2 | NM_002168.3:c.419G>A | NP_002159.2:p.Arg140Gln | 0.01068 | 345 | 176 | 117 | 27465 | 16480 | missense | 15 | 90631934 | C | T | Yes | 0.01164 |
| CH 012328 | 1 | IDH2 | NM_002168.3:c.419G>A | NP_002159.2:p.Arg140Gln | 0.03028 | 450 | 286 | 141 | 14624 | 9446 | missense | 15 | 90631934 | C | T | Yes | 0.02849 |
| CH 012362 | 1 | IDH2 | NM_002168.3:c.419G>A | NP_002159.2:p.Arg140Gln | 0.00763 | 154 | 98 | 75 | 19977 | 12842 | missense | 15 | 90631934 | C | T | Yes | 0.00900 |
| CH 012353 | 1 | IDH2 | NM_002168.3:c.419G>A | NP_002159.2:p.Arg140Gln | 0.00209 | 41 | 24 | 26 | 17842 | 11491 | missense | 15 | 90631934 | C | T | Yes | 0.00237 |
| CH 012365 | 1 | NPM1 | NM_002520.6:c.860_863dup | NP_002511.1:p.Trp288CysfsTer12 | 0.00120 | 46 | 7 | 23 | 38416 | 12620 | frameshift | 5 | 170837543 | C | CTCTG | Yes | 0.00114 |
| CH 012365 | 1 | FLT3 | NM_004119.2:c.1745_1825dup | NP_004110.2:p.Thr582_Glu608dup | 0.00046 | 45 | NA | 30 | 98157 | NA | FLT3-ITD | 13 | 28608231 | T | TTTCTCTTGGAAACTCCCATTTGAGATCATATTCATATTCTCTGAAATCAACGTAGAAGTACTCATTATCTGAGGAGCCGGT | Not Done | Not Done |
| CH 012361 | 1 | IDH2 | NM_002168.3:c.419G>A | NP_002159.2:p.Arg140Gln | 0.04372 | 684 | 434 | 188 | 15120 | 9927 | missense | 15 | 90631934 | C | T | Yes | 0.05181 |
| CH 012361 | 1 | NPM1 | NM_002520.6:c.860_863dup | NP_002511.1:p.Trp288CysfsTer12 | 0.00069 | 15 | 4 | 9 | 21732 | 6226 | frameshift | 5 | 170837543 | C | CTCTG | Yes | 0.00073 |
| CH 012363 | 1 | IDH1 | NM_005896.3:c.394C>T | NP_005887.2:p.Arg132Cys | 0.02007 | 1207 | 333 | 291 | 49346 | 16594 | missense | 2 | 209113113 | G | A | Yes | 0.02385 |
| CH 012382 | 1 | IDH1 | NM_005896.3:c.395G>T | NP_005887.2:p.Arg132Leu | 0.01580 | 673 | 230 | 205 | 40160 | 14561 | missense | 2 | 209113112 | C | A | Not Done | Not Done |
| CH 012401 | 1 | NPM1 | NM_002520.6:c.860_863dup | NP_002511.1:p.Trp288CysfsTer12 | 0.00173 | 54 | 2 | 33 | 31208 | 4425 | frameshift | 5 | 170837543 | C | CTCTG | Not Done | Not Done |
| CH 012401 | 1 | FLT3 | NM_004119.2:c.1837+2_1837+3insGGGTCAGATAATGAGTACTTCTACGTTGATTTCAGAGAATATGAATATGATCTCAAATGGGAGTTTCCAAGAGAAAATTTAGAGTTTGGT | p.? | 0.00065 | 71 | NA | 39 | 109700 | NA | FLT3-ITD | 13 | 28608217 | A | ACCAAACTCTAAATTTTCTCTTGGAAACTCCCATTTGAGATCATATTCATATTCTCTGAAATCAACGTAGAAGTACTCATTATCTGACCCA | Not Done | Not Done |
| CH 012428 | 1 | FLT3 | NM_004119.2:c.1837+1_1837+2insAAAAGATAATGAGTACTTCTACGTTGATTTCAGAGAATATGAATATGATCTCAAATGGGAGTTTCCAAGAGAAAATTTAGAGTTTGG | p.? | 0.00042 | 55 | NA | 31 | 130349 | NA | FLT3-ITD | 13 | 28608217 | A | ACCAAACTCTAAATTTTCTCTTGGAAACTCCCATTTGAGATCATATTCATATTCTCTGAAATCAACGTAGAAGTACTCATTATCTTTT | Not Done | Not Done |
| CH 012450 | 1 | NPM1 | NM_002520.6:c.860_863dup | NP_002511.1:p.Trp288CysfsTer12 | 0.00231 | 64 | 0 | 35 | 27703 | 2612 | frameshift | 5 | 170837543 | C | CTCTG | Yes | 0.00348 |
| CH 012451 | 1 | NPM1 | NM_002520.6:c.863_864insCATG | NP_002511.1:p.Trp288CysfsTer12 | 0.00011 | 4 | 2 | 3 | 37799 | 6468 | frameshift | 5 | 170837545 | C | CTGCA | Yes | 0.00014 |
| CH 012437 | 1 | IDH2 | NM_002168.3:c.419G>A | NP_002159.2:p.Arg140Gln | 0.07207 | 1530 | 911 | 302 | 21009 | 12641 | missense | 15 | 90631934 | C | T | Yes | 0.07500 |
| CH 012465 | 1 | IDH2 | NM_002168.3:c.419G>A | NP_002159.2:p.Arg140Gln | 0.34301 | 7533 | 4243 | 531 | 21804 | 12370 | missense | 15 | 90631934 | C | T | Yes | 0.34390 |
| CH 012435 | 1 | KIT | NM_000222.2:c.2447A>T | NP_000213.1:p.Asp816Val | 0.00059 | 23 | 10 | 18 | 38460 | 17006 | missense | 4 | 55599321 | A | T | Yes | 0.00070 |
| CH 012481 | 1 | IDH2 | NM_002168.3:c.419G>A | NP_002159.2:p.Arg140Gln | 0.01861 | 515 | 255 | 180 | 23955 | 13704 | missense | 15 | 90631934 | C | T | Yes | 0.02365 |
| CH 012479 | 1 | IDH2 | NM_002168.3:c.419G>A | NP_002159.2:p.Arg140Gln | 0.00173 | 30 | 18 | 16 | 18157 | 10383 | missense | 15 | 90631934 | C | T | Yes | 0.00218 |
| CH 012228 | 1 | IDH1 | NM_005896.3:c.394C>T | NP_005887.2:p.Arg132Cys | 0.00424 | 187 | 74 | 83 | 34772 | 17459 | missense | 2 | 209113113 | G | A | Yes | 0.00407 |
| CH 012203 | 1 | IDH2 | NM_002168.3:c.515G>A | NP_002159.2:p.Arg172Lys | 0.03023 | 805 | 491 | 240 | 24845 | 16244 | missense | 15 | 90631838 | C | T | Yes | 0.03595 |
| CH 012172 | 1 | IDH1 | NM_005896.3:c.394C>T | NP_005887.2:p.Arg132Cys | 0.00377 | 224 | 71 | 100 | 44961 | 18816 | missense | 2 | 209113113 | G | A | Yes | 0.00453 |
| CH 012212 | 1 | IDH2 | NM_002168.3:c.419G>A | NP_002159.2:p.Arg140Gln | 0.00774 | 184 | 120 | 87 | 22597 | 15494 | missense | 15 | 90631934 | C | T | Yes | 0.00920 |
| CH 012190 | 1 | FLT3 | NM_004119.2:c.1790_1791insCGGGACCGGCTCCTCAGATAATGAGTACTTCTACGTTGATTTCAGAGAATA | NP_004110.2:p.Tyr597_Glu598insGlyThrGlySerSerAspAsnGluTyrPheTyrValAspPheArgGluTyr | 0.00027 | 25 | NA | 15 | 93591 | NA | FLT3-ITD | 13 | 28608265 | A | ATATTCTCTGAAATCAACGTAGAAGTACTCATTATCTGAGGAGCCGGTCCCG | Not Done | Not Done |
| CH 014422 | 1 | FLT3 | NM_004119.2:c.1837+20_1837+21insGCCTTCAGAGAATATGAATATGATCTCAAATGGGAGTTTCCAAGAGAAAATTTAGAGTTTGGTAAGAATGGAATGTGCCA | p.? | 0.00018 | 20 | NA | 13 | 114277 | NA | FLT3-ITD | 13 | 28608198 | T | TTGGCACATTCCATTCTTACCAAACTCTAAATTTTCTCTTGGAAACTCCCATTTGAGATCATATTCATATTCTCTGAAGGC | Not Done | Not Done |
| CH 014416 | 1 | FLT3 | NM_004119.2:c.1781_1782insTGGCTCCTCAGATAATGAGTACTTCTACGTTGATTT | p.? | 0.00043 | 28 | NA | 16 | 64586 | NA | FLT3-ITD | 13 | 28608274 | G | GAAATCAACGTAGAAGTACTCATTATCTGAGGAGCCA | Not Done | Not Done |
| CH 012068 | 1 | IDH2 | NM_002168.3:c.419G>A | NP_002159.2:p.Arg140Gln | 0.00109 | 30 | 14 | 19 | 19553 | 12894 | missense | 15 | 90631934 | C | T | Yes | 0.00199 |
| CH 012068 | 1 | NPM1 | NM_002520.6:c.860_863dup | NP_002511.1:p.Trp288CysfsTer12 | 0.00100 | 29 | 4 | 16 | 29055 | 7072 | frameshift | 5 | 170837543 | C | CTCTG | Yes | 0.00104 |
| CH 016959 | 2 | FLT3 | NM_004119.2:c.1770_1793dup | NP_004110.2:p.Tyr597_Glu598insAspTyrValAspPheArgGluTyr | 0.00119 | 93 | NA | 44 | 78220 | NA | FLT3-ITD | 13 | 28608262 | T | TTCATATTCTCTGAAATCAACGTAG | Not Done | Not Done |
| CH 016959 | 2 | IDH2 | NM_002168.3:c.419G>A | NP_002159.2:p.Arg140Gln | 0.00253 | 75 | 37 | 41 | 22701 | 14617 | missense | 15 | 90631934 | C | T | Yes | 0.00242 |
| CH 016959 | 2 | NPM1 | NM_002520.6:c.860_863dup | NP_002511.1:p.Trp288CysfsTer12 | 0.00061 | 21 | 7 | 14 | 34554 | 11783 | frameshift | 5 | 170837543 | C | CTCTG | Not Done | Not Done |
| CH 016941 | 2 | FLT3 | NM_004119.2:c.1775_1795dup | NP_004110.2:p.Glu598_Tyr599insPheAspPheArgGluTyrGlu | 0.00023 | 17 | NA | 16 | 74735 | NA | FLT3-ITD | 13 | 28608260 | T | TATTCATATTCTCTGAAATCAA | Not Done | Not Done |
| CH 016940 | 2 | IDH2 | NM_002168.3:c.419G>A | NP_002159.2:p.Arg140Gln | 0.02787 | 640 | 385 | 211 | 22594 | 13812 | missense | 15 | 90631934 | C | T | Yes | 0.02673 |
| CH 016868 | 2 | IDH2 | NM_002168.3:c.419G>A | NP_002159.2:p.Arg140Gln | 0.00761 | 241 | 135 | 103 | 26455 | 17745 | missense | 15 | 90631934 | C | T | Yes | 0.01006 |
| CH 016873 | 2 | IDH2 | NM_002168.3:c.419G>A | NP_002159.2:p.Arg140Gln | 0.18720 | 4962 | 2373 | 443 | 26102 | 12676 | missense | 15 | 90631934 | C | T | Yes | 0.20223 |

| Subject ID | Cohort | Gene | HGVSc | HGVSp | VAF | Alternate Observations | Deep Alternate Observations | Unique Alternate Observations | Depth | Deep Depth | Variant Consequence | Chromosome | Position | Reference | Alternate | ddPCR Validated | ddPCR VAF |
|---|---|---|---|---|---|---|---|---|---|---|---|---|---|---|---|---|---|
| CH 016976 | 2 | NPM1 | NM_002520.6:c.860_863dup | NP_002511.1:p.Trp288CysfsTer12 | 0.00392 | 167 | 12 | 87 | 42589 | 8304 | frameshift | 5 | 170837543 | C | CTCTG | Yes | 0.00443 |
| CH 016952 | 2 | IDH2 | NM_002168.3:c.419G>A | NP_002159.2:p.Arg140Gln | 0.00070 | 31 | 14 | 22 | 29891 | 20082 | missense | 15 | 90631934 | C | T | Yes | 0.00070 |
| CH 016983 | 2 | IDH2 | NM_002168.3:c.419G>A | NP_002159.2:p.Arg140Gln | 0.00447 | 111 | 63 | 67 | 22503 | 14083 | missense | 15 | 90631934 | C | T | Yes | 0.00531 |
| CH 016763 | 2 | NPM1 | NM_002520.6:c.860_863dup | NP_002511.1:p.Trp288CysfsTer12 | 0.00010 | 4 | 1 | 3 | 38109 | 8035 | frameshift | 5 | 170837543 | C | CTCTG | Not Done | Not Done |
| CH 016833 | 2 | IDH2 | NM_002168.3:c.419G>A | NP_002159.2:p.Arg140Gln | 0.23850 | 5958 | 3779 | 537 | 24744 | 15845 | missense | 15 | 90631934 | C | T | Yes | 0.24327 |
| CH 016744 | 2 | IDH2 | NM_002168.3:c.419G>A | NP_002159.2:p.Arg140Gln | 0.01061 | 172 | 115 | 80 | 15732 | 10836 | missense | 15 | 90631934 | C | T | Yes | 0.01303 |
| CH 016883 | 2 | FLT3 | NM_004119.2:c.1812_1813insCAGGTACAGGTGACCGGCTCCTCAGATAATGAGTACTTCTACGTTGATTTCAGAGAATATGAATATGATCTCAAATGGGAG | NP_004110.2:p.Glu604_Phe605insGlnValGlnValThrGlySerSerAspAsnGluTyrPheTyrValAspPheArgGluTyrGluTyrAspLeuLysTrpGlu | 0.00224 | 150 | NA | 66 | 67092 | NA | FLT3-ITD | 13 | 28608243 | A | ACTCCCATTTGAGATCATATTCATATTCTCTGAAATCAACGTAGAAGTACTCATTATCTGAGGAGCCGGTCACCTGTACCTG | Not Done | Not Done |
| CH 016555 | 2 | FLT3 | NM_004119.2:c.1744_1837+14dup | p.? | 0.00954 | 1566 | NA | 224 | 164072 | NA | FLT3-ITD | 13 | 28608204 | A | ACATTCCATTCTTACCAAACTCTAAATTTTCTCTTGGAAACTCCCATTTGAGATCATATTCATATTCTCTGAAATCAACGTAGAAGTACTCATTATCTGAGGAGCCGGT | Not Done | Not Done |
| CH 016557 | 2 | IDH2 | NM_002168.3:c.515G>A | NP_002159.2:p.Arg172Lys | 0.03690 | 880 | 543 | 255 | 24362 | 14717 | missense | 15 | 90631838 | C | T | Yes | 0.04013 |
| CH 016536 | 2 | IDH2 | NM_002168.3:c.419G>A | NP_002159.2:p.Arg140Gln | 0.00426 | 95 | 59 | 54 | 19950 | 13857 | missense | 15 | 90631934 | C | T | Yes | 0.00416 |
| CH 016498 | 2 | IDH1 | NM_005896.3:c.394C>A | NP_005887.2:p.Arg132Ser | 0.00107 | 65 | 23 | 42 | 44701 | 21453 | missense | 2 | 209113113 | G | T | Yes | 0.00090 |
| CH 016988 | 2 | IDH2 | NM_002168.3:c.419G>A | NP_002159.2:p.Arg140Gln | 0.05984 | 1219 | 757 | 283 | 19461 | 12651 | missense | 15 | 90631934 | C | T | Yes | 0.06244 |
| CH 016515 | 2 | IDH2 | NM_002168.3:c.419G>A | NP_002159.2:p.Arg140Gln | 0.01069 | 215 | 124 | 82 | 20010 | 11602 | missense | 15 | 90631934 | C | T | Yes | 0.01397 |
| CH 016984 | 2 | IDH2 | NM_002168.3:c.419G>A | NP_002159.2:p.Arg140Gln | 0.27746 | 4632 | 3195 | 478 | 16518 | 11515 | missense | 15 | 90631934 | C | T | Yes | 0.27775 |
| CH 016522 | 2 | NPM1 | NM_002520.6:c.860_863dup | NP_002511.1:p.Trp288CysfsTer12 | 0.00010 | 3 | 0 | 3 | 29601 | 8396 | frameshift | 5 | 170837543 | C | CTCTG | Yes | 0.00009 |
| CH 016629 | 2 | IDH2 | NM_002168.3:c.419G>A | NP_002159.2:p.Arg140Gln | 0.00138 | 37 | 15 | 27 | 15900 | 10869 | missense | 15 | 90631934 | C | T | Yes | 0.00240 |
| CH 016518 | 2 | IDH2 | NM_002168.3:c.419G>A | NP_002159.2:p.Arg140Gln | 0.03027 | 436 | 285 | 140 | 13453 | 9414 | missense | 15 | 90631934 | C | T | Yes | 0.02724 |
| CH 016553 | 2 | IDH2 | NM_002168.3:c.419G>A | NP_002159.2:p.Arg140Gln | 0.00593 | 135 | 82 | 77 | 20332 | 13836 | missense | 15 | 90631934 | C | T | Yes | 0.00682 |
| CH 016509 | 2 | FLT3 | NM_004119.2:c.1780_1800dup | NP_004110.2:p.Phe594_Asp600dup | 0.00028 | 23 | NA | 18 | 82334 | NA | FLT3-ITD | 13 | 28608255 | G | GATCATATTCATATTCTCTGAA | Not Done | Not Done |
| CH 016509 | 2 | NPM1 | NM_002520.6:c.860_863dup | NP_002511.1:p.Trp288CysfsTer12 | 0.00116 | 42 | 2 | 21 | 36089 | 9716 | frameshift | 5 | 170837543 | C | CTCTG | Yes | 0.00134 |
| CH 016567 | 2 | IDH2 | NM_002168.3:c.419G>A | NP_002159.2:p.Arg140Gln | 0.19123 | 3809 | 2452 | 463 | 19628 | 12822 | missense | 15 | 90631934 | C | T | Yes | 0.20364 |
| CH 016521 | 2 | FLT3 | NM_004119.2:c.1787_1788insGTCCCATGGGCTTCATGATTTCAGAGA | NP_004110.2:p.Glu596_Tyr597insSerHisGlyLeuHisAspPheArgGlu | 0.01262 | 914 | NA | 232 | 72407 | NA | FLT3-ITD | 13 | 28608268 | T | TTCTCTGAAATCATGAAGCCCATGGGAC | Not Done | Not Done |
| CH 016521 | 2 | FLT3 | NM_004119.2:c.1833_1834insCCGAAATGGGAGTTTCCAAGAGAAAATTTAGAG | NP_004110.2:p.Glu611_Phe612insProLysTrpGluPheProArgGluAsnLeuGlu | 0.00271 | 269 | NA | 123 | 99195 | NA | FLT3-ITD | 13 | 28608222 | A | ACTCTAAATTTTCTCTTGGAAACTCCCATTTCGG | Not Done | Not Done |
| CH 016521 | 2 | NPM1 | NM_002520.6:c.860_863dup | NP_002511.1:p.Trp288CysfsTer12 | 0.01457 | 505 | 40 | 156 | 34669 | 7028 | frameshift | 5 | 170837543 | C | CTCTG | Yes | 0.01567 |
| CH 016660 | 2 | IDH2 | NM_002168.3:c.419G>A | NP_002159.2:p.Arg140Gln | 0.08519 | 1153 | 779 | 269 | 13413 | 9144 | missense | 15 | 90631934 | C | T | Yes | 0.08690 |
| CH 016624 | 2 | IDH2 | NM_002168.3:c.419G>A | NP_002159.2:p.Arg140Gln | 0.05055 | 620 | 440 | 187 | 11965 | 8705 | missense | 15 | 90631934 | C | T | Yes | 0.05416 |
| CH 016631 | 2 | IDH2 | NM_002168.3:c.419G>A | NP_002159.2:p.Arg140Gln | 0.00154 | 31 | 15 | 22 | 13537 | 9764 | missense | 15 | 90631934 | C | T | Yes | 0.00248 |
| CH 016695 | 2 | FLT3 | NM_004119.2:c.1778_1837dup | NP_004110.2:p.Asp593_Phe612dup | 0.00134 | 131 | NA | 92 | 97535 | NA | FLT3-ITD | 13 | 28608218 | C | CCAAACTCTAAATTTTCTCTTGGAAACTCCCATTTGAGATCATATTCATATTCTCTGAAAT | Not Done | Not Done |
| CH 016663 | 2 | IDH1 | NM_005896.3:c.394C>A | NP_005887.2:p.Arg132Ser | 0.01596 | 356 | 204 | 131 | 21023 | 12783 | missense | 2 | 209113113 | G | T | Yes | 0.01690 |
| CH 016870 | 2 | IDH1 | NM_005896.3:c.394C>G | NP_005887.2:p.Arg132Gly | 0.01058 | 298 | 166 | 133 | 26660 | 15691 | missense | 2 | 209113113 | G | C | Yes | 0.01208 |
| CH 016655 | 2 | FLT3 | NM_004119.2:c.1784_1804dup | NP_004110.2:p.Arg595_Leu601dup | 0.00055 | 32 | NA | 14 | 58566 | NA | FLT3-ITD | 13 | 28608251 | T | TTGAGATCATATTCATATTCTC | Not Done | Not Done |
| CH 016655 | 2 | NPM1 | NM_002520.6:c.863_864insCATG | NP_002511.1:p.Trp288CysfsTer12 | 0.00369 | 101 | 39 | 69 | 27360 | 9887 | frameshift | 5 | 170837545 | C | CTGCA | Yes | 0.00437 |
| CH 016753 | 2 | FLT3 | NM_004119.2:c.1788_1789insGGGGGCCCTTTCAGAGAA | NP_004110.2:p.Glu596_Tyr597insGlyGlyProPheArgGlu | 0.00137 | 36 | 22 | 26 | 26237 | 12547 | FLT3-ITD | 13 | 28608267 | A | ATTCTCTGAAAGGGCCCCC | Not Done | Not Done |
| CH 016684 | 2 | NPM1 | NM_002520.6:c.863_864insCCTG | NP_002511.1:p.Trp288CysfsTer12 | 0.00032 | 10 | 5 | 8 | 31398 | 11395 | frameshift | 5 | 170837544 | T | TCTGC | Yes | 0.00053 |
| CH 016686 | 2 | IDH2 | NM_002168.3:c.419G>A | NP_002159.2:p.Arg140Gln | 0.00559 | 90 | 52 | 49 | 13569 | 9295 | missense | 15 | 90631934 | C | T | Yes | 0.00602 |
| CH 016712 | 2 | IDH2 | NM_002168.3:c.419G>A | NP_002159.2:p.Arg140Gln | 0.22492 | 3081 | 2049 | 374 | 13518 | 9110 | missense | 15 | 90631934 | C | T | Yes | 0.22841 |
| CH 016643 | 2 | FLT3 | NM_004119.2:c.2503G>C | NP_004110.2:p.Asp835His | 0.19613 | 6687 | 3788 | 729 | 33706 | 19314 | missense | 13 | 28592642 | C | G | Yes | 0.20137 |
| CH 016683 | 2 | IDH1 | NM_005896.3:c.394C>A | NP_005887.2:p.Arg132Ser | 0.01033 | 239 | 147 | 118 | 23713 | 14224 | missense | 2 | 209113113 | G | T | Yes | 0.00980 |
| CH 016688 | 2 | NPM1 | NM_002520.6:c.860_863dup | NP_002511.1:p.Trp288CysfsTer12 | 0.00026 | 8 | 1 | 4 | 30320 | 11752 | frameshift | 5 | 170837543 | C | CTCTG | Yes | 0.00028 |
| CH 016673 | 2 | IDH1 | NM_005896.3:c.394C>G | NP_005887.2:p.Arg132Gly | 0.01945 | 600 | 345 | 221 | 30821 | 17734 | missense | 2 | 209113113 | G | C | Yes | 0.02029 |
| CH 016693 | 2 | FLT3 | NM_004119.2:c.1742_1786dup | NP_004110.2:p.Val581_Arg595dup | 0.00015 | 7 | NA | 6 | 45659 | NA | FLT3-ITD | 13 | 28608269 | T | TCTCTGAAATCAACGTAGAAGTACTCATTATCTGAGGAGCCGGTCA | Not Done | Not Done |
| CH 016691 | 2 | NPM1 | NM_002520.6:c.860_863dup | NP_002511.1:p.Trp288CysfsTer12 | 0.00016 | 4 | 1 | 2 | 24690 | 9693 | frameshift | 5 | 170837543 | C | CTCTG | Yes | 0.00021 |
| CH 016649 | 2 | IDH1 | NM_005896.3:c.394C>T | NP_005887.2:p.Arg132Cys | 0.02234 | 619 | 342 | 220 | 25060 | 15306 | missense | 2 | 209113113 | G | A | Yes | 0.02783 |
| CH 016764 | 2 | IDH1 | NM_005896.3:c.395G>A | NP_005887.2:p.Arg132His | 0.00215 | 83 | 37 | 52 | 28437 | 17182 | missense | 2 | 209113112 | C | T | Yes | 0.00320 |
| CH 016764 | 2 | IDH2 | NM_002168.3:c.419G>A | NP_002159.2:p.Arg140Gln | 0.40324 | 6319 | 4380 | 463 | 15760 | 10862 | missense | 15 | 90631934 | C | T | Yes | 0.39821 |
| CH 016702 | 2 | IDH2 | NM_002168.3:c.419G>A | NP_002159.2:p.Arg140Gln | 0.01002 | 162 | 104 | 81 | 15235 | 10380 | missense | 15 | 90631934 | C | T | Yes | 0.01291 |
| CH 016519 | 2 | FLT3 | NM_004119.2:c.1760_1819dup | NP_004110.2:p.Asn587_Pro606dup | 0.00016 | 15 | NA | 8 | 91408 | NA | FLT3-ITD | 13 | 28608236 | C | CTTGGAAACTCCCATTTGAGATCATATTCATATTCTCTGAAATCAACGTAGAAGTACTCAT | Not Done | Not Done |
| CH 016519 | 2 | NPM1 | NM_002520.6:c.860_863dup | NP_002511.1:p.Trp288CysfsTer12 | 0.00049 | 18 | 5 | 11 | 36560 | 13265 | frameshift | 5 | 170837543 | C | CTCTG | Yes | 0.00042 |
| CH 016501 | 2 | FLT3 | NM_004119.2:c.1789_1815dup | NP_004110.2:p.Tyr597_Phe605dup | 0.03411 | 2829 | NA | 413 | 82938 | NA | FLT3-ITD | 13 | 28608240 | G | GAAACTCCCATTTGAGATCATATTCATA | Not Done | Not Done |
| CH 016501 | 2 | FLT3 | NM_004119.2:c.1825_1826insGGGATGAGTACTTCTACGTTGATTTCAGAGAATATGAATATGATCTCAAATGGGAGTTTCCAAGAGAAA | NP_004110.2:p.Glu608_Asn609insArgAspGluTyrPheTyrValAspPheArgGluTyrGluTyrAspLeuLysTrpGluPheProArgGlu | 0.00187 | 135 | NA | 65 | 72355 | NA | FLT3-ITD | 13 | 28608230 | T | TTTTCTCTTGGAAACTCCCATTTGAGATCATATTCATATTCTCTGAAATCAACGTAGAAGTACTCATCCC | Not Done | Not Done |
| CH 016546 | 2 | FLT3 | NM_004119.2:c.1784_1804dup | NP_004110.2:p.Arg595_Leu601dup | 0.00035 | 30 | NA | 17 | 84776 | NA | FLT3-ITD | 13 | 28608251 | T | TTGAGATCATATTCATATTCTC | Not Done | Not Done |
| CH 016546 | 2 | NPM1 | NM_002520.6:c.860_863dup | NP_002511.1:p.Trp288CysfsTer12 | 0.00092 | 31 | 4 | 20 | 33577 | 10783 | frameshift | 5 | 170837543 | C | CTCTG | Yes | 0.00087 |
| CH 016550 | 2 | IDH2 | NM_002168.3:c.419G>A | NP_002159.2:p.Arg140Gln | 0.07247 | 1579 | 1003 | 336 | 20198 | 13841 | missense | 15 | 90631934 | C | T | Yes | 0.08521 |
| CH 016535 | 2 | IDH2 | NM_002168.3:c.419G>A | NP_002159.2:p.Arg140Gln | 0.33845 | 7093 | 4806 | 488 | 20994 | 14200 | missense | 15 | 90631934 | C | T | Yes | 0.33460 |
| CH 016535 | 2 | NPM1 | NM_002520.6:c.860_863dup | NP_002511.1:p.Trp288CysfsTer12 | 0.00026 | 10 | 0 | 5 | 38488 | 14072 | frameshift | 5 | 170837543 | C | CTCTG | Yes | 0.00014 |
| CH 016541 | 2 | FLT3 | NM_004119.2:c.1802_1803insTGGCTCCTCAGATAATGAGTACTTCTACGTTGATTTCAGAGAATATGAATATGATCT | NP_004110.2:p.Gly583_Leu601dup | 0.06522 | 3795 | NA | 417 | 58192 | NA | FLT3-ITD | 13 | 28608253 | G | GAGATCATATTCATATTCTCTGAAATCAACGTAGAAGTACTCATTATCTGAGGAGCCA | Not Done | Not Done |
| CH 016541 | 2 | NPM1 | NM_002520.6:c.863_864insCATG | NP_002511.1:p.Trp288CysfsTer12 | 0.03212 | 877 | 242 | 254 | 27303 | 9058 | frameshift | 5 | 170837545 | C | CTGCA | Yes | 0.03273 |
| CH 016600 | 2 | NPM1 | NM_002520.6:c.860_863dup | NP_002511.1:p.Trp288CysfsTer12 | 0.02163 | 655 | 98 | 182 | 30279 | 10638 | frameshift | 5 | 170837543 | C | CTCTG | Yes | 0.02041 |
| CH 016548 | 2 | NPM1 | NM_002520.6:c.860_863dup | NP_002511.1:p.Trp288CysfsTer12 | 0.00013 | 3 | 0 | 1 | 22233 | 8099 | frameshift | 5 | 170837543 | C | CTCTG | Yes | 0.00023 |
| CH 016610 | 2 | FLT3 | NM_004119.2:c.1770_1793dup | NP_004110.2:p.Tyr597_Glu598insAspTyrValAspPheArgGluTyr | 0.59543 | 39168 | NA | 1033 | 65781 | NA | FLT3-ITD | 13 | 28608262 | T | TTCATATTCTCTGAAATCAACGTAG | Not Done | Not Done |
| CH 016593 | 2 | NPM1 | NM_002520.6:c.860_863dup | NP_002511.1:p.Trp288CysfsTer12 | 0.00067 | 15 | 7 | 9 | 22254 | 11080 | frameshift | 5 | 170837543 | C | CTCTG | Yes | 0.00061 |
| CH 016563 | 2 | FLT3 | NM_004119.2:c.1802_1803insGGGTATGGTACAGGTGACCGGCTCCTCAGATAATGAGTACTTCTACGTTGATTTCAGAGAATATGAATATGATCT | NP_004110.2:p.Leu601_Lys602insGlyMetValGlnValThrGlySerSerAspAsnGluTyrPheTyrValAspPheArgGluTyrGluTyrAspLeu | 0.00209 | 99 | NA | 47 | 47319 | NA | FLT3-ITD | 13 | 28608253 | G | GAGATCATATTCATATTCTCTGAAATCAACGTAGAAGTACTCATTATCTGAGGAGCCGGTCACCTGTACCATACCC | Not Done | Not Done |
| CH 016951 | 2 | IDH2 | NM_002168.3:c.419G>A | NP_002159.2:p.Arg140Gln | 0.20220 | 3479 | 2282 | 414 | 16914 | 11286 | missense | 15 | 90631934 | C | T | Yes | 0.20290 |
| CH 016671 | 2 | NPM1 | NM_002520.6:c.860_863dup | NP_002511.1:p.Trp288CysfsTer12 | 0.00026 | 8 | 2 | 8 | 30958 | 13911 | frameshift | 5 | 170837543 | C | CTCTG | Yes | 0.00023 |
| CH 016832 | 2 | IDH1 | NM_005896.3:c.395G>A | NP_005887.2:p.Arg132His | 0.00269 | 118 | 56 | 60 | 36697 | 20801 | missense | 2 | 209113112 | C | T | Yes | 0.00287 |
| CH 016832 | 2 | IDH2 | NM_002168.3:c.515G>A | NP_002159.2:p.Arg172Lys | 0.00626 | 159 | 93 | 77 | 25181 | 14854 | missense | 15 | 90631838 | C | T | Yes | 0.00772 |
| CH 016751 | 2 | IDH2 | NM_002168.3:c.515G>A | NP_002159.2:p.Arg172Lys | 0.01613 | 359 | 215 | 140 | 20370 | 13333 | missense | 15 | 90631838 | C | T | Yes | 0.01791 |
| CH 016701 | 2 | NPM1 | NM_002520.6:c.860_863dup | NP_002511.1:p.Trp288CysfsTer12 | 0.00033 | 11 | 3 | 7 | 33127 | 15422 | frameshift | 5 | 170837543 | C | CTCTG | Yes | 0.00037 |
| CH 016767 | 2 | IDH2 | NM_002168.3:c.419G>A | NP_002159.2:p.Arg140Gln | 0.13338 | 1980 | 1267 | 359 | 14903 | 9499 | missense | 15 | 90631934 | C | T | Yes | 0.13745 |
| CH 016724 | 2 | IDH2 | NM_002168.3:c.419G>A | NP_002159.2:p.Arg140Gln | 0.43385 | 7310 | 4178 | 431 | 16908 | 9630 | missense | 15 | 90631934 | C | T | Yes | 0.42621 |

| Subject ID | Cohort | Gene | HGVSc | HGVSp | VAF | Alternate Observations | Deep Alternate Observations | Unique Alternate Observations | Depth | Deep Depth | Variant Consequence | Chromosome | Position | Reference | Alternate | ddPCR Validated | ddPCR VAF |
|---|---|---|---|---|---|---|---|---|---|---|---|---|---|---|---|---|---|
| CH 016696 | 2 | FLT3 | NM_004119.2:c.1832_1833insAACCGGCTCCTCAGATAATGAGTACTTCTACGTTGATTTCAGAGAATATGAATATGATCTCAAATGGGAGTTTCCAAGAGAAAATTTAGA | NP_004110.2:p.Thr582_Glu611dup | 0.00042 | 52 | NA | 26 | 123713 | NA | FLT3-ITD | 13 | 28608223 | C | CTCTAAATTTTCTCTTGGAAACTCCCATTTGAGATCATATTCATATTCTCTGAAATCAACGTAGAAGTACTCATTATCTGAGGAGCCGGTT | Not Done | Not Done |
| CH 016696 | 2 | NPM1 | NM_002520.6:c.860_863dup | NP_002511.1:p.Trp288CysfsTer12 | 0.00155 | 65 | 8 | 39 | 42037 | 12385 | frameshift | 5 | 170837543 | C | CTCTG | Yes | 0.00163 |
| CH 016989 | 2 | IDH1 | NM_005896.3:c.394C>T | NP_005887.2:p.Arg132Cys | 0.02924 | 1011 | 575 | 294 | 32448 | 19666 | missense | 2 | 209113113 | G | A | Yes | 0.03584 |
| CH 016769 | 2 | IDH2 | NM_002168.3:c.419G>A | NP_002159.2:p.Arg140Gln | 0.03210 | 612 | 411 | 188 | 18357 | 12805 | missense | 15 | 90631934 | C | T | Yes | 0.02933 |
| CH 016818 | 2 | FLT3 | NM_004119.2:c.1789_1824dup | NP_004110.2:p.Tyr597_Glu608dup | 0.00017 | 12 | NA | 11 | 69182 | NA | FLT3-ITD | 13 | 28608231 | T | TTTCTCTTGGAAACTCCCATTTGAGATCATATTCATA | Not Done | Not Done |
| CH 016741 | 2 | FLT3 | NM_004119.2:c.1780_1800dup | NP_004110.2:p.Phe594_Asp600dup | 0.00179 | 144 | NA | 67 | 80581 | NA | FLT3-ITD | 13 | 28608255 | G | GATCATATTCATATTCTCTGAA | Not Done | Not Done |
| CH 016748 | 2 | IDH1 | NM_005896.3:c.394C>T | NP_005887.2:p.Arg132Cys | 0.03547 | 1232 | 729 | 313 | 32558 | 20552 | missense | 2 | 209113113 | G | A | Yes | 0.03998 |
| CH 016748 | 2 | NPM1 | NM_002520.6:c.860_863dup | NP_002511.1:p.Trp288CysfsTer12 | 0.00050 | 17 | 3 | 10 | 34071 | 15101 | frameshift | 5 | 170837543 | C | CTCTG | Yes | 0.00019 |
| CH 016755 | 2 | IDH2 | NM_002168.3:c.419G>A | NP_002159.2:p.Arg140Gln | 0.01921 | 244 | 167 | 105 | 13044 | 8692 | missense | 15 | 90631934 | C | T | Yes | 0.01828 |
| CH 016826 | 2 | NPM1 | NM_002520.6:c.863_864insCATG | NP_002511.1:p.Trp288CysfsTer12 | 0.00090 | 21 | 10 | 19 | 23414 | 10167 | frameshift | 5 | 170837545 | C | CTGCA | Yes | 0.00052 |
| CH 016715 | 2 | IDH2 | NM_002168.3:c.419G>A | NP_002159.2:p.Arg140Gln | 0.02342 | 371 | 226 | 118 | 15474 | 9648 | missense | 15 | 90631934 | C | T | Yes | 0.02335 |
| CH 016725 | 2 | IDH2 | NM_002168.3:c.419G>A | NP_002159.2:p.Arg140Gln | 0.00946 | 209 | 107 | 81 | 19637 | 11307 | missense | 15 | 90631934 | C | T | Yes | 0.00907 |
| CH 016725 | 2 | NPM1 | NM_002520.6:c.860_863dup | NP_002511.1:p.Trp288CysfsTer12 | 0.00165 | 51 | 13 | 25 | 30945 | 12373 | frameshift | 5 | 170837543 | C | CTCTG | Yes | 0.00226 |
| CH 016757 | 2 | IDH2 | NM_002168.3:c.419G>A | NP_002159.2:p.Arg140Gln | 0.30099 | 6448 | 3738 | 492 | 21166 | 12419 | missense | 15 | 90631934 | C | T | Yes | 0.29870 |
| CH 016664 | 2 | IDH2 | NM_002168.3:c.418C>T | NP_002159.2:p.Arg140Trp | 0.47795 | 7527 | 4682 | 505 | 15721 | 9796 | missense | 15 | 90631935 | G | A | Yes | 0.47839 |
| CH 016992 | 2 | FLT3 | NM_004119.2:c.1775_1795dup | NP_004110.2:p.Glu598_Tyr599insPheAspPheArgGluTyrGlu | 0.00011 | 9 | NA | 5 | 78848 | NA | FLT3-ITD | 13 | 28608260 | T | TATTCATATTCTCTGAAATCAA | Not Done | Not Done |
| CH 016678 | 2 | IDH2 | NM_002168.3:c.419G>A | NP_002159.2:p.Arg140Gln | 0.01793 | 346 | 203 | 122 | 17453 | 11324 | missense | 15 | 90631934 | C | T | Yes | 0.01426 |
| CH 016678 | 2 | NPM1 | NM_002520.6:c.863_864insCCTG | NP_002511.1:p.Trp288CysfsTer12 | 0.01369 | 410 | 82 | 159 | 29941 | 11736 | frameshift | 5 | 170837544 | T | TCTGC | Yes | 0.01639 |
| CH 016348 | 2 | FLT3 | NM_004119.2:c.1742_1786dup | NP_004110.2:p.Val581_Arg595dup | 0.00016 | 8 | NA | 5 | 51606 | NA | FLT3-ITD | 13 | 28608269 | T | TCTCTGAAATCAACGTAGAAGTACTCATTATCTGAGGAGCCGGTCA | Not Done | Not Done |
| CH 016486 | 2 | IDH1 | NM_005896.3:c.394C>T | NP_005887.2:p.Arg132Cys | 0.00209 | 46 | 28 | 35 | 20823 | 13423 | missense | 2 | 209113113 | G | A | Yes | 0.00329 |
| CH 016981 | 2 | FLT3 | NM_004119.2:c.1770_1793dup | NP_004110.2:p.Tyr597_Glu598insAspTyrValAspPheArgGluTyr | 0.05936 | 3006 | NA | 337 | 50644 | NA | FLT3-ITD | 13 | 28608262 | T | TTCATATTCTCTGAAATCAACGTAG | Not Done | Not Done |
| CH 016981 | 2 | NPM1 | NM_002520.6:c.867_868insAGGT | NP_002511.1:p.Trp290ArgfsTer10 | 0.00613 | 148 | 58 | 87 | 24145 | 10523 | frameshift | 5 | 170837551 | G | GAGGT | Not Done | Not Done |
| CH 016461 | 2 | IDH2 | NM_002168.3:c.419G>A | NP_002159.2:p.Arg140Gln | 0.01008 | 303 | 169 | 120 | 25232 | 16774 | missense | 15 | 90631934 | C | T | Yes | 0.01103 |
| CH 016374 | 2 | FLT3 | NM_004119.2:c.1824_1825insTATCCAACCGACTCCAAATGGGAGTTTCCAAGAGAA | NP_004110.2:p.Glu608_Asn609insTyrProThrAspSerLysTrpGluPheProArgGlu | 0.01446 | 692 | NA | 243 | 47856 | NA | FLT3-ITD | 13 | 28608231 | T | TTTCTCTTGGAAACTCCCATTTGGAGTCGGTTGGATA | Not Done | Not Done |
| CH 016374 | 2 | IDH2 | NM_002168.3:c.419G>A | NP_002159.2:p.Arg140Gln | 0.04454 | 672 | 423 | 180 | 14478 | 9497 | missense | 15 | 90631934 | C | T | Yes | 0.04830 |
| CH 016365 | 2 | FLT3 | NM_004119.2:c.1777_1837+2dup | p.? | 0.00404 | 443 | NA | 191 | 109728 | NA | FLT3-ITD | 13 | 28608216 | T | TACCAAACTCTAAATTTTCTCTTGGAAACTCCCATTTGAGATCATATTCATATTCTCTGAAATC | Not Done | Not Done |
| CH 016365 | 2 | IDH2 | NM_002168.3:c.419G>A | NP_002159.2:p.Arg140Gln | 0.05360 | 1156 | 692 | 254 | 21720 | 12910 | missense | 15 | 90631934 | C | T | Yes | 0.05330 |
| CH 016462 | 2 | IDH2 | NM_002168.3:c.419G>A | NP_002159.2:p.Arg140Gln | 0.01420 | 202 | 141 | 77 | 14262 | 9929 | missense | 15 | 90631934 | C | T | Yes | 0.01495 |
| CH 016385 | 2 | FLT3 | NM_004119.2:c.1784_1804dup | NP_004110.2:p.Arg595_Leu601dup | 0.00013 | 8 | NA | 5 | 62213 | NA | FLT3-ITD | 13 | 28608251 | T | TTGAGATCATATTCATATTCTC | Not Done | Not Done |
| CH 016403 | 2 | IDH2 | NM_002168.3:c.419G>A | NP_002159.2:p.Arg140Gln | 0.00949 | 147 | 103 | 67 | 16298 | 10850 | missense | 15 | 90631934 | C | T | Yes | 0.00738 |
| CH 016466 | 2 | IDH1 | NM_005896.3:c.394C>T | NP_005887.2:p.Arg132Cys | 0.00265 | 84 | 54 | 54 | 32440 | 20414 | missense | 2 | 209113113 | G | A | Yes | 0.00252 |
| CH 016487 | 2 | IDH2 | NM_002168.3:c.515G>A | NP_002159.2:p.Arg172Lys | 0.00643 | 127 | 73 | 67 | 17827 | 11349 | missense | 15 | 90631838 | C | T | Yes | 0.00776 |
| CH 016440 | 2 | IDH2 | NM_002168.3:c.419G>A | NP_002159.2:p.Arg140Gln | 0.02493 | 700 | 434 | 196 | 26011 | 17410 | missense | 15 | 90631934 | C | T | Yes | 0.02713 |
| CH 016411 | 2 | FLT3 | NM_004119.2:c.1773_1820dup | NP_004110.2:p.Pro606_Arg607insSerValAspPheArgGluTyrGluTyrAspLeuLysTrpGluPhePro | 0.00062 | 32 | NA | 23 | 51537 | NA | FLT3-ITD | 13 | 28608235 | T | TCTTGGAAACTCCCATTTGAGATCATATTCATATTCTCTGAAATCAACG | Not Done | Not Done |
| CH 016406 | 2 | FLT3 | NM_004119.2:c.1744_1806dup | NP_004110.2:p.Thr582_Lys602dup | 0.00045 | 40 | NA | 20 | 89328 | NA | FLT3-ITD | 13 | 28608249 | A | ATTTGAGATCATATTCATATTCTCTGAAATCAACGTAGAAGTACTCATTATCTGAGGAGCCGGT | Not Done | Not Done |
| CH 016383 | 2 | FLT3 | NM_004119.2:c.1788_1811dup | NP_004110.2:p.Tyr597_Glu604dup | 0.00059 | 41 | NA | 34 | 69605 | NA | FLT3-ITD | 13 | 28608244 | C | CTCCCATTTGAGATCATATTCATAT | Not Done | Not Done |
| CH 016791 | 2 | IDH2 | NM_002168.3:c.419G>A | NP_002159.2:p.Arg140Gln | 0.41526 | 5469 | 3827 | 498 | 13190 | 9216 | missense | 15 | 90631934 | C | T | Yes | 0.40851 |
| CH 016787 | 2 | IDH2 | NM_002168.3:c.419G>A | NP_002159.2:p.Arg140Gln | 0.10845 | 1791 | 1177 | 323 | 16266 | 10853 | missense | 15 | 90631934 | C | T | Yes | 0.11318 |
| CH 016759 | 2 | NPM1 | NM_002520.6:c.860_863dup | NP_002511.1:p.Trp288CysfsTer12 | 0.00012 | 4 | 0 | 2 | 32192 | 11078 | frameshift | 5 | 170837543 | C | CTCTG | No | 0 |
| CH 016788 | 2 | IDH2 | NM_002168.3:c.419G>A | NP_002159.2:p.Arg140Gln | 0.40593 | 7342 | 4870 | 561 | 17918 | 11997 | missense | 15 | 90631934 | C | T | Yes | 0.40696 |
| CH 016799 | 2 | IDH1 | NM_005896.3:c.394C>G | NP_005887.2:p.Arg132Gly | 0.00190 | 54 | 25 | 41 | 23572 | 13146 | missense | 2 | 209113113 | G | C | Yes | 0.00150 |
| CH 017017 | 2 | IDH1 | NM_005896.3:c.394C>T | NP_005887.2:p.Arg132Cys | 0.00223 | 62 | 35 | 45 | 25384 | 15728 | missense | 2 | 209113113 | G | A | Yes | 0.00263 |
| CH 016850 | 2 | FLT3 | NM_004119.2:c.1837+1_1837+2insGGATTTCAGAGAATATGAATATGATCTCAAATGGGAGTTTCCAAGAGAAAATTTAGAGTTTGG | p.? | 0.01351 | 972 | NA | 294 | 71967 | NA | FLT3-ITD | 13 | 28608217 | A | ACCAAACTCTAAATTTTCTCTTGGAAACTCCCATTTGAGATCATATTCATATTCTCTGAAATCC | Not Done | Not Done |
| CH 016850 | 2 | NPM1 | NM_002520.6:c.860_863dup | NP_002511.1:p.Trp288CysfsTer12 | 0.01339 | 343 | 98 | 133 | 25615 | 11911 | frameshift | 5 | 170837543 | C | CTCTG | Yes | 0.01456 |
| CH 016956 | 2 | NPM1 | NM_002520.6:c.860_863dup | NP_002511.1:p.Trp288CysfsTer12 | 0.00101 | 22 | 7 | 16 | 21844 | 9691 | frameshift | 5 | 170837543 | C | CTCTG | Yes | 0.00072 |
| CH 016415 | 2 | FLT3 | NM_004119.2:c.1793_1794insTGGTAATGAGTACTTCTACGTTGATTTCAGAGAATATGA | NP_004110.2:p.Tyr597_Glu598insAspGlyAsnGluTyrPheTyrValAspPheArgGluTyr | 0.00155 | 100 | NA | 53 | 64646 | NA | FLT3-ITD | 13 | 28608262 | T | TTCATATTCTCTGAAATCAACGTAGAAGTACTCATTACCA | Not Done | Not Done |
| CH 016415 | 2 | FLT3 | NM_004119.2:c.1722_1784dup | NP_004110.2:p.Ser574_Phe594dup | 0.00093 | 63 | NA | 37 | 67940 | NA | FLT3-ITD | 13 | 28608271 | T | TCTGAAATCAACGTAGAAGTACTCATTATCTGAGGAGCCGGTCACCTGTACCATCTGTAGCTGG | Not Done | Not Done |
| CH 016415 | 2 | IDH2 | NM_002168.3:c.419G>A | NP_002159.2:p.Arg140Gln | 0.38303 | 7360 | 4550 | 506 | 19332 | 11879 | missense | 15 | 90631934 | C | T | Yes | 0.37251 |
| CH 016441 | 2 | FLT3 | NM_004119.2:c.1734_1787dup | NP_004110.2:p.Val579_Glu596dup | 0.00041 | 26 | NA | 10 | 63431 | NA | FLT3-ITD | 13 | 28608268 | T | TTCTCTGAAATCAACGTAGAAGTACTCATTATCTGAGGAGCCGGTCACCTGTACC | Not Done | Not Done |
| CH 016441 | 2 | FLT3 | NM_004119.2:c.1826_1827insCCTCAAGAGAGAATATGAATATGATCTCAAATGGGAGTTTCCAAGAGAAAA | NP_004110.2:p.Leu610_Glu611insLysArgGluTyrGluTyrAspLeuLysTrpGluPheProArgGluAsnLeu | 0.00015 | 9 | NA | 6 | 61579 | NA | FLT3-ITD | 13 | 28608229 | A | ATTTTCTCTTGGAAACTCCCATTTGAGATCATATTCATATTCTCTCTTGAGG | Not Done | Not Done |
| CH 016441 | 2 | NPM1 | NM_002520.6:c.863_864insCATG | NP_002511.1:p.Trp288CysfsTer12 | 0.00135 | 43 | 11 | 28 | 31966 | 11551 | frameshift | 5 | 170837545 | C | CTGCA | Yes | 0.00096 |
| CH 016464 | 2 | IDH2 | NM_002168.3:c.419G>A | NP_002159.2:p.Arg140Gln | 0.03277 | 738 | 439 | 211 | 20994 | 13397 | missense | 15 | 90631934 | C | T | Yes | 0.03447 |
| CH 016447 | 2 | IDH1 | NM_005896.3:c.394C>T | NP_005887.2:p.Arg132Cys | 0.02676 | 1085 | 608 | 293 | 37934 | 22717 | missense | 2 | 209113113 | G | A | Yes | 0.02671 |
| CH 016412 | 2 | KIT | NM_000222.2:c.2447A>T | NP_000213.1:p.Asp816Val | 0.00091 | 37 | 20 | 32 | 38227 | 21970 | missense | 4 | 55599321 | A | T | Yes | 0.00096 |
| CH 016545 | 2 | FLT3 | NM_004119.2:c.1821_1822insACAGATAATGAGTACTTCTACGTTGATTTCAGAGAATATGAATATGATCTCAAATGGGAGTTTCCAAGA | NP_004110.2:p.Arg607_Glu608insThrAspAsnGluTyrPheTyrValAspPheArgGluTyrGluTyrAspLeuLysTrpGluPheProArg | 0.00160 | 95 | NA | 57 | 59391 | NA | FLT3-ITD | 13 | 28608234 | C | CTCTTGGAAACTCCCATTTGAGATCATATTCATATTCTCTGAAATCAACGTAGAAGTACTCATTATCTGT | Not Done | Not Done |
| CH 016545 | 2 | IDH2 | NM_002168.3:c.419G>A | NP_002159.2:p.Arg140Gln | 0.01312 | 257 | 167 | 121 | 18340 | 12731 | missense | 15 | 90631934 | C | T | Yes | 0.01359 |
| CH 016478 | 2 | NPM1 | NM_002520.6:c.860_863dup | NP_002511.1:p.Trp288CysfsTer12 | 0.00028 | 9 | 0 | 6 | 31718 | 10937 | frameshift | 5 | 170837543 | C | CTCTG | Yes | 0.00054 |
| CH 016474 | 2 | FLT3 | NM_004119.2:c.2503G>T | NP_004110.2:p.Asp835Tyr | 0.00178 | 37 | 26 | 28 | 21114 | 14623 | missense | 13 | 28592642 | C | A | Yes | 0.00172 |
| CH 016463 | 2 | IDH2 | NM_002168.3:c.419G>A | NP_002159.2:p.Arg140Gln | 0.23142 | 4790 | 3076 | 447 | 20377 | 13292 | missense | 15 | 90631934 | C | T | Yes | 0.22202 |
| CH 016475 | 2 | NPM1 | NM_002520.6:c.863_864insCATG | NP_002511.1:p.Trp288CysfsTer12 | 0.00043 | 13 | 5 | 10 | 30025 | 11505 | frameshift | 5 | 170837545 | C | CTGCA | Yes | 0.00032 |
| CH 016489 | 2 | IDH2 | NM_002168.3:c.419G>A | NP_002159.2:p.Arg140Gln | 0.24724 | 4763 | 3161 | 453 | 19129 | 12785 | missense | 15 | 90631934 | C | T | Yes | 0.23721 |
| CH 016484 | 2 | IDH1 | NM_005896.3:c.394C>T | NP_005887.2:p.Arg132Cys | 0.00559 | 203 | 117 | 97 | 33383 | 20940 | missense | 2 | 209113113 | G | A | Yes | 0.00610 |
| CH 016495 | 2 | IDH2 | NM_002168.3:c.419G>A | NP_002159.2:p.Arg140Gln | 0.05485 | 1125 | 699 | 285 | 19490 | 12744 | missense | 15 | 90631934 | C | T | Yes | 0.05698 |
| CH 016672 | 2 | FLT3 | NM_004119.2:c.1781_1782insGCCCGGCTCCTCAGATAATGAGTACTTCTACGTTGATTT | NP_004110.2:p.Asp593_Phe594insLeuProGlySerSerAspAsnGluTyrPheTyrValAsp | 0.01842 | 990 | NA | 262 | 53753 | NA | FLT3-ITD | 13 | 28608274 | G | GAAATCAACGTAGAAGTACTCATTATCTGAGGAGCCGGGC | Not Done | Not Done |
| CH 016606 | 2 | IDH2 | NM_002168.3:c.515G>A | NP_002159.2:p.Arg172Lys | 0.00342 | 83 | 43 | 54 | 19375 | 12574 | missense | 15 | 90631838 | C | T | Yes | 0.00507 |
| CH 016589 | 2 | IDH1 | NM_005896.3:c.394C>T | NP_005887.2:p.Arg132Cys | 0.00250 | 105 | 50 | 60 | 32822 | 19978 | missense | 2 | 209113113 | G | A | Yes | 0.00343 |
| CH 016762 | 2 | IDH2 | NM_002168.3:c.419G>A | NP_002159.2:p.Arg140Gln | 0.00360 | 109 | 53 | 47 | 22281 | 14713 | missense | 15 | 90631934 | C | T | Yes | 0.00336 |

| Subject ID | Cohort | Gene | HGVSc | HGVSp | VAF | Alternate Observations | Deep Alternate Observations | Unique Alternate Observations | Depth | Deep Depth | Variant Consequence | Chromosome | Position | Reference | Alternate | ddPCR Validated | ddPCR VAF |
|---|---|---|---|---|---|---|---|---|---|---|---|---|---|---|---|---|---|
| CH 016604 | 2 | FLT3 | NM_004119.2:c.1830_1831insAACGTTGATTTCAGAGAATATGAATATGATCTCAAATGGGAGTTTCCAAGAGAAAATTTA | NP_004110.2:p.Leu610_Glu611insAsnValAspPheArgGluTyrGluTyrAspLeuLysTrpGluPheProArgGluAsnLeu | 0.00138 | 152 | NA | 105 | 109752 | NA | FLT3-ITD | 13 | 28608225 | C | CTAAATTTTCTCTTGGAAACTCCCATTTGAGATCATATTCATATTCTCTGAAATCAACGTT | Not Done | Not Done |
| CH 016604 | 2 | IDH2 | NM_002168.3:c.419G>A | NP_002159.2:p.Arg140Gln | 0.02952 | 732 | 470 | 203 | 24045 | 15922 | missense | 15 | 90631934 | C | T | Yes | 0.03346 |
| CH 016614 | 2 | FLT3 | NM_004119.2:c.1779_1832dup | NP_004110.2:p.Asp593_Leu610dup | 0.00883 | 927 | NA | 78 | 105025 | NA | FLT3-ITD | 13 | 28608223 | C | CTCTAAATTTTCTCTTGGAAACTCCCATTTGAGATCATATTCATATTCTCTGAAA | Not Done | Not Done |
| CH 016613 | 2 | IDH1 | NM_005896.3:c.395G>A | NP_005887.2:p.Arg132His | 0.02193 | 707 | 364 | 228 | 31069 | 16602 | missense | 2 | 209113112 | C | T | Yes | 0.02610 |
| CH 016613 | 2 | IDH2 | NM_002168.3:c.419G>A | NP_002159.2:p.Arg140Gln | 0.37570 | 7023 | 4765 | 518 | 18624 | 12683 | missense | 15 | 90631934 | C | T | Yes | 0.37459 |
| CH 016802 | 2 | NPM1 | NM_002520.6:c.860_863dup | NP_002511.1:p.Trp288CysfsTer12 | 0.00012 | 5 | 1 | 4 | 40005 | 13505 | frameshift | 5 | 170837543 | C | CTCTG | No | 0 |
| CH 016650 | 2 | IDH2 | NM_002168.3:c.419G>A | NP_002159.2:p.Arg140Gln | 0.32363 | 6062 | 4134 | 472 | 18676 | 12774 | missense | 15 | 90631934 | C | T | Yes | 0.31988 |
| CH 016801 | 2 | FLT3 | NM_004119.2:c.1787_1788insTGATAATGAGTACTTCTACGTTGATTTCAGAGA | NP_004110.2:p.Arg595_Glu596insAspAspAsnGluTyrPheTyrValAspPheArg | 0.00415 | 303 | NA | 128 | 72988 | NA | FLT3-ITD | 13 | 28608268 | T | TTCTCTGAAATCAACGTAGAAGTACTCATTATCA | Not Done | Not Done |
| CH 016801 | 2 | FLT3 | NM_004119.2:c.1778_1837dup | NP_004110.2:p.Asp593_Phe612dup | 0.00012 | 14 | NA | 8 | 120268 | NA | FLT3-ITD | 13 | 28608218 | C | CCAAACTCTAAATTTTCTCTTGGAAACTCCCATTTGAGATCATATTCATATTCTCTGAAAT | Not Done | Not Done |
| CH 016801 | 2 | NPM1 | NM_002520.6:c.860_863dup | NP_002511.1:p.Trp288CysfsTer12 | 0.01874 | 592 | 94 | 185 | 31583 | 10937 | frameshift | 5 | 170837543 | C | CTCTG | Yes | 0.01990 |
| CH 016844 | 2 | FLT3 | NM_004119.2:c.1792_1793insCCCTACCGGCTCCTCAGATAATGAGTACTTCTACGTTGATTTCAGAGAATATGA | NP_004110.2:p.Glu598delinsAlaLeuProAlaProGlnIleMetSerThrSerThrLeuIleSerGluAsnMetLys | 0.00045 | 23 | NA | 14 | 51499 | NA | FLT3-ITD | 13 | 28608262 | T | TTCATATTCTCTGAAATCAACGTAGAAGTACTCATTATCTGAGGAGCCGGTAGGG | Not Done | Not Done |
| CH 016844 | 2 | NPM1 | NM_002520.6:c.863_864insTTTG | NP_002511.1:p.Trp288CysfsTer12 | 0.00094 | 24 | 11 | 18 | 25609 | 11715 | frameshift | 5 | 170837545 | C | CTGTT | Not Done | Not Done |
| CH 016995 | 2 | IDH1 | NM_005896.3:c.394C>T | NP_005887.2:p.Arg132Cys | 0.02724 | 904 | 536 | 236 | 32960 | 19677 | missense | 2 | 209113113 | G | A | Yes | 0.02704 |
| CH 016871 | 2 | IDH2 | NM_002168.3:c.419G>A | NP_002159.2:p.Arg140Gln | 0.01451 | 427 | 237 | 166 | 26992 | 16330 | missense | 15 | 90631934 | C | T | Yes | 0.01772 |
| CH 016808 | 2 | IDH2 | NM_002168.3:c.419G>A | NP_002159.2:p.Arg140Gln | 0.00256 | 113 | 57 | 64 | 33136 | 22284 | missense | 15 | 90631934 | C | T | Yes | 0.00302 |
| CH 016857 | 2 | IDH2 | NM_002168.3:c.515G>A | NP_002159.2:p.Arg172Lys | 0.00067 | 30 | 13 | 25 | 35285 | 19504 | missense | 15 | 90631838 | C | T | Yes | 0.00063 |
| CH 016829 | 2 | IDH1 | NM_005896.3:c.394C>A | NP_005887.2:p.Arg132Ser | 0.00114 | 124 | 20 | 76 | 51081 | 17490 | missense | 2 | 209113113 | G | T | Yes | 0.00229 |
| CH 016843 | 2 | IDH2 | NM_002168.3:c.419G>A | NP_002159.2:p.Arg140Gln | 0.01880 | 616 | 345 | 190 | 29448 | 18351 | missense | 15 | 90631934 | C | T | Yes | 0.02560 |
| CH 016996 | 2 | NPM1 | NM_002520.6:c.863_864insCCTG | NP_002511.1:p.Trp288CysfsTer12 | 0.00115 | 42 | 4 | 27 | 36664 | 10022 | frameshift | 5 | 170837544 | T | TCTGC | Yes | 0.00102 |
| CH 016825 | 2 | IDH1 | NM_005896.3:c.395G>A | NP_005887.2:p.Arg132His | 0.30645 | 14168 | 7169 | 872 | 44813 | 23394 | missense | 2 | 209113112 | C | T | Yes | 0.31749 |
| CH 016582 | 2 | IDH2 | NM_002168.3:c.419G>A | NP_002159.2:p.Arg140Gln | 0.00095 | 58 | 19 | 35 | 31330 | 19911 | missense | 15 | 90631934 | C | T | Yes | 0.00131 |
| CH 016949 | 2 | IDH2 | NM_002168.3:c.419G>A | NP_002159.2:p.Arg140Gln | 0.07351 | 2284 | 1388 | 377 | 29986 | 18883 | missense | 15 | 90631934 | C | T | Yes | 0.08409 |
| CH 017013 | 2 | KIT | NM_000222.2:c.2446G>T | NP_000213.1:p.Asp816Tyr | 0.00043 | 22 | 13 | 16 | 53601 | 29946 | missense | 4 | 55599320 | G | T | Not Done | Not Done |
| CH 017037 | 2 | FLT3 | NM_004119.2:c.1756_1818dup | NP_004110.2:p.Asp586_Pro606dup | 0.00059 | 62 | NA | 37 | 104591 | NA | FLT3-ITD | 13 | 28608237 | T | TTGGAAACTCCCATTTGAGATCATATTCATATTCTCTGAAATCAACGTAGAAGTACTCATTATC | Not Done | Not Done |
| CH 017037 | 2 | NPM1 | NM_002520.6:c.860_863dup | NP_002511.1:p.Trp288CysfsTer12 | 0.00053 | 24 | 3 | 13 | 44978 | 15236 | frameshift | 5 | 170837543 | C | CTCTG | Yes | 0.00057 |
| CH 016969 | 2 | IDH1 | NM_005896.3:c.394C>T | NP_005887.2:p.Arg132Cys | 0.00994 | 555 | 237 | 200 | 47267 | 23840 | missense | 2 | 209113113 | G | A | Yes | 0.01298 |
| CH 016901 | 2 | FLT3 | NM_004119.2:c.1780_1800dup | NP_004110.2:p.Phe594_Asp600dup | 0.00012 | 13 | NA | 10 | 111256 | NA | FLT3-ITD | 13 | 28608255 | G | GATCATATTCATATTCTCTGAA | Not Done | Not Done |
| CH 016862 | 2 | FLT3 | c.? | p.? | 0.00259 | 475 | NA | 132 | 183073 | NA | FLT3-ITD | 13 | 28608065 | T | TTGCTAATTCCATAAGCTGTTGCGTTCATCACTTTTCCAAAAGCACCTGATCCTAGTACCTTCCCTGCAAAGACAAATGGTGAGTACGTGCATTTTAAAGATTTTCCAATGGAAAAGAAATGCTGCAGAAACATTTGGCACATTCCATTCTTACCAAACTCTAAATTTTCTCTTGGAAACTCCCATTTGAGATCATATTCATATTCTC | Not Done | Not Done |
| CH 016862 | 2 | NPM1 | NM_002520.6:c.860_863dup | NP_002511.1:p.Trp288CysfsTer12 | 0.00363 | 118 | 5 | 65 | 32507 | 6098 | frameshift | 5 | 170837543 | C | CTCTG | Yes | 0.00499 |
| CH 016877 | 2 | IDH2 | NM_002168.3:c.419G>A | NP_002159.2:p.Arg140Gln | 0.00123 | 53 | 16 | 38 | 29068 | 13031 | missense | 15 | 90631934 | C | T | Yes | 0.00166 |
| CH 016892 | 2 | IDH2 | NM_002168.3:c.419G>A | NP_002159.2:p.Arg140Gln | 0.14697 | 4728 | 2945 | 507 | 31527 | 20038 | missense | 15 | 90631934 | C | T | Yes | 0.16445 |
| CH 017043 | 2 | IDH1 | NM_005896.3:c.394C>T | NP_005887.2:p.Arg132Cys | 0.00655 | 275 | 135 | 136 | 36039 | 20601 | missense | 2 | 209113113 | G | A | Yes | 0.00789 |
| CH 017073 | 2 | IDH1 | NM_005896.3:c.394C>T | NP_005887.2:p.Arg132Cys | 0.04227 | 1356 | 764 | 330 | 30626 | 18074 | missense | 2 | 209113113 | G | A | Yes | 0.04679 |
| CH 016858 | 2 | IDH2 | NM_002168.3:c.419G>A | NP_002159.2:p.Arg140Gln | 0.01398 | 547 | 272 | 178 | 33308 | 19462 | missense | 15 | 90631934 | C | T | Yes | 0.01958 |
| CH 016869 | 2 | FLT3 | NM_004119.2:c.2505T>G | NP_004110.2:p.Asp835Glu | 0.00155 | 49 | 30 | 29 | 29919 | 19320 | missense | 13 | 28592640 | A | C | Not Done | Not Done |
| CH 016924 | 2 | IDH2 | NM_002168.3:c.516_518inv | NP_002159.2:p.Arg172_His173delinsSerAla | 0.05027 | 1517 | 965 | 342 | 29537 | 19197 | missense | 15 | 90631835 | TGC | GCA | Not Done | Not Done |
| CH 016924 | 2 | NPM1 | NM_002520.6:c.860_863dup | NP_002511.1:p.Trp288CysfsTer12 | 0.00040 | 17 | 3 | 10 | 42159 | 14323 | frameshift | 5 | 170837543 | C | CTCTG | Yes | 0.00043 |
| CH 016971 | 2 | FLT3 | NM_004119.2:c.1773_1799dup | NP_004110.2:p.Val592_As:600dup | 0.00064 | 68 | NA | 53 | 106434 | NA | FLT3-ITD | 13 | 28608256 | A | ATCATATTCATATTCTCTGAAATCAACG | Not Done | Not Done |
| CH 016971 | 2 | NPM1 | NM_002520.6:c.860_863dup | NP_002511.1:p.Trp288CysfsTer12 | 0.00847 | 356 | 35 | 129 | 42013 | 11972 | frameshift | 5 | 170837543 | C | CTCTG | Yes | 0.00894 |
| CH 016887 | 2 | NPM1 | NM_002520.6:c.863_864insCTTG | NP_002511.1:p.Trp288CysfsTer12 | 0.00070 | 25 | 2 | 18 | 35823 | 8060 | frameshift | 5 | 170837545 | C | CTGCT | Not Done | Not Done |
| CH 016893 | 2 | IDH1 | NM_005896.3:c.394C>T | NP_005887.2:p.Arg132Cys | 0.08262 | 4273 | 1703 | 568 | 44684 | 20612 | missense | 2 | 209113113 | G | A | Yes | 0.10241 |
| CH 016921 | 2 | NPM1 | NM_002520.6:c.863_864insCCTG | NP_002511.1:p.Trp288CysfsTer12 | 0.00112 | 38 | 4 | 22 | 33947 | 11383 | frameshift | 5 | 170837544 | T | TCTGC | Yes | 0.00117 |
| CH 016882 | 2 | IDH2 | NM_002168.3:c.419G>A | NP_002159.2:p.Arg140Gln | 0.27341 | 4766 | 3238 | 485 | 17605 | 11843 | missense | 15 | 90631934 | C | T | Yes | 0.27170 |
| CH 016916 | 2 | IDH2 | NM_002168.3:c.419G>A | NP_002159.2:p.Arg140Gln | 0.17988 | 4021 | 2691 | 495 | 22039 | 14960 | missense | 15 | 90631934 | C | T | Yes | 0.18173 |
| CH 016911 | 2 | FLT3 | NM_004119.2:c.2525A>G | NP_004110.2:p.Tyr842Cys | 0.50117 | 8994 | 6448 | 803 | 17818 | 12866 | missense | 13 | 28592620 | T | C | Not Done | Not Done |
| CH 016856 | 2 | FLT3 | NM_004119.2:c.1782_1829dup | NP_004110.2:p.Phe594_Asn609dup | 0.01589 | 1612 | NA | 382 | 101434 | NA | FLT3-ITD | 13 | 28608226 | T | TAAATTTTCTCTTGGAAACTCCCATTTGAGATCATATTCATATTCTCTG | Not Done | Not Done |
| CH 016856 | 2 | IDH2 | NM_002168.3:c.419G>A | NP_002159.2:p.Arg140Gln | 0.01306 | 412 | 240 | 148 | 28495 | 18376 | missense | 15 | 90631934 | C | T | Yes | 0.01695 |
| CH 016856 | 2 | NPM1 | NM_002520.6:c.863_864insCCTG | NP_002511.1:p.Trp288CysfsTer12 | 0.01110 | 402 | 47 | 138 | 36207 | 8685 | frameshift | 5 | 170837544 | T | TCTGC | Yes | 0.01413 |
| CH 017057 | 2 | IDH2 | NM_002168.3:c.419G>A | NP_002159.2:p.Arg140Gln | 0.01648 | 315 | 196 | 128 | 17778 | 11895 | missense | 15 | 90631934 | C | T | Yes | 0.01749 |
| CH 017057 | 2 | NPM1 | NM_002520.6:c.860_863dup | NP_002511.1:p.Trp288CysfsTer12 | 0.00011 | 3 | 1 | 2 | 26673 | 9008 | frameshift | 5 | 170837543 | C | CTCTG | No | 0 |
| CH 016999 | 2 | FLT3 | NM_004119.2:c.2503G>T | NP_004110.2:p.Asp835Tyr | 0.00103 | 14 | 11 | 12 | 16158 | 10631 | missense | 13 | 28592642 | C | A | Yes | 0.00077 |
| CH 016903 | 2 | IDH2 | NM_002168.3:c.419G>A | NP_002159.2:p.Arg140Gln | 0.00262 | 79 | 35 | 35 | 20756 | 13367 | missense | 15 | 90631934 | C | T | Yes | 0.00313 |
| CH 016946 | 2 | IDH1 | NM_005896.3:c.394C>T | NP_005887.2:p.Arg132Cys | 0.03361 | 1023 | 511 | 275 | 26841 | 15202 | missense | 2 | 209113113 | G | A | Yes | 0.03914 |
| CH 016958 | 2 | IDH1 | NM_005896.3:c.394C>G | NP_005887.2:p.Arg132Gly | 0.35310 | 6344 | 3132 | 534 | 18124 | 8870 | missense | 2 | 209113113 | G | C | Yes | 0.35364 |
| CH 016919 | 2 | FLT3 | NM_004119.2:c.1797_1798insTCCCCGACCGGCTCCTCAGATAATGAGTACTTCTACGTTGATTTCAGAGAATATGAATAT | NP_004110.2:p.Tyr599_Asp600insSerProThrGlySerSerAspAsnGluTyrPheTyrValAspPheArgGluTyrGluTyr | 0.00018 | 11 | NA | 7 | 62140 | NA | FLT3-ITD | 13 | 28608258 | C | CATATTCATATTCTCTGAAATCAACGTAGAAGTACTCATTATCTGAGGAGCCGGTCGGGGA | Not Done | Not Done |
| CH 016919 | 2 | IDH2 | NM_002168.3:c.419G>A | NP_002159.2:p.Arg140Gln | 0.02537 | 526 | 325 | 170 | 19894 | 12812 | missense | 15 | 90631934 | C | T | Yes | 0.02890 |
| CH 016937 | 2 | IDH2 | NM_002168.3:c.419G>A | NP_002159.2:p.Arg140Gln | 0.00112 | 29 | 9 | 19 | 15569 | 8043 | missense | 15 | 90631934 | C | T | Yes | 0.00100 |
| CH 016964 | 2 | IDH2 | NM_002168.3:c.419G>A | NP_002159.2:p.Arg140Gln | 0.00284 | 40 | 27 | 26 | 13565 | 9511 | missense | 15 | 90631934 | C | T | Not Done | Not Done |
| CH 016962 | 2 | IDH2 | NM_002168.3:c.419G>A | NP_002159.2:p.Arg140Gln | 0.29249 | 4087 | 2783 | 437 | 13819 | 9515 | missense | 15 | 90631934 | C | T | Yes | 0.28943 |
| CH 017026 | 2 | IDH2 | NM_002168.3:c.419G>A | NP_002159.2:p.Arg140Gln | 0.08160 | 1770 | 1163 | 311 | 21411 | 14252 | missense | 15 | 90631934 | C | T | Yes | 0.08612 |
| CH 017056 | 2 | FLT3 | NM_004119.2:c.1742_1838-2dup | p.? | 0.00186 | 208 | NA | 53 | 112100 | NA | FLT3-ITD | 13 | 28608129 | C | CTGCAAAGACAAATGGTGAGTACGTGCATTTTAAAGATTTTCCAATGGAAAAGAAATGCTGCAGAAACATTTGGCACATTCCATTCTTACCAAACTCTAAATTTTCTCTTGGAAACTCCCATTTGAGATCATATTCATATTCTCTGAAATCAACGTAGAAGTACTCATTATCTGAGGAGCCGGTCA | Not Done | Not Done |
| CH 017008 | 2 | IDH1 | NM_005896.3:c.395G>A | NP_005887.2:p.Arg132His | 0.01655 | 587 | 253 | 200 | 34690 | 15290 | missense | 2 | 209113112 | C | T | Yes | 0.01861 |
| CH 016991 | 2 | FLT3 | NM_004119.2:c.1806_1807insACCCCTCGTTTAGGTTTCAGAGAATATGAATATGATCTCAAA | NP_004110.2:p.Lys602_Trp603insThrProArgLeuGlyPheArgGluTyrGluTyrAspLeuLys | 0.00223 | 169 | NA | 90 | 75937 | NA | FLT3-ITD | 13 | 28608249 | A | ATTTGAGATCATATTCATATTCTCTGAAACCTAAACGAGGGGT | Not Done | Not Done |
| CH 017019 | 2 | NPM1 | NM_002520.6:c.863_864insCATG | NP_002511.1:p.Trp288CysfsTer12 | 0.00016 | 5 | 0 | 4 | 31099 | 6031 | frameshift | 5 | 170837545 | C | CTGCA | Yes | 0.00018 |
| CH 017036 | 2 | NPM1 | NM_002520.6:c.860_863dup | NP_002511.1:p.Trp288CysfsTer12 | 0.00013 | 5 | 0 | 2 | 37559 | 9258 | frameshift | 5 | 170837543 | C | CTCTG | Yes | 0.00006 |
| CH 017078 | 2 | IDH1 | NM_005896.3:c.394C>T | NP_005887.2:p.Arg132Cys | 0.00241 | 120 | 42 | 56 | 33768 | 17434 | missense | 2 | 209113113 | G | A | Yes | 0.00373 |

| Subject ID | Cohort | Gene | HGVSc | HGVSp | VAF | Alternate Observations | Deep Alternate Observations | Unique Alternate Observations | Depth | Deep Depth | Variant Consequence | Chromosome | Position | Reference | Alternate | ddPCR Validated | ddPCR VAF |
|---|---|---|---|---|---|---|---|---|---|---|---|---|---|---|---|---|---|
| CH 017003 | 2 | FLT3 | NM_004119.2:c.1759_1800dup | NP_004110.2:p.Asn587_Asp600dup | 0.01070 | 686 | NA | 159 | 64131 | NA | FLT3-ITD | 13 | 28608255 | G | GATCATATTCATATTCTCTGAAATCAACGTAGAAG TACTCATT | Not Done | Not Done |
| CH 017003 | 2 | IDH2 | NM_002168.3:c.419G>A | NP_002159.2:p.Arg140Gln | 0.00408 | 94 | 47 | 61 | 19298 | 11513 | missense | 15 | 90631934 | C | T | Yes | 0.00614 |
| CH 017022 | 2 | IDH1 | NM_005896.3:c.395G>A | NP_005887.2:p.Arg132His | 0.48460 | 25024 | 7806 | 884 | 51843 | 16108 | missense | 2 | 209113112 | C | T | Yes | 0.48050 |
| CH 017027 | 2 | FLT3 | NM_004119.2:c.1751_1810dup | NP_004110.2:p.Trp603_Glu604insAlaSerAspAsnGluTyrP heTyrValAspPheArgGluTyrGluTyrAspLeuLysTrp | 0.02031 | 1631 | NA | 166 | 80310 | NA | FLT3-ITD | 13 | 28608245 | T | TCCCATTTGAGATCATATTCATATTCTCTGAAATC AACGTAGAAGTACTCATTATCTGAGG | Not Done | Not Done |
| CH 017053 | 2 | IDH1 | NM_005896.3:c.394C>G | NP_005887.2:p.Arg132Gly | 0.00102 | 31 | 17 | 22 | 34789 | 16631 | missense | 2 | 209113113 | G | C | Yes | 0.00050 |
| CH 017091 | 2 | IDH1 | NM_005896.3:c.394C>G | NP_005887.2:p.Arg132Gly | 0.01210 | 479 | 224 | 158 | 35214 | 18509 | missense | 2 | 209113113 | G | C | Yes | 0.00598 |
| CH 017031 | 2 | FLT3 | NM_004119.2:c.1804_1805insCTGTTGGTAATGAG TACTTCTACGTTGATTTCAGAGAATATGAATATGA TCTCA | NP_004110.2:p.Leu601_Lys602insThrValGlyAsnGluTyrP heTyrValAspPheArgGluTyrGluTyrAspLeu | 0.00171 | 141 | NA | 63 | 82519 | NA | FLT3-ITD | 13 | 28608251 | T | TTGAGATCATATTCATATTCTCTGAAATCAACGTA GAAGTACTCATTACCAACAG | Not Done | Not Done |
| CH 017104 | 2 | IDH2 | NM_002168.3:c.419G>A | NP_002159.2:p.Arg140Gln | 0.12163 | 3209 | 1952 | 394 | 25304 | 16049 | missense | 15 | 90631934 | C | T | Yes | 0.12794 |
| CH 017495 | 2 | IDH1 | NM_005896.3:c.394C>G | NP_005887.2:p.Arg132Gly | 0.12252 | 5616 | 1950 | 555 | 43459 | 15916 | missense | 2 | 209113113 | G | C | Yes | 0.12944 |
| CH 017055 | 2 | FLT3 | NM_004119.2:c.2503G>T | NP_004110.2:p.Asp835Tyr | 0.01151 | 305 | 167 | 138 | 24289 | 14509 | missense | 13 | 28592642 | C | A | Yes | 0.01248 |
| CH 017103 | 2 | IDH2 | NM_002168.3:c.419G>A | NP_002159.2:p.Arg140Gln | 0.00078 | 21 | 12 | 18 | 24169 | 15386 | missense | 15 | 90631934 | C | T | Yes | 0.00054 |
| CH 017051 | 2 | FLT3 | NM_004119.2:c.1799_1800insAACTCAAAGCCAGCT ACAGATGGTACAGGTGACCGGCTCCTCAGATAA TGAGTACTTCTACGTTGATTTCAGAGAATATGAAT ATGA | NP_004110.2:p.Tyr599_Asp600insGluThrGlnSerGlnLeu GlnMetValGlnValThrGlySerSerAspAsnGluTyrPheTyrValA spPheArgGluTyrGluTyr | 0.00232 | 153 | NA | 65 | 65982 | NA | FLT3-ITD | 13 | 28608256 | A | ATCATATTCATATTCTCTGAAATCAACGTAGAAGT ACTCATTATCTGAGGAGCCGGTCACCTGTACCAT CTGTAGCTGGCTTTGAGTT | Not Done | Not Done |
| CH 017051 | 2 | IDH2 | NM_002168.3:c.419G>A | NP_002159.2:p.Arg140Gln | 0.00081 | 18 | 11 | 12 | 20148 | 13651 | missense | 15 | 90631934 | C | T | Yes | 0.00092 |
| CH 017059 | 2 | IDH2 | NM_002168.3:c.419G>A | NP_002159.2:p.Arg140Gln | 0.00233 | 67 | 34 | 41 | 22900 | 14572 | missense | 15 | 90631934 | C | T | Yes | 0.00397 |
| CH 017116 | 2 | IDH2 | NM_002168.3:c.419G>A | NP_002159.2:p.Arg140Gln | 0.00575 | 123 | 76 | 61 | 20693 | 13214 | missense | 15 | 90631934 | C | T | Yes | 0.00577 |
| CH 017054 | 2 | IDH2 | NM_002168.3:c.515G>A | NP_002159.2:p.Arg172Lys | 0.08083 | 1707 | 1107 | 319 | 20929 | 13695 | missense | 15 | 90631838 | C | T | Yes | 0.09242 |
| CH 017058 | 2 | FLT3 | NM_004119.2:c.1748_1795dup | NP_004110.2:p.Glu598_Tyr599insCysSerSerAspAsnGlu TyrPheTyrValAspPheArgGluTyrGlu | 0.01288 | 882 | NA | 229 | 68464 | NA | FLT3-ITD | 13 | 28608260 | T | TATTCATATTCTCTGAAATCAACGTAGAAGTACTC ATTATCTGAGGAGC | Not Done | Not Done |
| CH 017058 | 2 | NPM1 | NM_002520.6:c.860_863dup | NP_002511.1:p.Trp288CysfsTer12 | 0.00680 | 189 | 26 | 74 | 27780 | 9303 | frameshift | 5 | 170837543 | C | CTCTG | Yes | 0.00769 |
| CH 017093 | 2 | IDH2 | NM_002168.3:c.419G>A | NP_002159.2:p.Arg140Gln | 0.02229 | 676 | 385 | 209 | 28982 | 17272 | missense | 15 | 90631934 | C | T | Yes | 0.02612 |
| CH 016646 | 2 | NPM1 | NM_002520.6:c.860_863dup | NP_002511.1:p.Trp288CysfsTer12 | 0.00042 | 10 | 2 | 5 | 23985 | 4442 | frameshift | 5 | 170837543 | C | CTCTG | Yes | 0.00050 |
| CH 016646 | 2 | FLT3 | NM_004119.2:c.1799_1800insAGATTTCAGAGAATA TGAATATGA | NP_004110.2:p.Tyr599_Asp600insGluAspPheArgGluTyr GluTyr | 0.00016 | 9 | NA | 7 | 55399 | NA | FLT3-ITD | 13 | 28608256 | A | ATCATATTCATATTCTCTGAAATCT | Not Done | Not Done |

VAF, variant allele frequency; ddPCR, digital droplet PCR; NA, not applicable
